# Immunotherapy and Cannabis: A Harmful Drug Interaction or Reefer Madness?

**DOI:** 10.1101/2024.01.26.24301817

**Authors:** Brian J. Piper, Maria Tian, Pragosh Saini, Ahmad Higazy, Jason Graham, Christian J. Carbe, Michael Bordonaro

## Abstract

A retrospective (N=140) and a prospective (N=102) observational Israeli study by Bar-Sela and colleagues about cannabis potentially adversely impacting the response to immunotherapy have together been cited 191 times including by clinical practice guidelines. There have also been reports on PubPeer outlining unverifiable information in their statistics and numerous discrepancies calculating percentages. This report attempted to replicate the data-analysis including non-parametric statistics. Table 1 of the corrected prospective report contained 22 p-values but only one (4.5%) could be verified, despite the authors being transparent about the N and statistics employed. Cannabis users were significantly (p < .0025) younger than non-users but this was not reported in the retrospective report. There were also errors in percentage calculations (e.g. 13/34 reported as 22.0% instead of 38.2%). Overall, these observational investigations, and especially the prospective, appear to contain gross inaccuracies which could impact the statistical decisions (i.e. significant findings reported as non-significant or vice-versa). Although it is mechanistically plausible that cannabis could have immunosuppressive effects which inhibit the response to immuno-therapy, these two reports should be viewed cautiously. Larger prospective studies of this purported drug interaction that account for potential confounds (e.g. greater nicotine smoking among cannabis users) may be warranted. 198 / 200 words

**Simple Summary:** Two Israeli studies about medical marijuana potentially interfering with immunotherapies like nivolumab for cancer treatment have received substantial attention. However, there have been anonymous but detailed concerns about these reports on PubPeer. This team attempted to verify the data-analysis and statistics of these two reports and the published correction. Many findings including some that could impact the statistical conclusions could not be verified. Of the 22 statistical tests on Table 1 of the prospective report, six could not be repeated using the same statistics and with the provided N. The p-value on 15 corresponded with that of a different statistical test than was listed in the methods. Re-analysis also identified some previously unreported significant differences (e.g. age) between cannabis users and non-users at baseline. Further study of the safety of immunotherapy and cannabis combination may be warranted using patient groups that have been matched on key demographic and medical variables. 150 / 150 words

**Graphical abstract:** 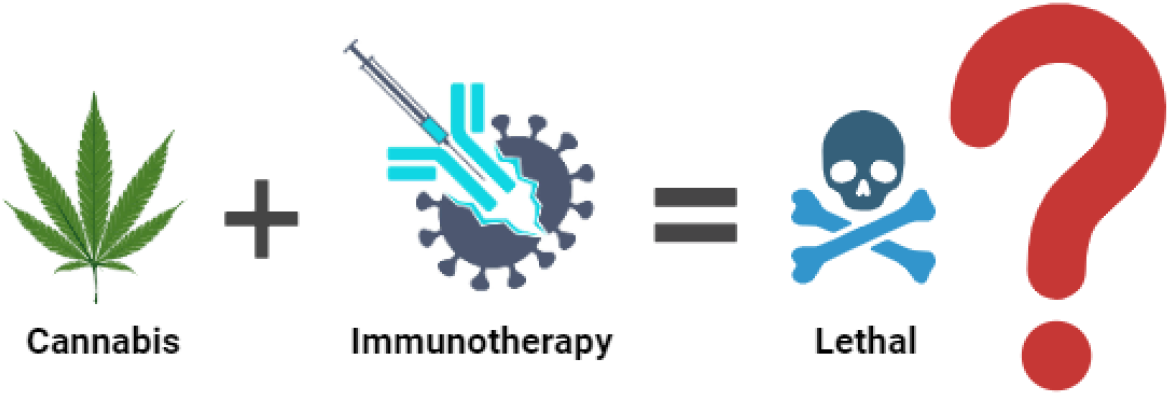

## 1. Introduction

The National Academy of Sciences (NAS) 2017 cannabis report [1] provides key information for oncology patients who would like their decision of whether to use medical marijuana to be empirically informed. There was substantial or conclusive evidence that oral cannabinoids were effective for chemotherapy induced nausea and vomiting. Similarly, the NAS rated the strength of the evidence of cannabis or cannabinoids being effective for the treatment of chronic pain as substantial/conclusive. However, evidence was rated as limited that cannabis was effective for improving anxiety symptoms. There was moderate evidence that cannabis and cannabinoids caused a small increased risk for the development of depressive disorders [1]. Evidence was rated insufficient to support or refute cannabinoids as an effective treatment for cancer-associated anorexia-cachexia syndrome [1]. There was also limited evidence of a statistical association between cannabis smoking and decreased production of inflammatory cytokines in healthy individuals [1]. The American Cancer Society does not take position for, or against, use of medical cannabis [2].

**Table 1.** Results of re-analysis of the Bar-Sela correction [4] with total N = 102 evaluating potential adverse effects of cannabis on immunotherapy using ^C^chi-square, ^F^Fisher’s or ^Y^Yates non-parametric statistics. Minimum expected cell N determined with [20]. The British Medical Journal (BMJ) guidance on the recommend non-parametric statistic is reported [11]. Atezo: atezolizumab, COPD: Chronic Obstructive Pulmonary Disease, ECOG: Eastern Cooperative Oncology Group, LC: Lung Cancer, nivo: nivolumab.

| Variable | Smallest Cell N observed<br>(expected) | BMJ statistic | Reported p | Calculated p | Inter |
| --- | --- | --- | --- | --- | --- |
| Gender | 10 (10.67) | chi-square | .9399<sup>C or F</sup> | .9399<sup>Y</sup> | Mis |
| ECOG | 10 (7.67) | chi-square | .3568<sup>C or F</sup> | .3568<sup>Y</sup> | Mis |
| Chronic diseases = 0 | 13 (11.67) | chi-square | .7124<sup>C or F</sup> | .4908<sup>C</sup>, .5136<sup>F</sup>, .6398<sup>Y</sup> | Un |
| Chronic diseases = 1 | 7 (7.67) | chi-square | .9332<sup>C or F</sup> | .9332<sup>Y</sup> | Mis |
| Chronic diseases = 2+ | 14 (14.67) | chi-square | .9437<sup>C or F</sup> | .9437<sup>Y</sup> | Mis |
| Chronic heart disease | 5 (7.67) | chi-square | .2762<sup>C or F</sup> | .2762<sup>Y</sup> | Mis |
| Diabetes | 6 (7.67) | chi-square | .5576<sup>C or F</sup> | .5576<sup>Y</sup> | Mis |
| High blood pressure | 13 (15.67) | chi-square | .3612<sup>C or F</sup> | .3612<sup>Y</sup> | Mis |
| COPD | 3 (4.00) | chi-square | 1<sup>C or F</sup> | .5145<sup>C</sup>, .7463<sup>F</sup>, .7445<sup>Y</sup> | Un |
| Hyperlipidemia | 7 (10.00) | chi-square | .2491<sup>C or F</sup> | .1436<sup>C</sup>, .1737<sup>F</sup>, .2714<sup>Y</sup> | Un |
| Other disease | 0 (-) | chi-square | 1<sup>C or F</sup> | .3125<sup>C</sup>, .5512<sup>F</sup>, .8007<sup>Y</sup> | Un |
| Non-small cell LC | 14 (15.00) | chi-square | .8325<sup>C or F</sup> | .8325<sup>Y</sup> | Mis |
| Melanoma | 9 (11.33) | chi-square | .414<sup>C or F</sup> | .4140<sup>Y</sup> | Mis |
| Renal Cell Carcinoma | 2 (2.00) | chi-square | 1<sup>C or F</sup> | 1.000<sup>C,F,Y</sup> | V |
| Other malignancy | 3 (1.67) | chi-square | 1<sup>C or F</sup> | .1946<sup>C</sup>, .3300<sup>F</sup>, .4175<sup>Y</sup> | Un |
| Brain metastasis | 8 (6.67) | chi-square | .6993<sup>C or F</sup> | .6993<sup>Y</sup> | Mis |
| Lungs metastasis | 11 (13.33) | chi-square | .4303<sup>C or F</sup> | .4303<sup>Y</sup> | Mis |
| Liver metastasis | 11 (8.00) | chi-square | .2157<sup>C or F</sup> | .2157<sup>Y</sup> | Mis |
| Immunotherapy 1<sup>st</sup> line | 8 (13.00) | chi-square | .05178<sup>C or F</sup> | .0518<sup>Y</sup> | Mis |
| Pembrolizumab or nivo | 5 (8.67) | chi-square | .127<sup>C or F</sup> | .1270<sup>Y</sup> | Mis |
| Ipilimumab & nivo | 4 (6.67) | chi-square | .2517<sup>C or F</sup> | .2517<sup>Y</sup> | Mis |
| Durvalumab or atezo | 1 (2.00) | chi-square | 1<sup>C or F</sup> | .3720<sup>C</sup>, .6607<sup>F</sup>, .6554<sup>Y</sup> | Un |

Cancer is a complex disease process traditionally managed with medical treatments that have progressed from surgical resection, radiation therapy, chemotherapy, and targeted drug therapy to most recently immunotherapy [3, 4]. Unlike traditional therapy, immunotherapy utilizes monoclonal antibodies, small molecule drugs, adoptive cell therapy, oncolytic viruses, and cancer vaccines to activate the body’s innate and adaptive immune responses to produce anticancer effects to kill and eliminate tumor cells [4]. However, tumor cells have adapted the ability to express inhibitory “checkpoint” proteins (i.e., cell surface signaling molecules normally expressed in healthy cells that safeguard against dysregulated immunity capable of subjecting the host to immunodeficiency, autoimmunity, malignancy, or harmful immune responses to infectious agents), resulting in decreased function of antigen-specific T cells and ultimately preventing T cells from recognizing and attacking cancer cells [3, 5]. Checkpoint inhibition therapy is a form of cancer immunotherapy that employs antibodies against T cell or antigen presenting cell surface regulators of immune cell inhibition, (i.e., checkpoint inhibitors), to then activate cytotoxic T cells to assist in the killing of tumor cells [5]. Two major immune checkpoint pathways that are currently targeted in oncologic immunotherapeutics are the cytotoxic T-lymphocyte–associated antigen 4 (CTLA-4) which regulate T-cell proliferation primarily in lymph nodes early in an immune response, and the programmed cell death protein (PD-1) pathways which suppresses T cells in peripheral tissues later the immune response [5, 6]. Inhibition of these targets, resulting in increased activation of the immune system, has led to new immunotherapies for melanoma, non–small cell lung cancer, and several other cancers [6]. Ipilimumab, an inhibitor of CTLA-4, is approved for the treatment of advanced or unresectable melanoma [3, 7]. Nivolumab and pembrolizumab, both PD-1 inhibitors, are approved to treat patients with advanced or metastatic melanoma, patients with metastatic refractory non-small cell lung cancer, patients with renal cell carcinoma, or patients with Hodgkin’s lymphoma (pembrolizumab), or head and neck squamous cell carcinoma (pembrolizumab), or hepatocellular carcinoma (nivolumab) [1, 7]. Atezolizumab, avelumab, and durvamulab are PD-1 ligand (PDL-1) inhibitors approved to treat urothelial carcinoma (all), non-small cell lung cancer (atezolizumab and durvalumab), or Merkel cell carcinoma (avelumab), [3,7].

Two observational studies conducted in Israel and published in 2019 [8] and 2020 [9] identified a potential pharmacodynamic drug interaction between immunotherapy and cannabis. The first, a retrospective report, had a moderate sized sample and compared 89 patients that received the anti-programmed death-1 (PD-1) monoclonal antibody immunotherapy nivolumab to 51 that received nivolumab and cannabis [8]. Multivariate analyses determined that cannabis was associated with a decreased response rate to nivolumab with an Odds Ratio of 3.1. A Google Scholar search conducted 1/25/2024 found that this paper [8] has received 121 citations including by two clinical practice guidelines [10,11] (Table S1). However, despite the results section noting that “As shown in Table 2, no significant difference was found between the two groups in aspects of demographic and medical characteristics.” [8], an anonymous PubPeer posting in December of 2023 reanalyzed Table 2 using the same statistic and claimed to find four significant unreported baseline group differences, one-trend (p = .06) towards a baseline group difference in smoking (a well-established risk factor for a variety of cancers) [12], five mistakes calculating percentages and four rounding oversights [13]. A subsequent prospective observational report compared 68 immunotherapy patients with 34 immunotherapy and cannabis. Median overall survival was 6.4 months among cannabis users versus 28.5 months for cannabis non-users (p < .001) [9]. A Google Scholar search completed in 12/28/2023 determined that this study [9] has received 71 citations (Table S1). However, there have again been detailed anonymous PubPeer postings [14, 15] regarding both the original report [9] and the published correction [16]. These claim many difficulties repeating the non-parametric analyses, three errors in determining percentages, and 19 rounding issues [15]. The purported challenges in transparently calculating and interpreting the bi-variate analysis could challenge the conclusion that “no statistically significant differences were found in the baseline demographic and clinical variables between the study groups” [9].

Two research design issues in this area may warrant particular attention. First, observational studies with self-selected participants may have important confounds. Many studies have reported that marijuana users have an increased risk for of nicotine use [17]. The prospective study [9] did not contain any information on nicotine smoking and whether the groups were similar on this variable. It would be unfortunate to conclude that marijuana had negative health outcomes if these were driven by the well-established consequences of tobacco smoking [12]. Second, analysis of a 2 x 2 table for non-parametric variables is often completed with a chi-square test [18] although other statistics were developed for when there is a small N. Different practices have evolved since Karl Pearson developed the chi-square in 1900 including using Fisher’s exact test (henceforth Fisher’s, developed in 1930) or chi-square with Yates correction for continuity (henceforth Yates, developed in 1934 [19]) when the smallest cell has an observed value < 5 [20]. Yates has been criticized for over correcting and providing a p value that is too large (i.e. overly ‘conservative’) [18].The *British Medical Journal* (BMJ) offers more precise guidance [20] “In fourfold tables (i.e. a 2 x 2 contingency table), a *χ*^2^ test is inappropriate if the total of the table is less than 20, or if the total lies between 20 and 40 and the smallest expected (not observed) value is less than 5. … An alternative to the *χ*^2^ test is known as Fisher’s” (Supplemental Appendix 1). As immunotherapy is a first-line or cofirst line treatment for advanced non-small-cell lung cancer [21], metastatic colorectal cancer [22], advanced cutaneous melanoma [23], and advanced kidney cancer [24]) and medical cannabis is often used by cancer patients [25], the goal of this report was to reanalyze the data from these two reports [8, 9] including the correction [16].

## 2. Materials and Methods

This investigation involved an effort to verify the statistical analysis based on the information contained in the retrospective report [8], the prospective report [9], and the corresponding correction of Table 1 [16]. Most of the re-analyses were completed on the 2 x 2 nonparametric tests and determined whether using the reported N and same test would result in the same p-value. If the reported p value could be reproduced using the same statistic as was in the original [8, 9] methods, this was interpreted as verified. If the reported p value could only be reproduced using a different statistic as was listed in the methods, this was interpreted as misreported. If the p-value could not be reproduced with at least three-different statistics (chi-square, Fisher’s, or Yates), this was interpreted as unverified. Information about the smallest expected N and the total N was obtained to apply the BMJ guidance [20] on which non-parametric statistic was recommended. Additional analyses were also completed on whether the percentages were accurately reported using Microsoft Excel. Errors were categorized as: 1) general, defined as the reported and calculated differing by > 1%; 2) floor-rounding: defined as the reported being lower than the calculated; and 3) ceiling-rounding: defined as the reported being higher than the calculated. Nonparametric analyses were conducted with GraphPad Prism [26] with the smallest cell expected values determined with [27]. Pilot testing determined that 2x2 nonparametric analyses produced identical p-values for Prism, SPSS, and SAS. A between groups t-test was completed with the mean age, SD, and N provided [8] using GraphPad Prism [28].

## 3. Results

The original report provided sufficient information that an attempt could be made to verify Tables 1-4 [8], Tables 1-3 [9], and the corrected Table 1 [16].

### 3.1. Taha et al. *The Oncologist* 2019;24:549–554 [8]

The statistical analysis section noted that “Chi-square test was used to determine the difference between patients’ characteristics in both groups” and “Two-tailed p values of .05 were considered statistically significant” [8]. The original Table 1 [8] for THC ≥ 10 (yes or no) by progressive disease (yes or no) reported a chi-square p = .393. Reanalysis revealed a chi-square p = .2165. However, Fisher’s p = .3932. Similarly, CBD ≥ 1 (yes or no) by progressive disease reported a chi-square p = .116. Reanalysis showed a chi-square = .0885 but a Fisher’s p = .1161 indicating that both p values on Table 1 [8] were misreported.

Although not reported as significant in Table 2 [8], the immunotherapy + cannabis group were 5.7 years and significantly younger then the immunotherapy only group (*t*(138) = 3.137, p = .0021, Figure 1A). Smoking was 16.5% more frequent in the immunotherapy + cannabis (56.9%) than the immunotherapy only group (40.4%) but this difference was not quite significant (*χ*^2^(1) = 3.512, p = .0609, Figure 1B).

**Figure 1.**
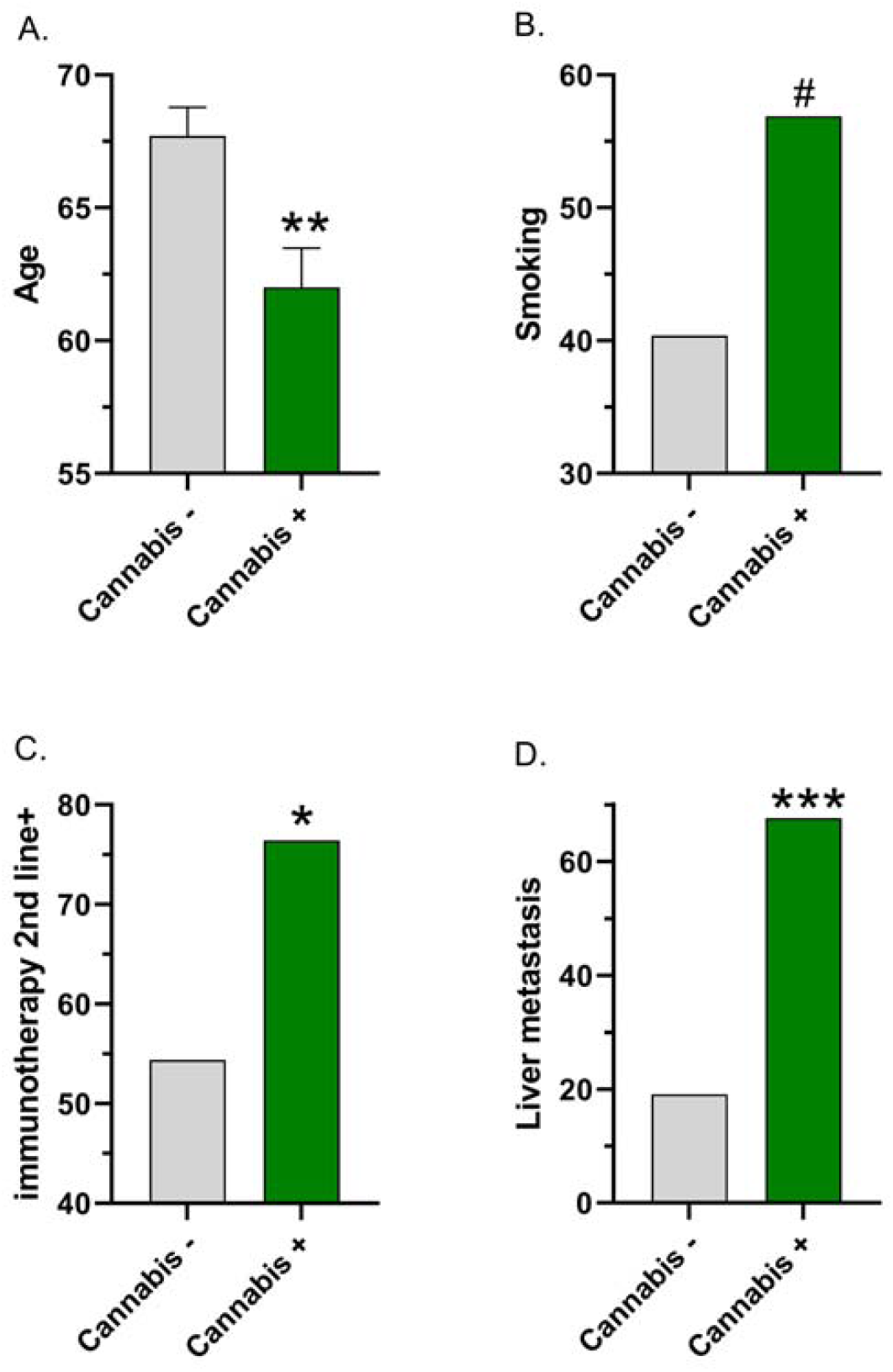
Baseline group differences between cannabis non-users (-) and users (+) based on re-analysis of the reported data in [8] (A, B) and [9] (C, D) ^#^chi-square p ≤ .061, ^*^chi-square p < .05, ^**^t-test p < .005, ^***^chi-square p < .0001.

**Table S1.**
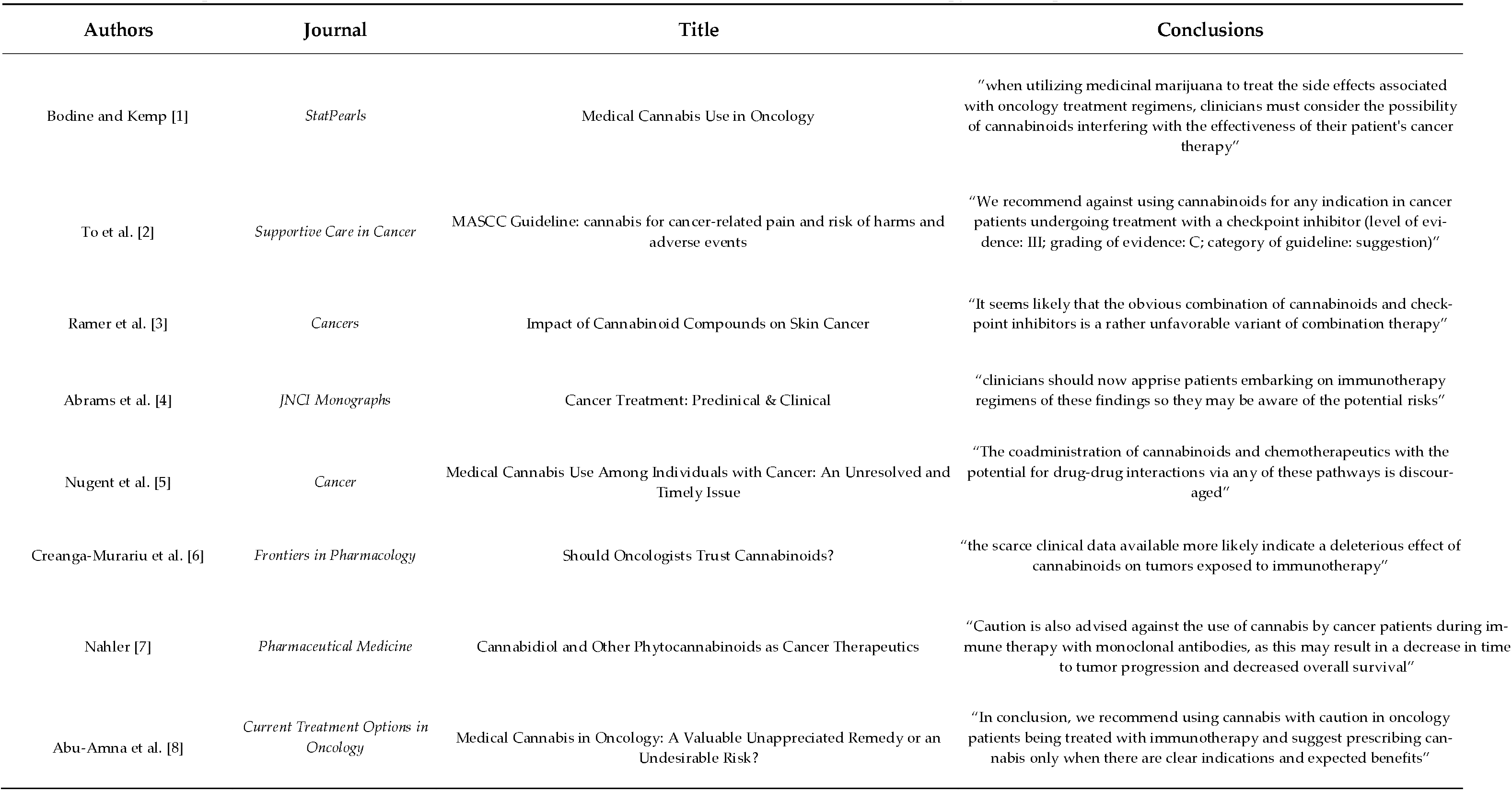
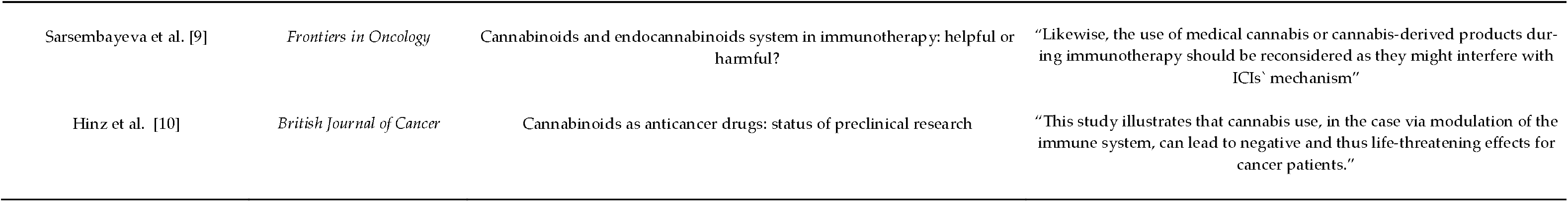
Examples of studies that have cited the Teha [8] and the Bar-Sela [9] cannabis and immunotherapy manuscripts.

| Authors | Journal | Title | Conclusions |
| --- | --- | --- | --- |
| Bodine and Kemp [1] | *StatPearls* | Medical Cannabis Use in Oncology | "when utilizing medicinal marijuana to treat the side effects associated with oncology treatment regimens, clinicians must consider the possibility of cannabinoids interfering with the effectiveness of their patient's cancer therapy" |
| To et al. [2] | *Supportive Care in Cancer* | MASCC Guideline: cannabis for cancer-related pain and risk of harms and adverse events | "We recommend against using cannabinoids for any indication in cancer patients undergoing treatment with a checkpoint inhibitor (level of evidence: III; grading of evidence: C; category of guideline: suggestion)" |
| Ramer et al. [3] | *Cancers* | Impact of Cannabinoid Compounds on Skin Cancer | "It seems likely that the obvious combination of cannabinoids and check-point inhibitors is a rather unfavorable variant of combination therapy" |
| Abrams et al. [4] | *JNCI Monographs* | Cancer Treatment: Preclinical & Clinical | "clinicians should now apprise patients embarking on immunotherapy regimens of these findings so they may be aware of the potential risks" |
| Nugent et al. [5] | *Cancer* | Medical Cannabis Use Among Individuals with Cancer: An Unresolved and Timely Issue | "The coadministration of cannabinoids and chemotherapeutics with the potential for drug-drug interactions via any of these pathways is discouraged" |
| Creanga-Murariu et al. [6] | *Frontiers in Pharmacology* | Should Oncologists Trust Cannabinoids? | "the scarce clinical data available more likely indicate a deleterious effect of cannabinoids on tumors exposed to immunotherapy" |
| Nahler [7] | *Pharmaceutical Medicine* | Cannabidiol and Other Phytocannabinoids as Cancer Therapeutics | "Caution is also advised against the use of cannabis by cancer patients during immune therapy with monoclonal antibodies, as this may result in a decrease in time to tumor progression and decreased overall survival" |
| Abu-Amna et al. [8] | *Current Treatment Options in Oncology* | Medical Cannabis in Oncology: A Valuable Unappreciated Remedy or an Undesirable Risk? | "In conclusion, we recommend using cannabis with caution in oncology patients being treated with immunotherapy and suggest prescribing cannabis only when there are clear indications and expected benefits" |
| Sarsembayeva et al. [9] | *Frontiers in Oncology* | Cannabinoids and endocannabinoids system in immunotherapy: helpful or harmful? | “Likewise, the use of medical cannabis or cannabis-derived products during immunotherapy should be reconsidered as they might interfere with ICIs’ mechanism” |
| Hinz et al. [10] | *British Journal of Cancer* | Cannabinoids as anticancer drugs: status of preclinical research | “This study illustrates that cannabis use, in the case via modulation of the immune system, can lead to negative and thus life-threatening effects for cancer patients.” |

### 3.2. Bar-Sela et al. *Cancers* 2020:12:2447 [9]

The statistical analysis section noted “A series of *χ*^2^ tests or Fisher’s exact tests … were conducted to analyze the differences between patients’ characteristics in both groups. … We computed 2-tailed p-values, where p < 0.05 was considered a statistically significant result” [9]. Table 1 [9] reported a non-significant p-value of .05178 for the non-parametric analysis (chi-square or Fisher’s) of whether immunotherapy was received as first line vs second+ line. However, recalculation determined that chi-square p = .0307 (Figure 1C) and Fisher’s p = .0334. The cannabis users (76.4%) were significantly more likely at baseline to receive immunotherapy as 2^nd^ line or later treatment than the cannabis non-users (54.4%). As the total N exceeded 40 and the minimum expected cell value was 13, the BMJ guidance [20] indicates that chi-square was the appropriate analysis (Table 1). This result was classified as misreported as the recalculated Yates p = .0518. Further information on Table 1 may be found in Section 3.3.

The results section [9] noted “liver metastasis of the immunotherapy group (I-G) (I-G 19%) vs. the immunotherapy-cannabis group (IC-G) (67%, p = 0.89)” which, based on this p-value, would be interpreted as non-significant. However, reanalysis revealed that this difference was significant (p < .0001 for chi-square, Fisher’s, and Yates, Figure 1D).

Table 2 [9] reported two trends in p-values of abnormal laboratory tests but both were unverified. For lymphocytes, this was reported as p = .08 but the calculated two-tailed chi-square was p = .1199, Fisher’s p = .1412, and Yates p = .1793. As the results section [9] mentioned a one-tailed p value, calculation of one-tailed values were as follows: chi-square p = .0600, Fisher’s p = .0890, and Yates p = .0896. Similarly, for Alkaline Phophatase, the reported p value (p = .09) could not be verified despite six attempts (two-tailed: chi-square = .1374, Fisher’s = .1468, Yates = .2157; one-tailed chi-square = .0687, Fisher’s = .1089, Yates = .1079). Further, of the twelve reported percentages, two (16.7%) contained minor rounding issues (12 / 68 reported as 17 but calculated as 17.6% which would round to 18, and 23 / 34 reported as 67 but calculated as 67.6% which would round to 68).

### 3.3. Correction to Bar-Sela et al. *Cancers* 2020:12:2447 [16]

The correction [16] published in April of 2022 to [9] contained a new Table 1. Of the 22 p-values reported from 2 x 2 analyses, 6 (27.3%) could not be replicated with chi-square or Fisher’s exact test (i.e. the non-parametric statistics listed in the methods) [9] or with chi-square with Yates correction. However, the p-value on 15 statistical tests did correspond to four decimal places with that of different statistic (Yates) than was listed in the methods. Finally, for renal cell carcinoma, the percentages (+ / total) were equal in both groups (5.9%) and the p-value (1.000) was the same for the chi-square, Yates, and Fisher’s tests (Table 1). Overall, 4.5% of the p-values from Table 1 [16] were verified. Table 1 also shows that four analyses had a minimum expected cell value < 5. However, as the total N (102) was well (> 2.5 fold) above 40, the BMJ guidance [21] indicates that chi-square would be the appropriate analysis.

Table 1 [16] did not list a p-value for the age comparison. However, the data was reported in a format (median, min – max) that precluded reanalysis of whether the cannabis users being three years younger was a significant difference. There were three errors in calculating percentages among cannabis users. Chronic diseases = 0 (13/34) was reported as 22.0% but calculated as 38.2%. High blood pressure (13/34) was reported as 34.1% but calculated as 38.2%. Brain metastasis (8/34) was reported as 13.2% but calculated as 23.5% (Table S2). The corrected table [16] contained 19 rounding errors, mostly floor rounding but there was one case of ceiling rounding of 0.1% (e.g. 55/68 reported as 80.8% but calculated as 80.9%, Table S2).

## Supporting information

Bar-Sela statistical concerns

Taha statistical concerns

---

**Table S2.** Percentages reported from [9] and calculated for Cannabis Users (CU) and Cannabis Non Users (CNU). ECOG: Eastern Conference Oncology Group.

| <u>Variable</u> | <u>Numerator</u> | <u>Denominator</u> | <u>Reported %</u> | <u>Calculated %</u> | <u>Difference</u> |
| --- | --- | --- | --- | --- | --- |
| Gender-Female (CU) | 10 | 34 | 29.5% | 29.4% | -0.1% |
| Gender-Male (CU) | 24 | 34 | 70.5% | 70.6% | +0.1% |
| ECOG ≤ 1 (CNU) | 55 | 68 | 80.8% | 80.9% | +0.1% |
| ECOG ≤ 1 (CU) | 24 | 34 | 70.5% | 70.6% | +0.1% |
| Chronic diseases = 0 (CNU) | 22 | 68 | 32.3% | 32.4% | +0.1% |
| Chronic diseases = 0 (CU) | 13 | 34 | 22.0% | 38.2% | +16.2% |
| Chronic diseases = 1 (CU) | 7 | 34 | 20.5% | 20.6% | +0.1% |
| Chronic diseases = 2 (CU) | 14 | 34 | 41.1% | 41.2% | +0.1% |
| Chronic heart disease (CNU) | 18 | 68 | 26.4% | 26.5% | +0.1% |
| High blood pressure (CU) | 13 | 34 | 34.1% | 38.2% | +4.1% |
| Melanoma (CNU) | 25 | 68 | 36.7% | 36.8% | +0.1% |
| Melanoma (CU) | 9 | 34 | 26.4% | 26.5% | +0.1% |
| Renal cell carcinoma (CNU) | 4 | 68 | 5.8% | 5.9% | +0.1% |
| Renal cell carcinoma (CU) | 2 | 34 | 5.8% | 5.9% | +0.1% |
| Brain metastasis (CU) | 8 | 34 | 13.2% | 23.5% | +10.3% |
| Lungs metastasis (CNU) | 39 | 68 | 57.3% | 57.4% | +0.1% |
| Liver metastasis (CU) | 11 | 34 | 32.3% | 32.4% | +0.1% |
| Immunotherapy 1<sup>st</sup> line (CNU) | 31 | 68 | 45.5% | 45.6% | +0.1% |
| Immunotherapy 2<sup>nd</sup> line (CU) | 26 | 34 | 76.4% | 76.5% | +0.1% |
| Pembrolizumab or nivolumab (CU) | 29 | 34 | 85.2% | 85.3% | +0.1% |
| Ipilimumab or nivolumab (CU) | 4 | 34 | 11.7% | 11.8% | +0.1% |
| Durvalumab or atezolizumab (CNU) | 5 | 68 | 7.3% | 7.4% | +0.1% |

## Supplementary Appendix 1

British Medical Journal guidance [20] on the selection of non-parametric statistics for small N.

“When the numbers in a 2 x 2 contingency table are small, the χ2 approximation becomes poor. The following recommendations may be regarded as a sound guide. In fourfold tables a χ2 test is inappropriate if the total of the table is less than 20, or if the total lies between 20 and 40 and the smallest expected (not observed) value is less than 5; in contingency tables with more than one degree of freedom it is inappropriate if more than about one fifth of the cells have expected values less than 5 or any cell an expected value of less than 1. An alternative to the χ2 test for fourfold tables is known as Fisher’s Exact test and is described in Chapter 9

When the values in a fourfold table are fairly small a “correction for continuity” known as the “Yates’ correction” may be applied. Although there is no precise rule defining the circumstances in which to use Yates’ correction, a common practice is to incorporate it into χ2 calculations on tables with a total of under 100 or with any cell containing a value less than 10.”

## 4. Discussion

The observational studies [8, 9] identifying a potential drug interaction between cannabis and immunotherapy have been widely cited including in clinical practice guidelines [10, 11] and in a publication geared to the general public [2] (Table S1). This report determined that an appreciable subset of the statistics contained in [8, 9] could not be verified and was generally consistent with the PubPeer reports [13-15]. There are a few possibilities that might be able to account for only 4.5% of the p-values in the correction [4] being verified and the many inconsistencies. Analyses in the retrospective report were completed with SPSS, version 21 [8]. The prospective study listed both SAS and R [9]. Although we used GraphPad for our analyses, we feel that it is unlikely that different software would produce such disparate results. Another possibility is that research with humans is challenging and there was a small amount of missing data. We feel this explanation is also unlikely because the correction listed the N for each cell for four variables [9] and there was no missing data. Similarly, verification of the percentages (Table S3) does not indicate that the denominator was decreased. However, we can not dis-count this possibility for other variables or tables. Another possibility, which we feel is more likely because the p-values correspond to four decimal places on fifteen occasions (Table 1) is that the methods section listed one statistic (chi-square) but a slightly different statistic (chi-square with Yates) was completed. The PubPeer response by a middle author also indicates that Yates was completed [29]. Notably, the BMJ guidance [20] attempted to clarify when to use chi-square, Fisher’s, and Yates. Table 1 suggests that Yates may have been uniformly used, independent of the minimum expected value or the total N. Although the methods section listed “We computed 2-tailed p-values”, as the attempted verification produced p-values that were approximately twice as large (and the results section alluded to a one-tailed test), it is also possible that the reported p-values were one-tailed on Table 2 [4]. Similarly, while the methods section [8] lists “Chi-square test was used to determine the difference between patients’ characteristics in both groups.”, it is likely that some of the reported p-values were from Fisher’s. It is important for the readers to be informed of which nonparametric statistic was completed so that they can assess the Type I error rate. There is a general consensus that Yates is overly cautious in its desire to avoid a type I error [30]. Fisher’s should not be reported if the total N of all four cells is above 40 (the N was 140 in [Teha]) and the expected N was ≥ 5 [20]. Although choosing which non-parametric statistic to use can be challenging when different resources have contradictory recommendations [31, Supplemental Appendix 1], at the very least, we believe that telling that audience in the methods that you will run statistic A but then reporting in the results the findings from statistic B is a non-trivial oversight.

Smoking is a well-established factor for the development of different cancers [12] which should not be overlooked [9]. In colorectal cancer (CRC), smoking increases the risk by 59% (OR = 1.59, 95% CI: 1.30–1.94) [31]. Additionally, over 29 pack years of smoking was linked to 61% increased CRC risk (OR = 1.61, 95% CI: 1.31–1.99) compared to those who never smoked [32]. Similarly, smokers carry a high risk of developing early-onset colorectal neoplasms (EoCRN) (OR, 1.33; 95% confidence interval [CI], 1.17–1.52) compared to those who never smoked [33]. Another study analyzed 13,169 cases and 16,010 controls from Europe and Canada and found that male smokers with an average daily dose of >30 cigarettes had ORs of 103.5 (95% CI 74.8-143.2) for squamous cell carcinoma (SqCC), 111.3 (95% CI 69.8-177.5) for small cell lung cancer (SCLC), and 21.9 (95% CI 16.6-29.0) for adenocarcinoma (AdCa). In women, the corresponding ORs were 62.7 (95% CI 31.5-124.6), 108.6 (95% CI 50.7-232.8), and 16.8 (95% CI 9.2-30.6), respectively. [34] Interestingly, both inherent and acquired attributes of a person such as sex or medical conditions can also affect the degree of cancer development from smoking. In one study, patients with hyperglycemia and smoked ≥20 pack-years had a synergistic effect on the gallbladder cancer (GBC) risk (all P < 0.01) [35]. Additionally, those with diabetes and smoked ≥20 pack-years had the highest risk of GBC compared to those with normoglycemia and never smoked (hazard ratio (HR), 1.658; 95% CI, 1.437–1.914) [36]. Another study using multivariate Cox regression models suggested that male smokers had a 39% higher risk (HR = 1.39, 95% confidence interval (CI): 1.16, 1.67) of cancer of the left (distal or descending) colon but not of the right (proximal or ascending) colon (HR = 1.03, 95% CI: 0.89, 1.18), while female ever smokers had a 20% higher risk (HR = 1.20, 95% CI: 1.06, 1.36) of cancer of the right colon but not of the left colon (HR = 0.96, 95% CI: 0.80, 1.15) [35]. Compared with male smokers, female smokers also had a greater risk of rectal cancer (P for heterogeneity = 0.03) [35].

Although there were many differences between our findings are those reported earlier [8, 9, 16], we are not suggesting that anyone has engaged in anything nefarious. We commend the earlier reports [8, 9] for addressing a timely and important topic. However, it may also be valuable to prevent situations like this in the future. If the authors are going to do engage in an atypical practice (e.g. floor rounding [14]), it would be beneficial to briefly document this in the methods section. It is also possible that floor rounding was employed for reporting the p-values. About one-third of leading biomedical journals reported rarely or never using specialized statistical reviews [36]. As the statistical analysis section contained “A series of *χ* ^2^tests or Fisher’s exact tests (when the assumptions of the parametric *χ* ^2^ test (sic) were not met)” [9], if there was a statistical reviewer, this person may not have been reading carefully. Emails to the editors asking if these manuscripts [8, 9] received a statistical review were not answered. Further, it would be well beyond standard practice to expect an (often volunteer) reviewer to re-run all the analyses. In psychology, it is common to list the test statistic, the degrees of freedom and the p-value. An analysis of a quarter million psychology papers revealed that half contained inconsistencies where the reported test statistic and p-value did not correspond. Further, one-seventh of papers included gross inconsistences defined as “the reported p-value was significant and the computed p-value was not, or vice versa” [37]. Some experimental psychology journals use software to identify statistical irregularities [37]. We are not aware of any biomedical journals which are employing similar software to assess percentage calculations including rounding errors or non-parametric analyses.

Age was not reported as statistically significant in [8] but had a significant difference (p < .002) in our analysis (Figure 1A). This age difference could reflect that cannabis is psychoactive and providers in Israel may be less likely to prescribe cannabis to older patients [38]. Alternatively, cannabis users could have had a more aggressive or more advanced form of cancer at baseline. As the cannabis users were much more likely to have liver metastasis at baseline (Figure 1D), cannabis may simply be a proxy for a high burden symptomatic disease [38]. Whether immunotherapy was received first or second-line was reported as non-significant (p > .050) in Table 1 [8]) and again in [9]. However, re-analysis revealed that this baseline difference was significant (Figure 1C). Both of these findings could be interpreted as gross inaccuracies. In one high-profile instance of a manuscript with multiple issues, there was a twelve-year interval between the original MMR vaccine and autism study [39] and the retraction [40]. Journals, including those with reasonable open-access fees (e.g. *Cancers* is currently 2,900 CHF [41], *The Oncologist* is currently $2,500 [42]) need to have sufficient staff to deal with these concerns in a timely fashion. A practice that might be helpful moving forward is to include the full data and a detailed data dictionary as a supplementary material, either with the published manuscript or with a preprint, to allow for a second party to verify all analyses. The author instructions of *Cancers* includes *“*We encourage all authors of articles published in MDPI journals to share their research data.” [26]. Similarly, the author guidelines for the retrospective journal notes “*The Oncologist* strongly encourages authors to make all data and software code on which the conclusions of the paper rely available to readers.” An email to the corresponding author requesting the original data was not responded to. It would also be informative to list the N for all cells in each analysis in a supplementary appendix to facilitate statistical verification.

Basic science investigations are crucial as they can avoid the many potential confounds (e.g. age, disease severity, smoking, Figure 1) that are challenging to overcome with observational reports. A recent report attempted to replicate and extend [8, 9] in two ways. First, tetrahydrocannabinol did not impact the enhanced survivability to anti-programmed death ligand 1 antibody treatments in a murine colorectal model. Second, although advanced non-small cell lung cancer patients using cannabis (N = 102) were significantly younger, more likely to have brain metastasis, and marginally more likely to have liver metastasis (p = .06) at baseline than those that did not (N = 99), cannabis did not significantly impact the survivability following pembrolizumab [39].

In balancing the benefits and harms of cannabis for cancer patients [1, 2, 39], a broader perspective is useful. There has been appreciable work focused on anti-cancer activity of cannabinoids, including possible use in potentiating immunotherapy [46-50]. Cannabonoids, including cannabigerol (CBG), cannabidiol (CBD), and tetrahydrocannabinol, have been shown, in vitro and in vivo, to inhibit cancer cell proliferation and metastasis, while also promoting apoptosis and suppressing cancer-related angiogenesis [47-49]. Cannabinoids may also play a positive role in regulating aberrant cellular metabolism characteristic of cancer [50]. With respect to the role of cannabinoids in potentially potentiating immunotherapy in colorectal cancer, these agents may boost immunogenicity through cytotoxic effects on cancer cells and the subsequent release of antigens, as well as reprograming immune cells to target tumors [49]. Given the various mechanisms by which cannabinoids may have anti-cancer effects, it is important not to dismiss potential benefits based on possibly unverifiable statistical interpretation of data. In addition, the Taha [8] and Bar-Sela [9, 16] studies involved a limited number of cancer types, which should not be considered universally applicable for cancer immunotherapy in general. It should also be noted that effects of cannabinoids on immunotherapy may not only be influenced by cancer type but by the concentration of the relevant agents. For example, while higher concentrations of THC had immunosuppressive effects in vitro and in vivo, lower concentrations were immunostimulatory [26].

All of these facts recommend caution in the interpretation of the Taha [8] and Bar-Sela [9, 16] studies as well as other naturalistic investigations where the cannabis route of administration and THC concentration are not homogenous.

One limitation of this report as that only a subset of the originally reported findings could be assessed for veracity because the full raw data was unavailable. Therefore, we can not infer whether the figures contain verifiable, misreported, or unverifiable information. As noted above, we can not discount the possibility that the Israeli team [29] also used floor rounding to report their p-values (e.g. p = .0896 reported as .08) so some analysis that were classified as unverified may in fact be misreported instead. There are many statisticians that would argue that it is ridiculous to interpret the results of a study with p = .055 differently than p = .045. Dividing results of hypothesis tests into ‘significant’ and ‘non-significant’ is both unhelpful and outdated [51]. Although we would concur with the thrust of this argument, findings like Table 1 [8] where age is described as non-significant (i.e. p > .05) but then has a recalculated p-value of .0021 or the liver metastasis which was reported as “p = .89” but was calculated as p ≤ .0001 are more concerning as they obscure the reader’s ability to accurately determine whether the cannabis users and non-user groups were equivalent at baseline.

## 5. Conclusions

In conclusion, reanalysis of a subset of the reported information in the very influential observational reports purporting to show a harm inducing drug interaction between immunotherapy and cannabis [8, 9, 15], was unable to verify many of the analyses and there were some gross inaccuracies. This reanalysis and Figure 1 calls into question the conclusion that “no statistically significant differences were found in the baseline demographic and clinical variables between the study groups” [9]. Future prospective studies on this topic could consider matching on key demographic or medical condition variables. Due to the well-known contribution of smoking to cancer in a variety of tissues [12], excluding participants who are current or former nicotine smokers, or at least stratifying a homogenous sample with one cancer type accordingly, might be informative. Perhaps more important would be strengthening practices [51] to prevent similar situations [52, 53] from occurring. This could include registering the methods, making the raw data publicly available, and developing and implementing statistical software that could assist manuscript reviewers in identifying irregularities in the reporting of descriptive and bivariate analyses.

## 6. Patents

Not applicable.

## Supplementary Materials

The following supporting information can be downloaded at: www.mdpi.com/xxx/s1, Table S1, S2, and Supplementary Appendix.

## Author Contributions

Conceptualization, B.J.P.; methodology, B.J.P.; software, M.T.; validation, X.X., Y.Y. and Z.Z.; formal analysis, B.J.P.; investigation, B.J.P, M.T.; resources, X.X.; data curation, X.X.; writing—original draft preparation, B.J.P., C.J.C., A.H, J.G., M.T., & M.B.; writing—review and editing, B.J.P. C.J.C, & M.B.; visualization, B.J.P, M.T.; supervision, X.X.; project administration, X.X.; funding acquisition, B.J.P. All authors have read and agreed to the published version of the manuscript.

## Institutional Review Board Statement

Not applicable.

## Informed Consent Statement

Not applicable.

## Data Availability Statement

All of the information used in this manuscript is publicly available at [8. 9, 16].

## Acknowledgments

Thanks to Amy Allison, MLS for technical support, Christina M. Gregor and Alysha Lopez, PharmD for feedback on an earlier version of this manuscript, and Mellar Davis, MD for his deep insights on this topic.

