## Supplementary material for "Immunotherapy and Cannabis: A Harmful Drug Interaction or Reefer Madness?": Bar-Sela statistical concerns

### Summary of Table 1

- Appropriate analysis of Table 1 is crucial for any RCT or retrospective study. Unfortunately, this "correction" should be viewed with substantial skepticism.
- Of the 22 p values that were listed, 16 could **not** be replicated using the N provided and using the statistics listed in the methods. 6 of the 22 p values could also not be verified using the N reported and three different nonparametric statistics (chi-square, Fisher's, and chi-square with Yates correction)
- The authors appear to have inappropriately reported the p value for another statistic than was listed in their methods. As the Yates correction is typically only run with N per cell of < 5[1] (see also BMJ guidance), the preponderance of the information in this "correction" is factually incorrect. This statistic was misapplied in many situations where the N was sufficiently large.
- It also shows exceedingly poor attention to detail that percentages appear to be calculated incorrectly three times and there were 19 rounding errors on this "corrected" table.

cancers MDPI

*Correction*

**Correction: Bar-Sela et al. Cannabis Consumption Used by Cancer Patients during Immunotherapy Correlates with Poor Clinical Outcome. *Cancers* 2020, *12*, 2447**

Gil Bar-Sela [1,2,*], Idan Cohen [1], Salvatore Campisi-Pinto [3], Gil M. Lewitus [3], Lanuel Oz-Ari [2], Ayellet Jehassi [4], Avivit Peer [5], Ilit Turgeman [5], Olga Vernicova [1], Paula Berman [3], Mira Wollner [5], Mor Moskovitz [5] and David Meiri [3,*]

1. See Wikipedia or most standard statistics textbooks

4.4 Statistical analysis (from Bar-Sala et al. *Cancers* 2020; 12:2447)

A series of $\chi^2$ tests or Fisher's exact tests (when the assumptions of the parametric $\chi^2$ test were not met) and nonparametric Mann–Whitney U tests were conducted to analyze the differences between patients' characteristics in both groups. Time to tumor progression (TTP) and overall survival (OS) was estimated using the Kaplan-Meier survival curve by group, and the log-rank test was computed to differentiate the survival curves between groups. Hazard ratios and the corresponding 95% CIs based on a Cox proportional hazards regression model were provided for multivariate analyses. We computed 2-tailed $p$-values, where $p < 0.05$ was considered a statistically significant result. Statistical analyses were performed using the SAS software package version 9.4 (SAS Institute, Cary, NC, USA).

Note that the methods section does **not** mention running chi-square with Yates correction. According to standard convention, this would be completed when the N for a cell was < 5.
https://en.wikipedia.org/wiki/Yates%27s_correction_for_continuity

I attempted to replicate the nonparametric statistics using:
https://www.graphpad.com/quickcalcs/contingency2/
**Bold** p-values on the next page (right 3 columns) correspond with those reported in the "corrected" table

### BMJ's guidance on small N

#### Small numbers

When the numbers in a 2 x 2 contingency table are small, the $\chi^2$ approximation becomes poor. The following recommendations may be regarded as a sound guide. (2) In fourfold tables a $\chi^2$ test is inappropriate if the total of the table is less than 20, or if the total lies between 20 and 40 and the smallest expected (not observed) value is less than 5; in contingency tables with more than one degree of freedom it is inappropriate if more than about one fifth of the cells have expected values less than 5 or any cell an expected value of less than 1. An alternative to the $\chi^2$ test for fourfold tables is known as Fisher's Exact test and is described in Chapter 9

Here is a description from the *British Medical Journal (*BMJ*)* on when the N expected per cell is so small as to be concerning. They note a total of all 4 cells should be between 20 and 40. This study has a total N of 68 + 34 = 102 so alternatives to chi-square would generally **not** be warranted.  My reading of the above is that when more than one-fifth of cells (i.e. two of four) have expected (not observed) values of < 5, then Fisher's should be run. The corrected table contains observed (instead of expected) values but the 2x2 analyses for the data in K, M & O (label on 1st column of page 5) would be situation where Fisher's would be warranted.

https://www.bmj.com/about-bmj/resources-readers/publications/statistics-square-one/8-chi-squared-tests

### < 10 guidance for use of Yates

When the values in a fourfold table are fairly small a "correction for continuity" known as the "Yates' correction" may be applied (3). Although there is no precise rule defining the circumstances in which to use Yates' correction, a common practice is to incorporate it into $\chi^2$ calculations on tables with a total of under 100 or with any cell containing a value less than 10. The $\chi^2$ test on a fourfold table is then modified as follows:

As will be seen, it does not appear that Fisher's was ever reported. Yates correction would be applied when the value of a single cell is between 0 and 9. This would apply to 2x2 tables from D, F, G, I, J, K, M, N, O, P, S, U, V.

Note that, even if the authors did run Yates, the circumstances of when they ran a chi-square and when they ran Yates should have been clearly but briefly described in the methods. Unfortunately, they were not.

**Table 1.** Demographics and medical conditions. (page 1)

| | Characteristics | Cannabis Non-Users N = 68 | | Cannabis Users N = 34 | | p-Value | Chi-sq | Fishers | Chi-sq Yates |
|---|---|---|---|---|---|---|---|---|---|
| | Age in years median (range) | 69 (18–92) | | 66 (37–85) | | Why no t p-value? | | | |
| | Gender—N (%) | | | | | | | | |
| | Female | 22 (32.4) | 32.4 | 10 (29.5) | 29.4 | | | | |
| A | Male | 46 (67.6) | 67.6 | 24 (70.5) | 70.6 | 0.9399 | .7628 | .8240 | **.9399** |
| B | Performance Status Eastern Cooperative Oncology Group (ECOG)—N (%) | | | | | | | | |
| | 1≤ | 55 (80.8) | 80.9 | 24 (70.5) | 70.6 | 0.3568 | | | |
| | ≥2 | 13 (19.1) | 19.1 | 10 (29.4) | 29.4 | 0.3568 | .2409 | .3152 | **.3568** |
| | Chronic diseases per patient—N (%) | | | | | | | | |
| C | 0 | 22 (32.3) | 32.4 | 13 (22.0) | 38.2 | 0.7124 | .4908 | .5136 | .6398 |
| D | 1 | 16 (23.5) | **23.5** | 7 (20.5) | 20.6 | 0.9332 | .7376 | .8062 | **.9332** |
| E | 2 or more | 30 (44.1) | **44.1** | 14 (41.1) | 41.2 | 0.9437 | .7774 | .8341 | **.9437** |
| | Background diseases—N (%) | | | | | | | | |
| F | Chronic heart disease | 18 (26.4) | 26.5 | 5 (14.7) | 14.7 | 0.2762 | .1802 | .2162 | **.2762** |
| G | Diabetes | 17 (25.0) | 25.0 | 6 (17.6) | 17.6 | 0.5576 | .4022 | .4606 | **.5576** |
| H | High blood pressure | 34 (50.0) | 50.0 | 13 (34.1) | 38.2 | 0.3612 | .2611 | .2971 | **.3612** |
| I | Chronic obstructive pulmonary disease (COPD) | 9 (13.2) | 13.2 | 3 (8.8) | 8.8 | 1 | .5145 | .7463 | .7445 |
| J | Hyperlipidemia | 23 (33.8) | 33.8 | 7 (20.5) | 20.6 | 0.2491 | .1436 | .1737 | .2174 |
| K | Other | 2 (2.9) | 2.9 | 0 (0.0) | 0.0 | 1 | .3125 | .5512 | .8007 |
| | Type of malignancy—N (%) | | | | | | | | |
| L | Non-small cell lung cancer | 37 (54.4) | 54.4 | 20 (58.8) | 58.8 | 0.8325 | .6723 | .8327 | **.8325** |
| M | Melanoma | 25 (36.7) | 36.8 | 9 (26.4) | 26.5 | 0.414 | .2985 | .3750 | **.4140** |
| N | Renal cell carcinoma | 4 (5.8) | 5.9 | 2 (5.8) | 5.9 | 1 | **1.000** | **1.000** | **1.000** |
| O | Other | 2 (2.9) | 2.9 | 3 (8.8) | 8.8 | 1 | .1946 | .3300 | .4175 |
| | Main site of metastasis—N (%) | | | | | | | | |
| P | Brain | 12 (17.6) | 17.6 | 8 (13.2) | 23.5 | 0.6593 | .4806 | .5976 | **.6593** |
| Q | Lungs | 39 (57.3) | 57.4 | 23 (67.6) | 67.6 | 0.4303 | .3155 | .3914 | **.4303** |
| R | Liver | 13 (19.1) | 19.1 | 11 (32.3) | 32.4 | 0.2157 | .1374 | .1468 | **.2157** |

Green = correctly reported %, Red = rounding error, Yellow highlights = percentage reporting error

**Bold** matches reported p-value
red = incorrect test, green = correct test

### Summary of Page 1 of Table 1

Of 18 provided p values, only 1 could be verified using the statistics specified in the methods section.

Of the 18 provided p values, 12 had p values that could be replicated but only by using a **different statistic** than was described in the methods section (chi-square with Yates correction).

Of the 18 provided p values, 5 could not be verified using either chi-sq or Fisher's (i.e. the statistics in the methods) or with chi-square with Yates correction.

There were 3 percentages whose calculations were incorrectly reported by at least 4.0% (Min = 4.1%, Max = 13.2%)

There were 14 errors of rounding (difference always = 0.1%).

A minor omission is that it would be more conventional to list a p-value (e.g. t-test) for the group comparison for age.

**Table 1.** *Cont.* (page 2)

| | Characteristics | Cannabis Non-Users N = 68 | | Cannabis Users N = 34 | | *p*-Value | P values: Chi-sq | P values: Fisher's | P values: Chi-sq w/Yates |
|---|---|---|---|---|---|---|---|---|---|
| | Immunotherapy given as—N (%) | | | | | | | | |
| S | First line of treatment | 31 (45.5) | 45.6 | 8 (23.5) | 23.5 | 0.05178 | .0307 | .0334 | .0518 |
| | Second line of treatment or more | 37 (54.4) | 54.4 | 26 (76.4) | 76.5 | 0.05178 | | | |
| | Checkpoint therapy—N (%) | | | | | | | | |
| T | Anti PD1: Pembrolizumab or Nivolumab | 47 (69.1) | 69.1 | 29 (85.2) | 85.3 | 0.127 | .0772 | .0945 | .1270 |
| U | Ipilimumab and Nivolumab | 16 (23.5) | 23.5 | 4 (11.7) | 11.8 | 0.2517 | .1583 | .1934 | .2517 |
| V | Anti PDL-1: Durvalumab or Atezolizumab | 5 (7.3) | 7.4 | 1 (2.9) | 2.9 | 1 | ..3720 | .6607 | .6554 |

Key: green = % calculated (rounded) correctly
red = % calculated (rounding) incorrectly

P values calculated with GraphPad Prism. Although it is theoretically possible that the differences are due to software (Bel-Sala used SAS), we think this is exceedingly unlikely for simple non-parametric statistic like this. It is much more likely (based on values corresponding to 4 decimal places) that the authors simply ran a different statistic than was reported. Unfortunately, the statistic that was run (chi-square with Yates correction) is (again) only appropriate with a cell N of < 5.

**Summary of Page 2 of Table 2**

Table 1 tests for the equivalence between groups which is a core component of any observational study.

Importantly, the results section does **not** note that the two groups were statistically different at baseline regarding whether immunotherapy was first or second line. This is a major oversight that substantially impacts the interpretation of this study.

Of the 4 statistics on this page, only 1 was "correct".

2 statistics could not be verified using the statistics specified in the methods. However, the p-value could be obtained using a different statistic (chi-square with Yates correction) than was in their methods for 3 p values.

1 statistic could not be verified.

Although minor, of the 10 percentages that were listed, 5 had rounding errors.

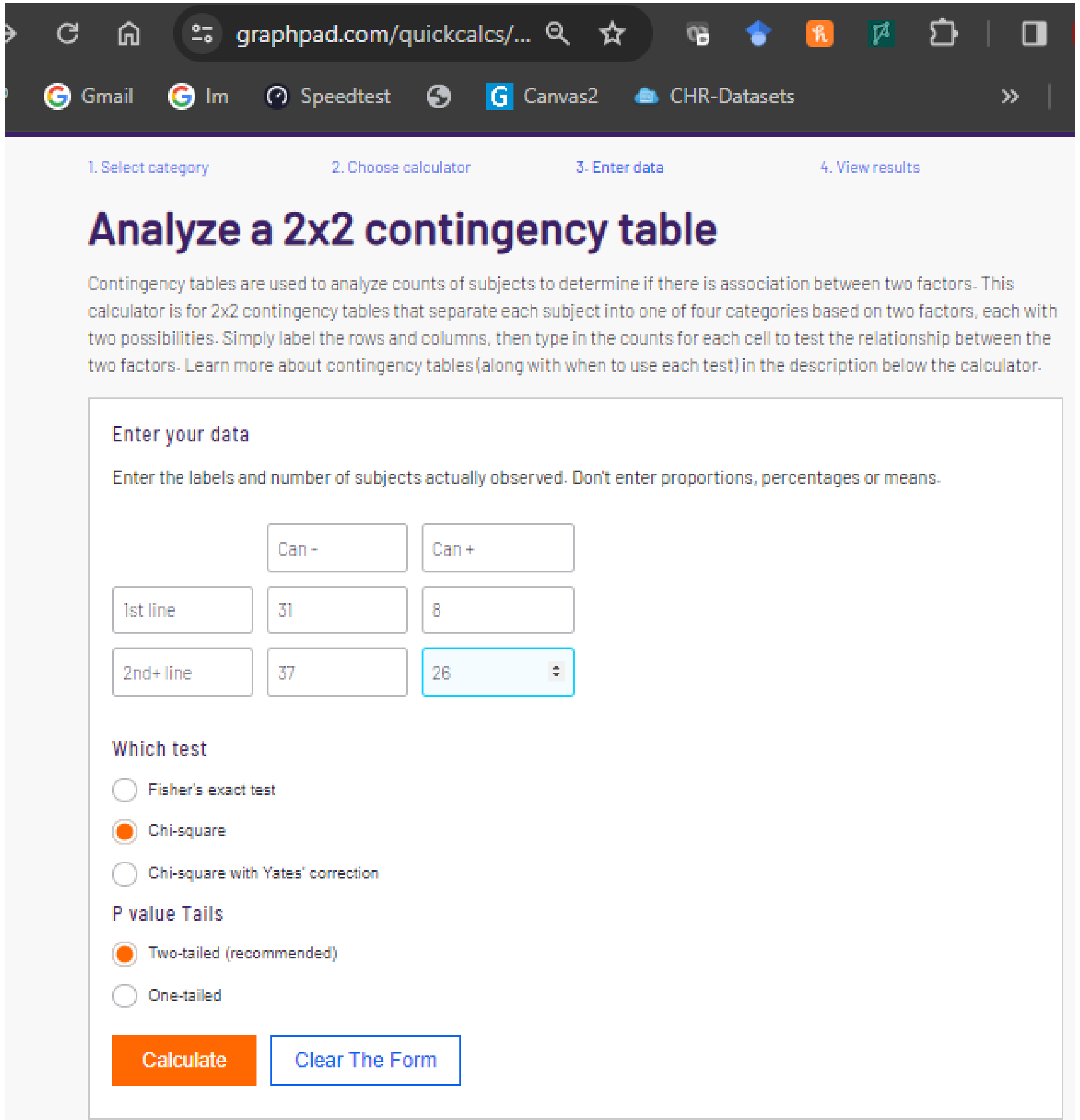


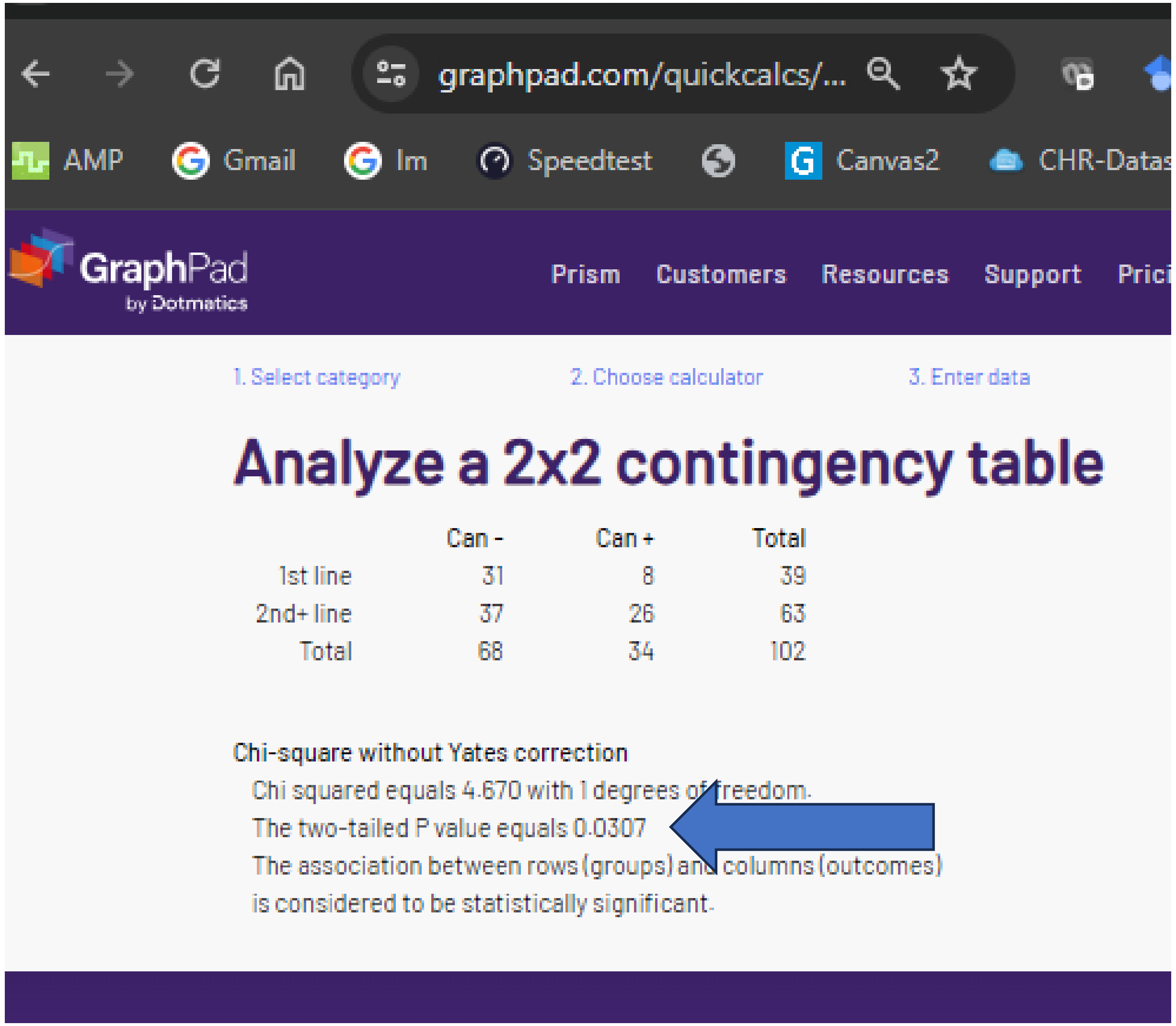


Here are the screen captures of the immunotherapy as first vs second line analysis.
The “Correction” reports the p-value as .05178 (i.e. non-significant). GraphPad show a p-value of .0317 (i.e. significant).

### A. “corrected” chi-sq or Fishers p = .9399

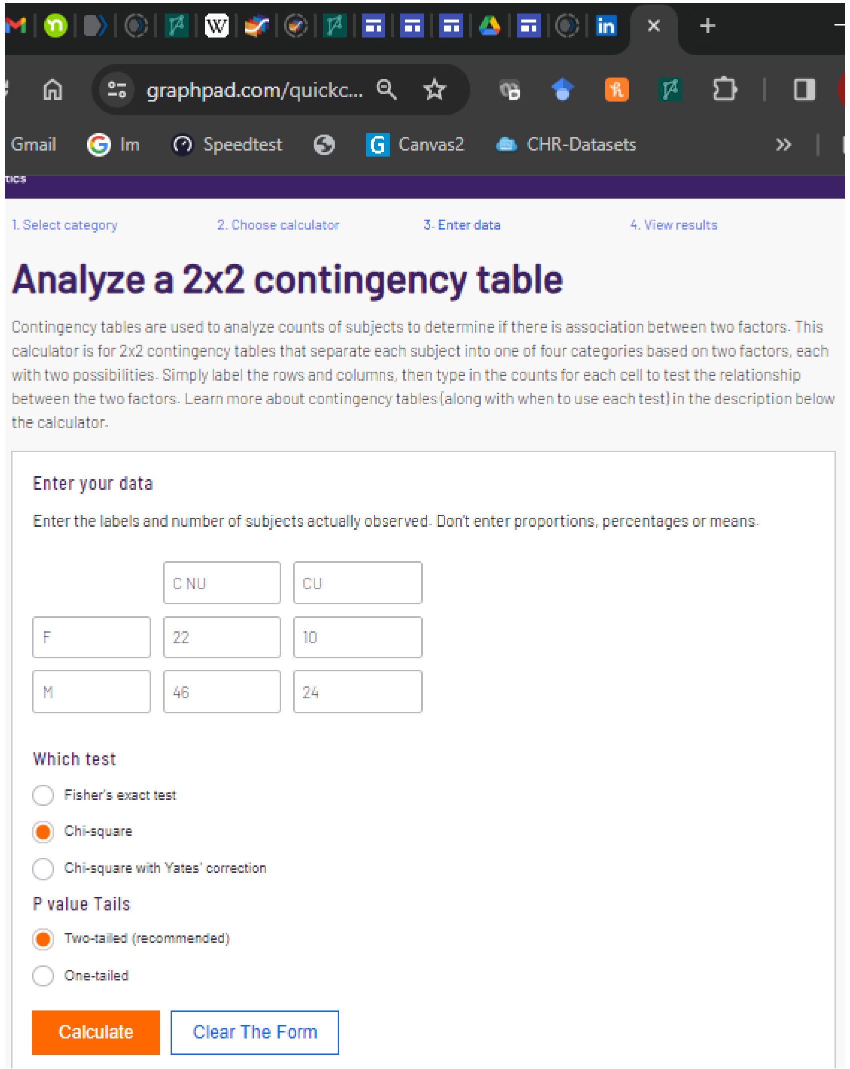


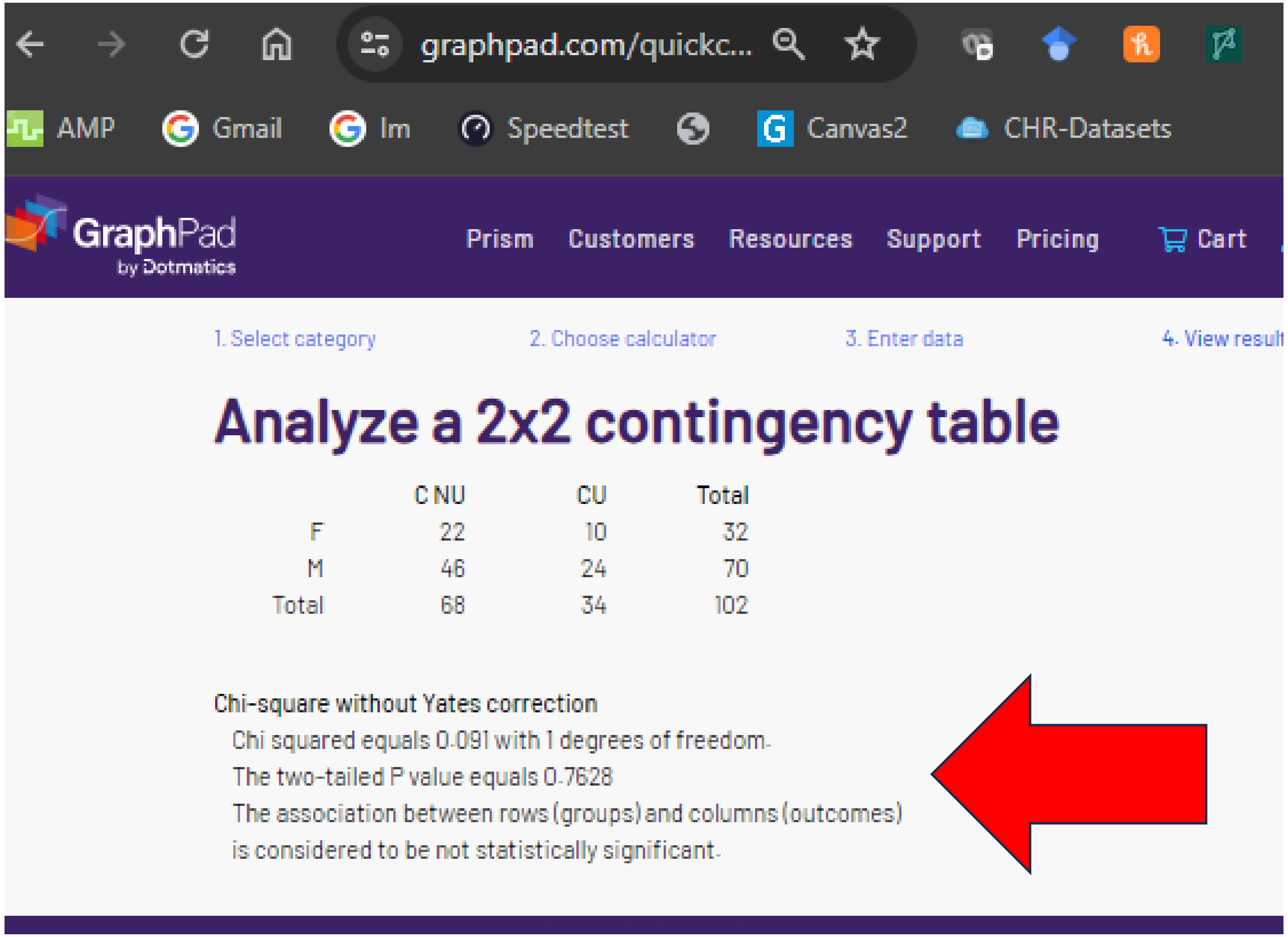

### A. “corrected” chi-sq or Fishers p = .9399

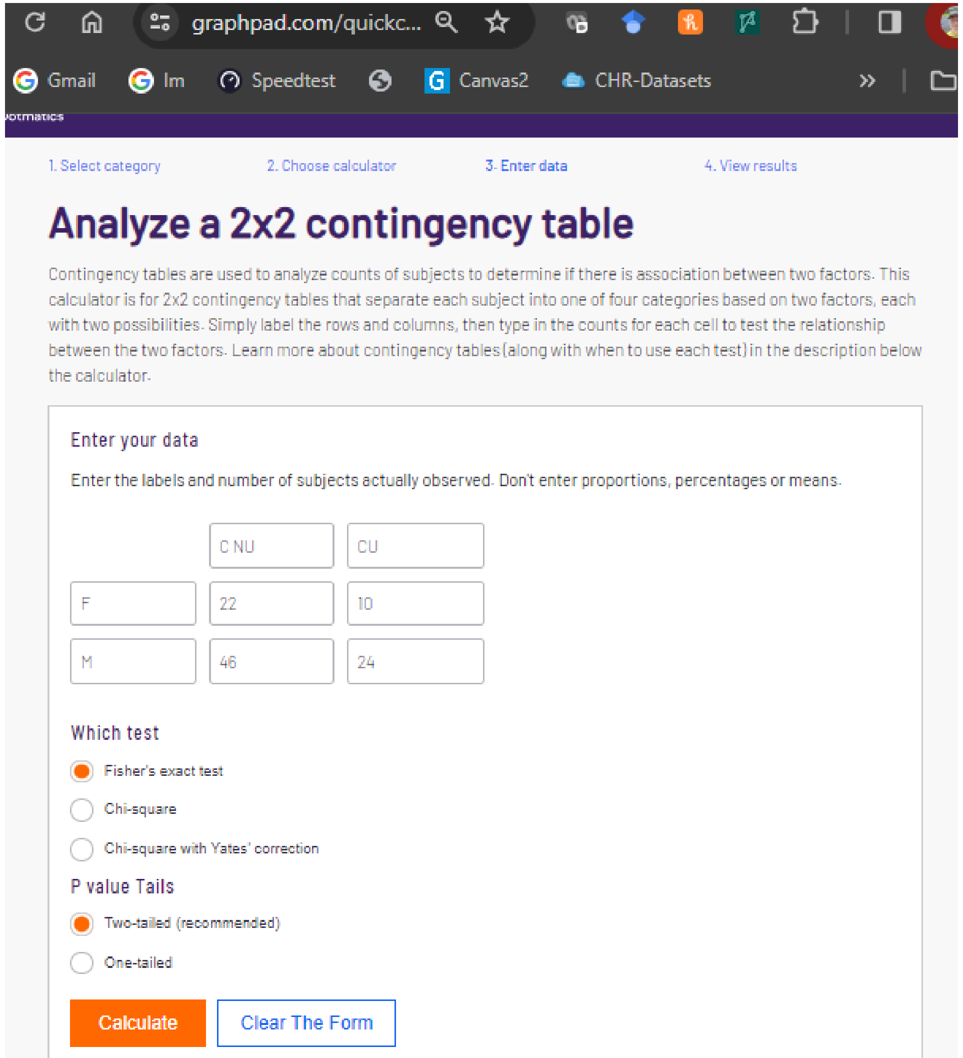


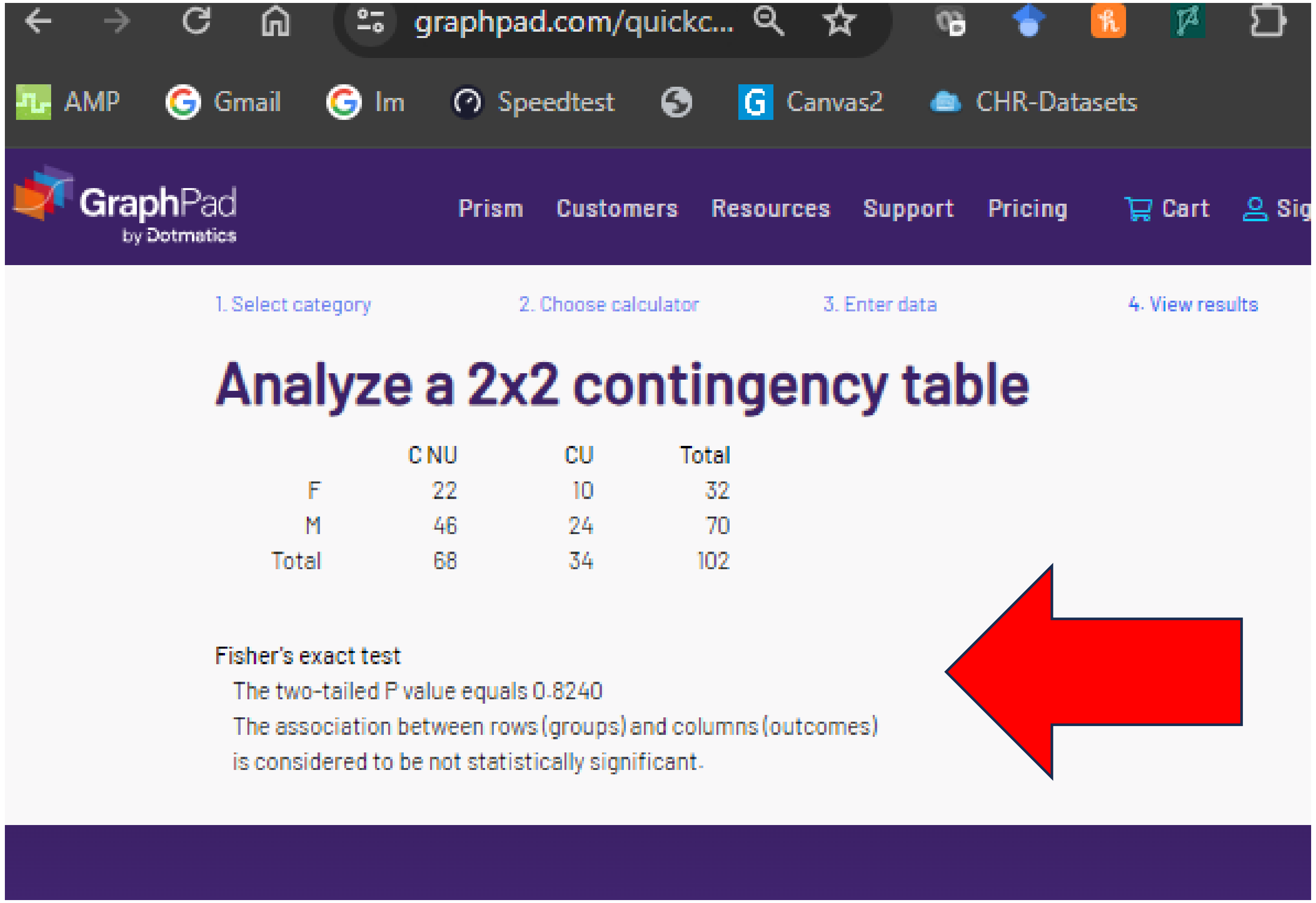


The “corrected” p value could not be verified for gender.

### A. "corrected" chi-sq or Fishers p = .9399

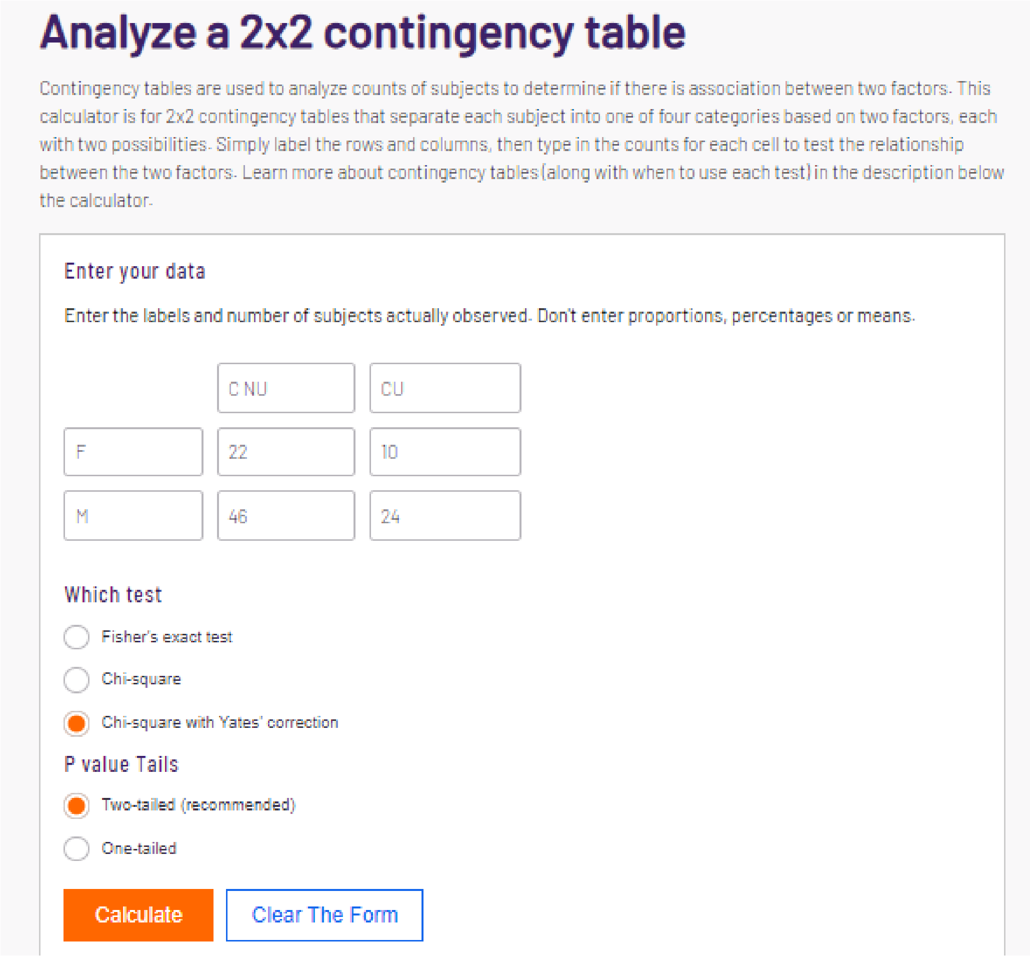


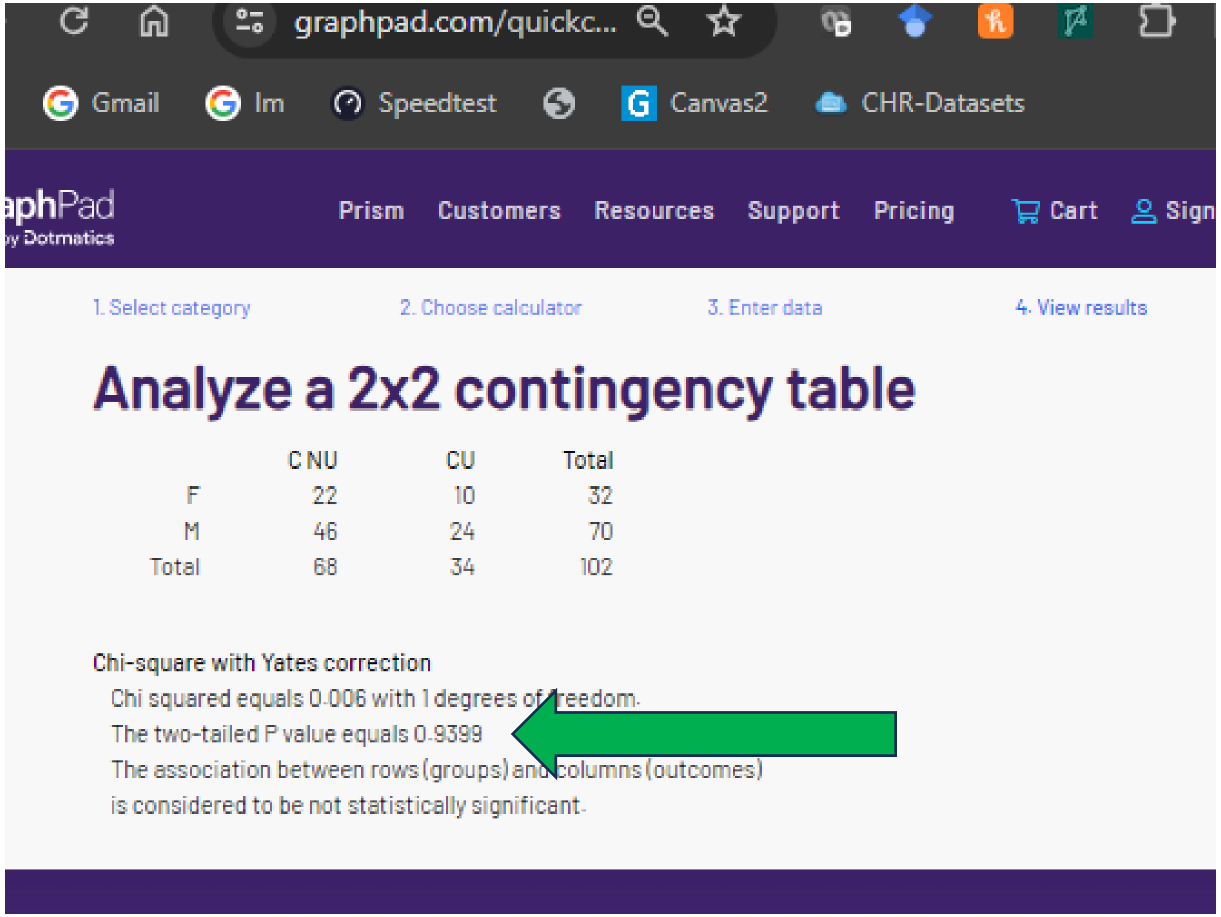


Note large (> 10) cell size

The "corrected" p value for gender **does** correspond with that of another (arguably inappropriate, not in methods) test.

### B. “corrected” chi-sq or Fisher’s p = .3568

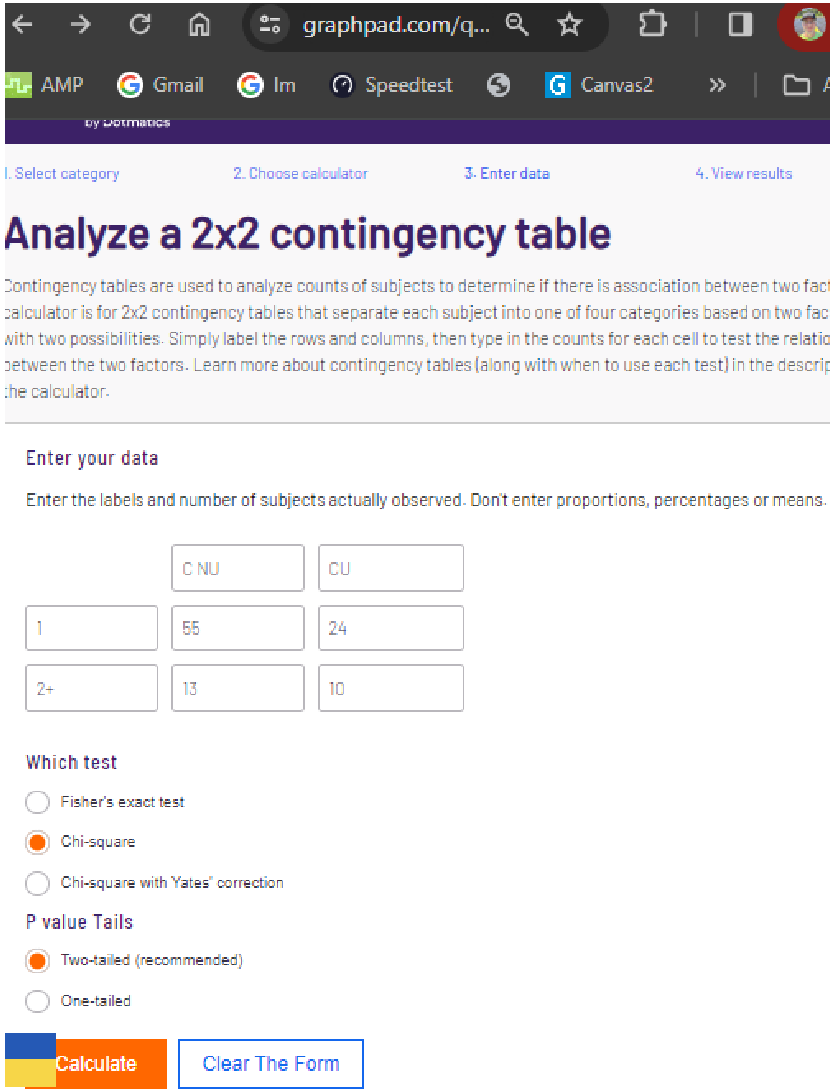


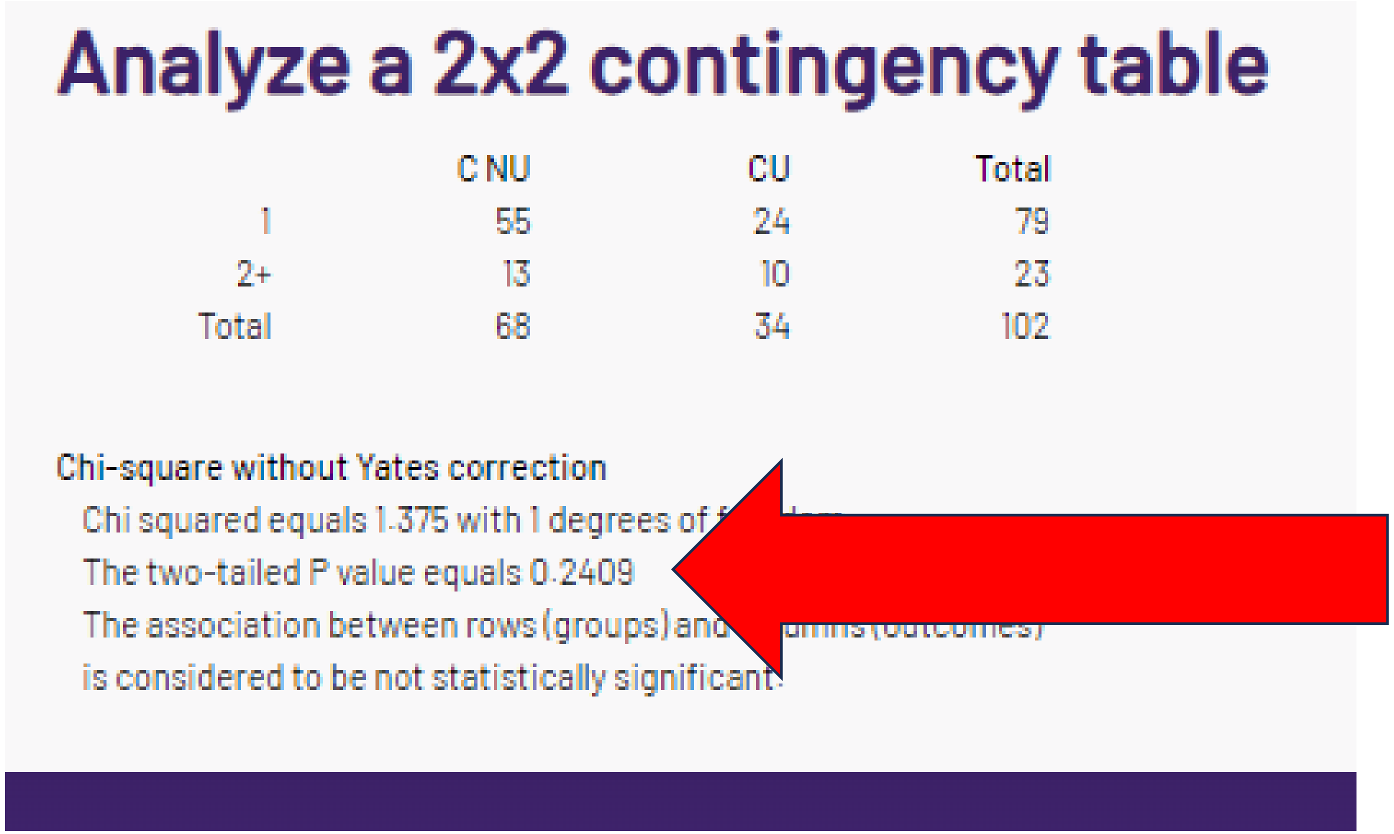

### B. “corrected” chi-sq or Fishers p = .3568

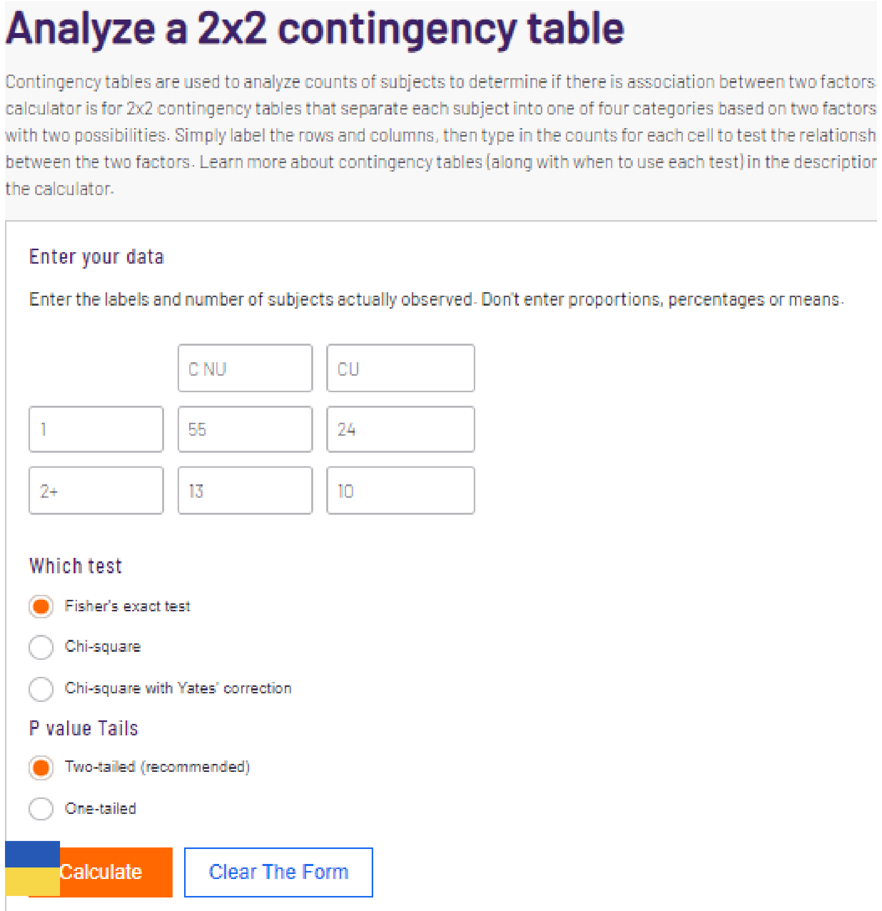


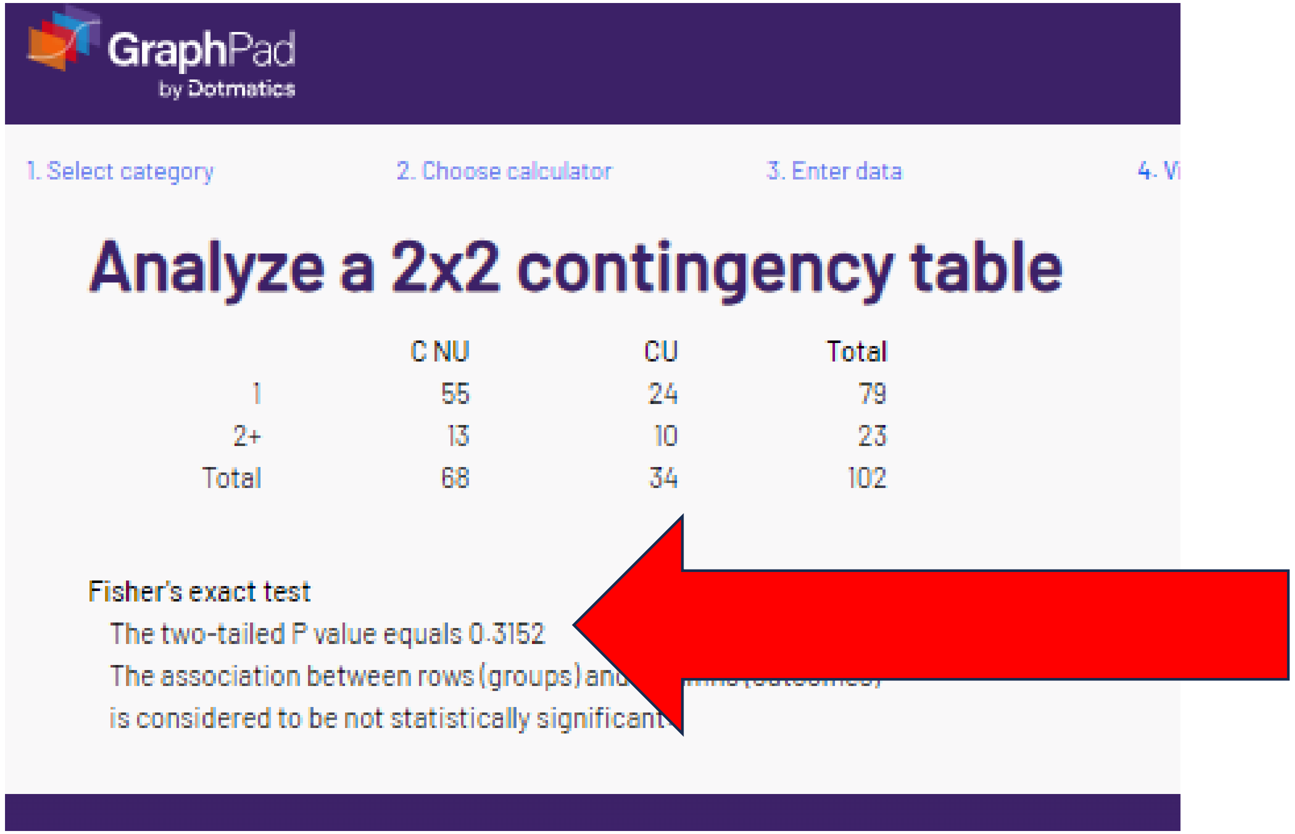


The reported ECOG p values could not be verified with chi-square or Fisher’s The cell size >= so Yates would not be warranted according to the BMJ guidance.

### B. “corrected” chi-sq or Fishers p = .3568

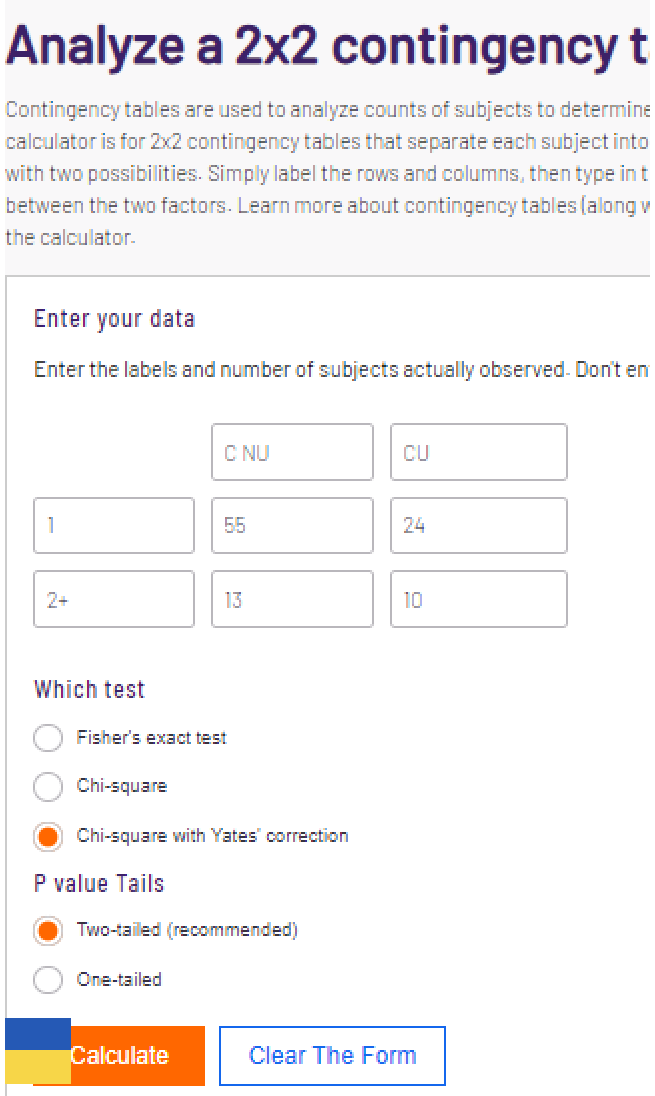


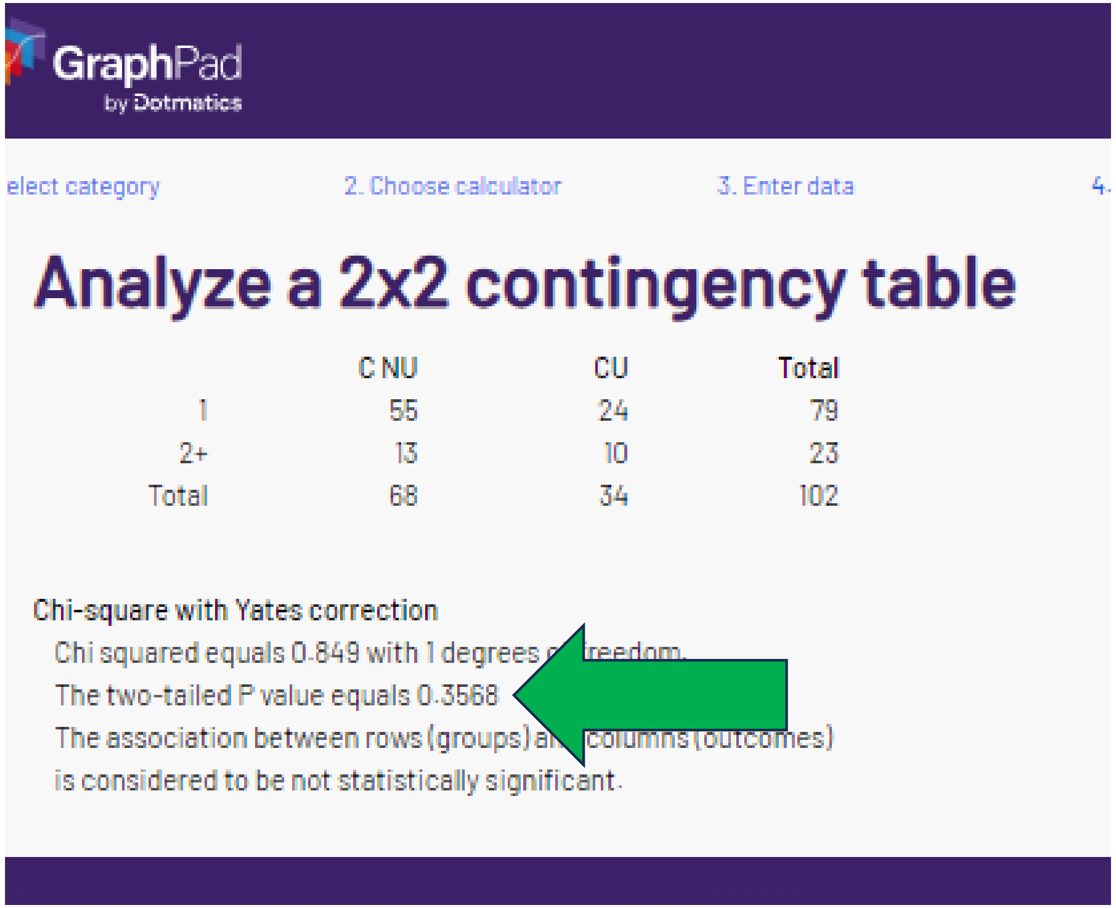


The p-value can be repeated with the **wrong** statistic.

Note that the N per cell is >= 10 so chi-square with Yates is **not** the appropriate statistic to run here.

### C. “corrected” chi-sq or Fishers p = .7124

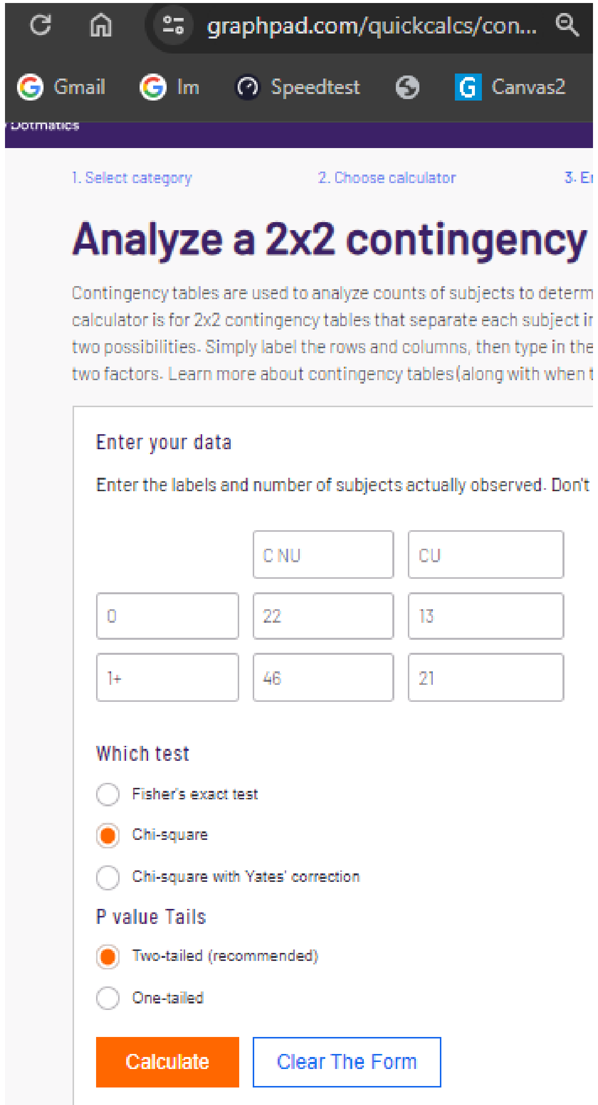


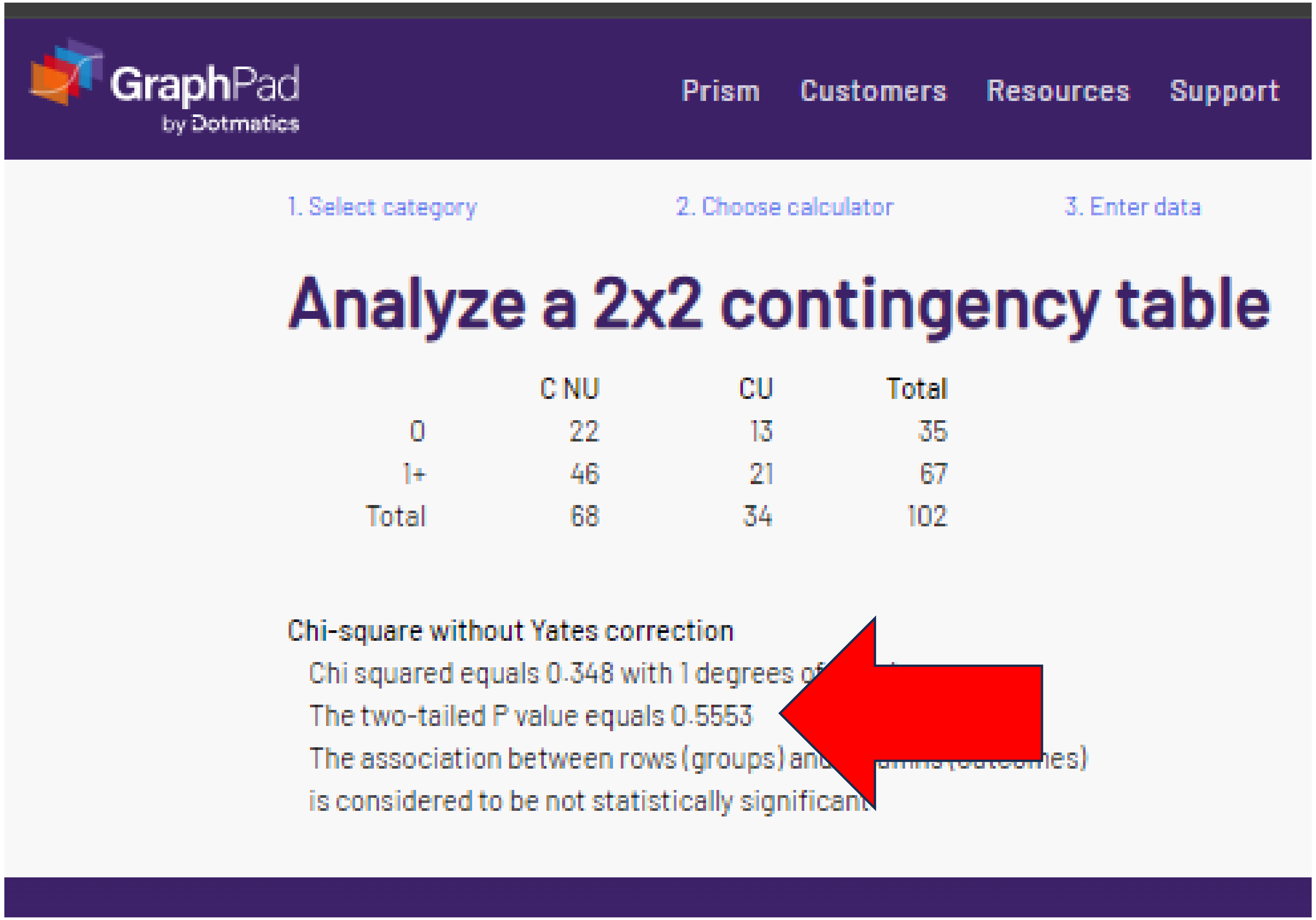


The 1+ is calculated as sum of 1 and 2+ groups

### C. “corrected” chi-sq or Fishers p = .7124

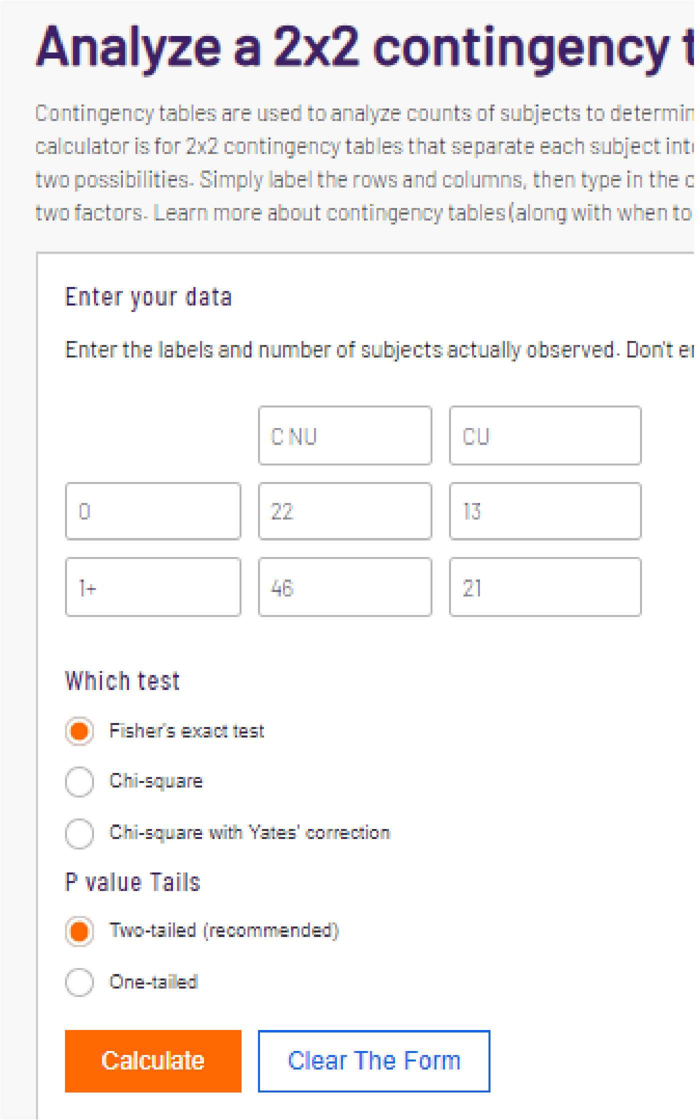

#### Analyze a 2x2 contingency t

Contingency tables are used to analyze counts of subjects to determin
calculator is for 2x2 contingency tables that separate each subject int
two possibilities. Simply label the rows and columns, then type in the c
two factors. Learn more about contingency tables (along with when to

Enter your data

Enter the labels and number of subjects actually observed. Don't er

| | | |
|---|---|---|
| | C NU | CU |
| 0 | 22 | 13 |
| 1+ | 46 | 21 |

Which test

- (selected) Fisher's exact test
- Chi-square
- Chi-square with Yates' correction

P value Tails

- (selected) Two-tailed (recommended)
- One-tailed

Calculate | Clear The Form

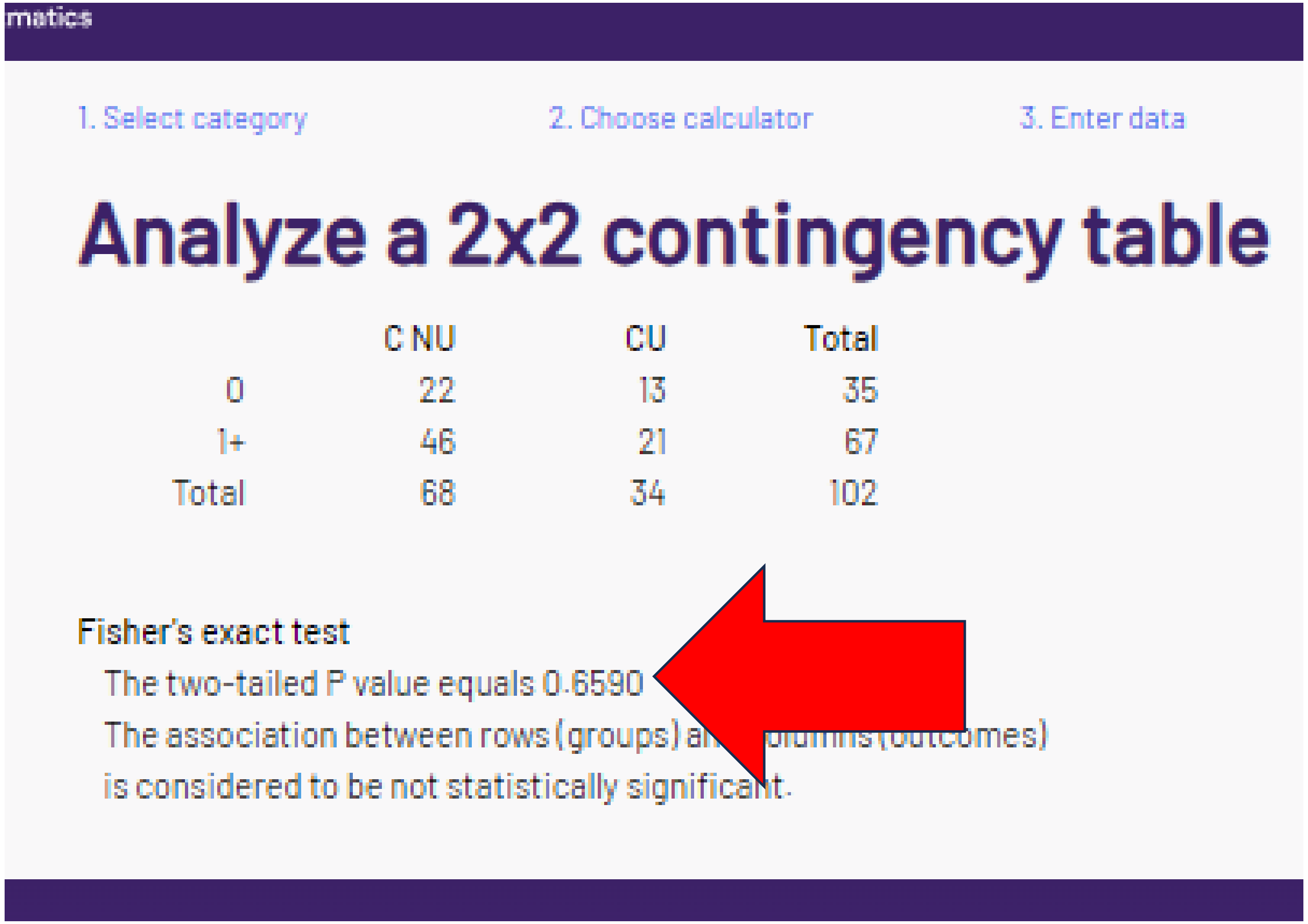

matics

1. Select category    2. Choose calculator    3. Enter data

#### Analyze a 2x2 contingency table

| | C NU | CU | Total |
|---|---|---|---|
| 0 | 22 | 13 | 35 |
| 1+ | 46 | 21 | 67 |
| Total | 68 | 34 | 102 |

Fisher's exact test

The two-tailed P value equals 0.6590

The association between rows (groups) a[illegible]olumns (outcomes)

is considered to be not statistically significant.

The Chronic Diseases / patient (0 vs 1+) could not be verified with chi-square or Fisher’s.

### C. “corrected” chi-sq or Fishers p = .7124

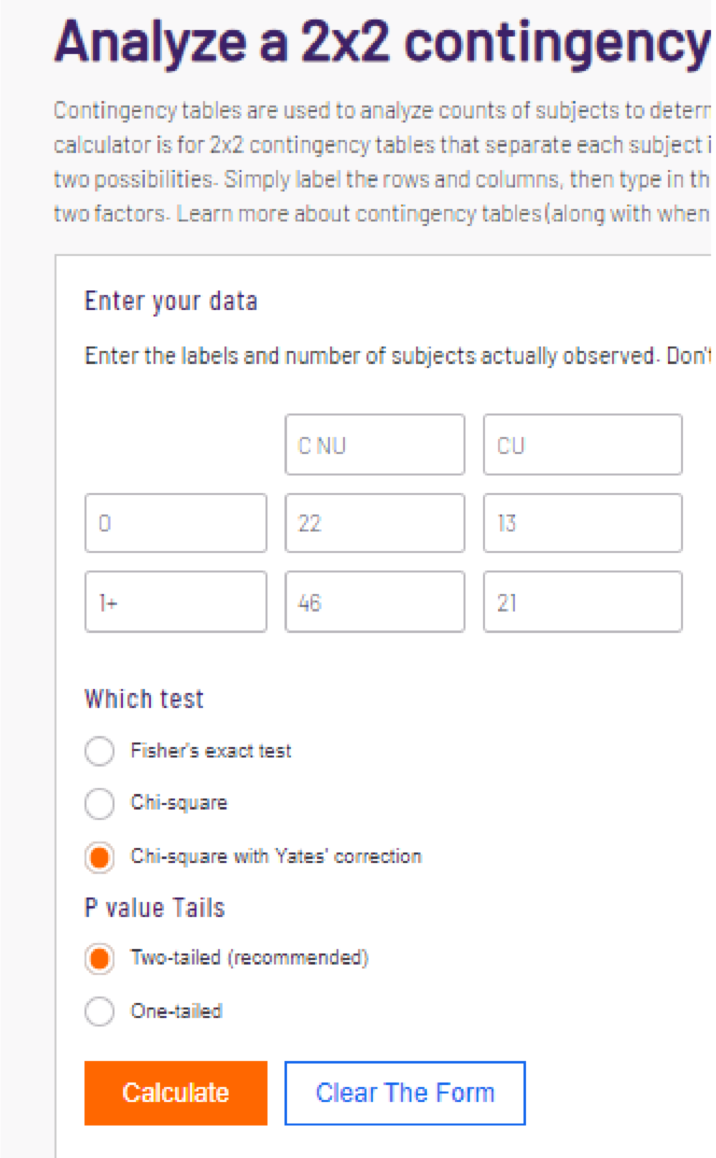


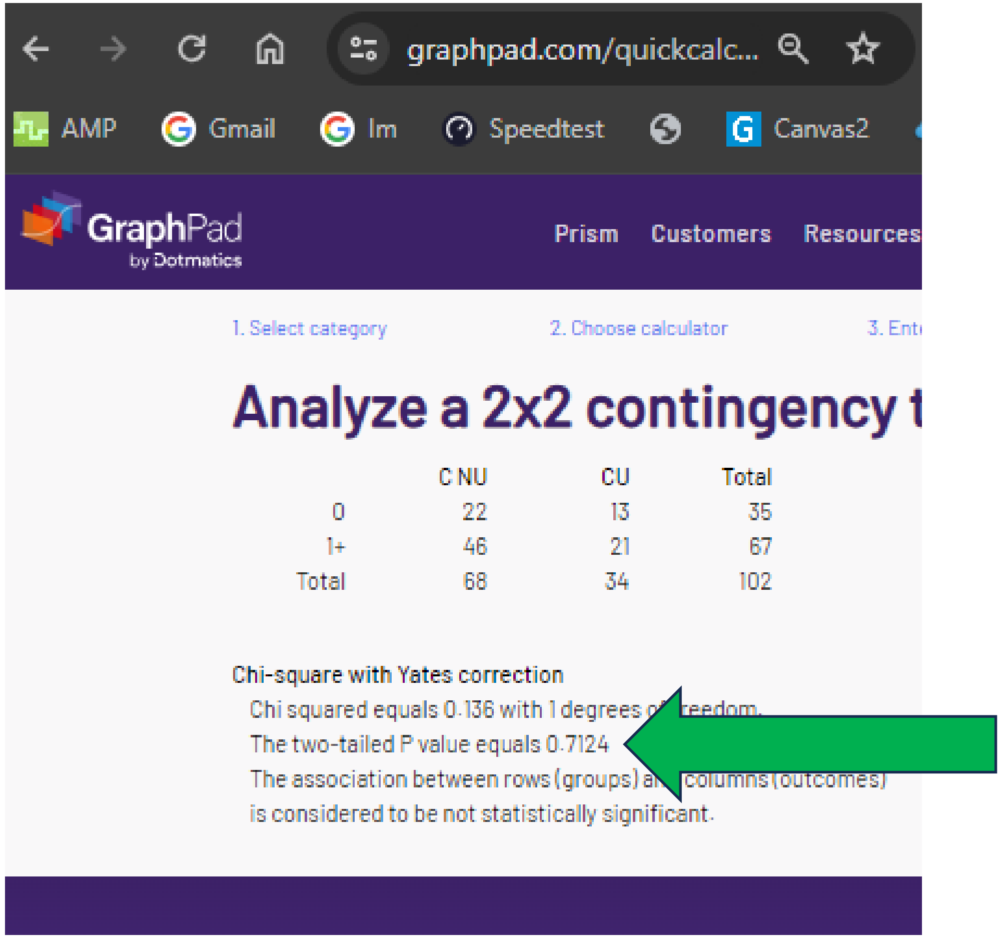


The Chronic Diseases / patient (0 vs 1+) does correspond with that of the wrong statistic.

Note that N per cell > 20 so Yates is **inappropriate** and should **not** be reported here.

### D. “corrected” chi-sq or Fishers p = .9332

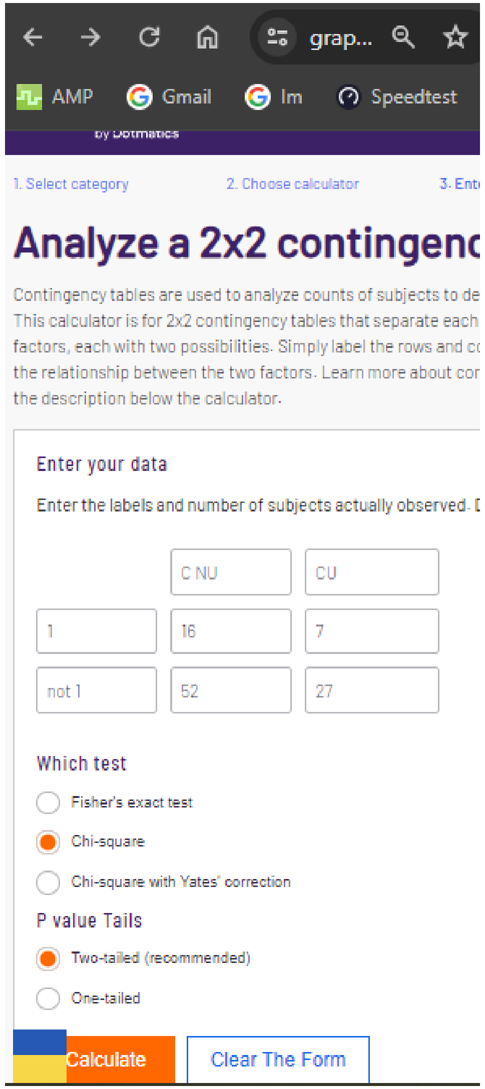


‘Not 1’ calculated as sum of 0 and 2+

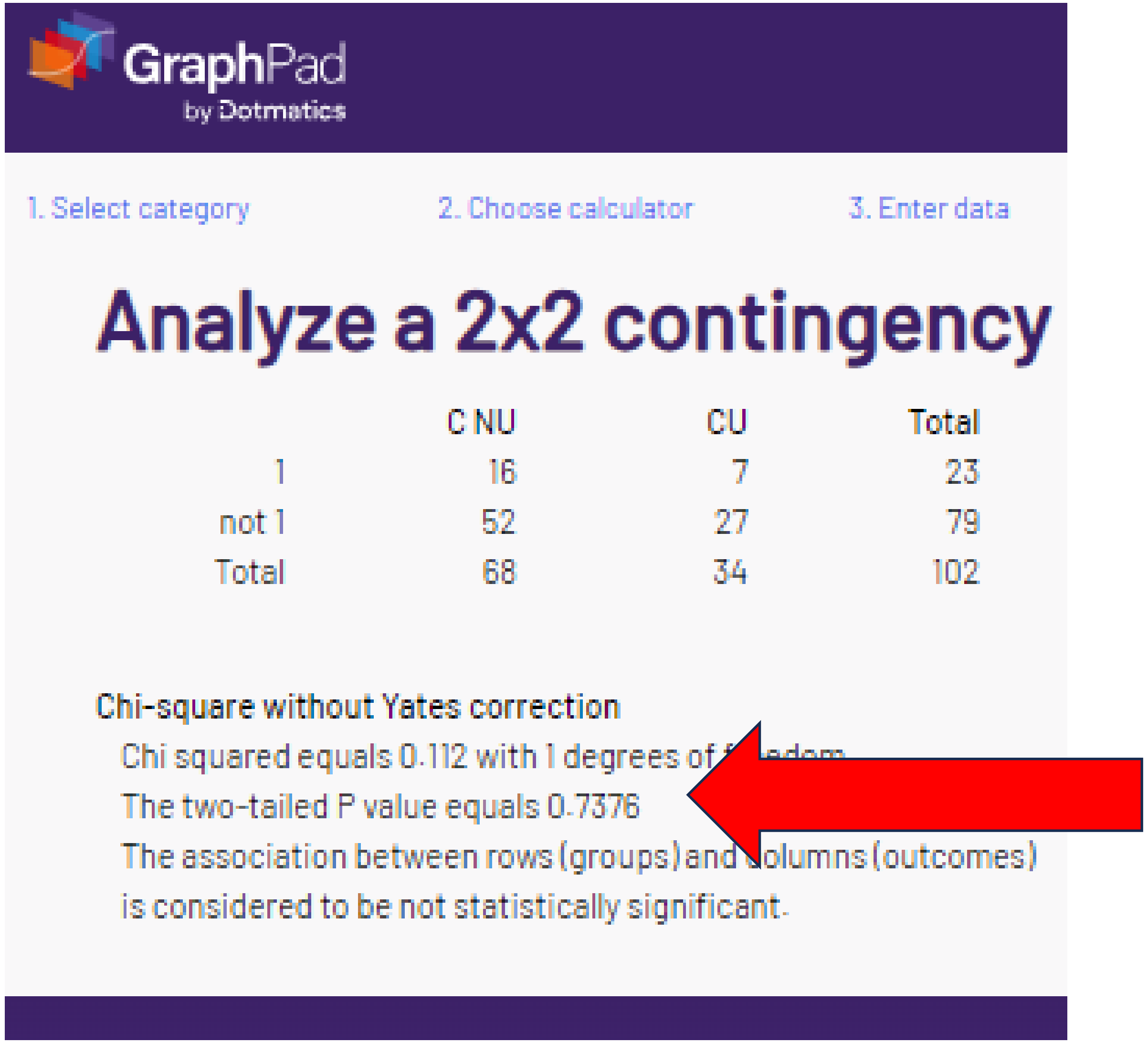

### D. “corrected” chi-sq or Fishers p = .9332

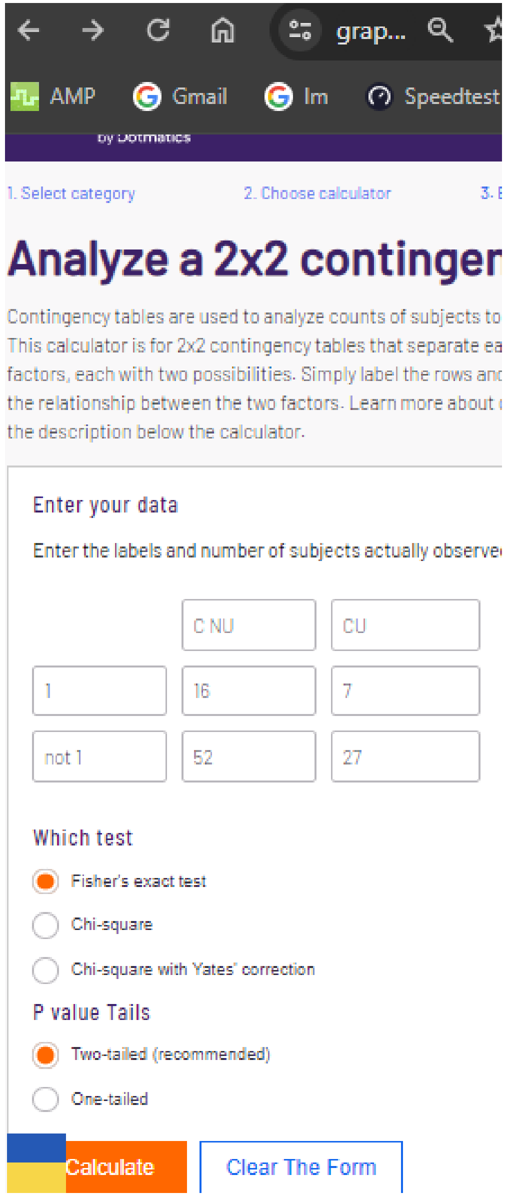


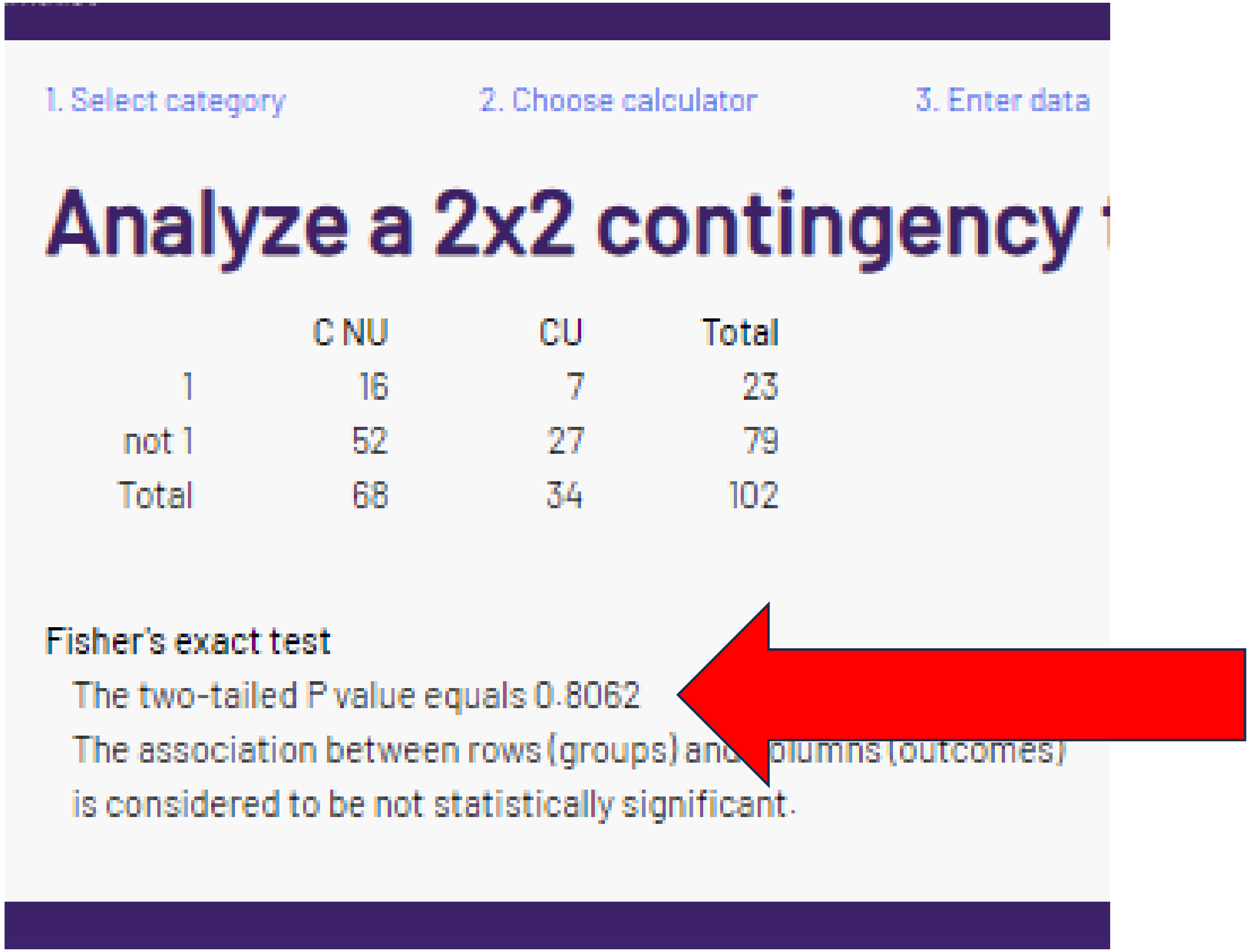


P value for chronic diseases = 1 **not verified** with chi-square or Fisher’s.

### D. “corrected” chi-sq or Fishers p = .9332

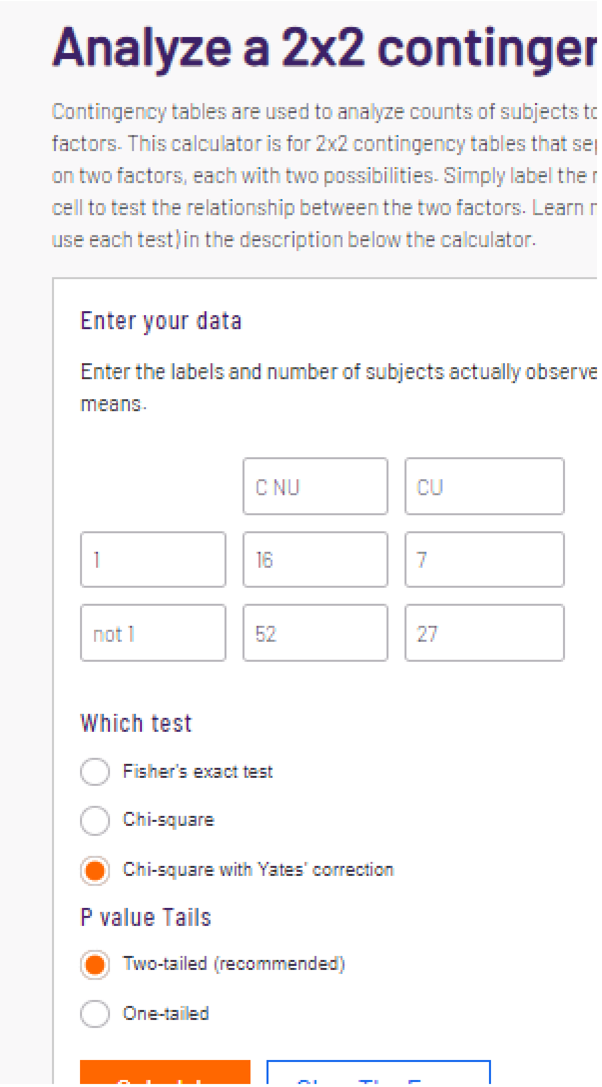


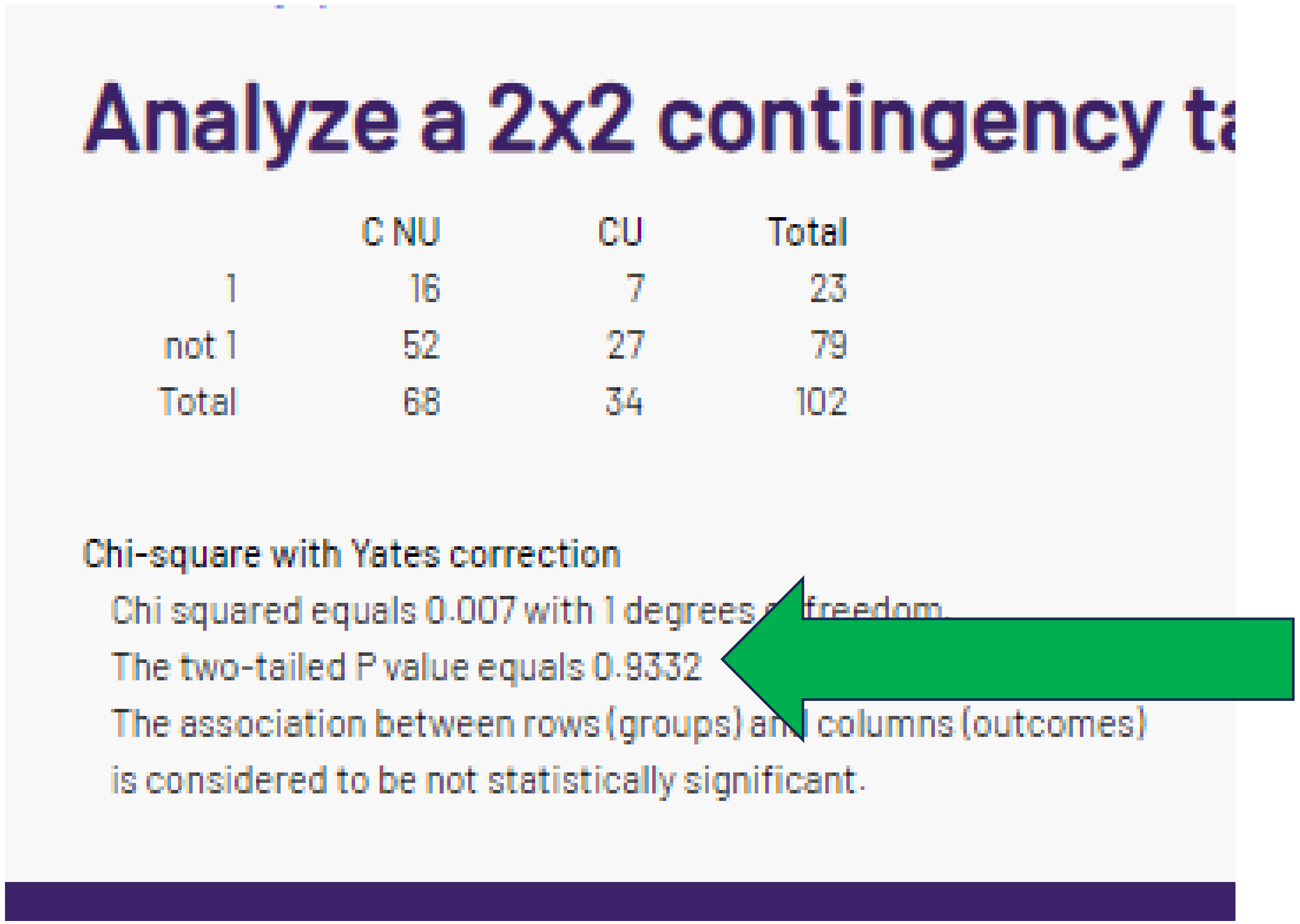


P value for chronic diseases = 1 can be reproduced but only using an **inappropriate statistic** (not in methods).

Note that N per cell is > 5 so Yates is the wrong statistic according to the Wikipedia criteria.
It could be applied according to BMJ guidance (although, again, only if specified in the methods).

### E. “corrected” chi-sq or Fishers p = .9437

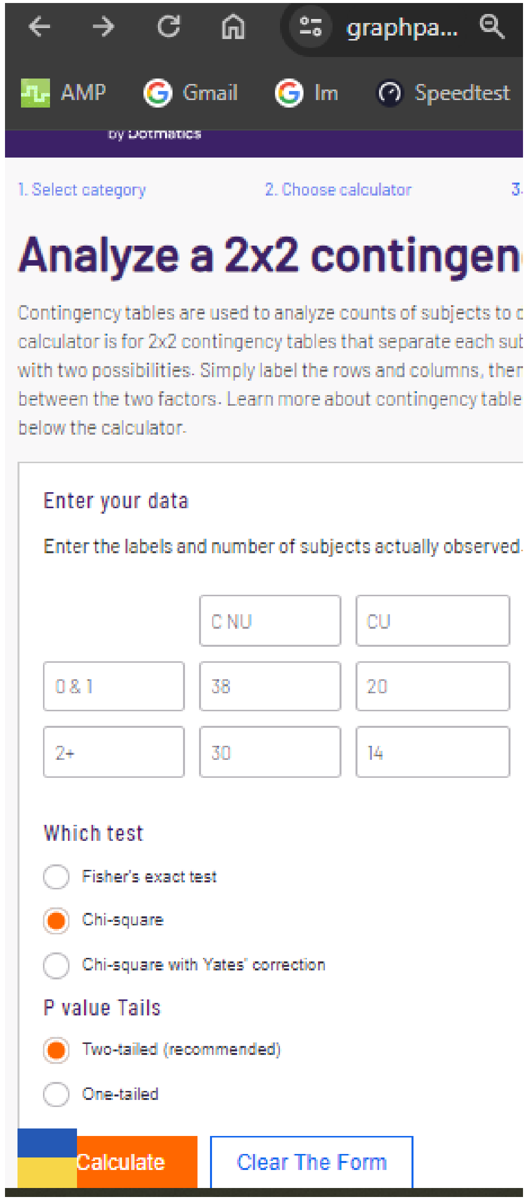


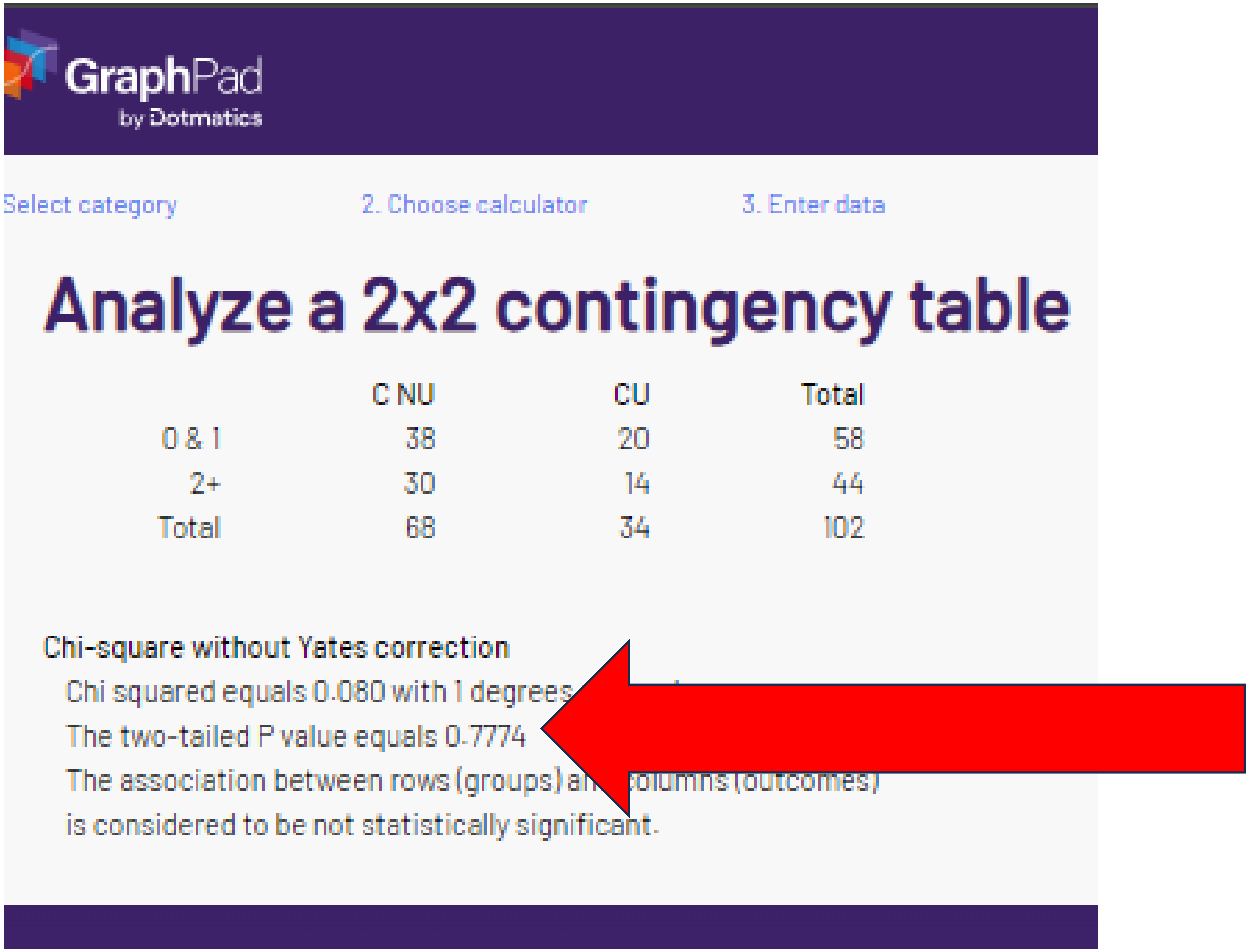

### E. “corrected” chi-sq or Fishers p = .9437

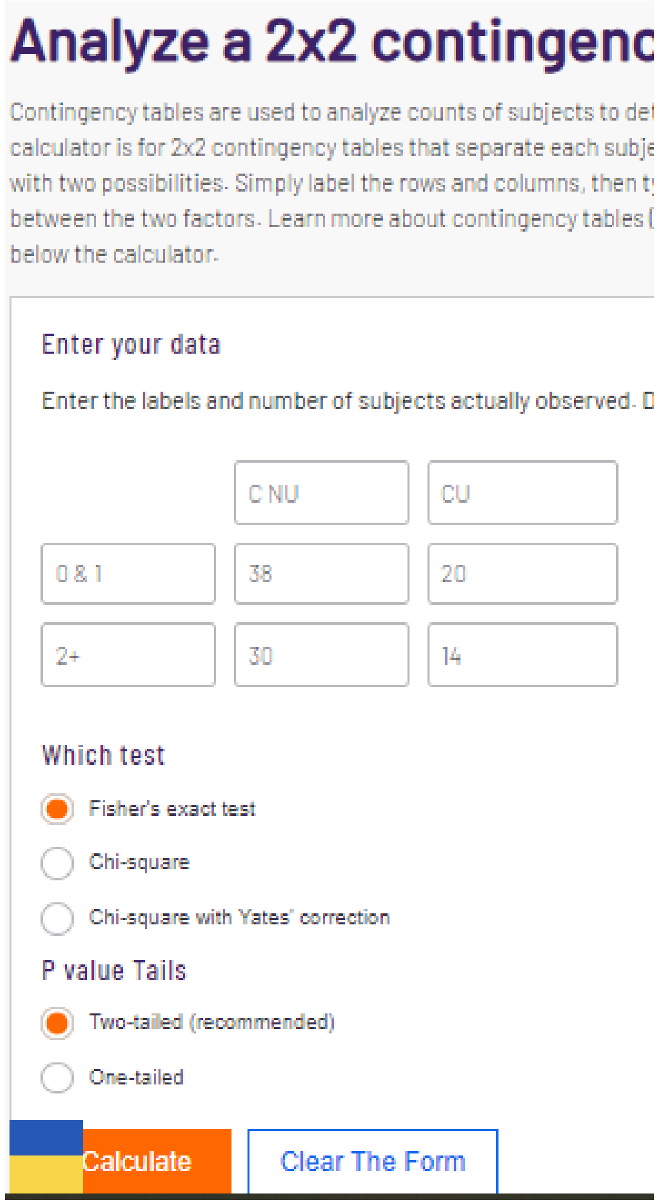

#### Analyze a 2x2 contingenc

Contingency tables are used to analyze counts of subjects to det
calculator is for 2x2 contingency tables that separate each subje
with two possibilities. Simply label the rows and columns, then t
between the two factors. Learn more about contingency tables (
below the calculator.

Enter your data

Enter the labels and number of subjects actually observed. D

| | | |
|---|---|---|
| | C NU | CU |
| 0 & 1 | 38 | 20 |
| 2+ | 30 | 14 |

Which test

- (selected) Fisher's exact test
- Chi-square
- Chi-square with Yates' correction

P value Tails

- (selected) Two-tailed (recommended)
- One-tailed

Calculate | Clear The Form

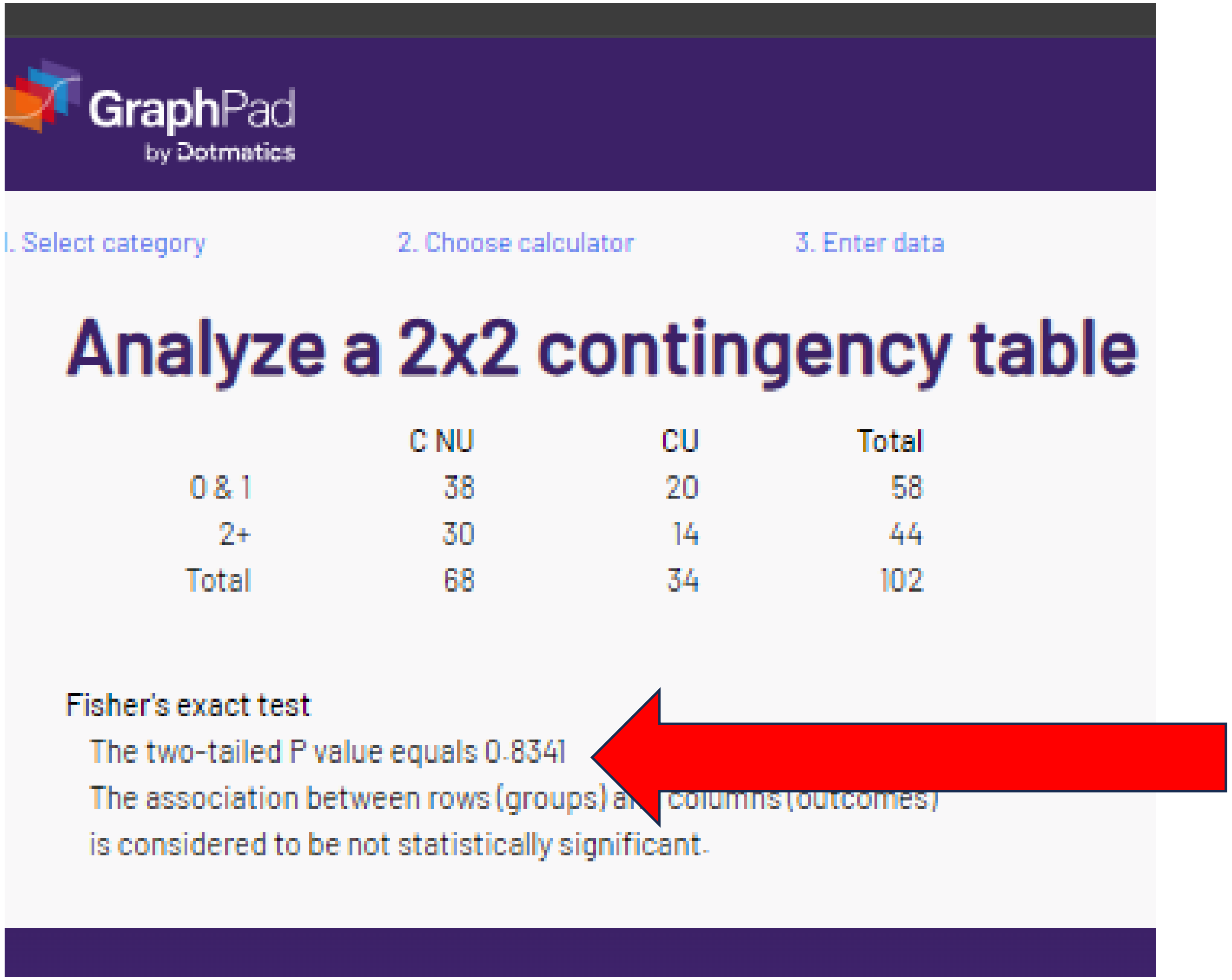

GraphPad by Dotmatics

1. Select category | 2. Choose calculator | 3. Enter data

#### Analyze a 2x2 contingency table

| | C NU | CU | Total |
|---|---|---|---|
| 0 & 1 | 38 | 20 | 58 |
| 2+ | 30 | 14 | 44 |
| Total | 68 | 34 | 102 |

Fisher's exact test

The two-tailed P value equals 0.8341
The association between rows (groups) a[illegible] columns (outcomes)
is considered to be not statistically significant.

For chronic diseases 2+, the “corrected” statistics (chi-sq or Fisher’s) could **not** be verified.

### E. “corrected” chi-sq or Fishers p = .9437

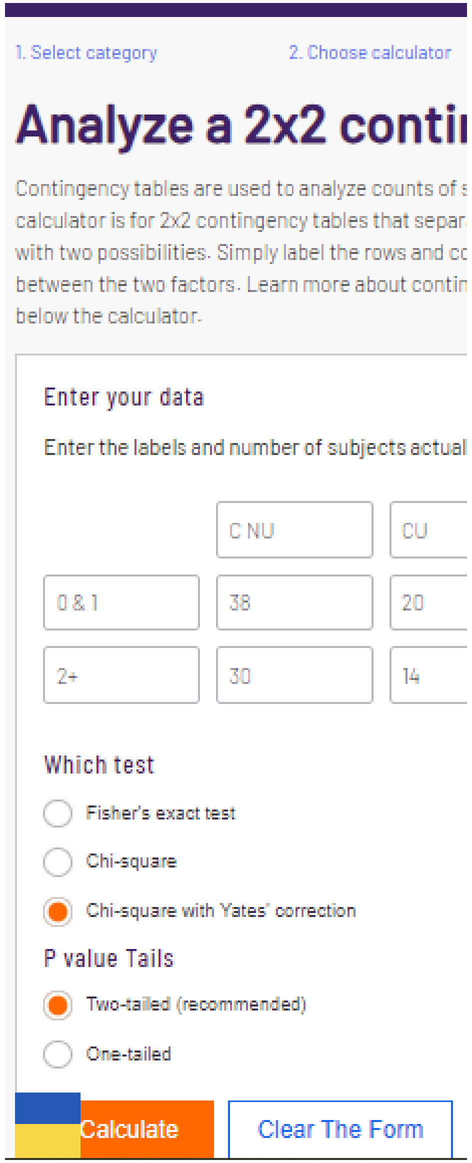


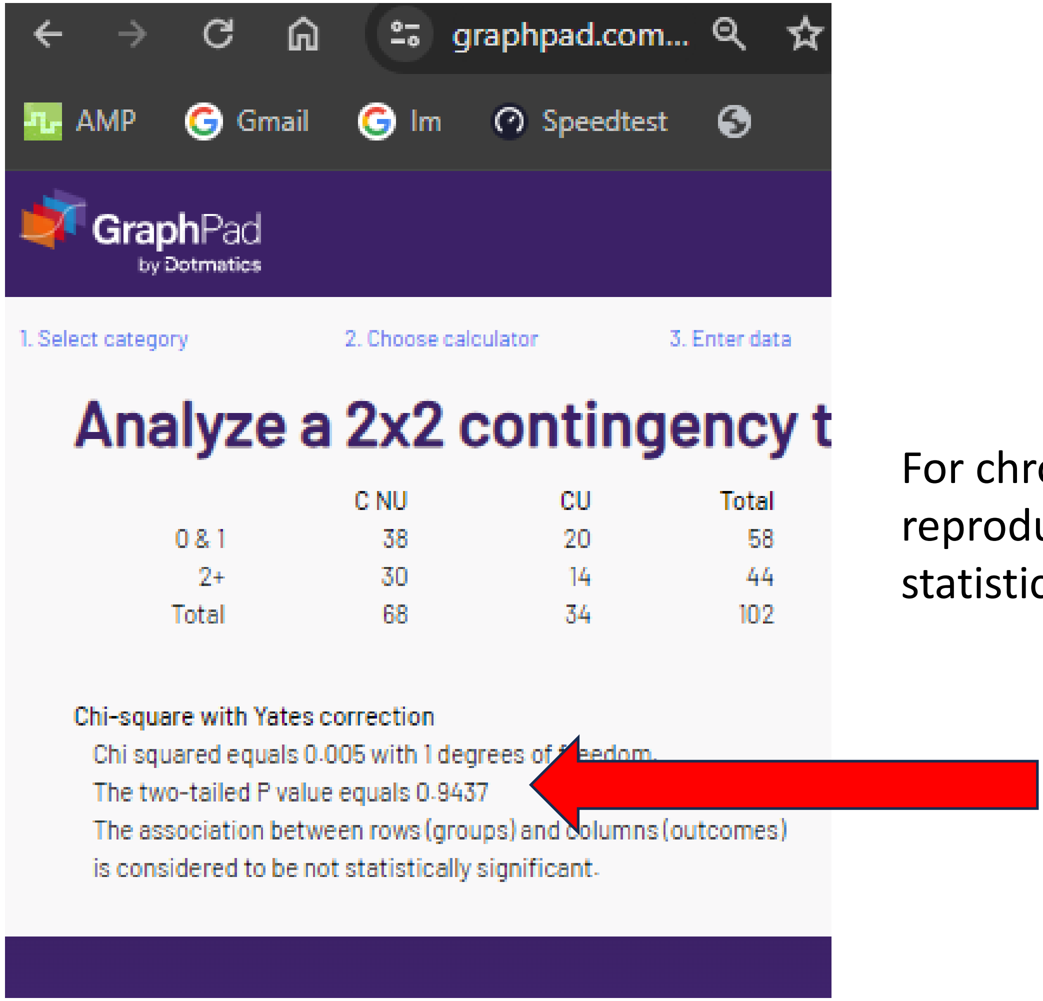


Note large N.

For chronic diseases 2+, the p value could be reproduced but only with an Inappropriate statistic.

As the N per cell is large (>13), Yates is an inappropriate statistic and should not be reported.

### F. “corrected” chi-sq or Fishers p = .2762

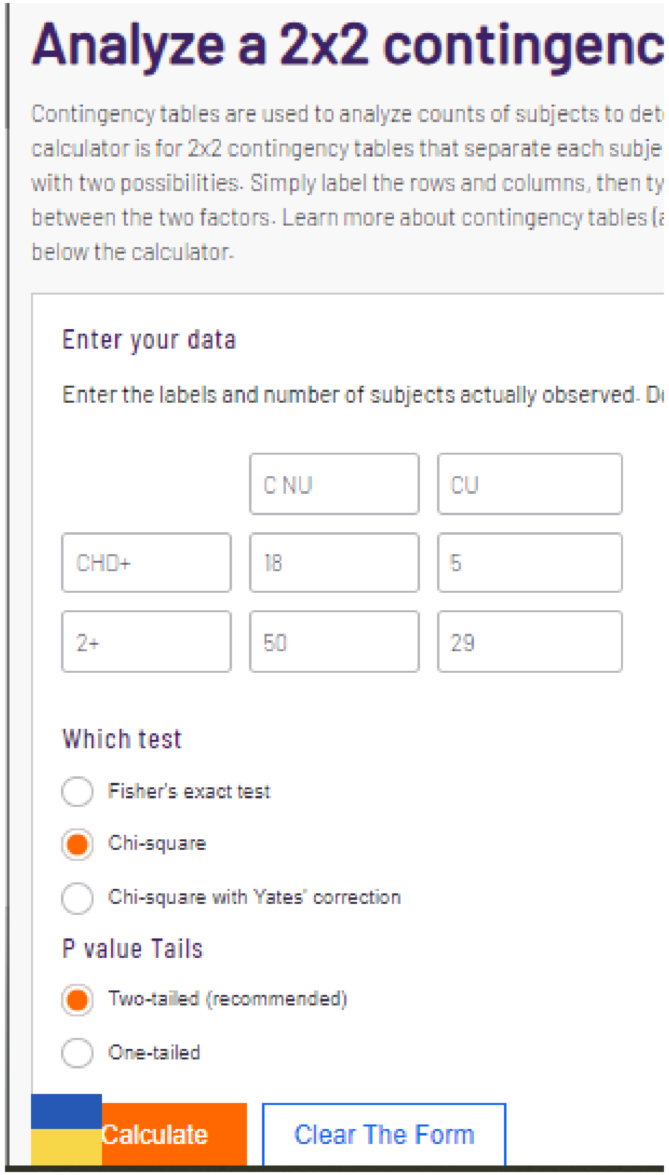


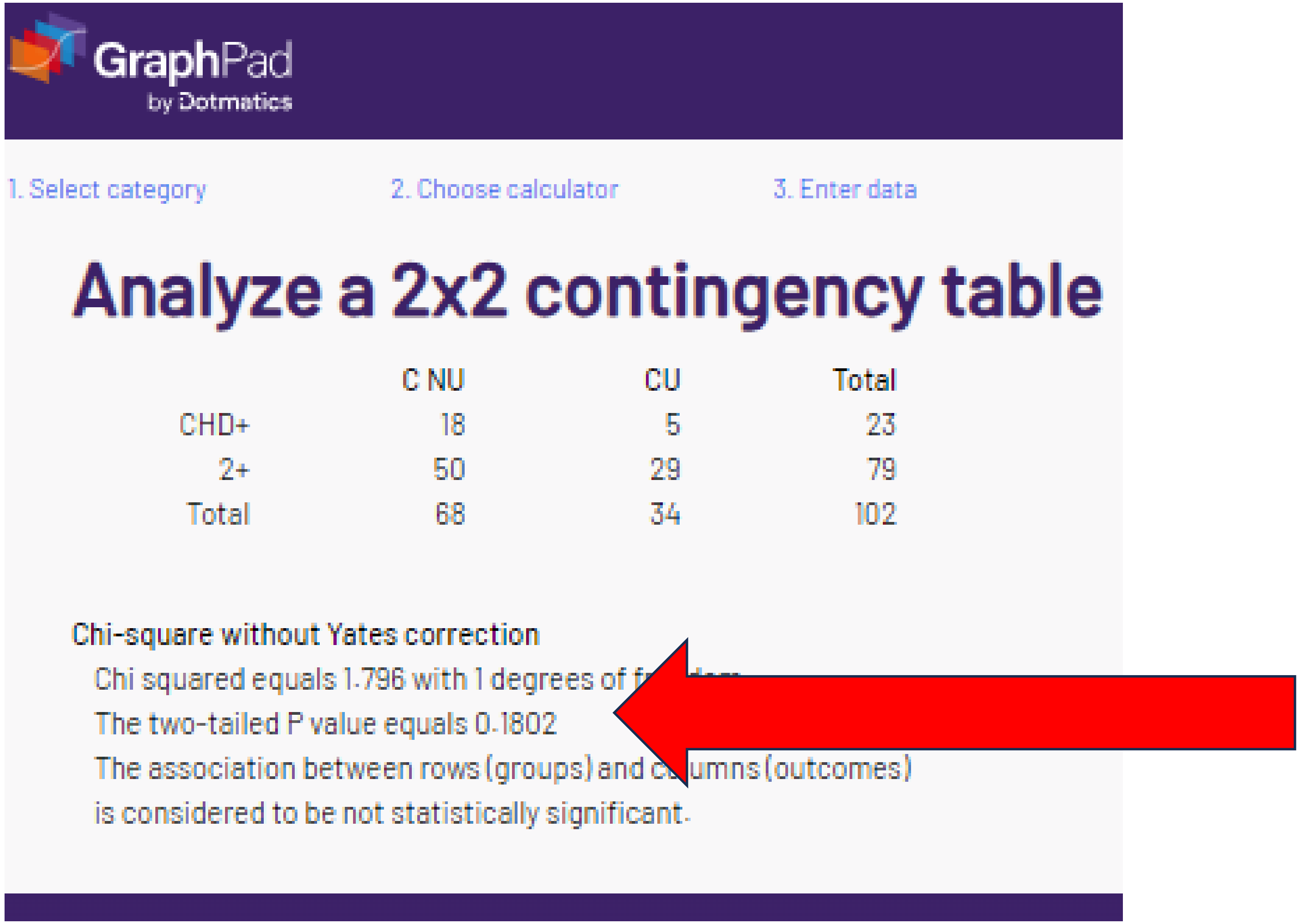

### F. “corrected” chi-sq or Fishers p = .2762

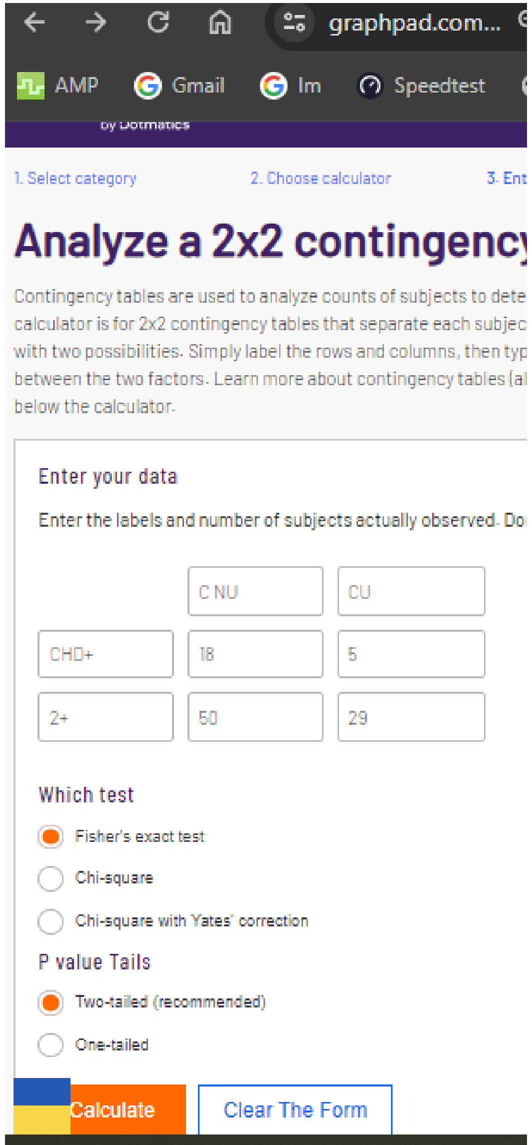


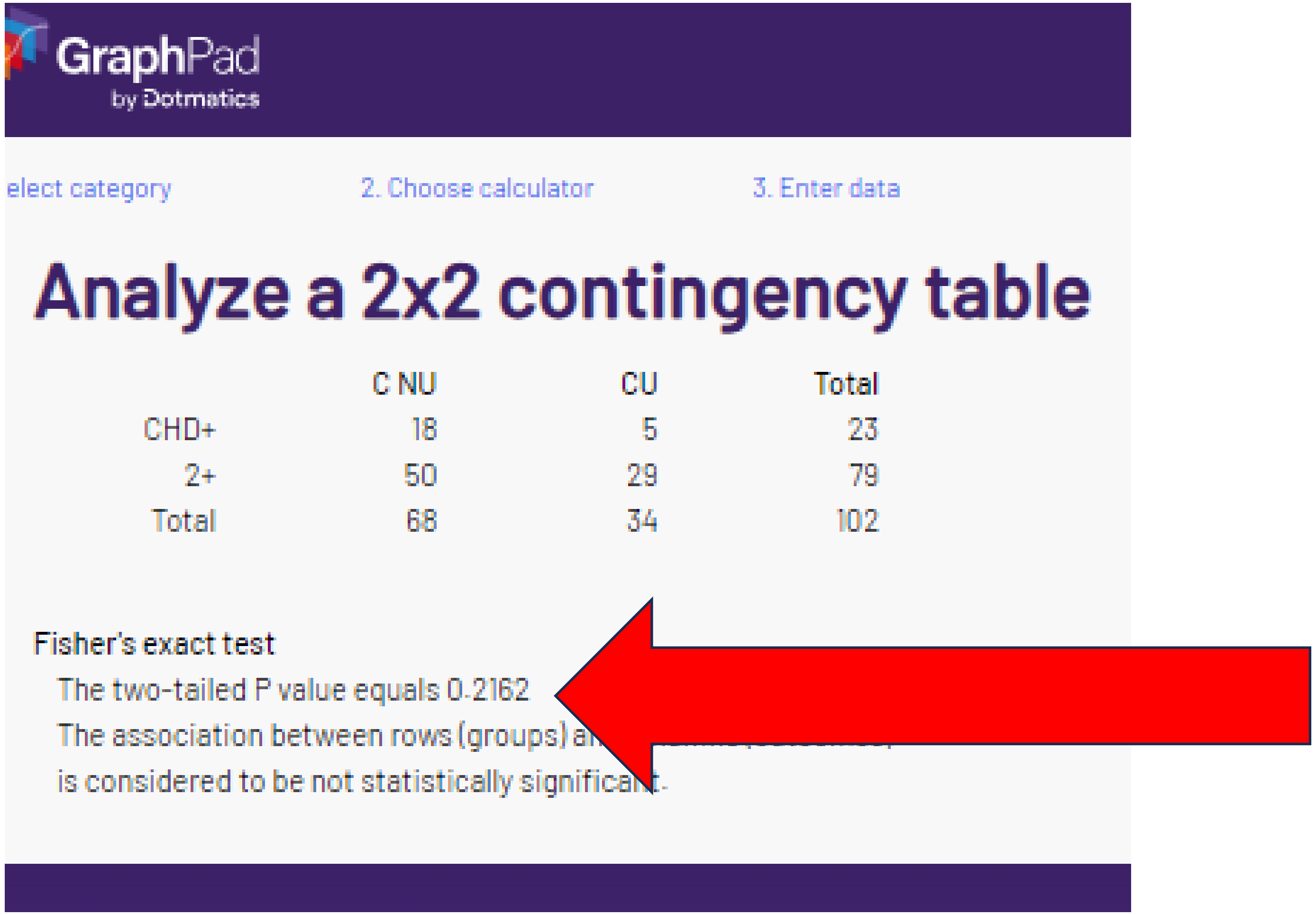


The reported “corrected” p value could not be verified with chi-square or Fisher’s.

### F. "corrected" chi-sq or Fishers p = .2762

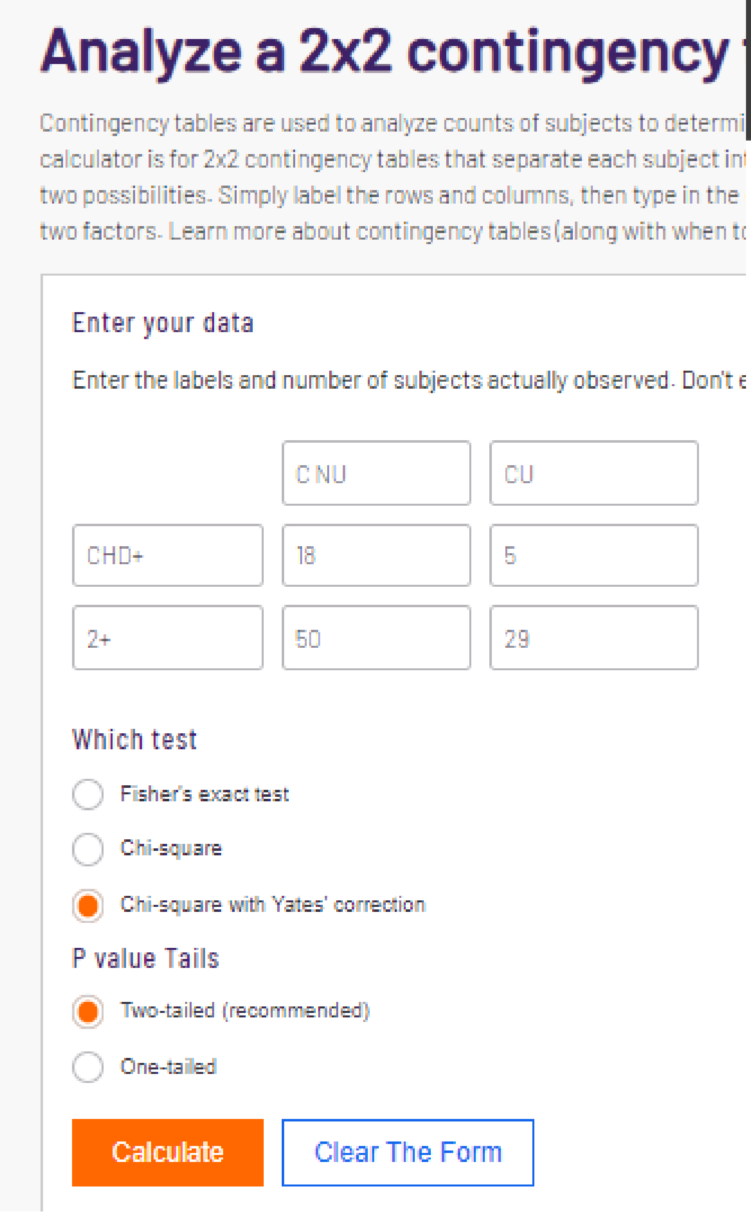


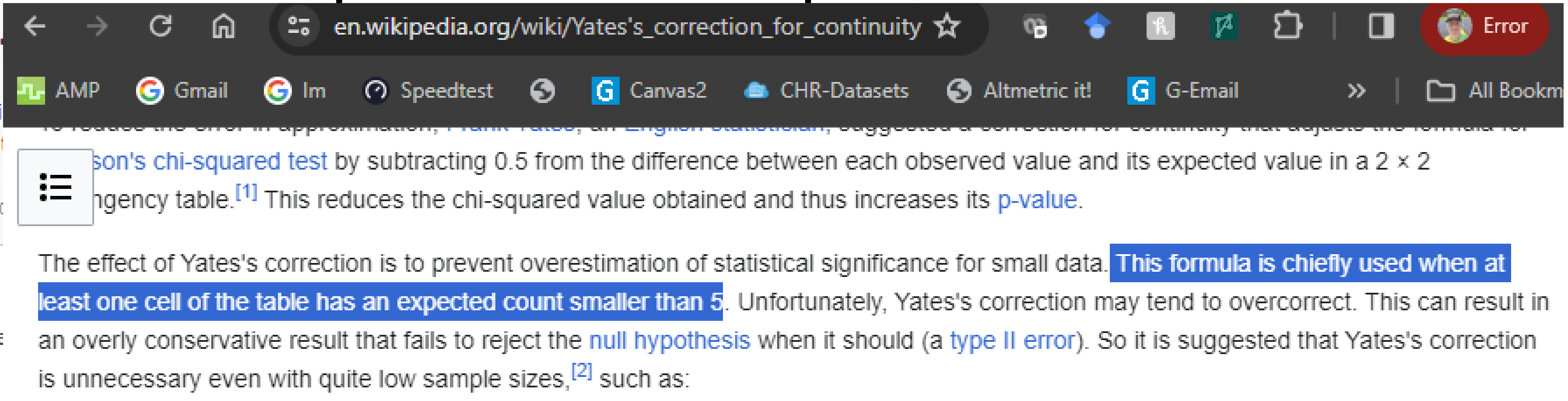


According to Wikipedia (above), an N < 5 might warrant Yates correction. However, as the smallest cell observed = 5, it would not be warranted here according to the Wiki convention. It could be used (when in methods) according to the BMJ Guidance.

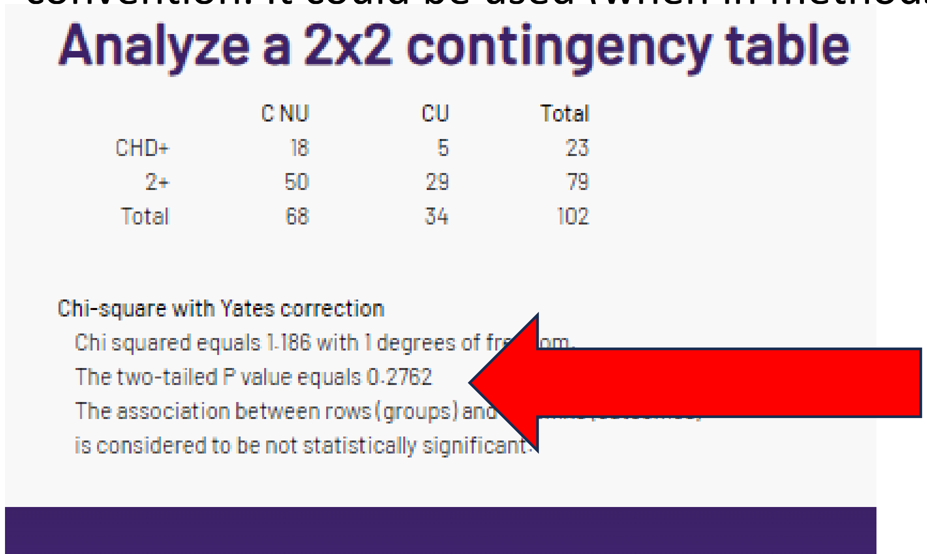
Analyze a 2x2 contingency table

| | C NU | CU | Total |
|---|---|---|---|
| CHD+ | 18 | 5 | 23 |
| 2+ | 50 | 29 | 79 |
| Total | 68 | 34 | 102 |

Chi-square with Yates correction
Chi squared equals 1.186 with 1 degrees of fr[illegible]om.
The two-tailed P value equals 0.2762
The association between rows (groups) and [illegible]
is considered to be not statistically significant.

P value is reproduced but with (arguably) the wrong statistic (not in methods section too).

### G. “corrected” chi-sq or Fishers p = .5576

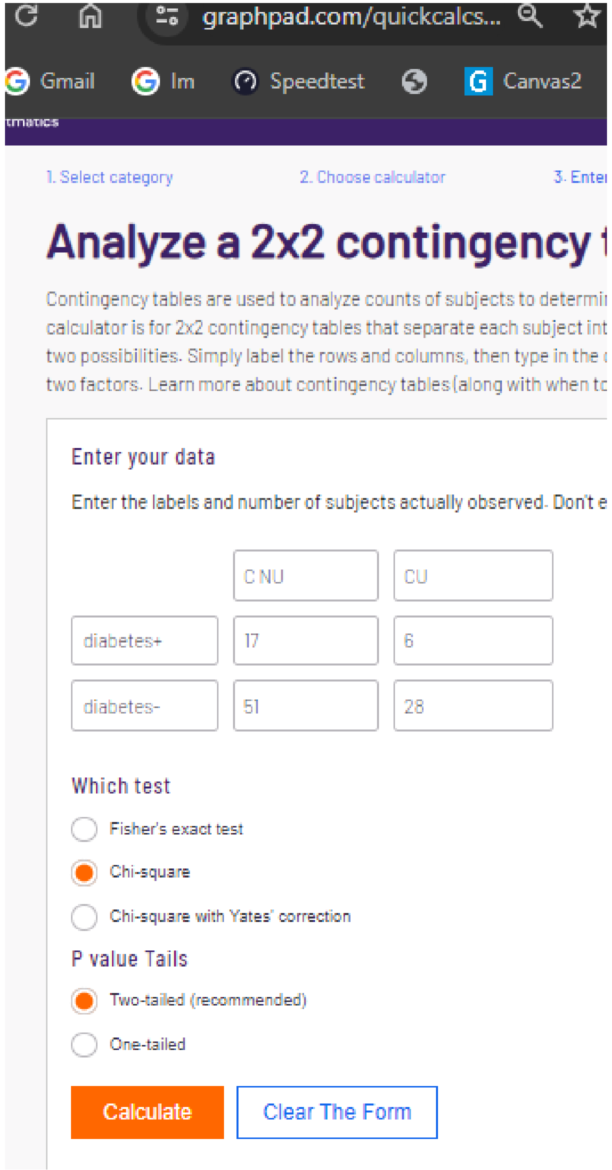


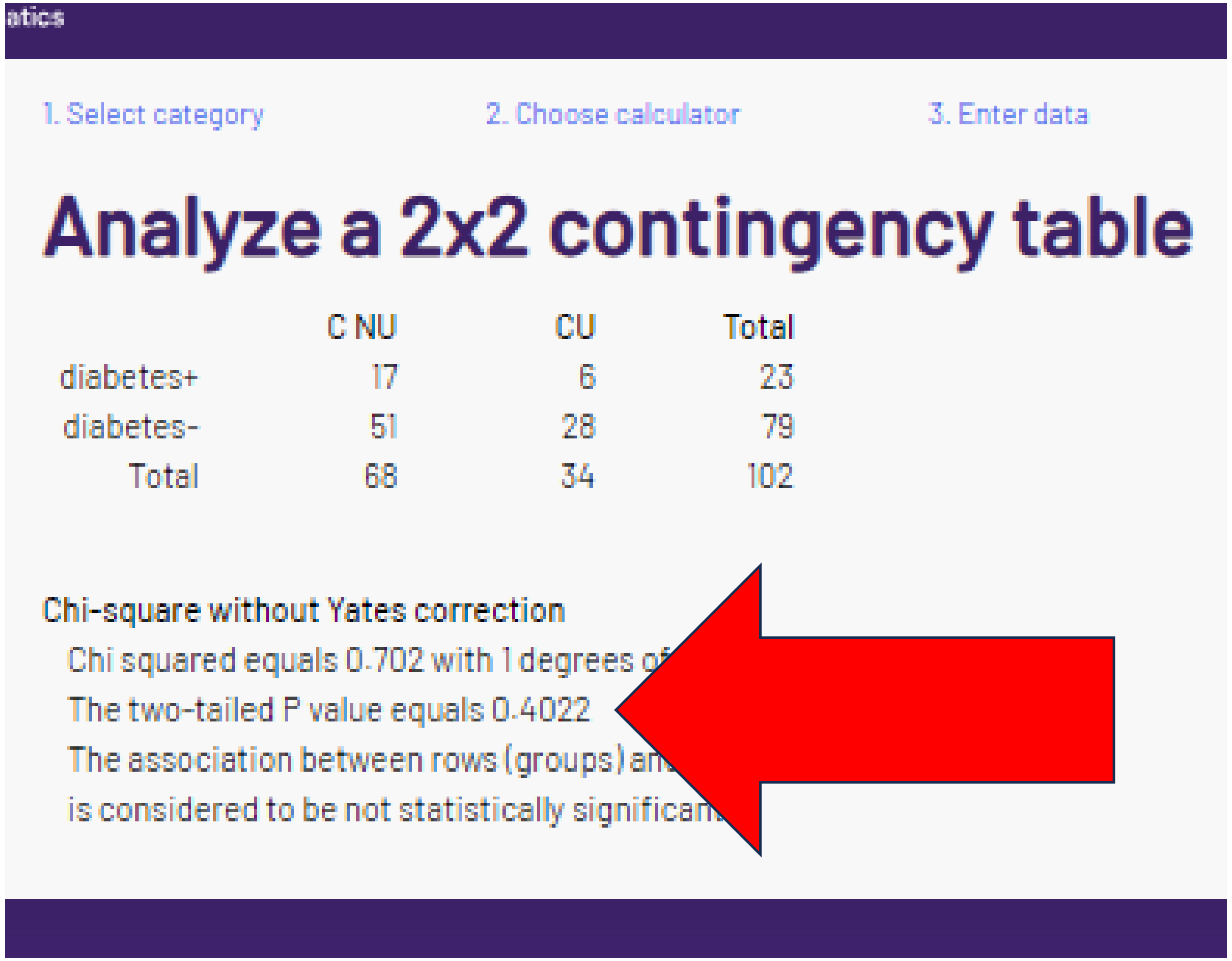

### G. “corrected” chi-sq or Fisher’s p = .5576

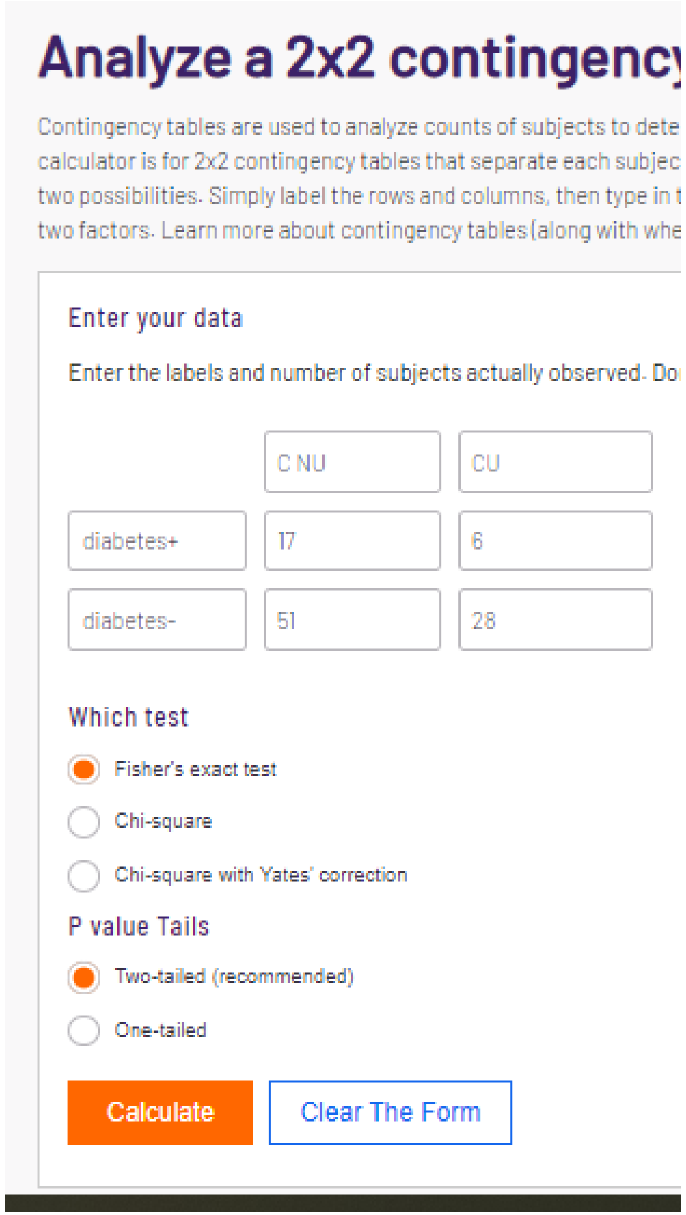

#### Analyze a 2x2 contingency

Contingency tables are used to analyze counts of subjects to dete
calculator is for 2x2 contingency tables that separate each subjec
two possibilities. Simply label the rows and columns, then type in t
two factors. Learn more about contingency tables (along with whe

Enter your data

Enter the labels and number of subjects actually observed. Do

| | C NU | CU |
|---|---|---|
| diabetes+ | 17 | 6 |
| diabetes- | 51 | 28 |

Which test

- Fisher's exact test
- Chi-square
- Chi-square with Yates' correction

P value Tails

- Two-tailed (recommended)
- One-tailed

Calculate | Clear The Form

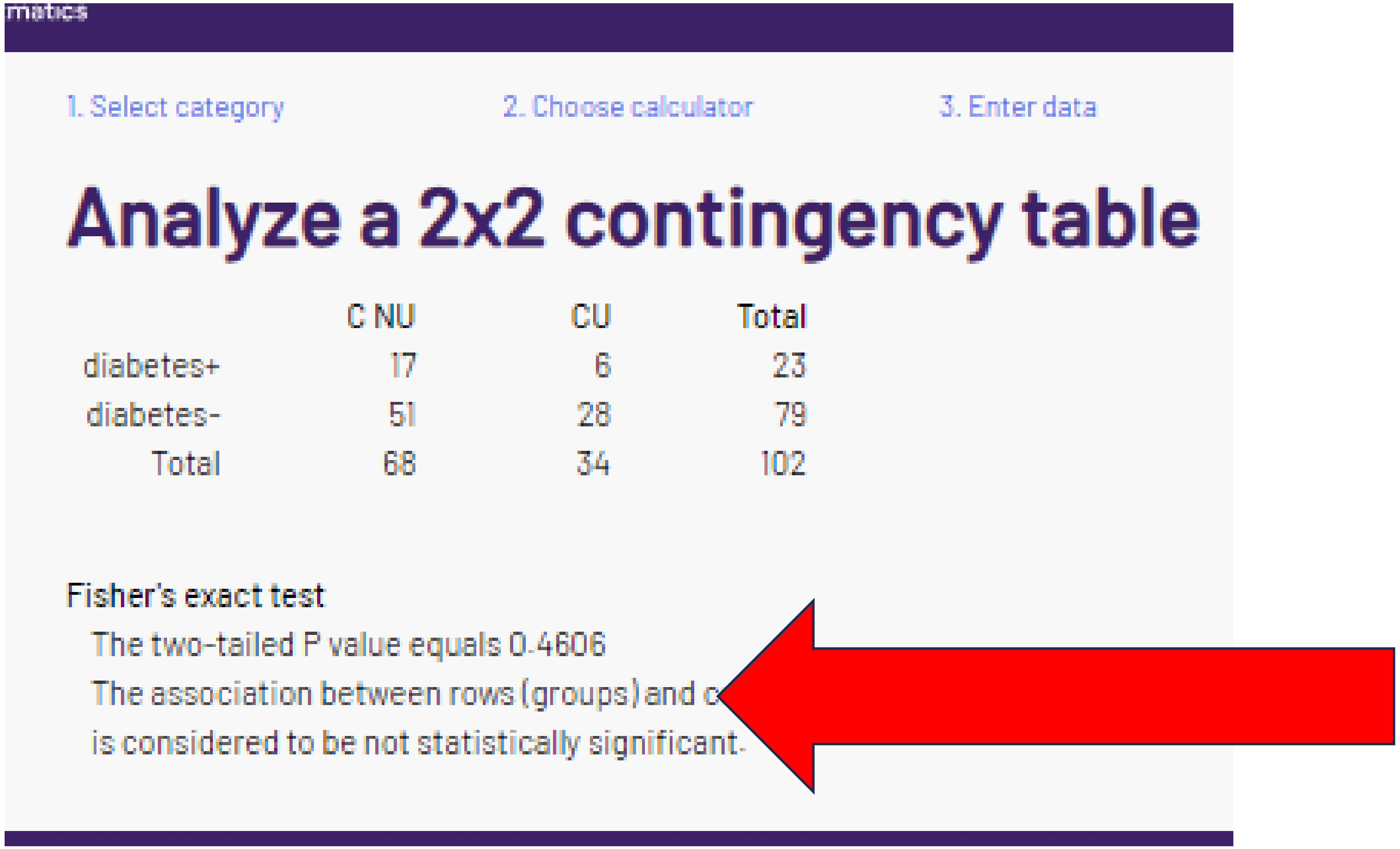

matics

1. Select category 2. Choose calculator 3. Enter data

#### Analyze a 2x2 contingency table

| | C NU | CU | Total |
|---|---|---|---|
| diabetes+ | 17 | 6 | 23 |
| diabetes- | 51 | 28 | 79 |
| Total | 68 | 34 | 102 |

Fisher's exact test
The two-tailed P value equals 0.4606
The association between rows (groups) and c
is considered to be not statistically significant.

The “corrected” p value is not reproduced for chi-square or Fisher’s for diabetes.

### G. “corrected” chi-sq or Fishers p = .5576

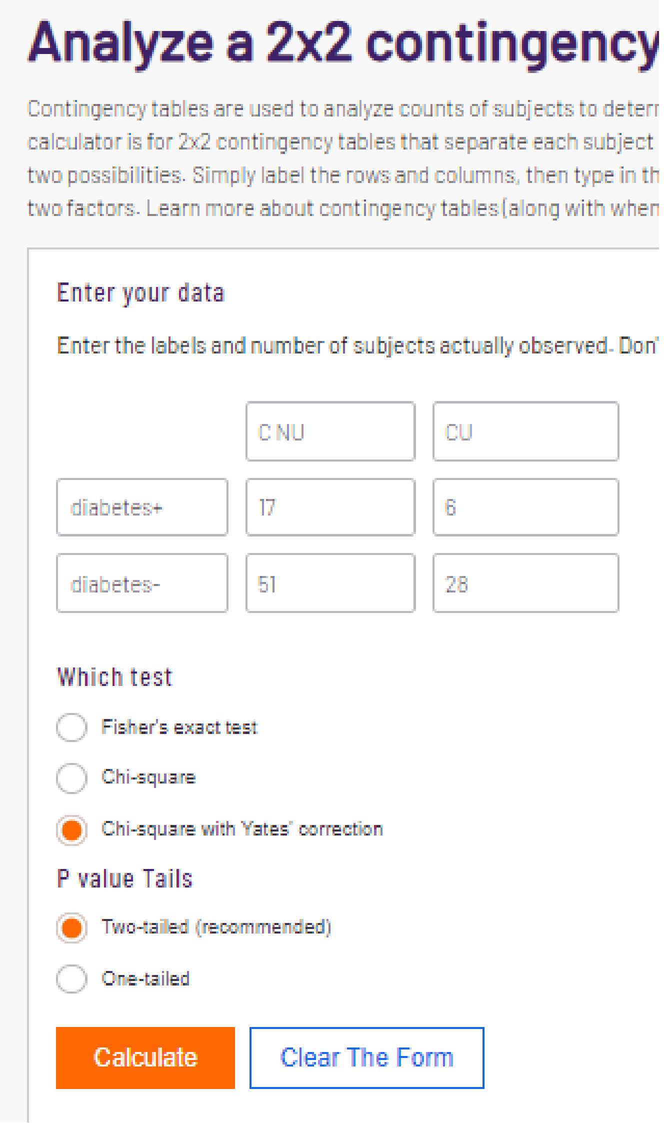


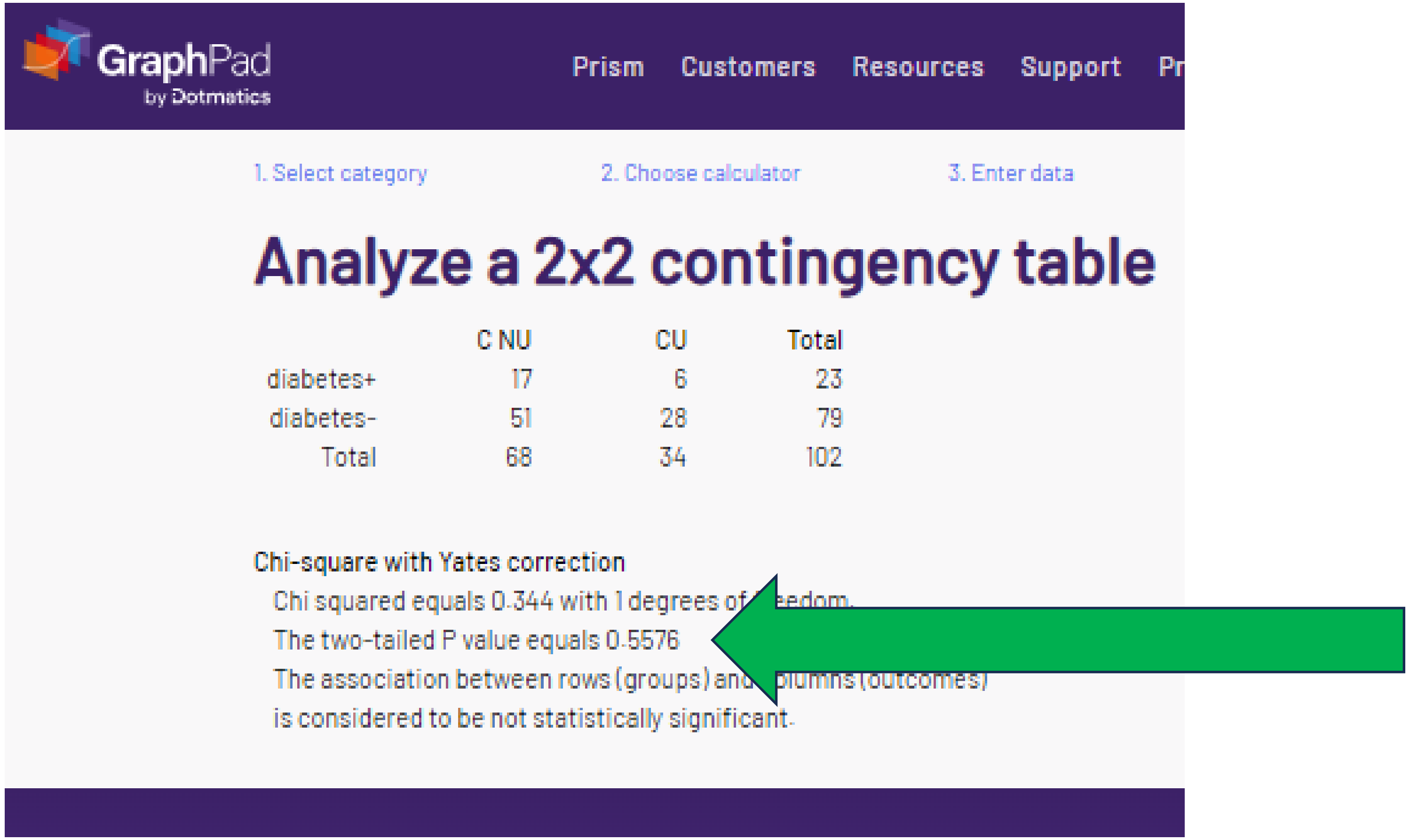


The smallest cell = 6 so Yates is **not** appropriate (Wikipedia convention).
The p values is reproduced with an incorrect statistic (not in methods).

### H. “corrected” chi-sq or Fishers p = .3612

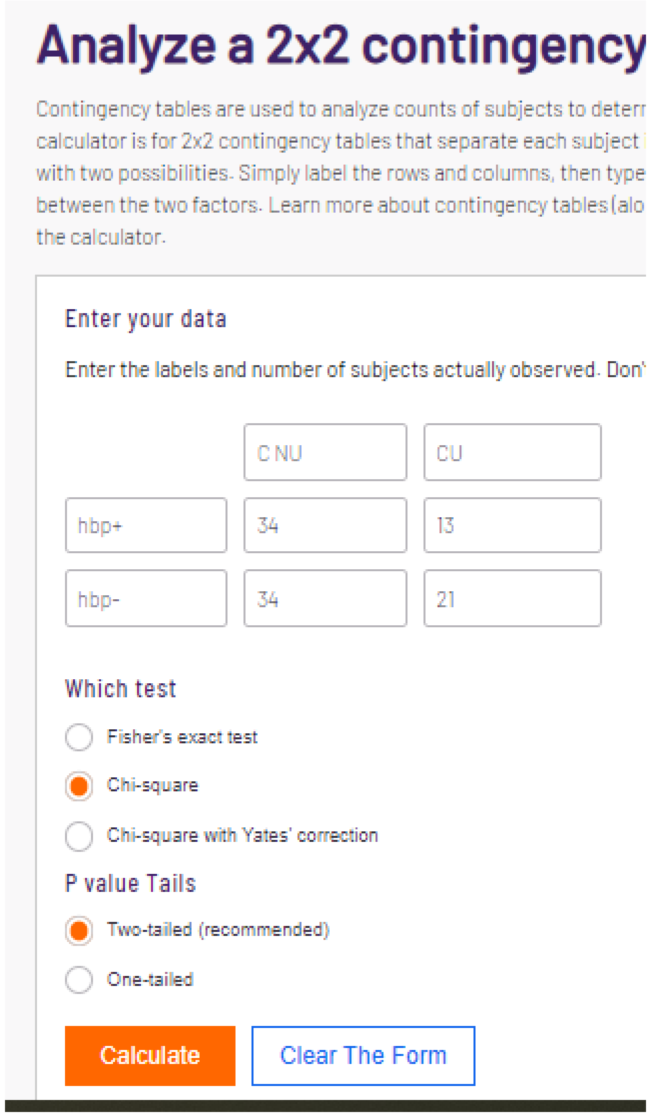


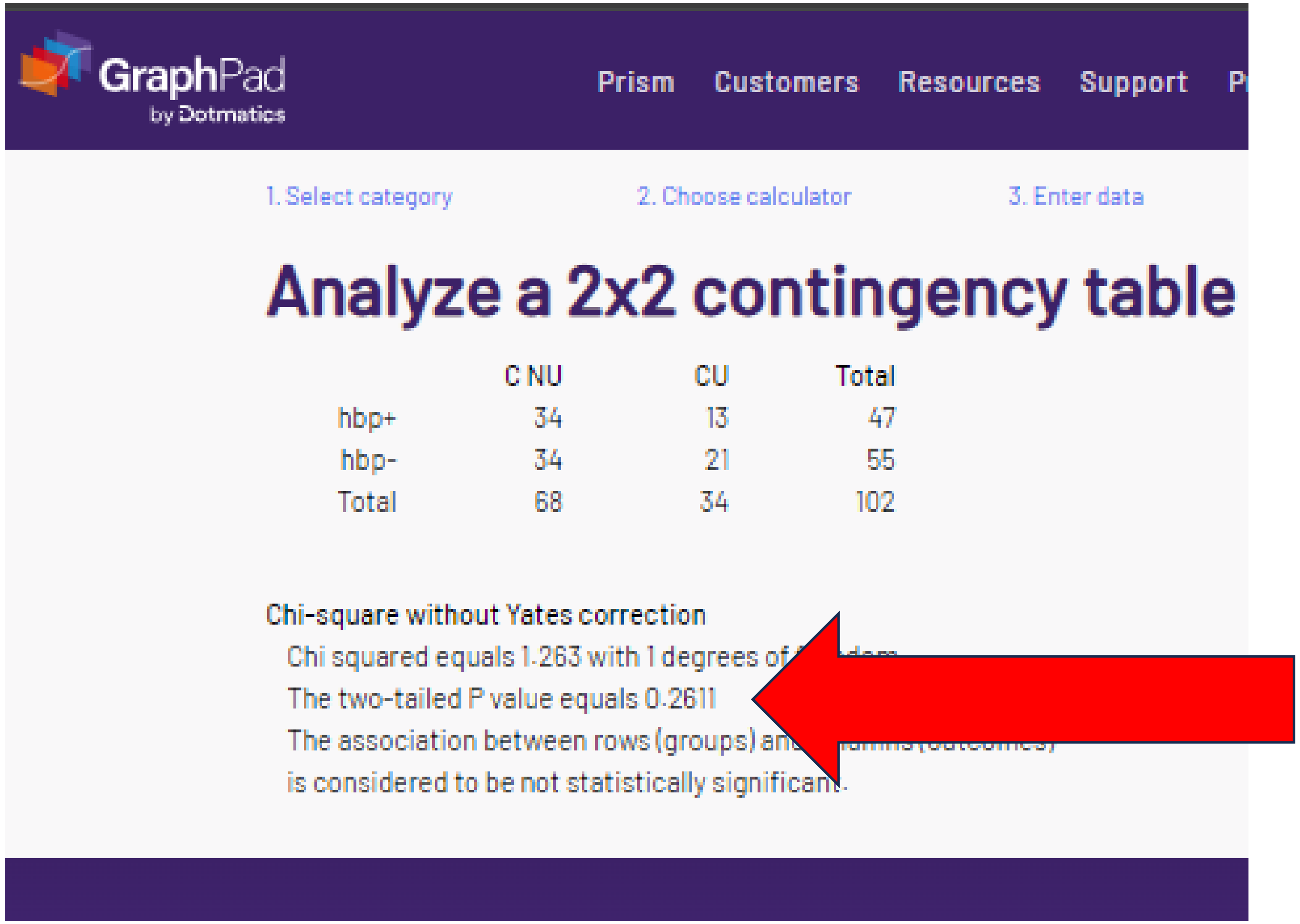


Note large (>12) N per cell.

### H. “corrected” chi-sq or Fishers p = .3612

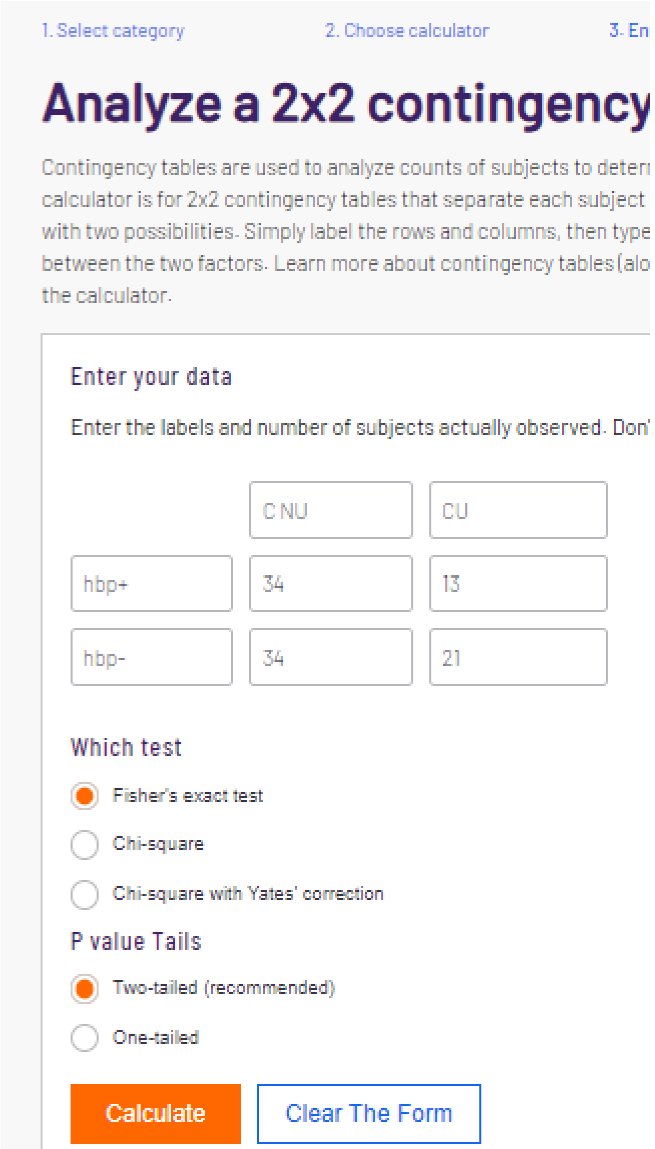


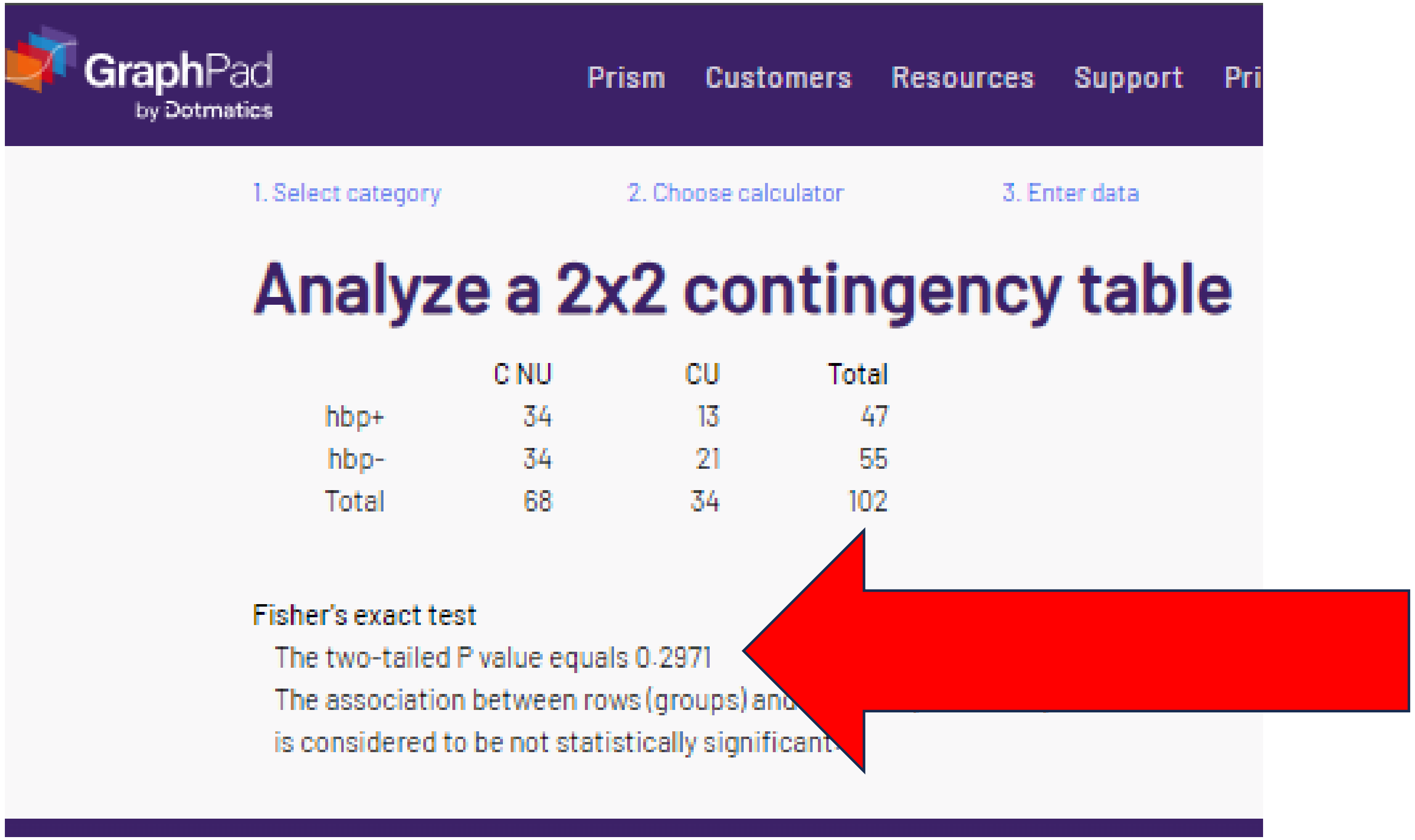


The analysis is **not** verified for chi-square or Fisher’s for high blood pressure.

### H. “corrected” chi-sq or Fishers p = .3612

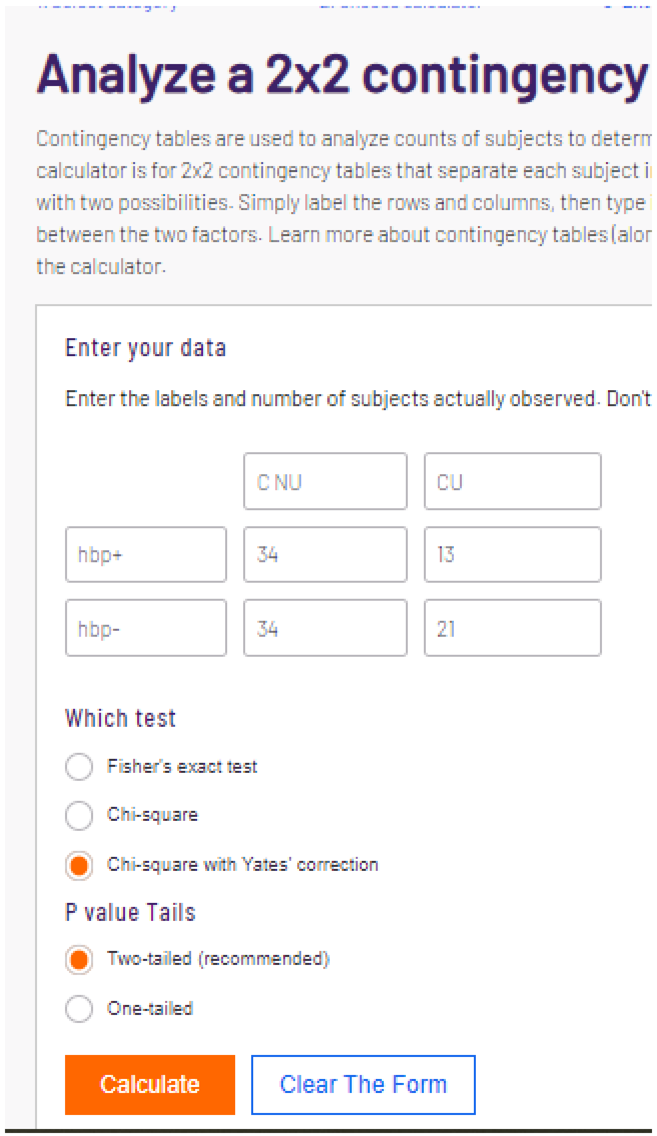


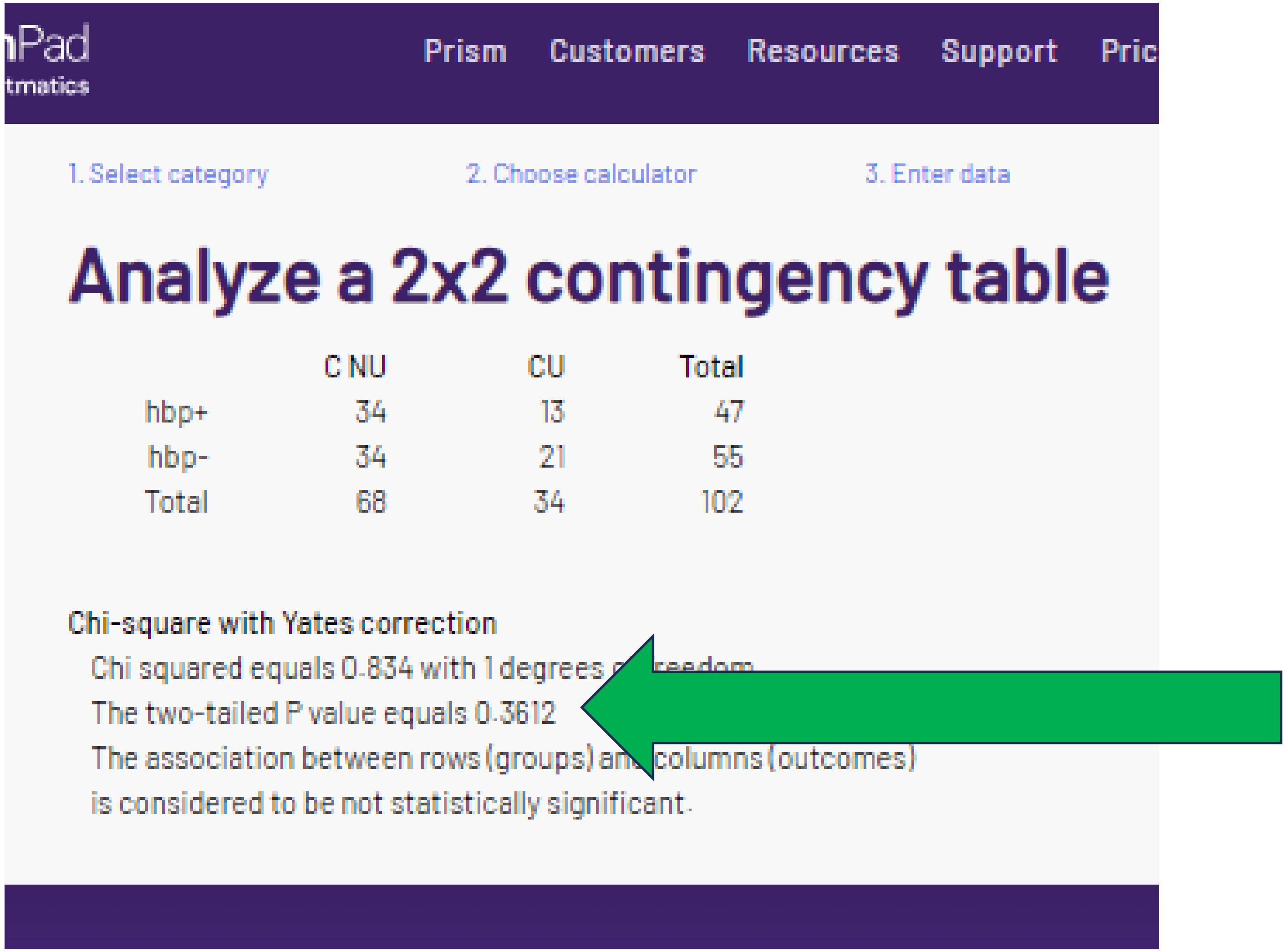


The smallest hypertension cell = 13 so there was **no reason** to run Yates. However, the p value was reproduced (with an incorrect statistic).

### I. “corrected” chi-sq or Fishers p = “1”

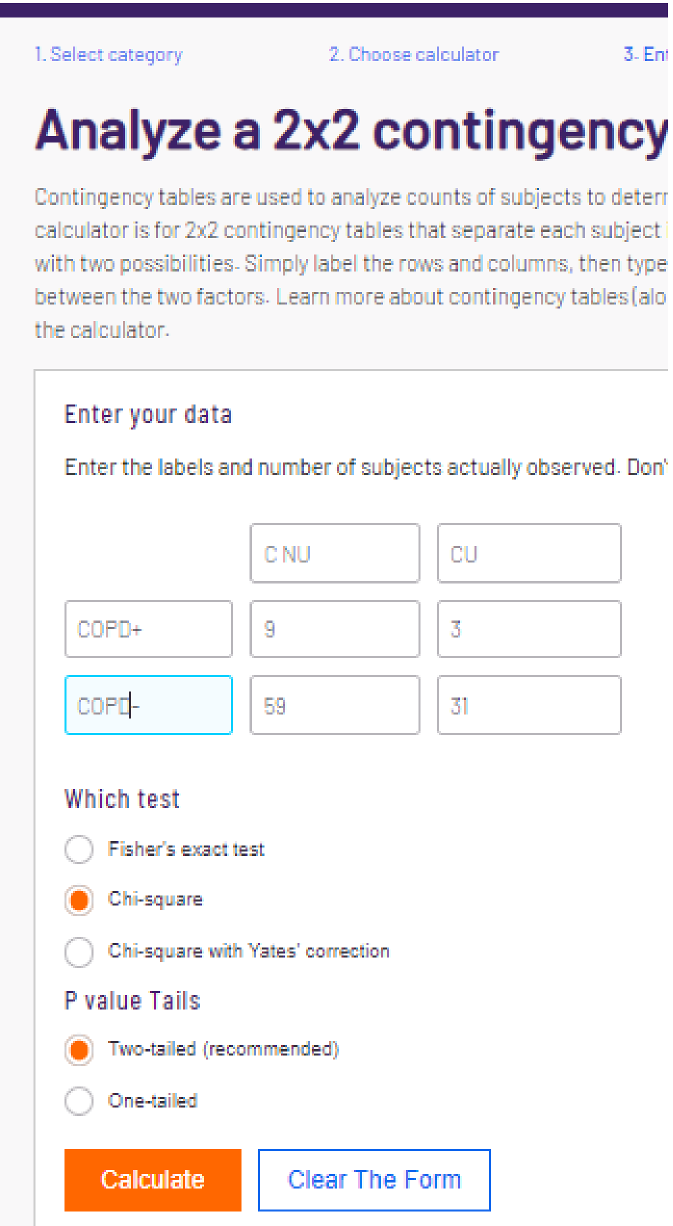


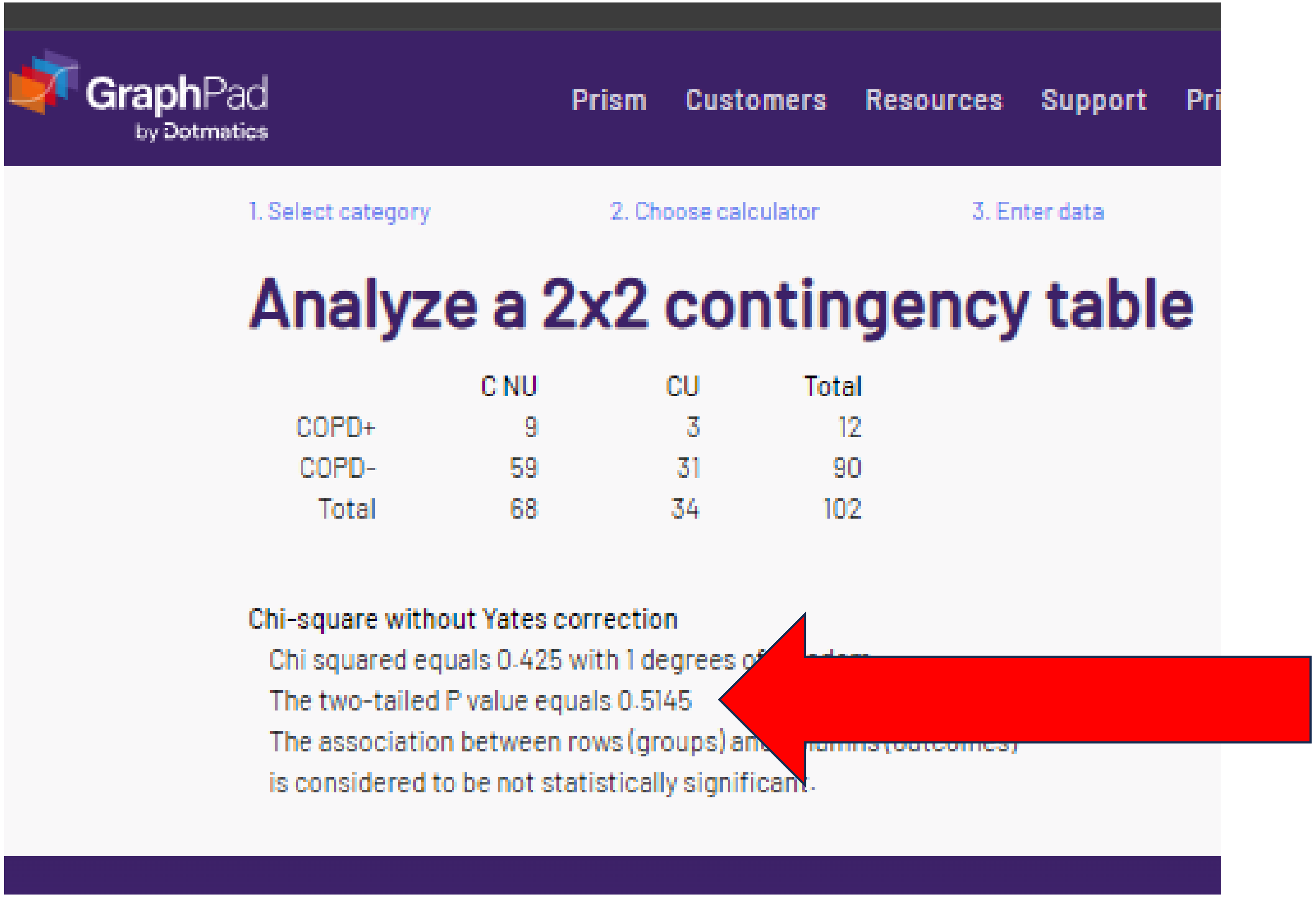

### I. "corrected" chi-sq or Fishers p = "1"

1. Select category 2. Choose calculator 3. Er

#### Analyze a 2x2 contingency

Contingency tables are used to analyze counts of subjects to deter
calculator is for 2x2 contingency tables that separate each subject
with two possibilities. Simply label the rows and columns, then typ
between the two factors. Learn more about contingency tables (al
the calculator.

**Enter your data**

Enter the labels and number of subjects actually observed. Dor

| | | |
|---|---|---|
| | C NU | CU |
| COPD+ | 9 | 3 |
| COPD- | 59 | 31 |

**Which test**

- (●) Fisher's exact test
- ( ) Chi-square
- ( ) Chi-square with Yates' correction

**P value Tails**

- (●) Two-tailed (recommended)
- ( ) One-tailed

Calculate | Clear The Form

GraphPad by Dotmatics — Prism Customers Resources Support Pric

1. Select category 2. Choose calculator 3. Enter data

#### Analyze a 2x2 contingency table

| | C NU | CU | Total |
|---|---|---|---|
| COPD+ | 9 | 3 | 12 |
| COPD- | 59 | 31 | 90 |
| Total | 68 | 34 | 102 |

**Fisher's exact test**

The two-tailed P value equals 0.7463

The association between rows (groups) and columns (outcomes)
is considered to be not statistically significant.

### I. "corrected" chi-sq or Fishers p = "1"

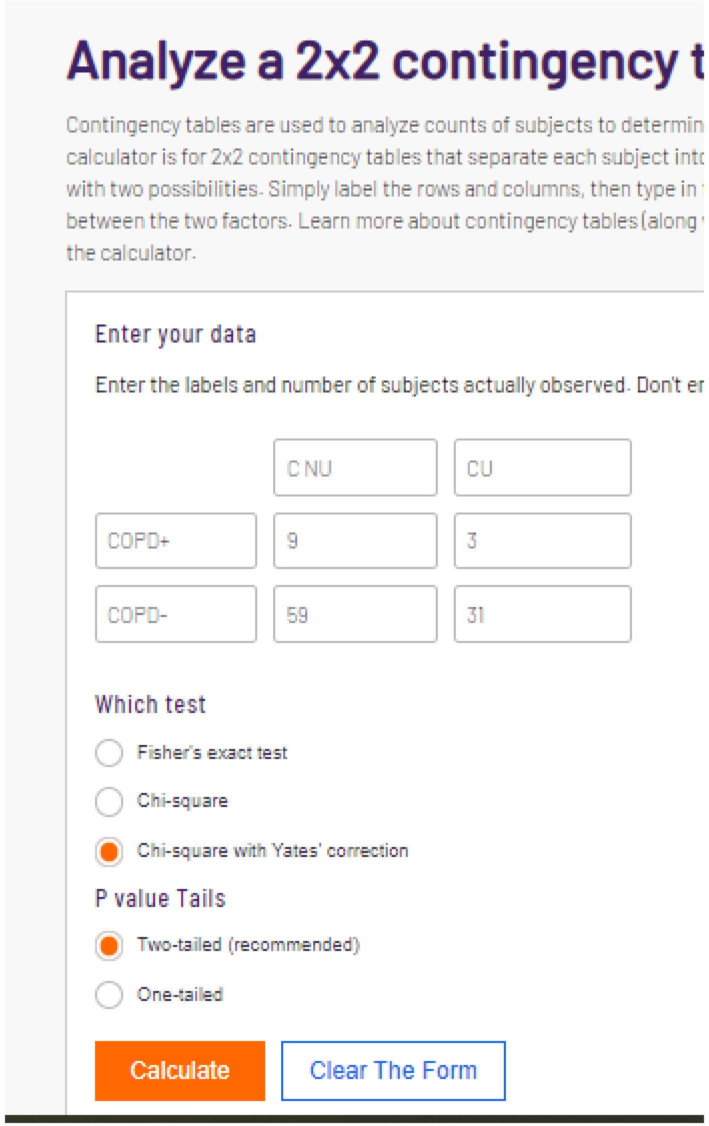

#### Analyze a 2x2 contingency t

Contingency tables are used to analyze counts of subjects to determin
calculator is for 2x2 contingency tables that separate each subject int
with two possibilities. Simply label the rows and columns, then type in
between the two factors. Learn more about contingency tables (along
the calculator.

Enter your data

Enter the labels and number of subjects actually observed. Don't er

| | | |
|---|---|---|
| | C NU | CU |
| COPD+ | 9 | 3 |
| COPD- | 59 | 31 |

Which test

- Fisher's exact test
- Chi-square
- Chi-square with Yates' correction

P value Tails

- Two-tailed (recommended)
- One-tailed

Calculate | Clear The Form

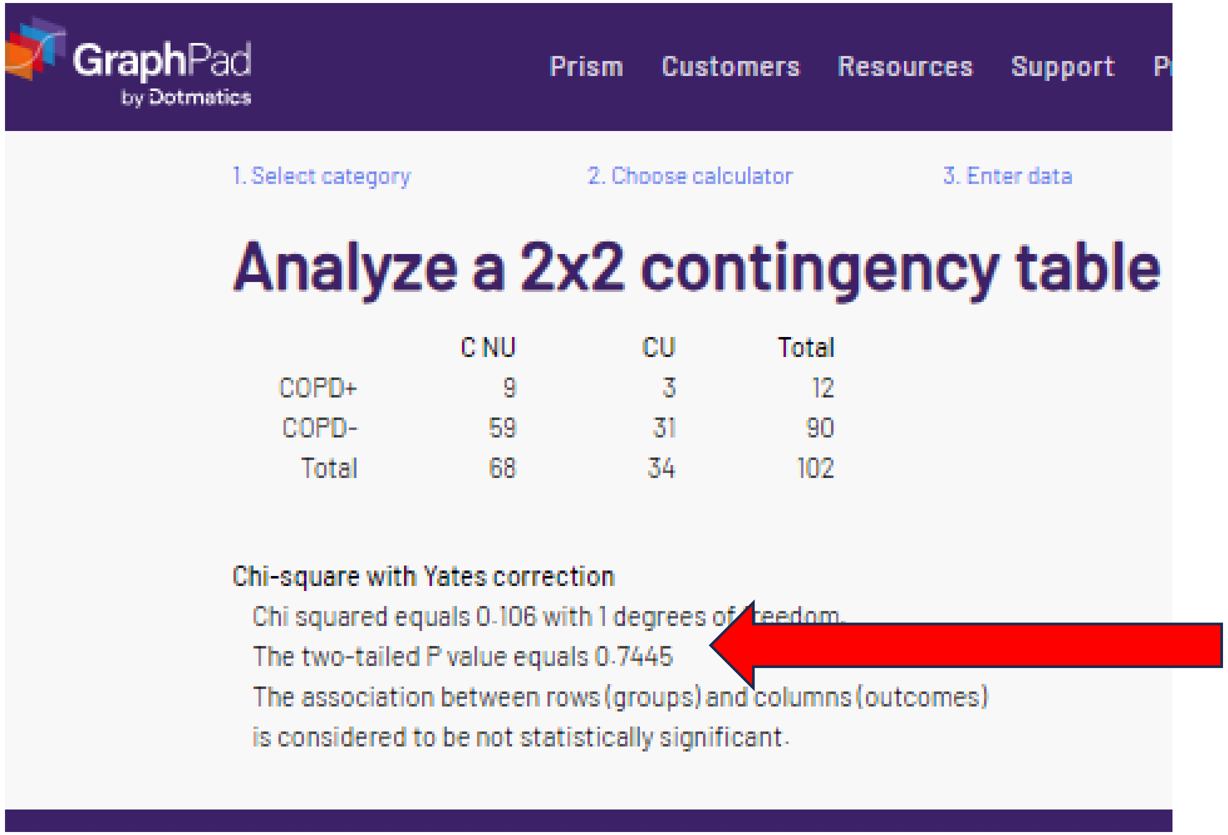

GraphPad by Dotmatics | Prism | Customers | Resources | Support | P

1. Select category | 2. Choose calculator | 3. Enter data

#### Analyze a 2x2 contingency table

| | C NU | CU | Total |
|---|---|---|---|
| COPD+ | 9 | 3 | 12 |
| COPD- | 59 | 31 | 90 |
| Total | 68 | 34 | 102 |

Chi-square with Yates correction

Chi squared equals 0.106 with 1 degrees of freedom.
The two-tailed P value equals 0.7445
The association between rows (groups) and columns (outcomes)
is considered to be not statistically significant.

The reported p value for COPD was **not verified** with either chi-square, Fisher's, or Yates. Note that for the small (< 5) values per cell, many statisticians would argue that Yates would be an **appropriate** analysis here. For p to = 1.000, the proportion would have to be identical in both groups. If 9 per total 68 in Cannabis Non Users, then 4.5 per 34 in Cannabis Users. Obviously, this is an impossible N so there is some error in reporting.

### J. "corrected" chi-sq or Fishers p = .2491

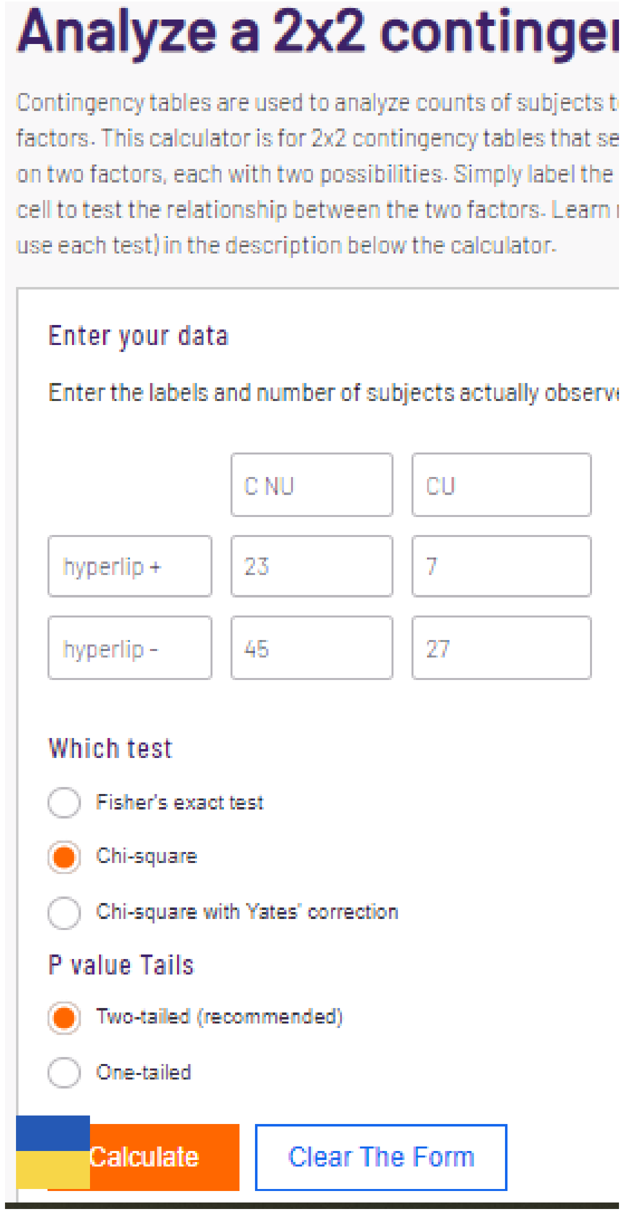

#### Analyze a 2x2 continge

Contingency tables are used to analyze counts of subjects t
factors. This calculator is for 2x2 contingency tables that se
on two factors, each with two possibilities. Simply label the
cell to test the relationship between the two factors. Learn
use each test) in the description below the calculator.

Enter your data

Enter the labels and number of subjects actually observ

| | C NU | CU |
|---|---|---|
| hyperlip + | 23 | 7 |
| hyperlip - | 45 | 27 |

Which test

- Fisher's exact test
- Chi-square
- Chi-square with Yates' correction

P value Tails

- Two-tailed (recommended)
- One-tailed

Calculate | Clear The Form

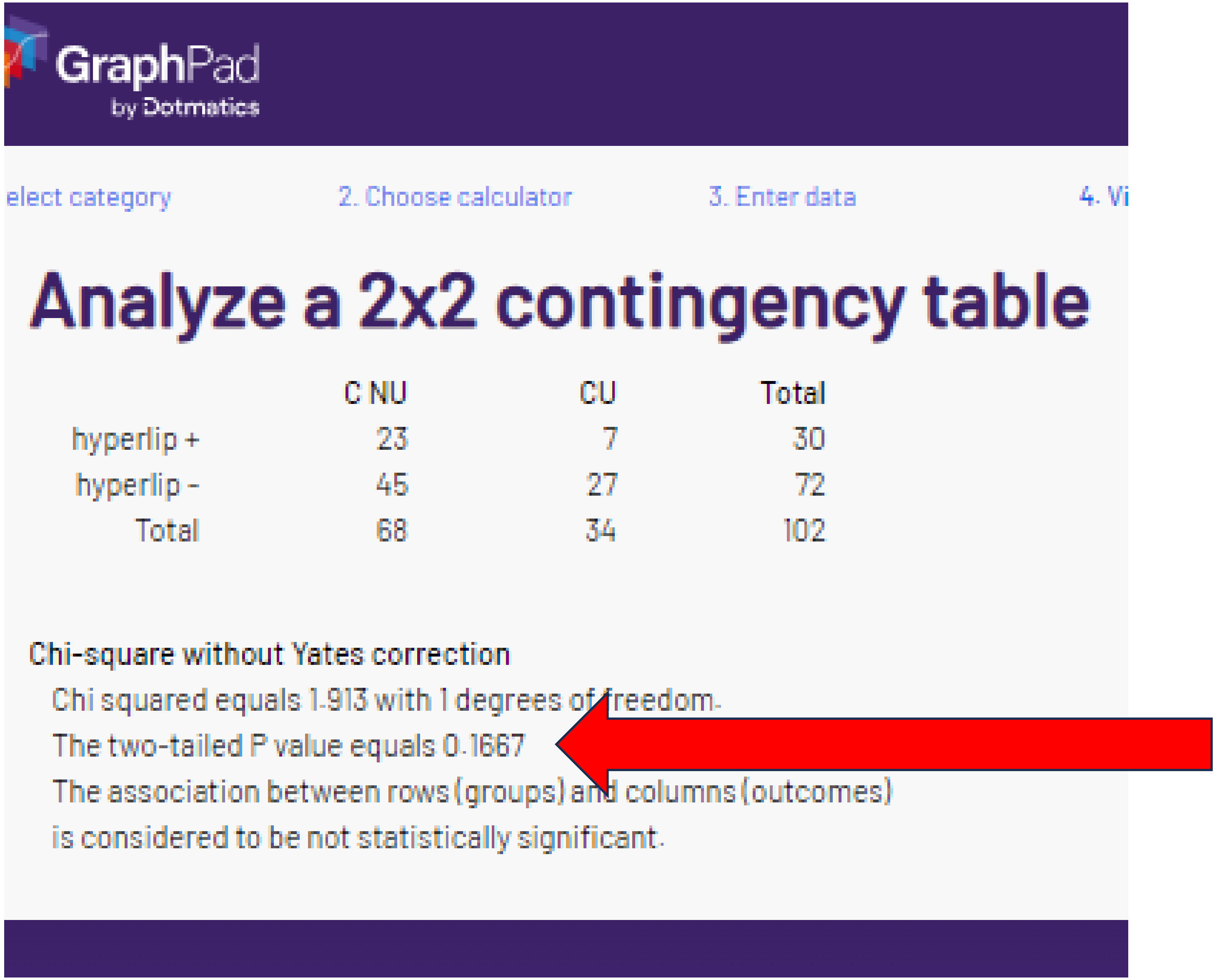

GraphPad by Dotmatics

elect category | 2. Choose calculator | 3. Enter data | 4. Vi

#### Analyze a 2x2 contingency table

| | C NU | CU | Total |
|---|---|---|---|
| hyperlip + | 23 | 7 | 30 |
| hyperlip - | 45 | 27 | 72 |
| Total | 68 | 34 | 102 |

Chi-square without Yates correction

Chi squared equals 1.913 with 1 degrees of freedom.

The two-tailed P value equals 0.1667

The association between rows (groups) and columns (outcomes)

is considered to be not statistically significant.

### J. “corrected” chi-sq or Fishers p = .2491

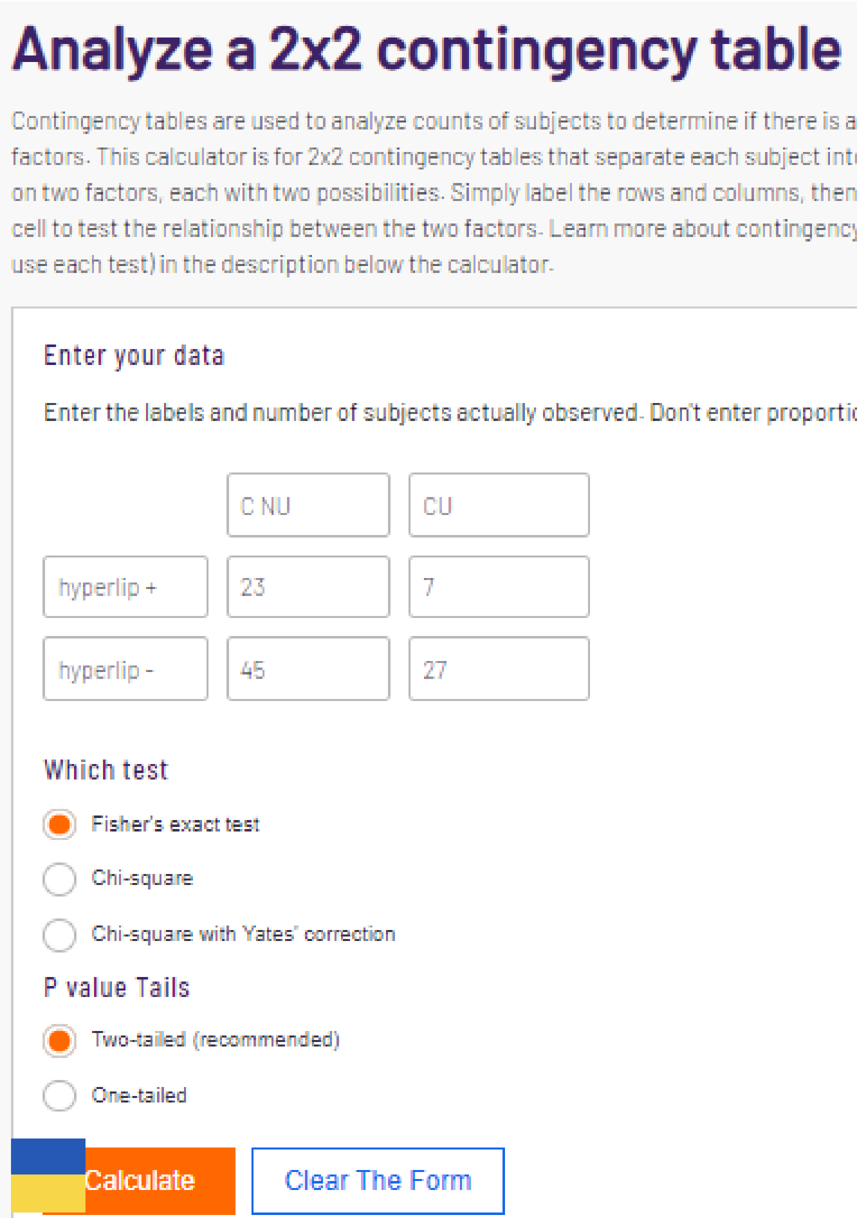


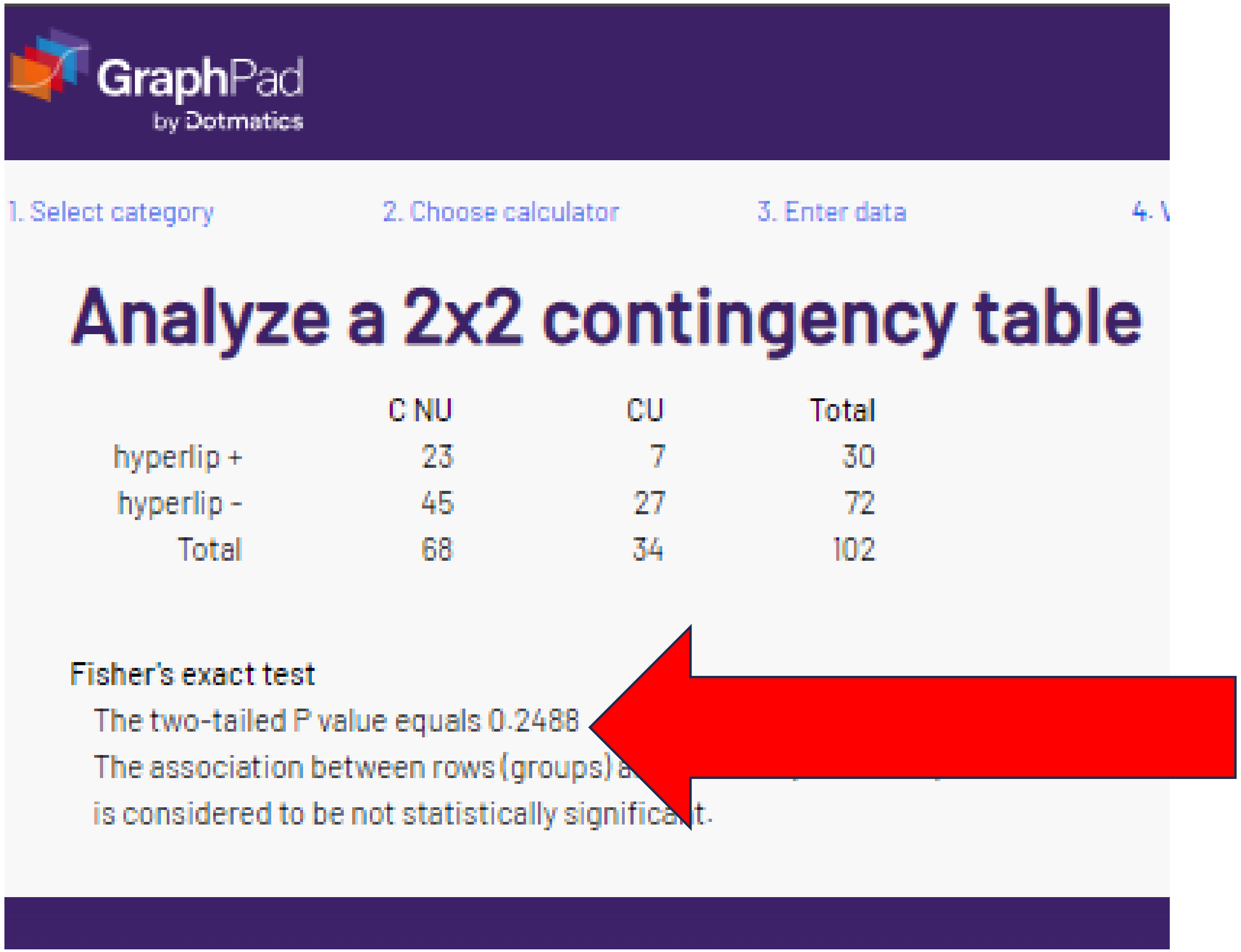

### J. “corrected” chi-sq or Fisher’s p = .2491

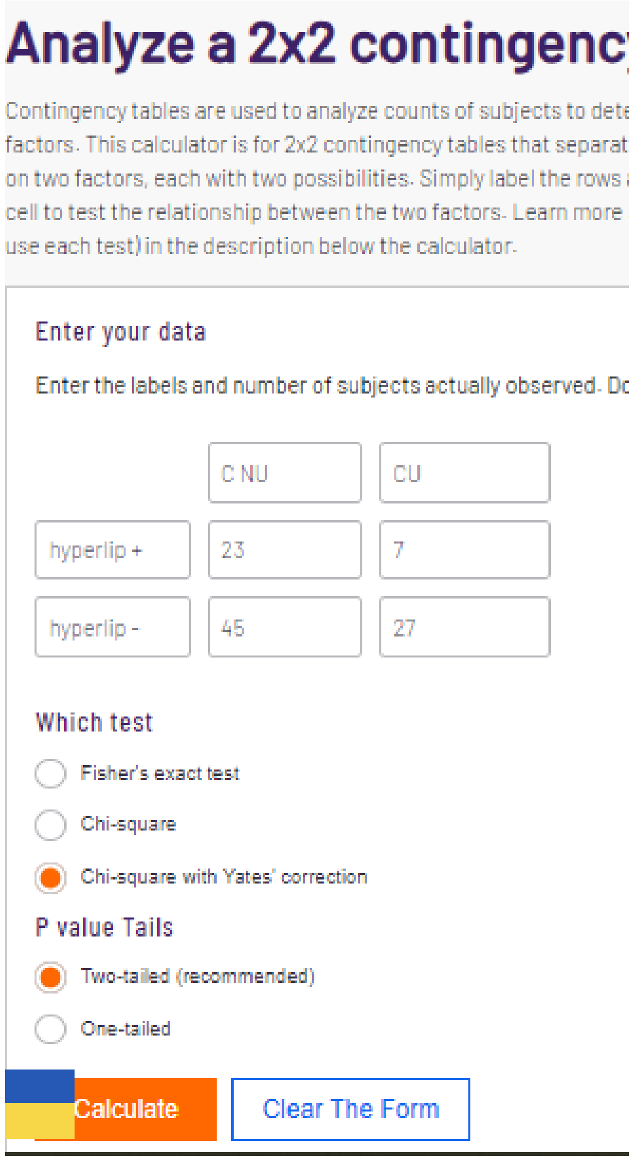


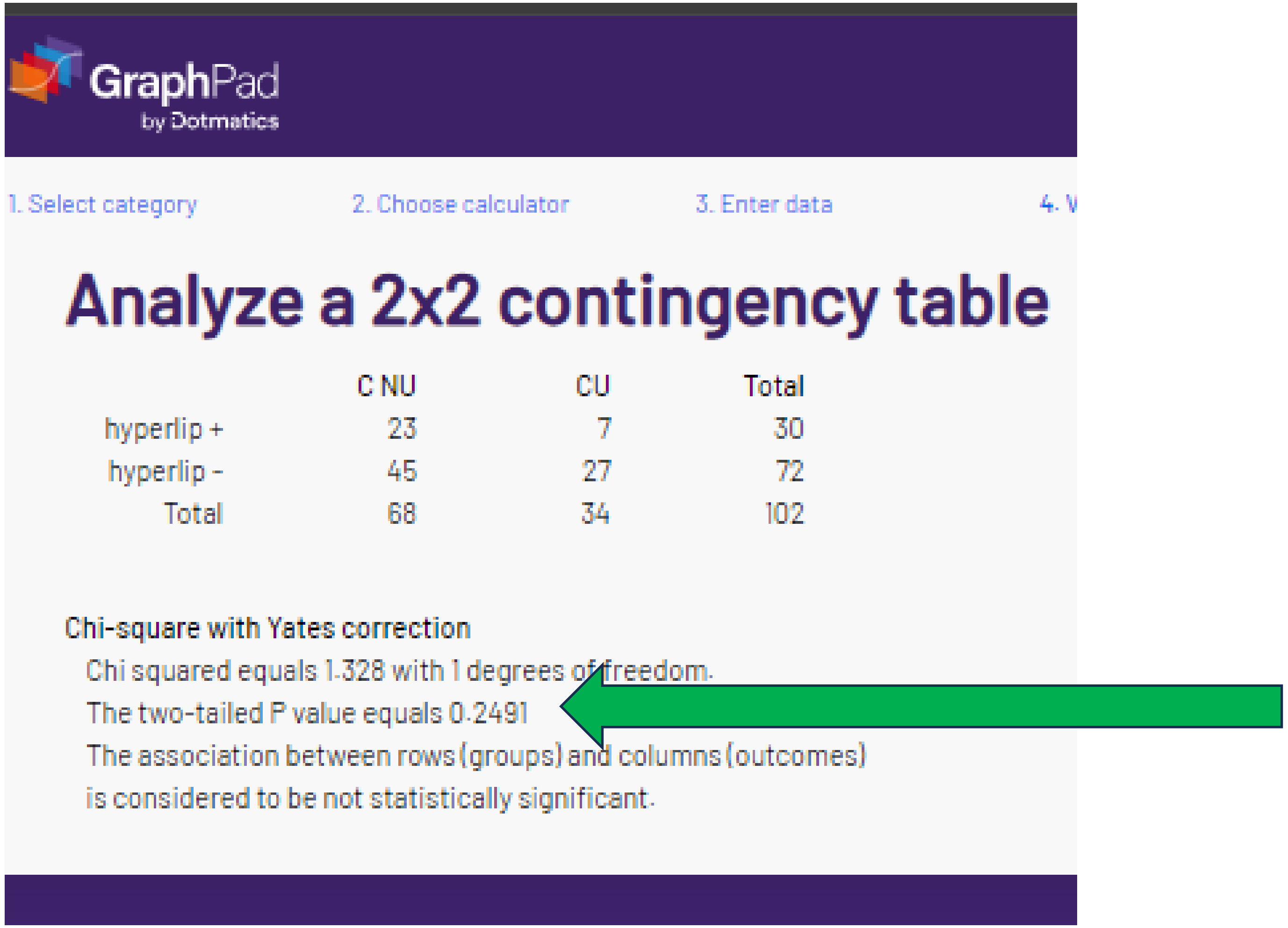


The smallest cell = 7 so Yates would not be appropriate for hyperlipidemia (Wiki criteria). However, the p value was reproduced (albeit with the wrong-not in methods, statistic).

### K. “corrected” chi-sq or Fisher’s p = “1”

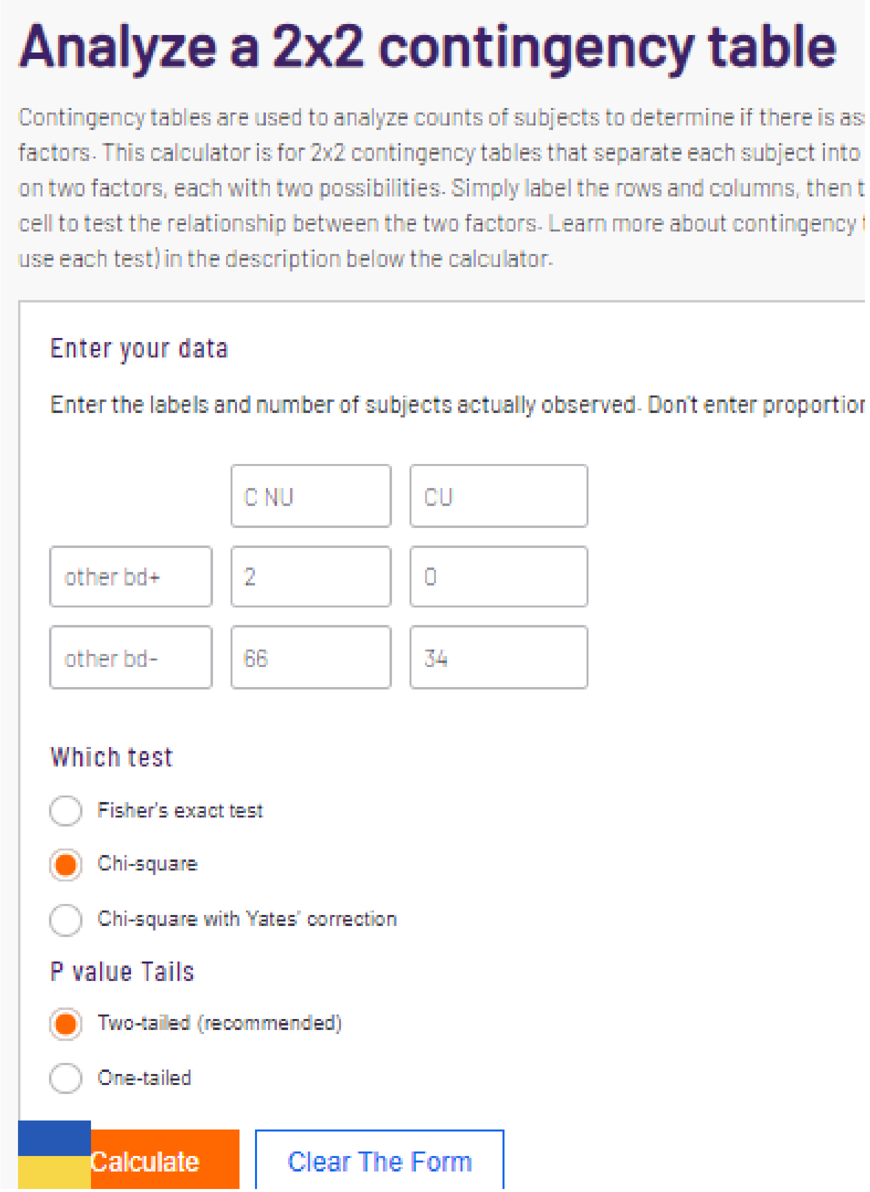


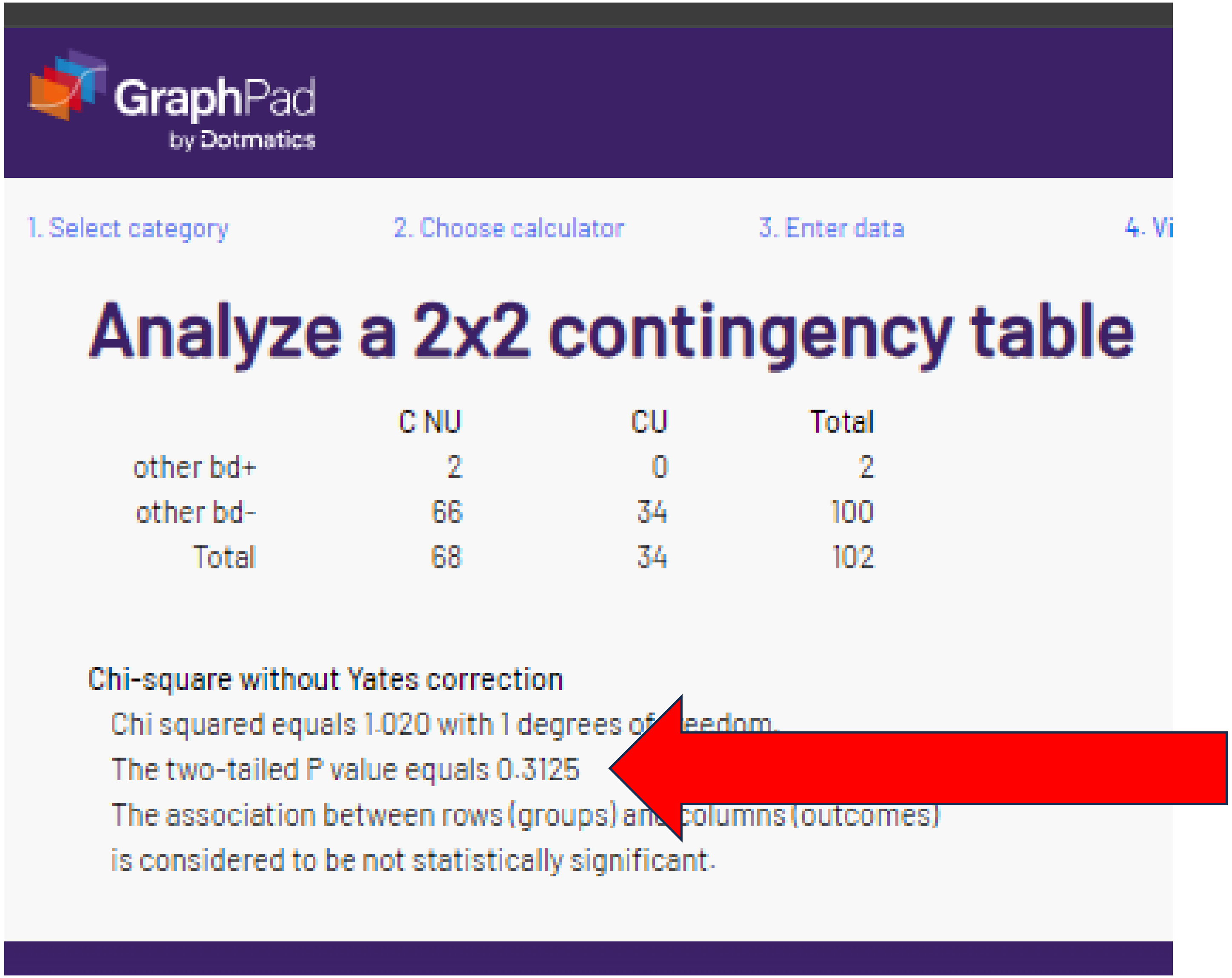


| | C NU | CU | Total |
|---|---|---|---|
| other bd+ | 2 | 0 | 2 |
| other bd- | 66 | 34 | 100 |
| Total | 68 | 34 | 102 |

Chi-square without Yates correction

Chi squared equals 1.020 with 1 degrees of [illegible]eedom.

The two-tailed P value equals 0.3125

The association between rows (groups) an[illegible] columns (outcomes)

is considered to be not statistically significant.

### K. "corrected" chi-sq or Fisher's p = "1"

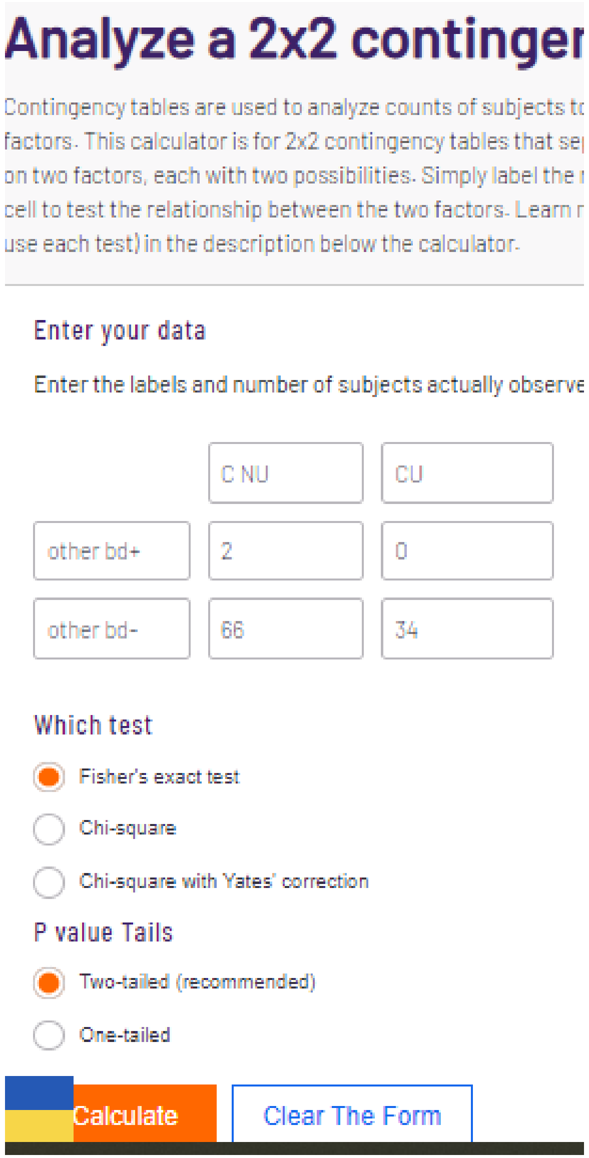


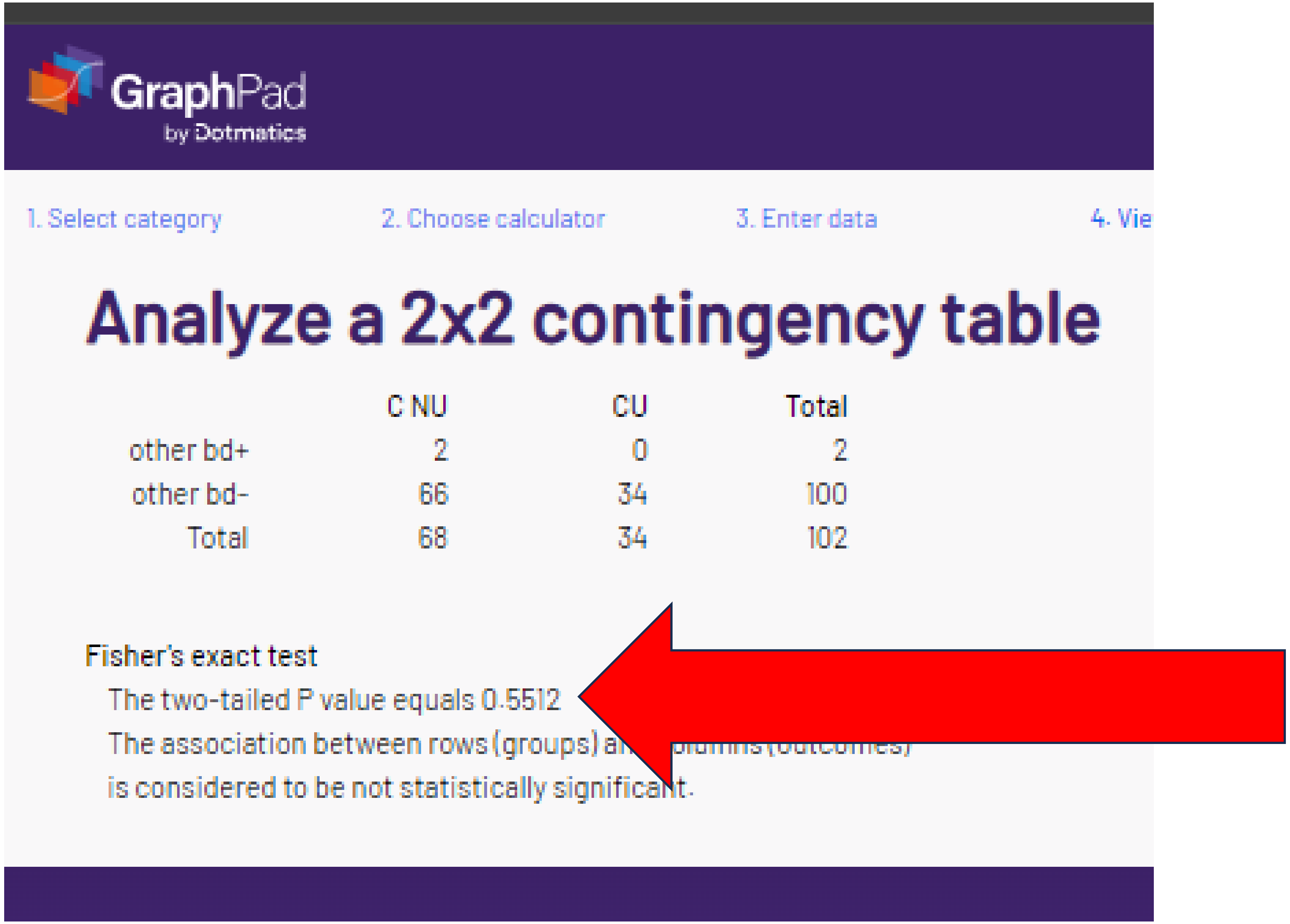


For other background diseases, the "corrected" reported p-value was not verified with either Chi-square or Fisher's.

### K. “corrected” chi-sq or Fishers p = “1”

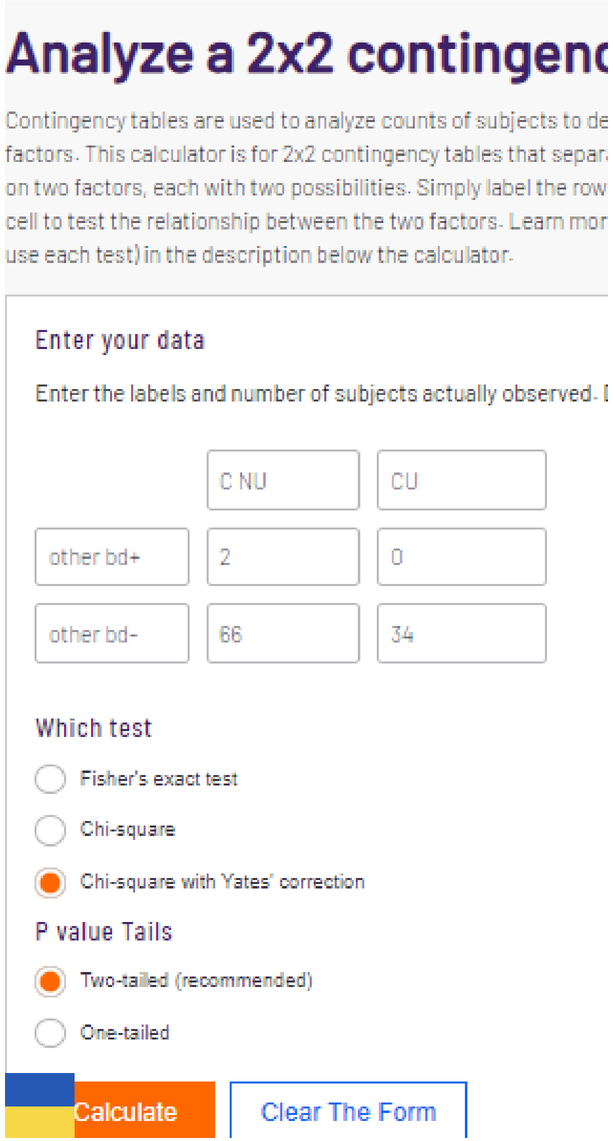


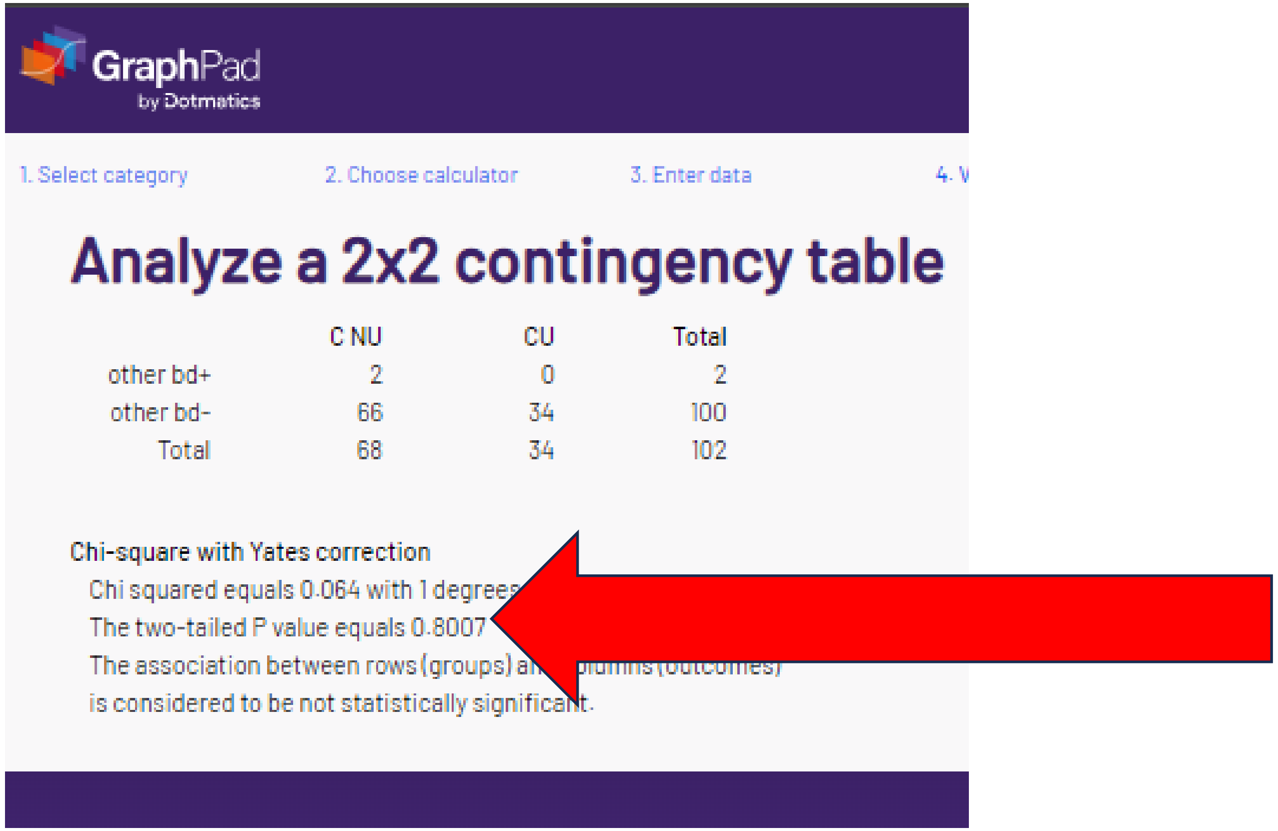


For other background diseases, the reported p-value was not verified with either chi-square, Fisher’s, or Yates. For p = 1.000, the % would have to be identical for both groups. If 2 per 68 total for CNU, then 1 per 34 for the CU group. As this was not listed, there appears to be an error in the original data.

### L. “corrected” chi-sq or Fishers p = .8325

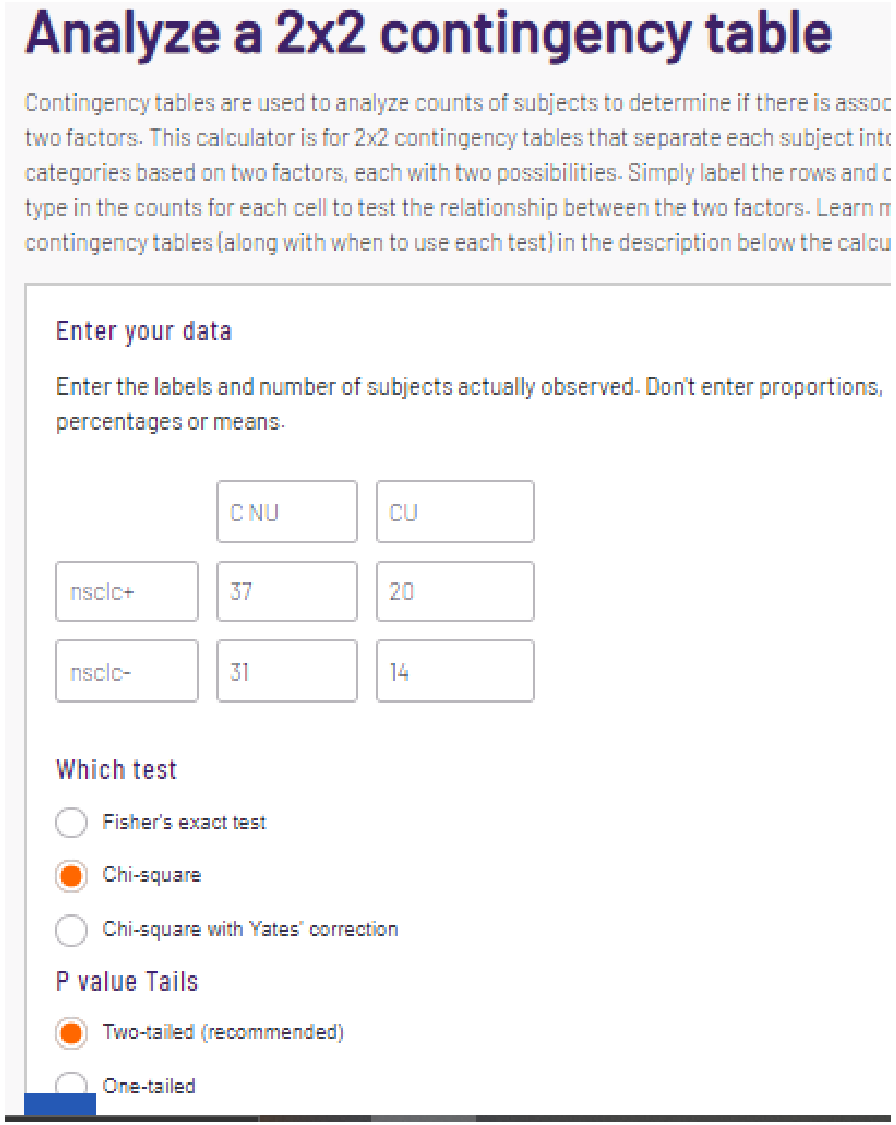


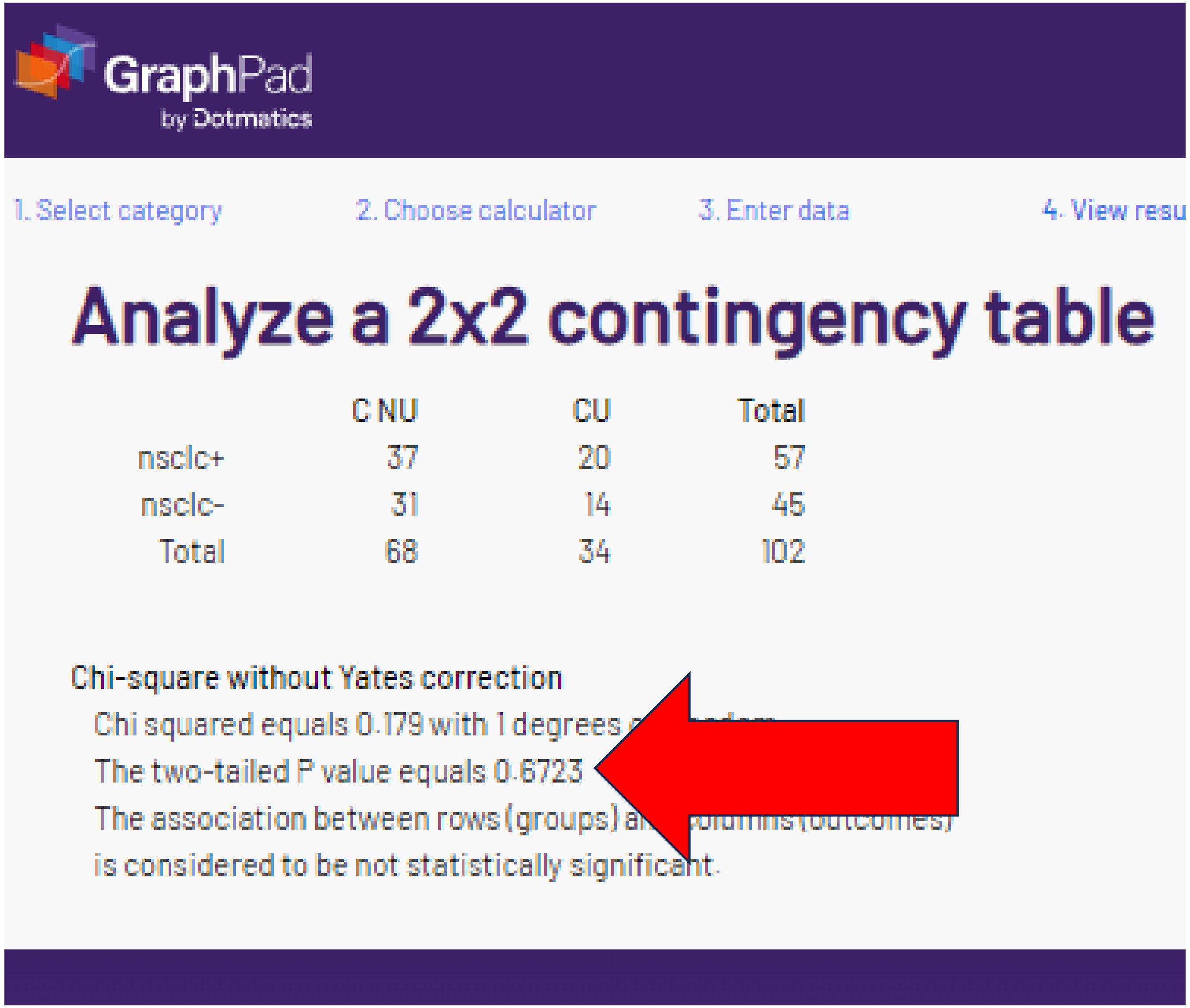

### L. “corrected” chi-sq or Fishers p = .8325

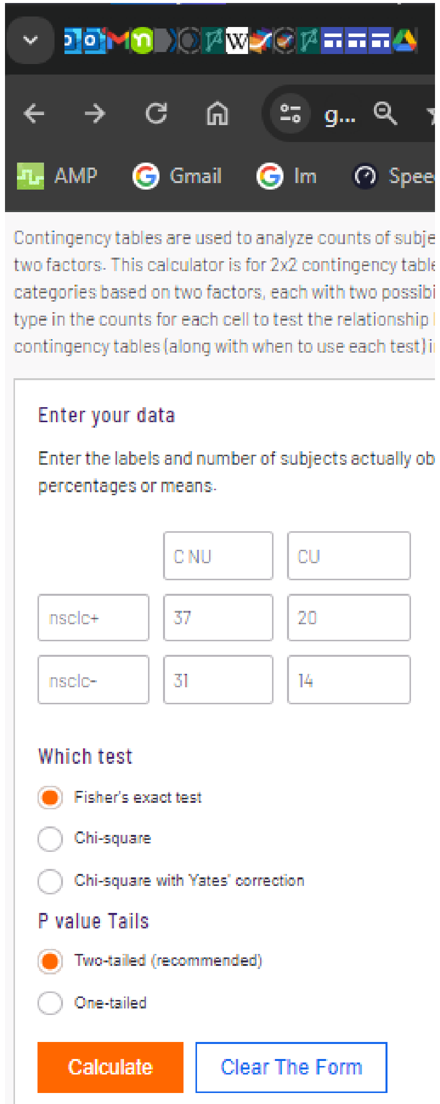


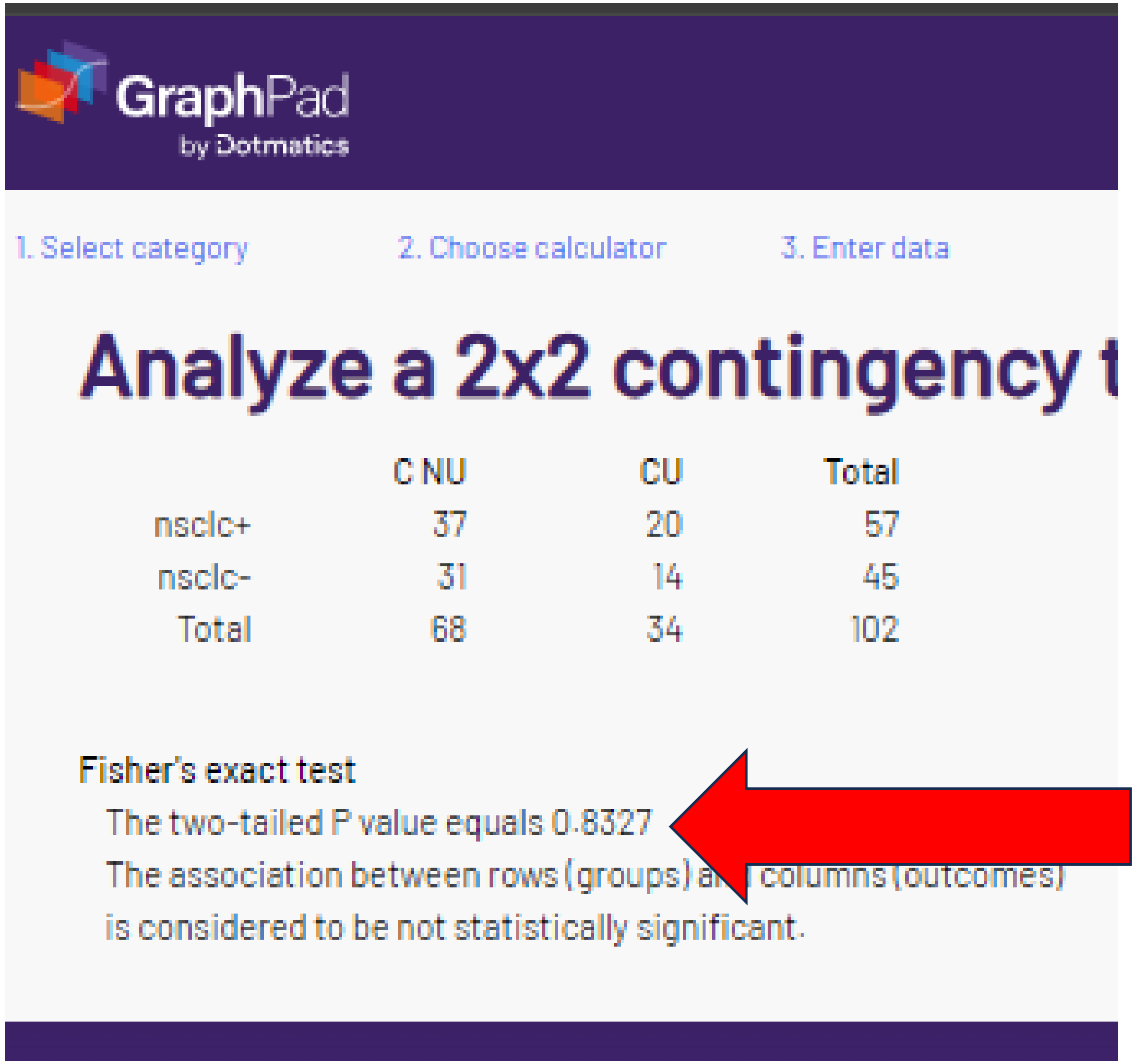


Not verified.

### L. “corrected” chi-sq or Fishers p = .8325

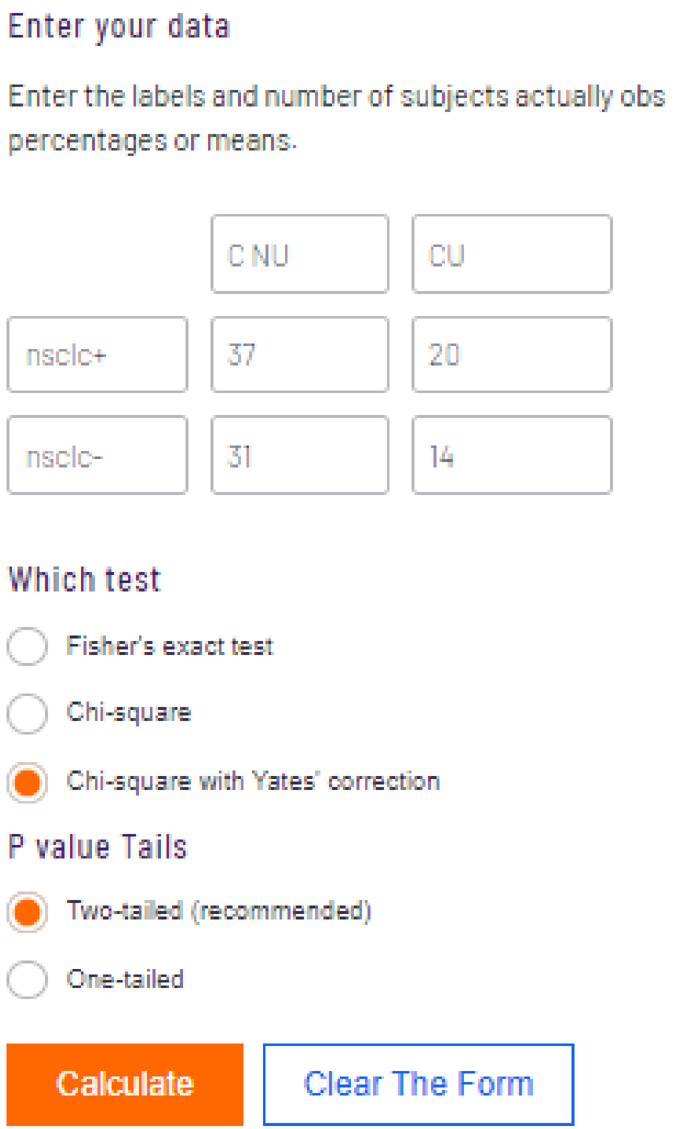

#### Enter your data

Enter the labels and number of subjects actually obs percentages or means.

| | | |
|---|---|---|
| | C NU | CU |
| nsclc+ | 37 | 20 |
| nsclc- | 31 | 14 |

#### Which test

- Fisher's exact test
- Chi-square
- Chi-square with Yates' correction (selected)

#### P value Tails

- Two-tailed (recommended) (selected)
- One-tailed

Calculate | Clear The Form

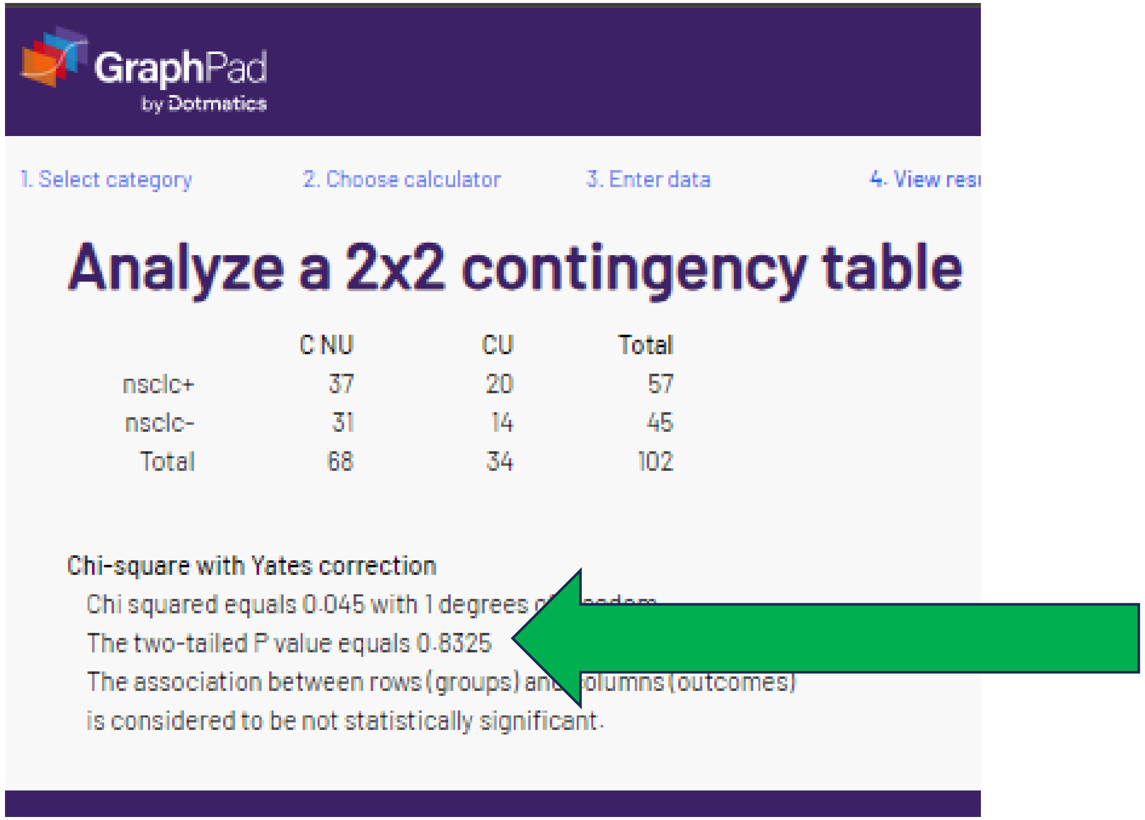

GraphPad by Dotmatics

1. Select category 2. Choose calculator 3. Enter data 4. View resu

#### Analyze a 2x2 contingency table

| | C NU | CU | Total |
|---|---|---|---|
| nsclc+ | 37 | 20 | 57 |
| nsclc- | 31 | 14 | 45 |
| Total | 68 | 34 | 102 |

**Chi-square with Yates correction**

Chi squared equals 0.045 with 1 degrees o[illegible]eedom.
The two-tailed P value equals 0.8325
The association between rows (groups) an[illegible]olumns (outcomes)
is considered to be not statistically significant.

With the smallest cell = 14, Yates is the **wrong** statistic.
However, the p-value is reproduced for non small cell lung cancer.

### M. “corrected” chi-sq or Fishers p = .414

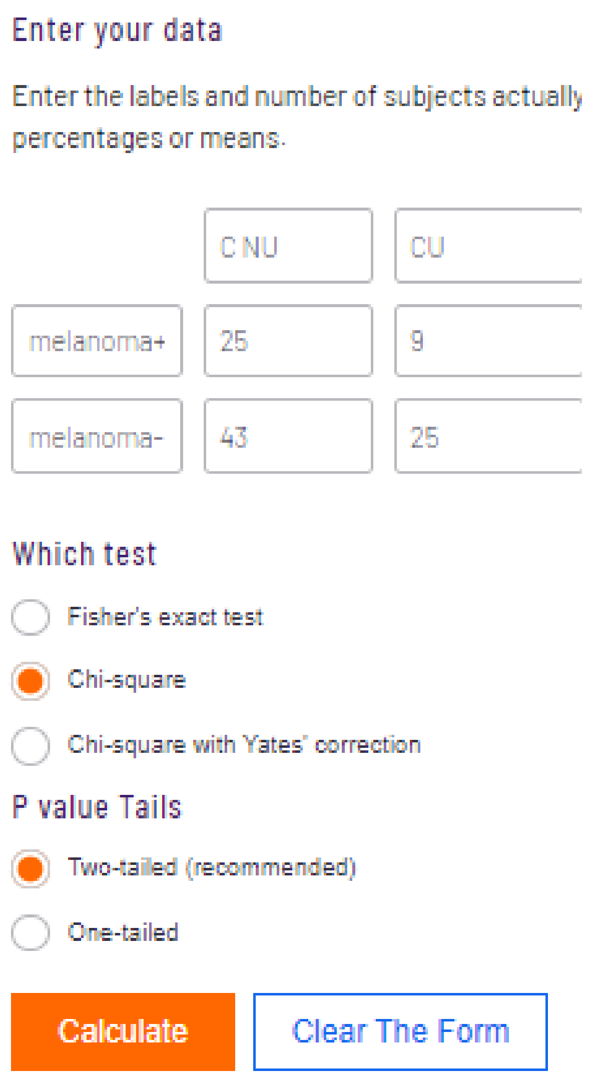


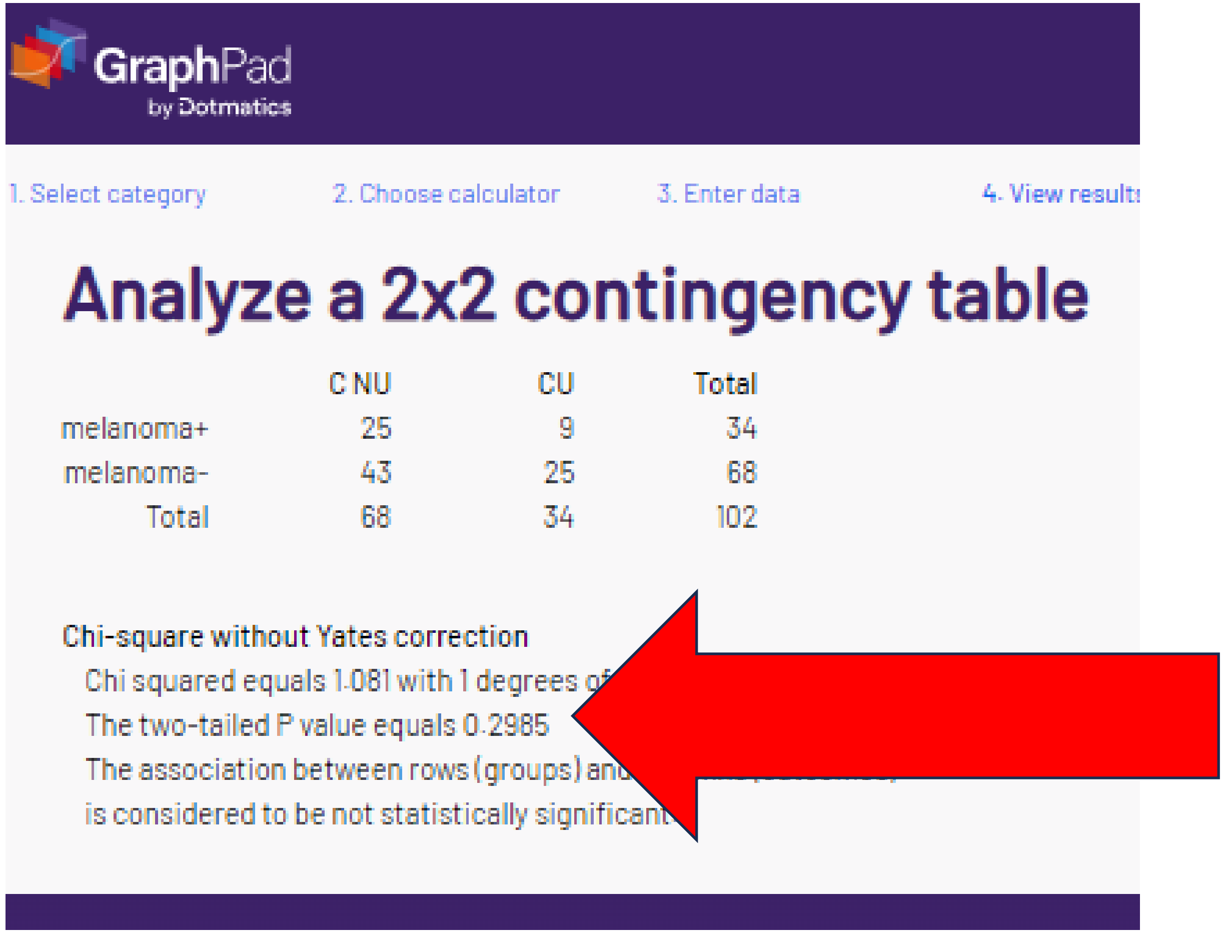

### M. “corrected” chi-sq or Fishers p = .414

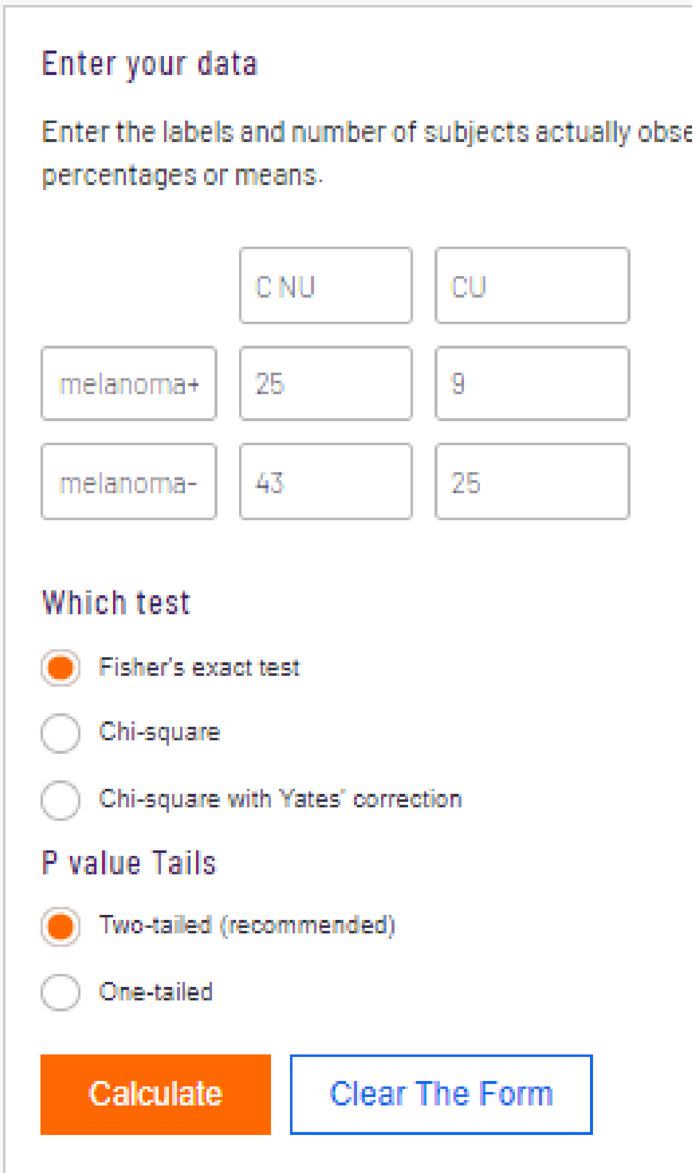
Enter your data

Enter the labels and number of subjects actually obse

percentages or means.

| | C NU | CU |
|---|---|---|
| melanoma+ | 25 | 9 |
| melanoma- | 43 | 25 |

Which test

Fisher's exact test

Chi-square

Chi-square with Yates' correction

P value Tails

Two-tailed (recommended)

One-tailed

Calculate

Clear The Form

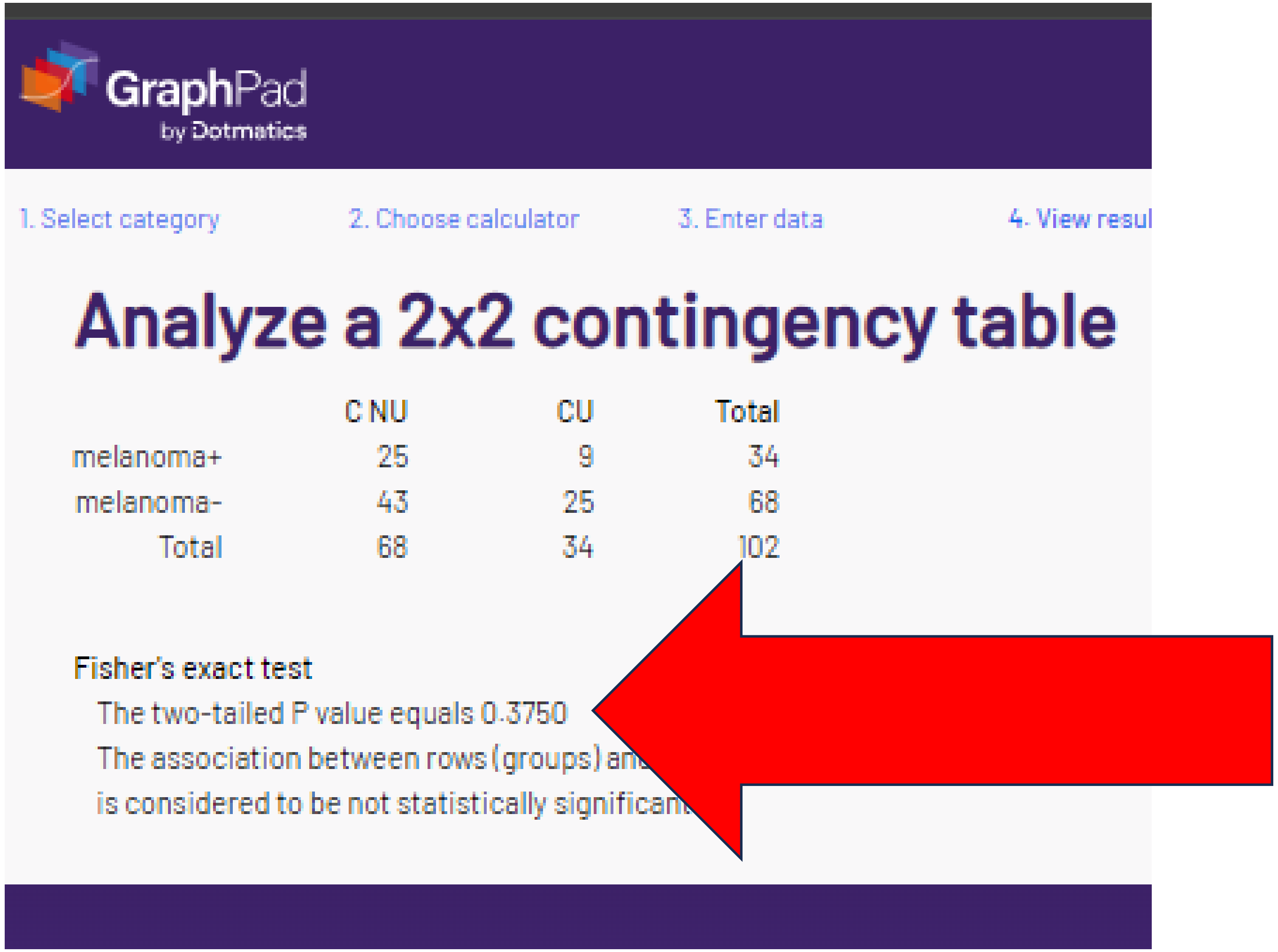

1. Select category 2. Choose calculator 3. Enter data 4. View resul

Analyze a 2x2 contingency table

| | C NU | CU | Total |
|---|---|---|---|
| melanoma+ | 25 | 9 | 34 |
| melanoma- | 43 | 25 | 68 |
| Total | 68 | 34 | 102 |

Fisher's exact test

The two-tailed P value equals 0.3750

The association between rows (groups) an

is considered to be not statistically significan

The reported “corrected” p values for melanoma were not verified with chi-square or Fisher’s.

### M. “corrected” chi-sq or Fishers p = .414

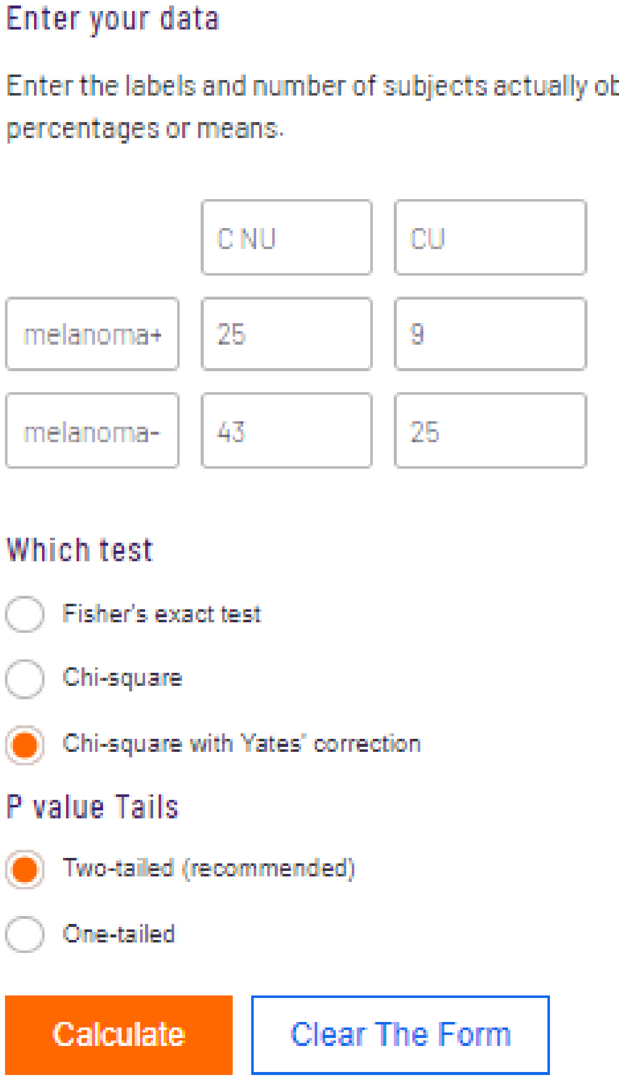


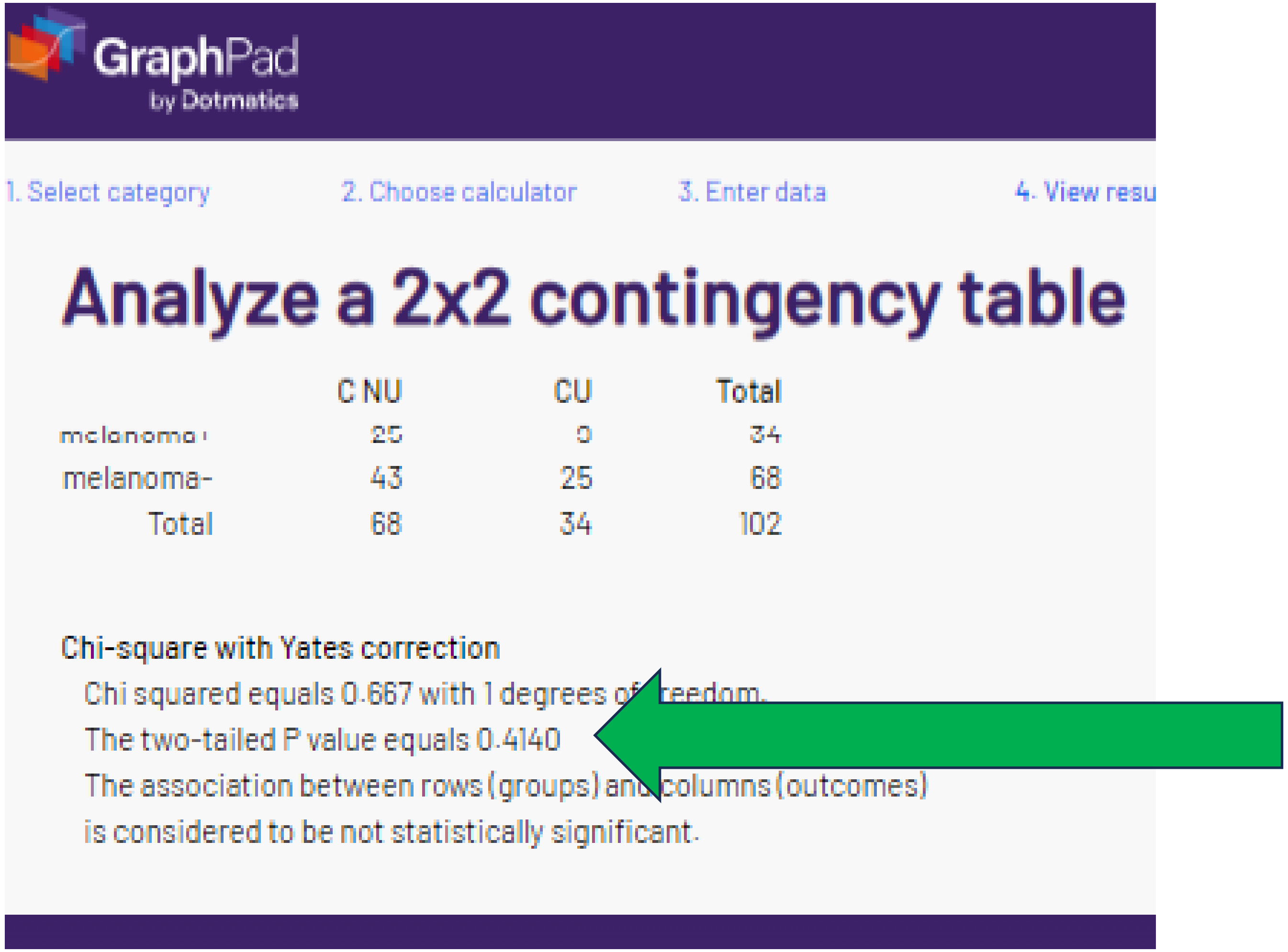


With a smallest N / cell of 9, Yates is an **inappropriate** statistic.
However, this corresponds with the p-value reported.

### N. “corrected” chi-sq or Fishers p = “1”

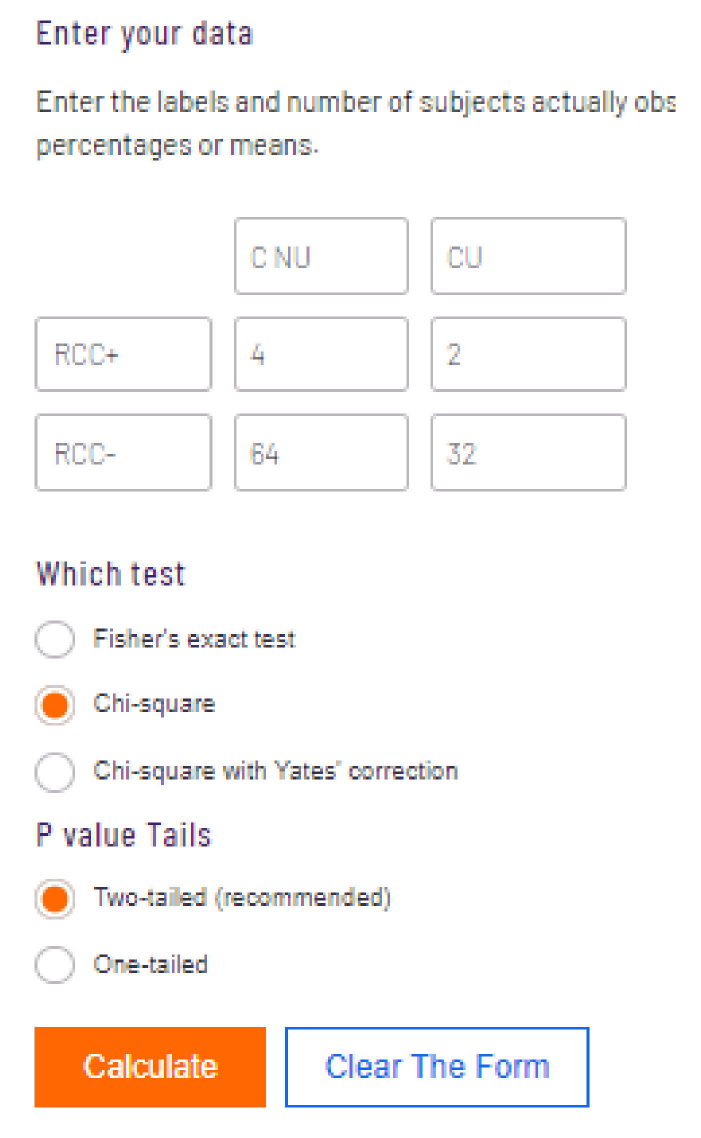


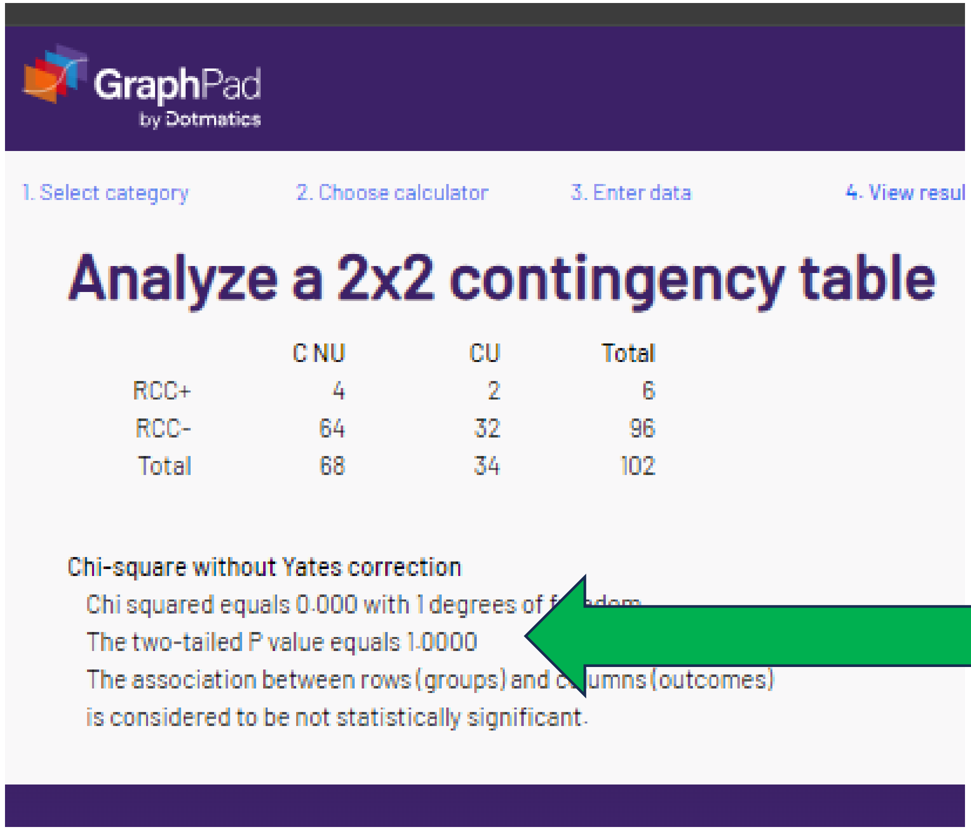


| | C NU | CU | Total |
|---|---|---|---|
| RCC+ | 4 | 2 | 6 |
| RCC- | 64 | 32 | 96 |
| Total | 68 | 34 | 102 |

Chi-square without Yates correction
Chi squared equals 0.000 with 1 degrees of f[illegible]dom
The two-tailed P value equals 1.0000
The association between rows (groups) and c[illegible]umns (outcomes)
is considered to be not statistically significant.

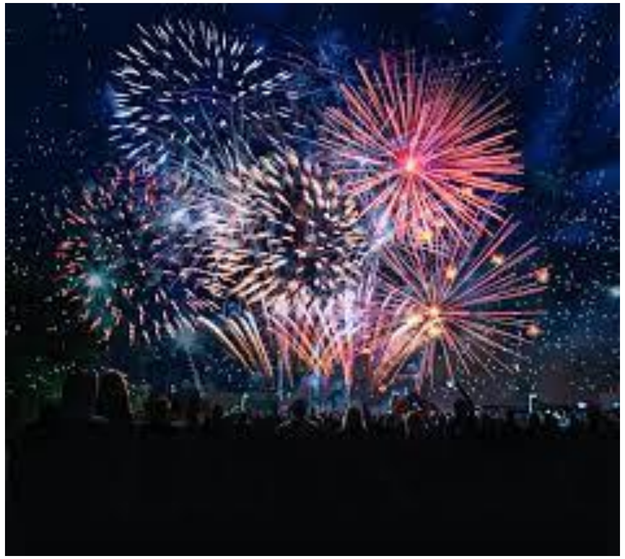

The reported p value for Renal Cell Carcinoma is verified!

### N. “corrected” chi-sq or Fishers p = “1”

Enter your data

Enter the labels and number of subjects actually observed. Don'

| | C NU | CU |
|---|---|---|
| RCC+ | 4 | 2 |
| RCC- | 64 | 32 |

Which test

- (•) Fisher's exact test
- ( ) Chi-square
- ( ) Chi-square with Yates' correction

P value Tails

- (•) Two-tailed (recommended)
- ( ) One-tailed

Calculate | Clear The Form

#### Analyze a 2x2 contingency table

| | C NU | CU | Total |
|---|---|---|---|
| RCC+ | 4 | 2 | 6 |
| RCC- | 64 | 32 | 96 |
| Total | 68 | 34 | 102 |

Fisher's exact test

The two-tailed P value equals 1.0000

The association between rows (groups) a[illegible]

is considered to be not statistically significa[illegible]t.

The result has been verified!

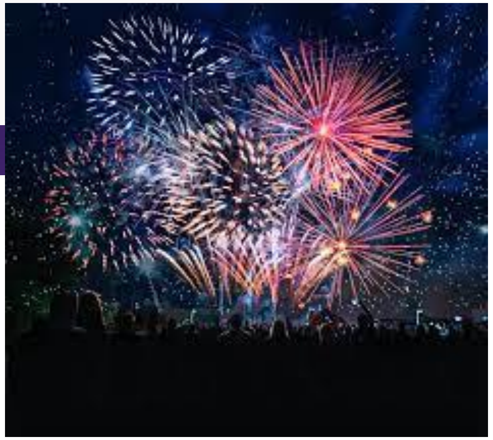

### N. “corrected” chi-sq or Fishers p = “1”

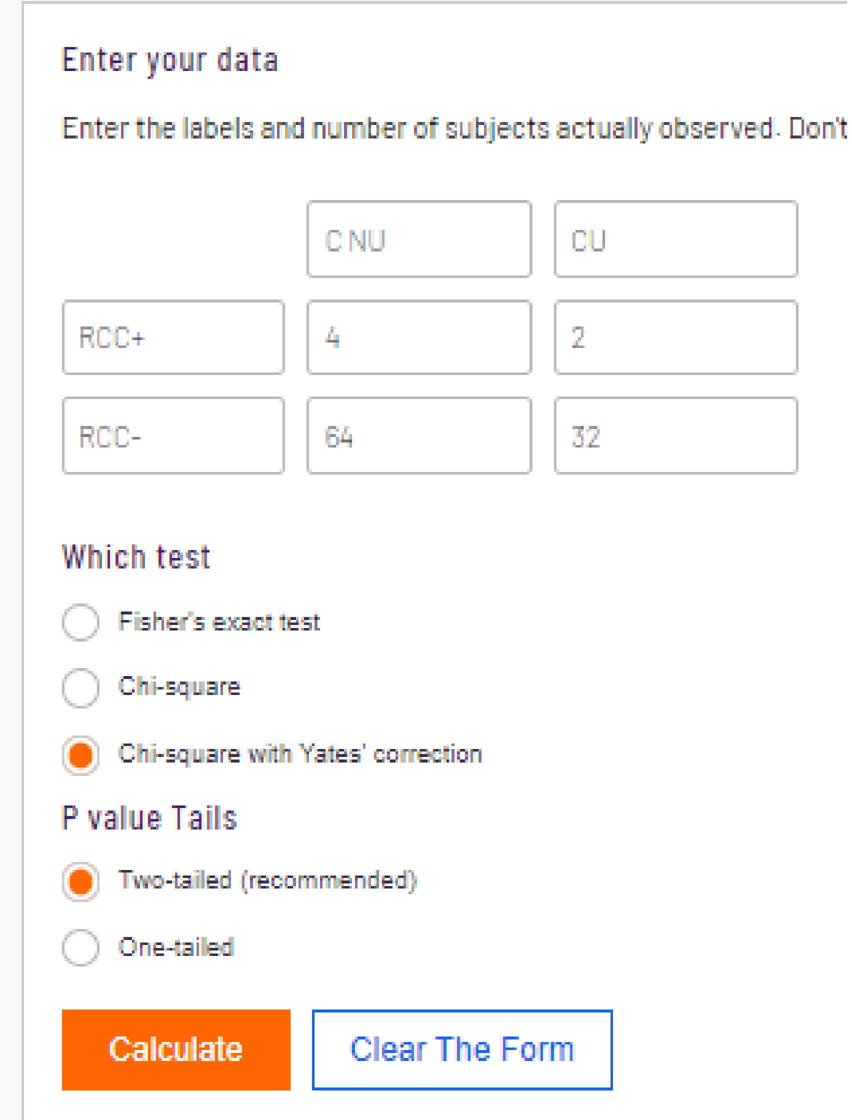


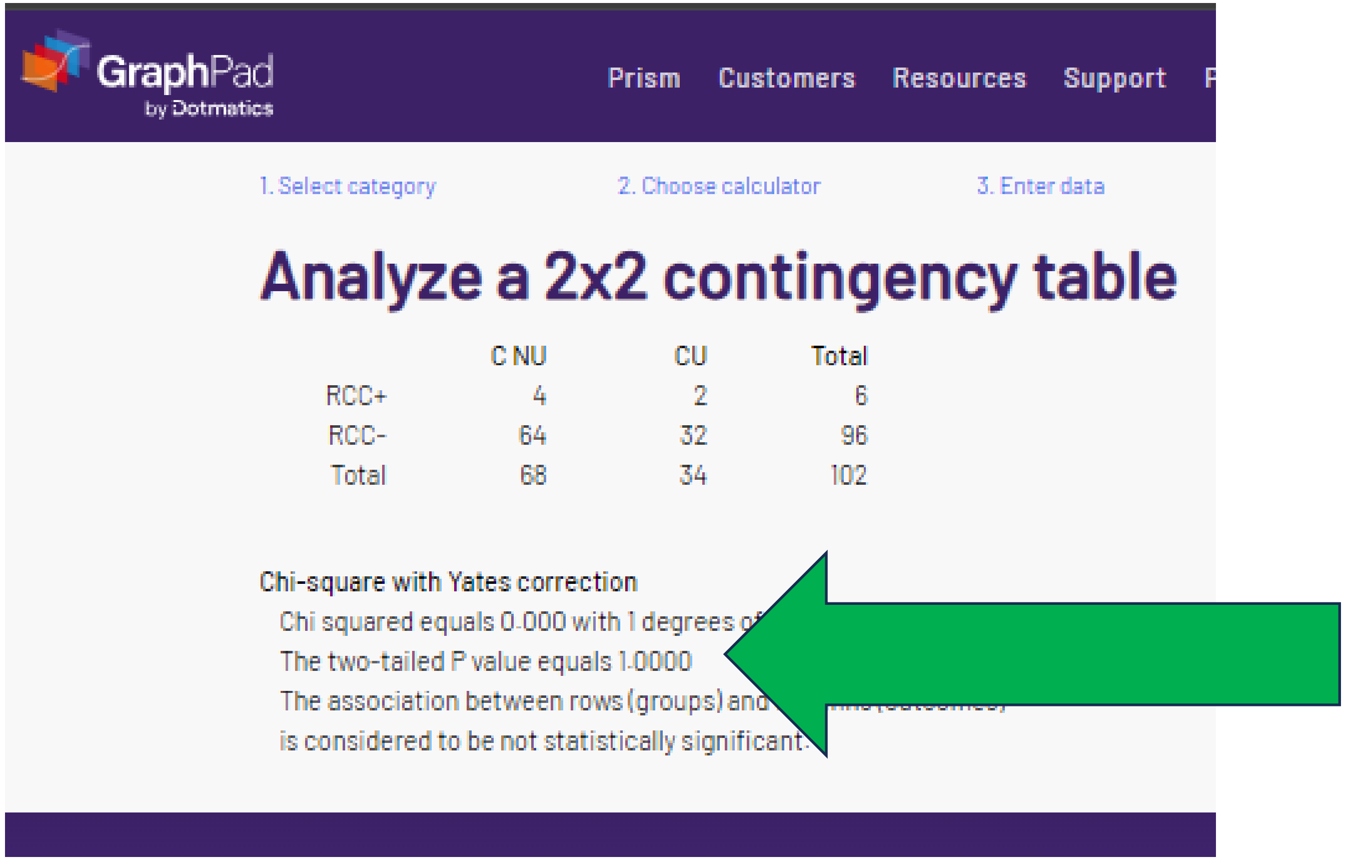


In this circumstance, all three statistics give the same result.

### O. “corrected” chi-sq or Fishers p = “1”

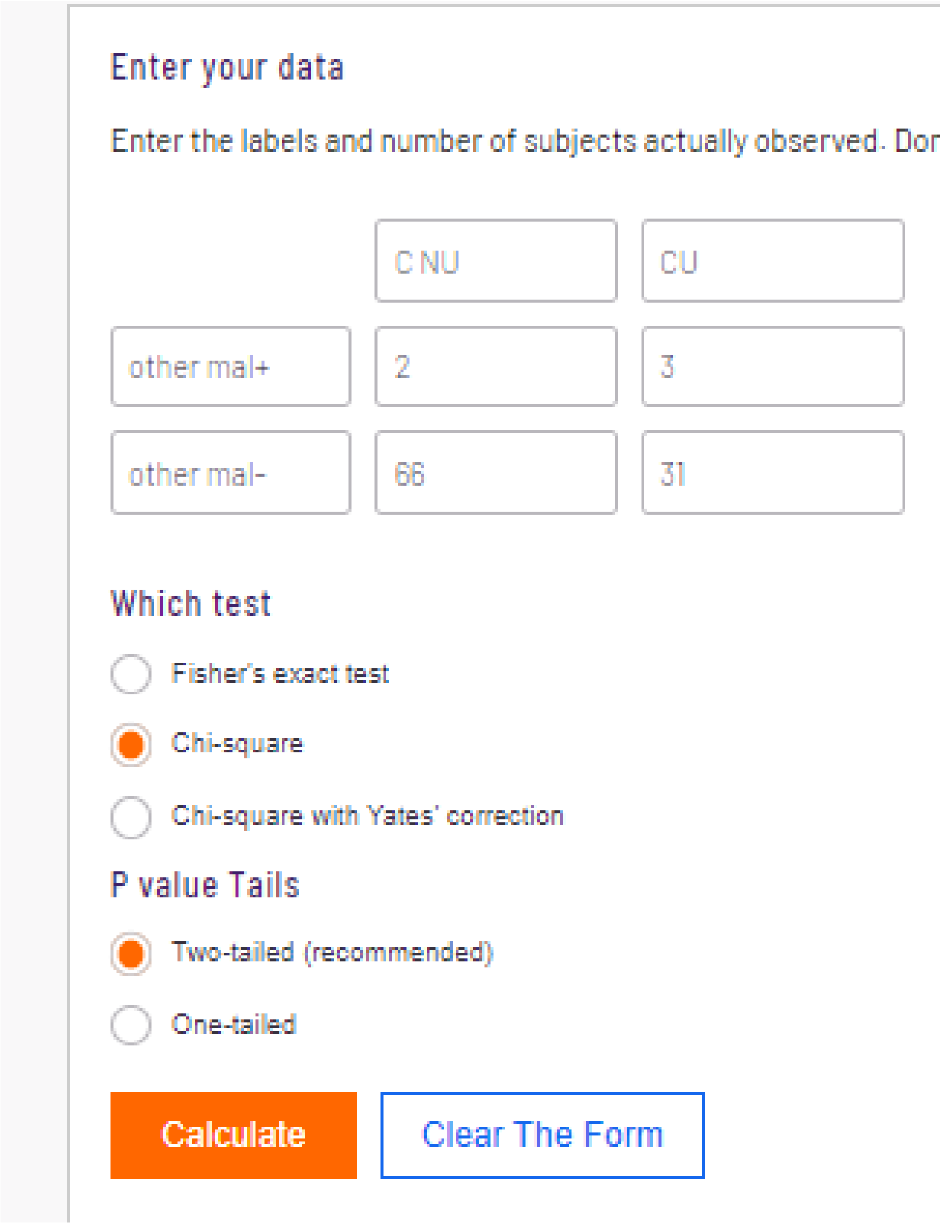


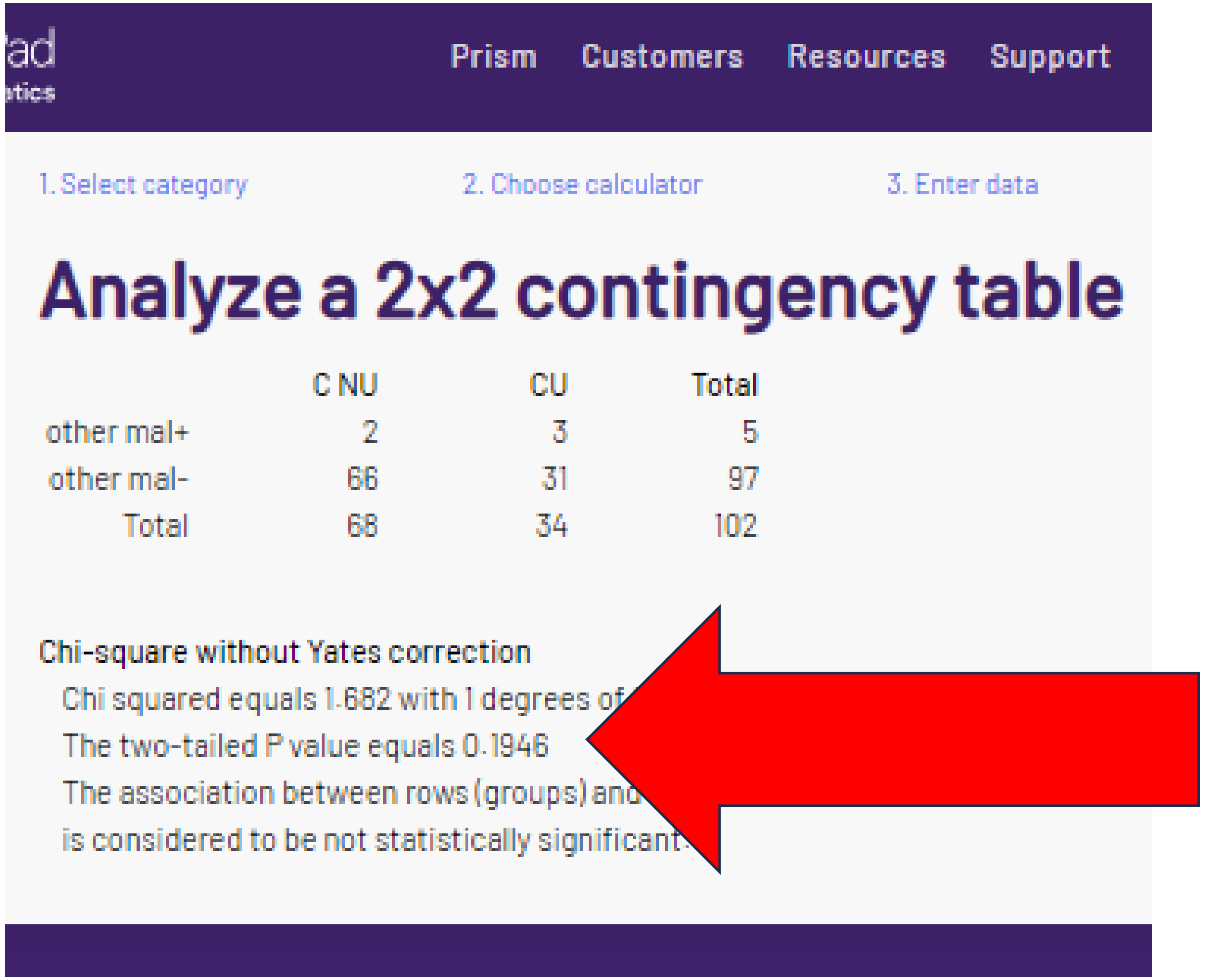

### O. "corrected" chi-sq or Fishers p = "1"

Enter your data

Enter the labels and number of subjects actually observed. Don't e

| | C NU | CU |
|---|---|---|
| other mal+ | 2 | 3 |
| other mal- | 66 | 31 |

Which test

- (●) Fisher's exact test
- ( ) Chi-square
- ( ) Chi-square with Yates' correction

P value Tails

- (●) Two-tailed (recommended)
- ( ) One-tailed

Calculate | Clear The Form

ad
tics

Prism Customers Resources Support P

1. Select category 2. Choose calculator 3. Enter data

#### Analyze a 2x2 contingency table

| | C NU | CU | Total |
|---|---|---|---|
| other mal+ | 2 | 3 | 5 |
| other mal- | 66 | 31 | 97 |
| Total | 68 | 34 | 102 |

Fisher's exact test

The two-tailed P value equals 0.3300

The association between rows (groups) an[illegible]

is considered to be not statistically significan[illegible]

The reported p-value was **not** verified.

### O. “corrected” chi-sq or Fishers p = “1”

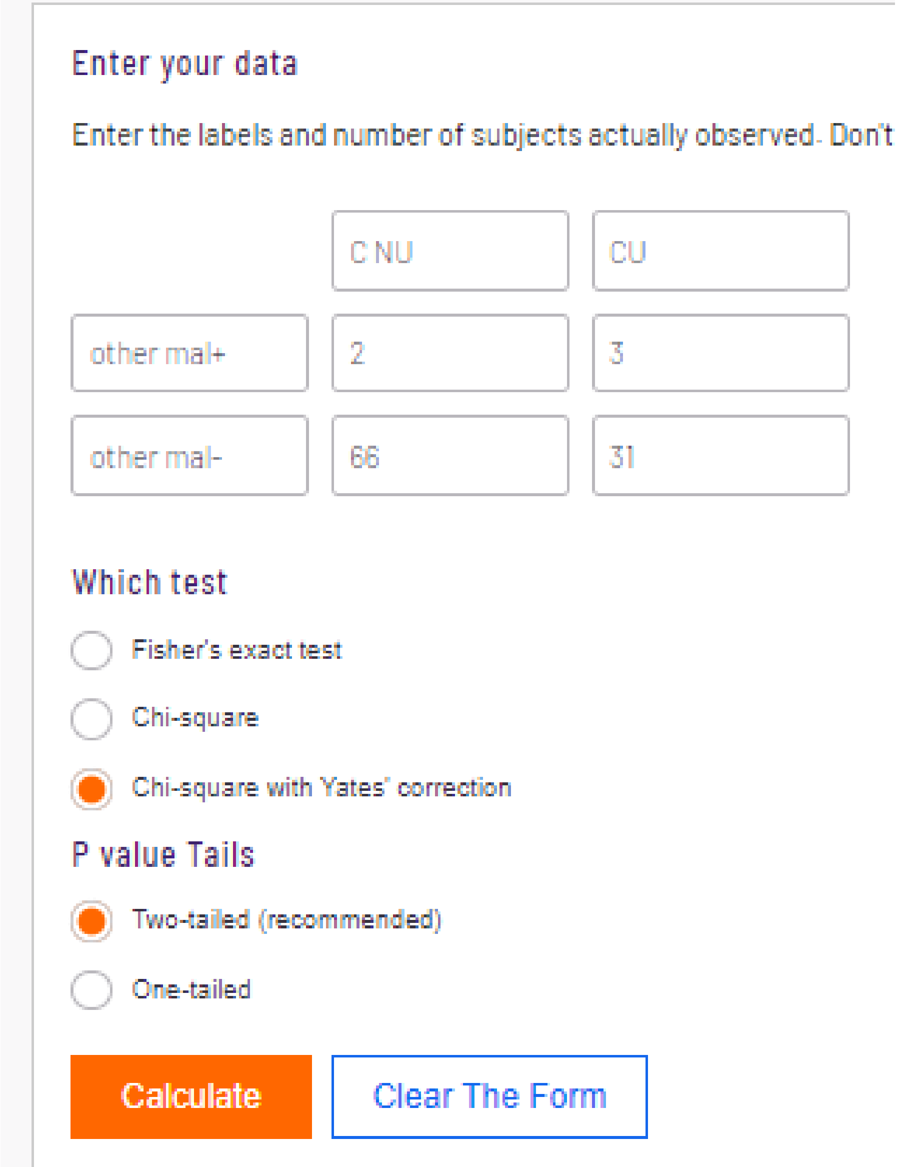


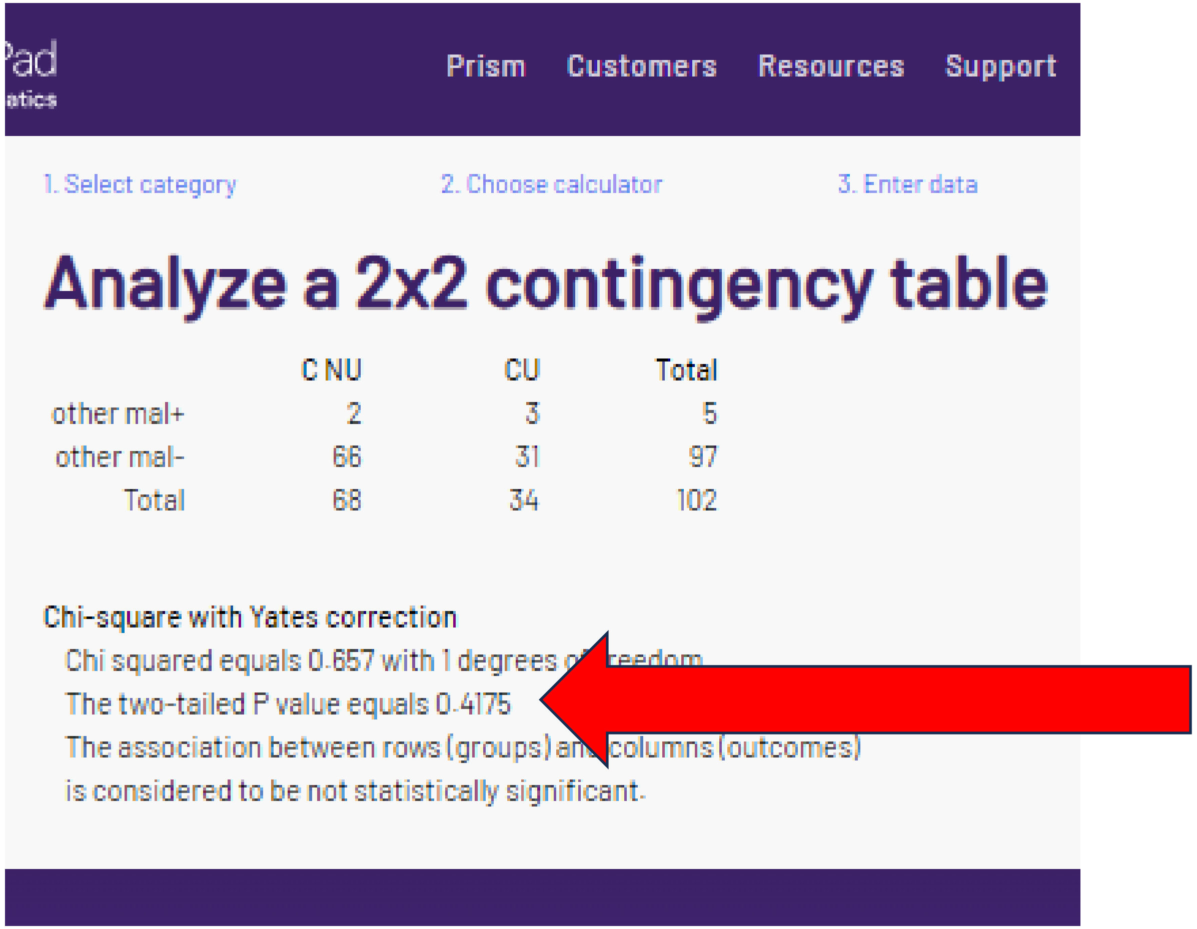


The reported p value for other malignancies was **not** verified.

### P. “corrected” chi-sq or Fishers p = .6593

Enter your data

Enter the labels and number of subjects actually observed. Don

| | C NU | CU |
|---|---|---|
| brain + | 12 | 8 |
| brain- | 56 | 26 |

Which test

- Fisher's exact test
- Chi-square
- Chi-square with Yates' correction

P value Tails

- Two-tailed (recommended)
- One-tailed

Calculate    Clear The Form

1. Select category    2. Choose calculator    3. Enter data

#### Analyze a 2x2 contingency table

| | C NU | CU | Total |
|---|---|---|---|
| brain + | 12 | 8 | 20 |
| brain- | 56 | 26 | 82 |
| Total | 68 | 34 | 102 |

Chi-square without Yates correction

Chi squared equals 0.498 with 1 degrees [illegible] freedom.

The two-tailed P value equals 0.4806

The association between rows (groups) a[illegible] columns (outcomes)

is considered to be not statistically significant.

### P. “corrected” chi-sq or Fishers p = .6593

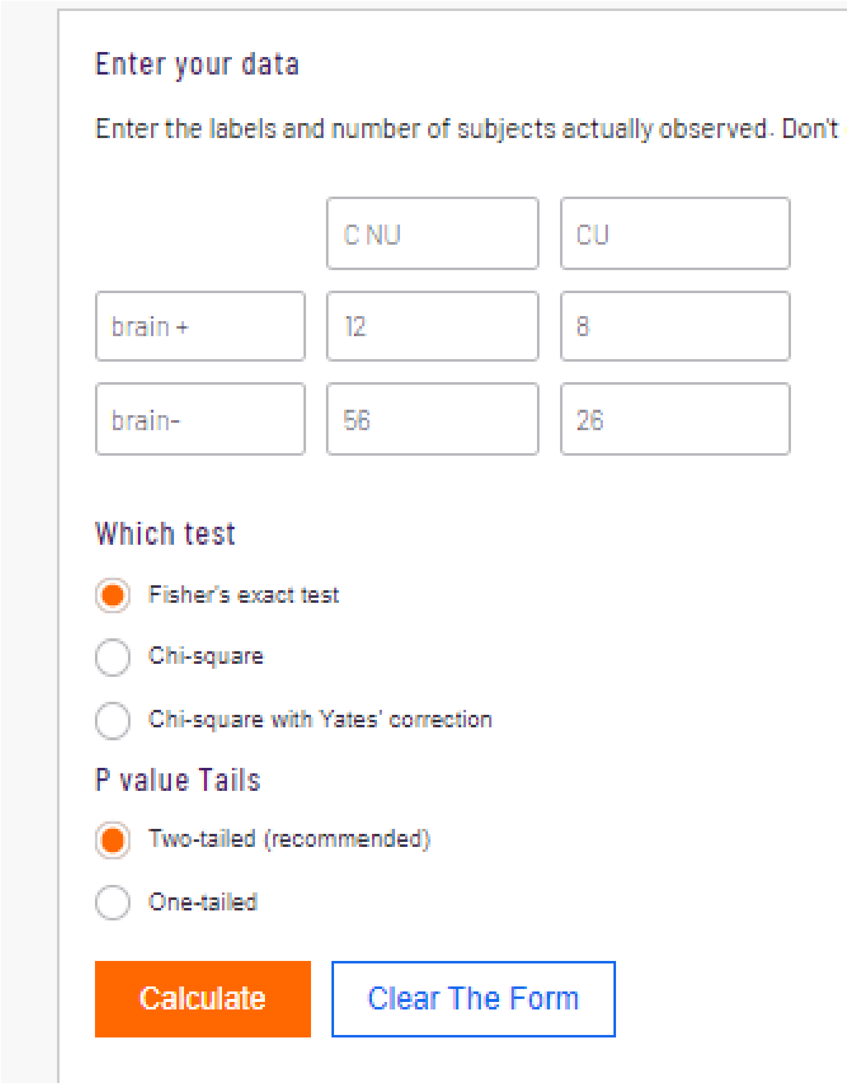


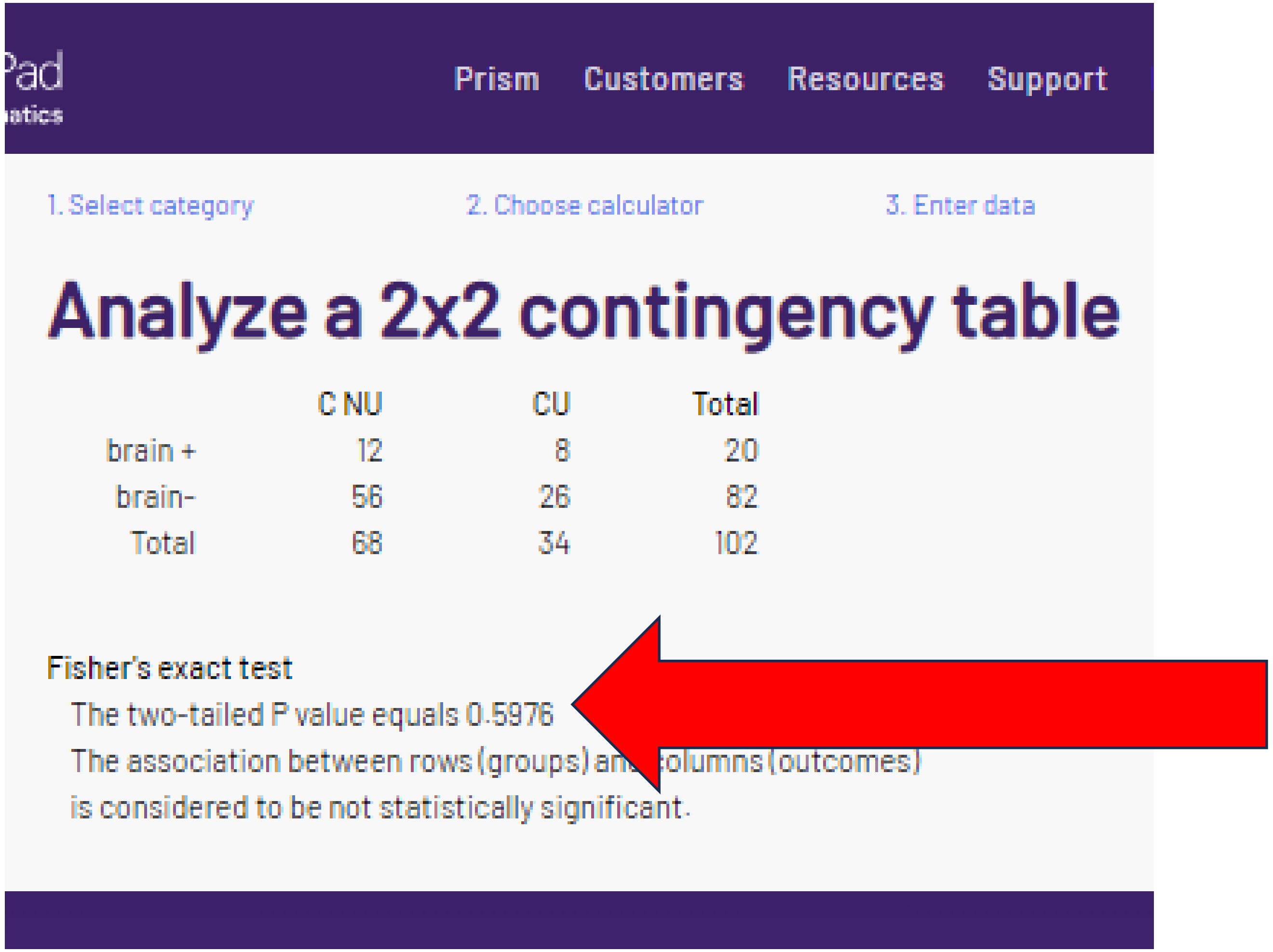


| | C NU | CU | Total |
|---|---|---|---|
| brain + | 12 | 8 | 20 |
| brain- | 56 | 26 | 82 |
| Total | 68 | 34 | 102 |

The reported p-values for brain metastasis were **not** verified.

### P. “corrected” chi-sq or Fishers p = .6593

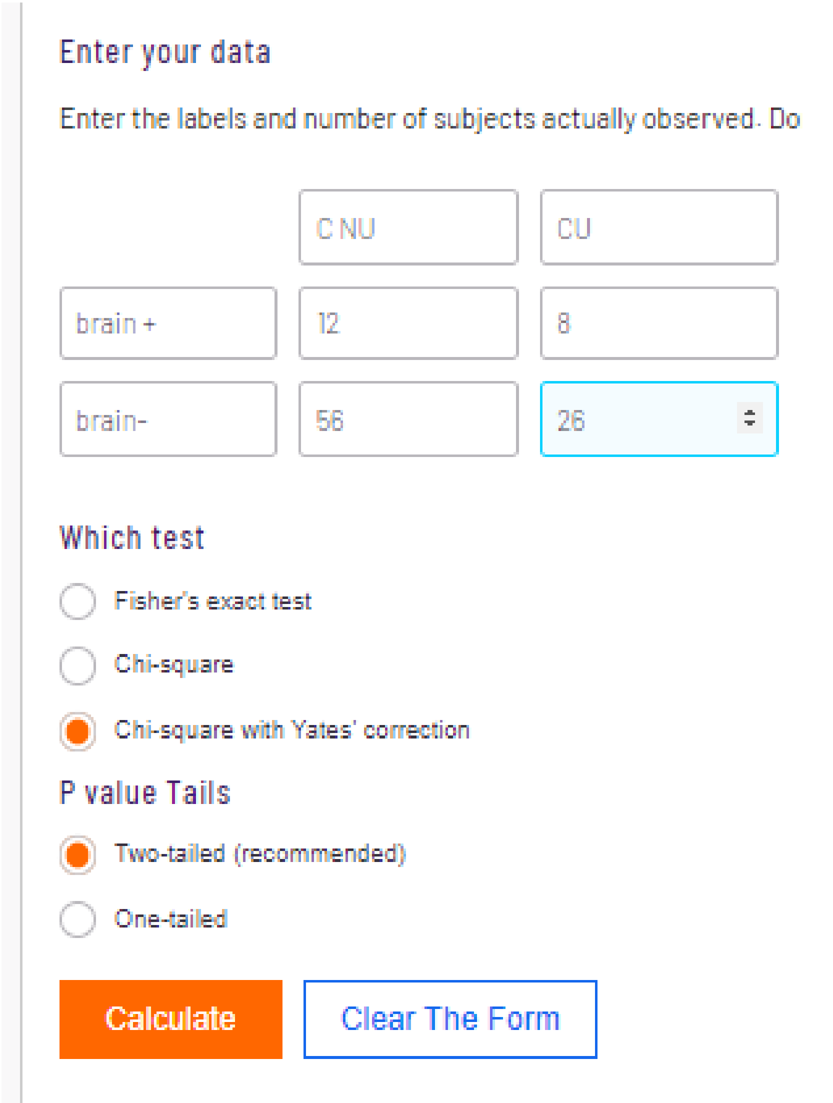

Enter your data

Enter the labels and number of subjects actually observed. Do

| | | |
|---|---|---|
| | C NU | CU |
| brain + | 12 | 8 |
| brain- | 56 | 26 |

Which test

- Fisher's exact test
- Chi-square
- Chi-square with Yates' correction

P value Tails

- Two-tailed (recommended)
- One-tailed

Calculate | Clear The Form

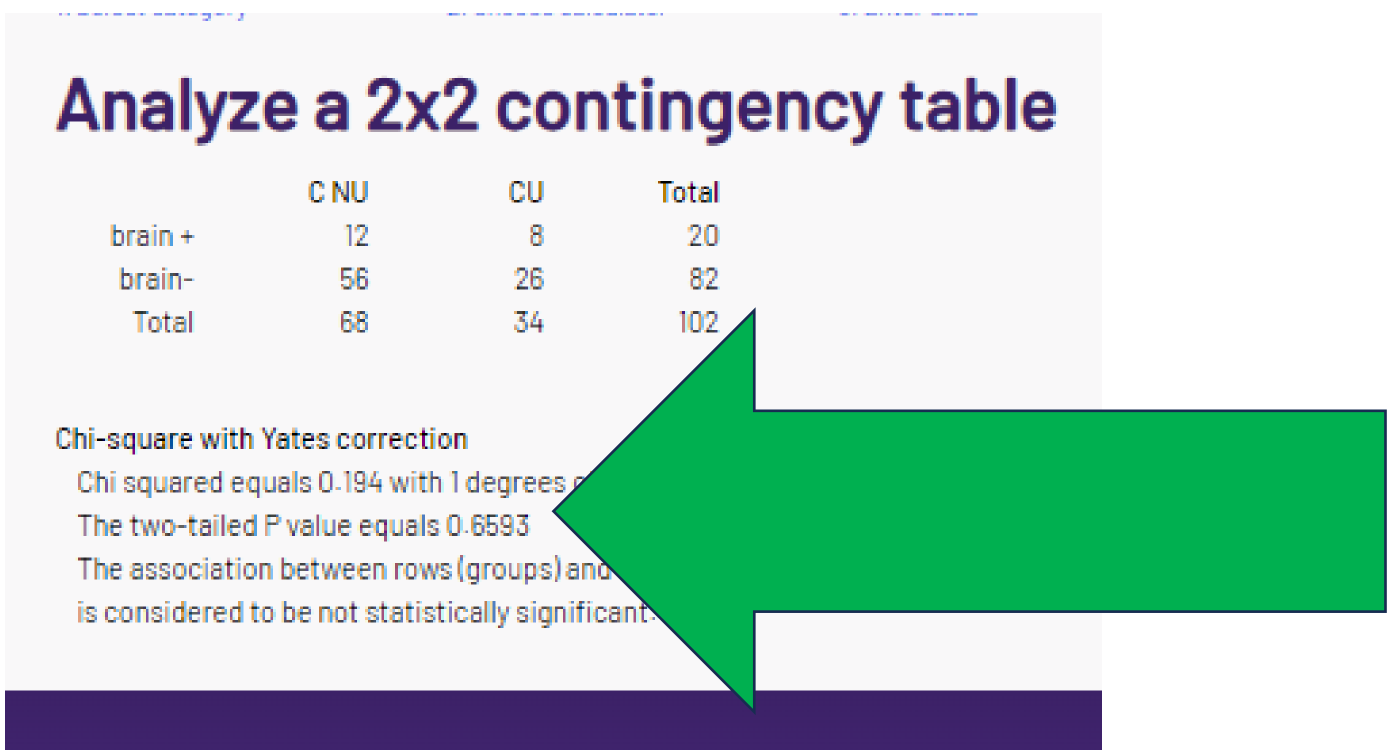

Analyze a 2x2 contingency table

| | C NU | CU | Total |
|---|---|---|---|
| brain + | 12 | 8 | 20 |
| brain- | 56 | 26 | 82 |
| Total | 68 | 34 | 102 |

Chi-square with Yates correction

Chi squared equals 0.194 with 1 degrees [illegible]

The two-tailed P value equals 0.6593

The association between rows (groups) and [illegible]

is considered to be not statistically significant.

For the smallest N per cell = 8, Yates would typically not be considered the right statistic. However, the reported p value corresponds with Yates.

### Q. “corrected” chi-sq or Fishers p = .4303

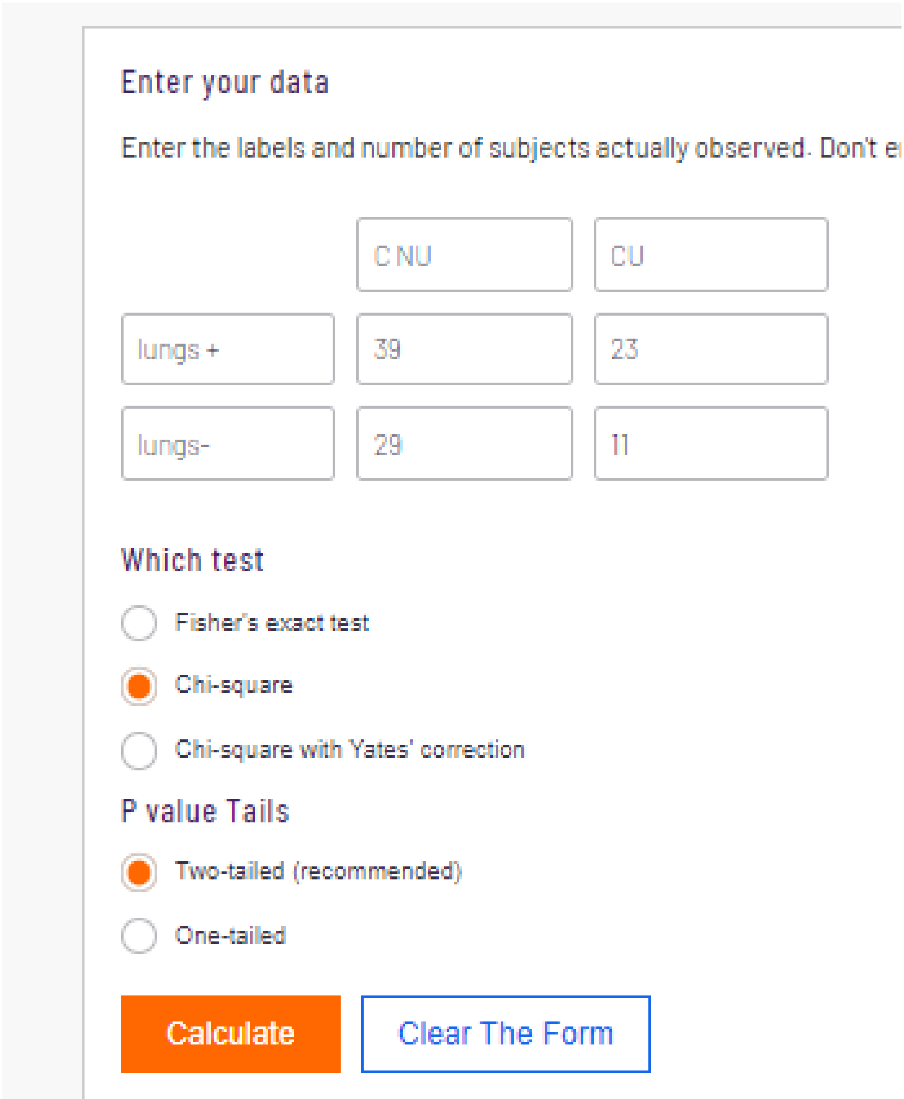

Enter your data

Enter the labels and number of subjects actually observed. Don't e[illegible]

| | | |
|---|---|---|
| | C NU | CU |
| lungs + | 39 | 23 |
| lungs- | 29 | 11 |

Which test

- Fisher's exact test
- Chi-square
- Chi-square with Yates' correction

P value Tails

- Two-tailed (recommended)
- One-tailed

Calculate | Clear The Form

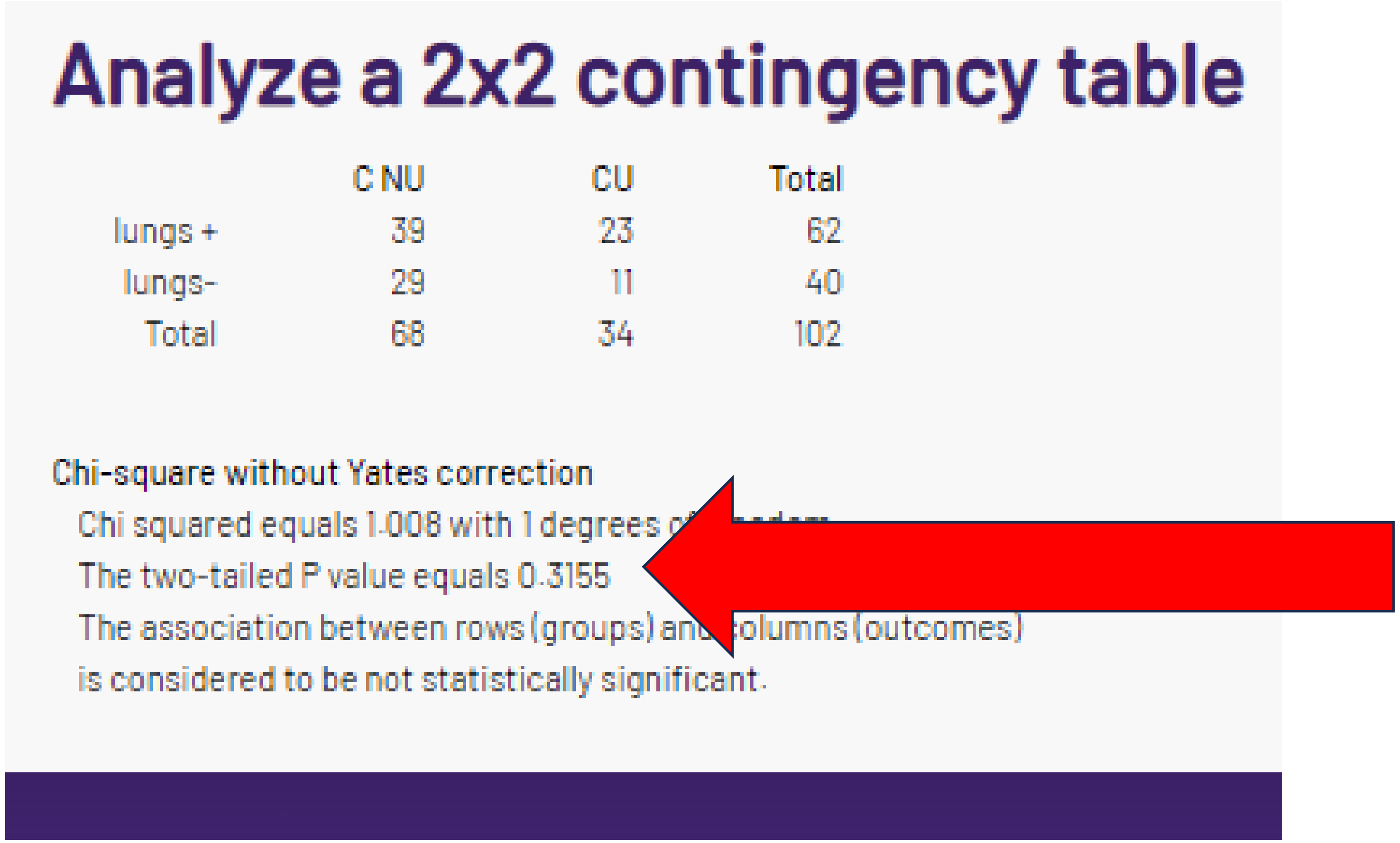

#### Analyze a 2x2 contingency table

| | C NU | CU | Total |
|---|---|---|---|
| lungs + | 39 | 23 | 62 |
| lungs- | 29 | 11 | 40 |
| Total | 68 | 34 | 102 |

Chi-square without Yates correction

Chi squared equals 1.008 with 1 degrees o[illegible]

The two-tailed P value equals 0.3155

The association between rows (groups) an[illegible]olumns (outcomes)

is considered to be not statistically significant.

### Q. “corrected” chi-sq or Fishers p = .4303

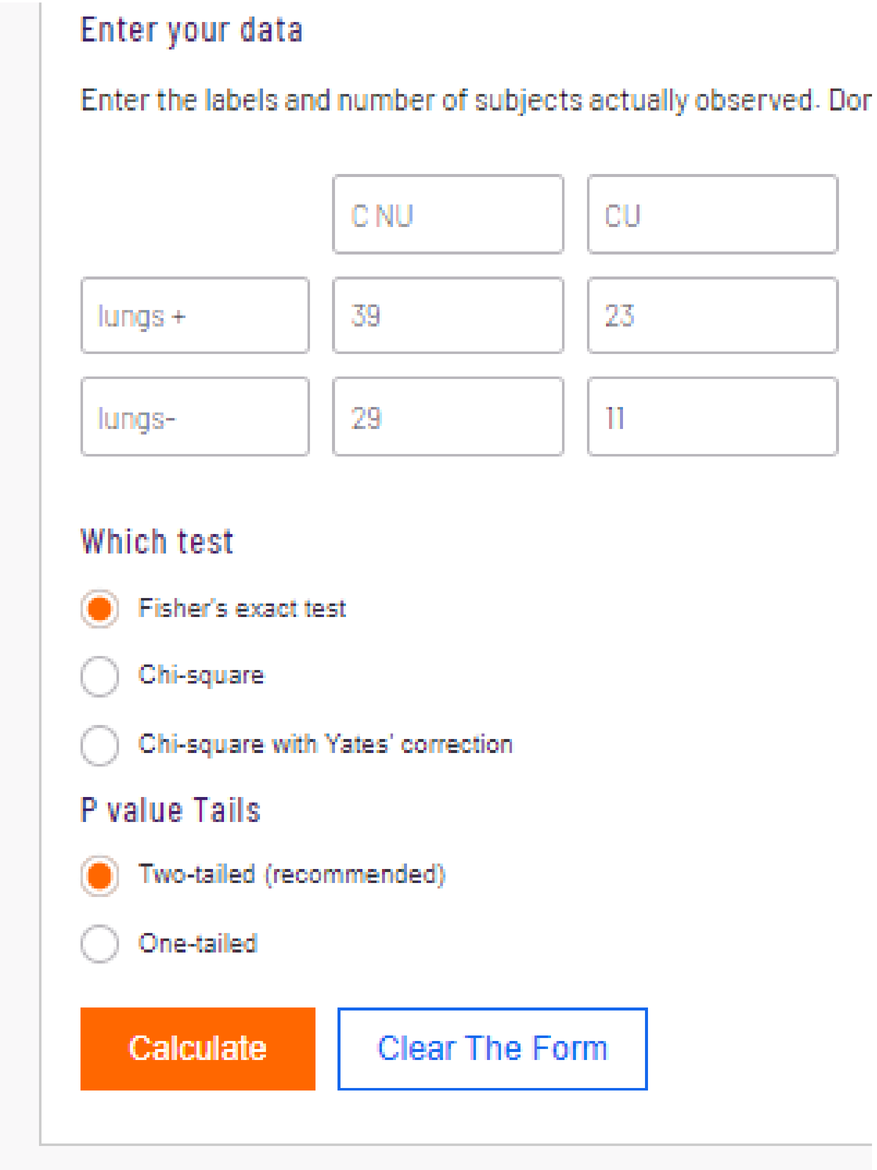


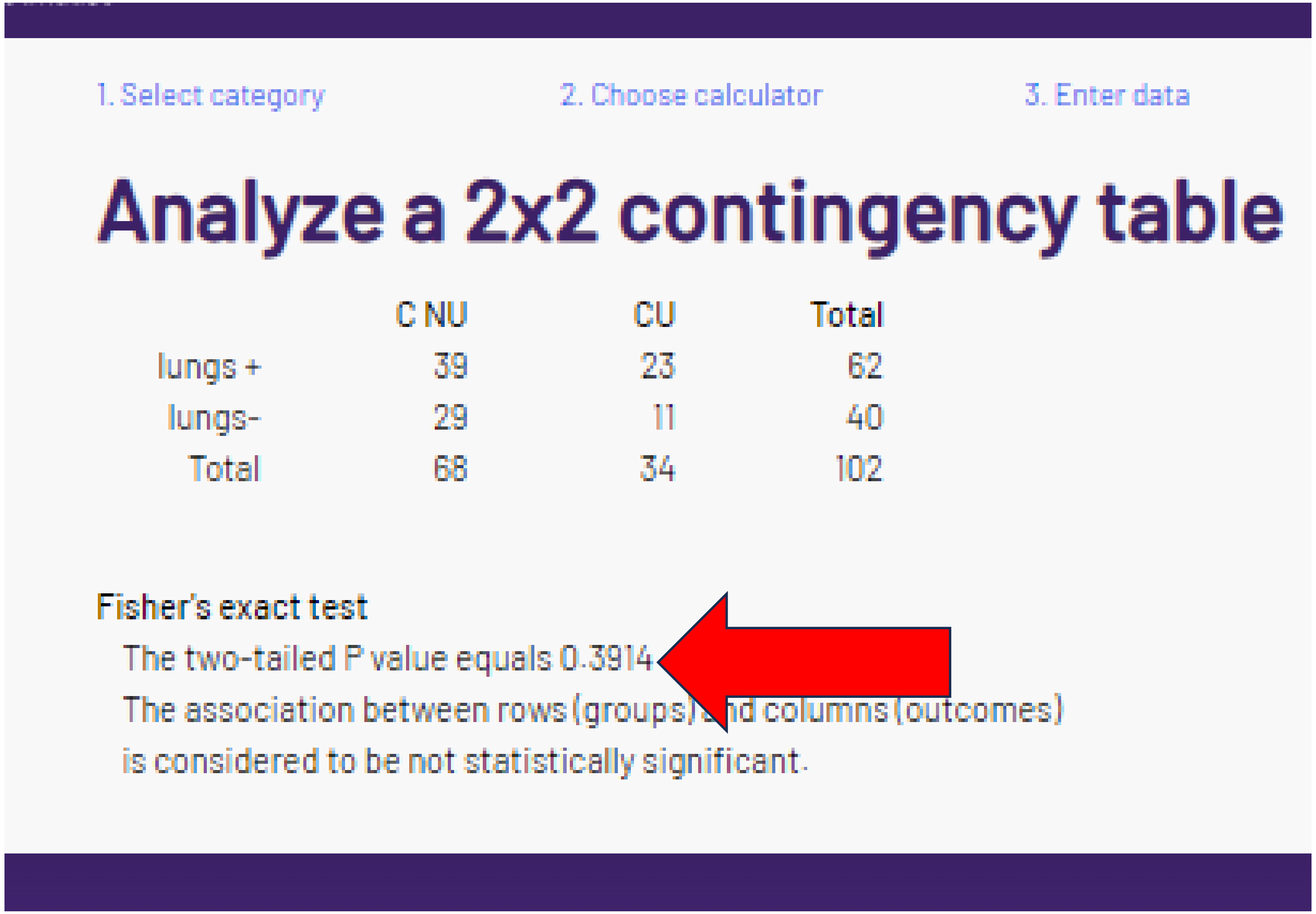


The reported p value for lung metastasis could **not** be verified.

### Q. “corrected” chi-sq or Fisher’s p = .4303

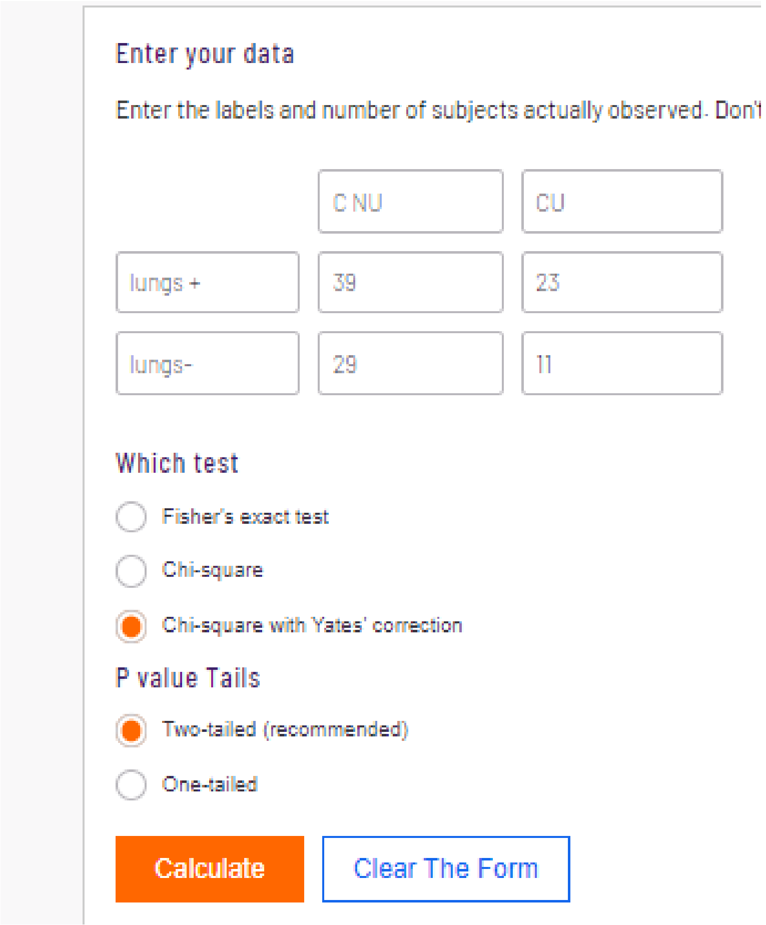


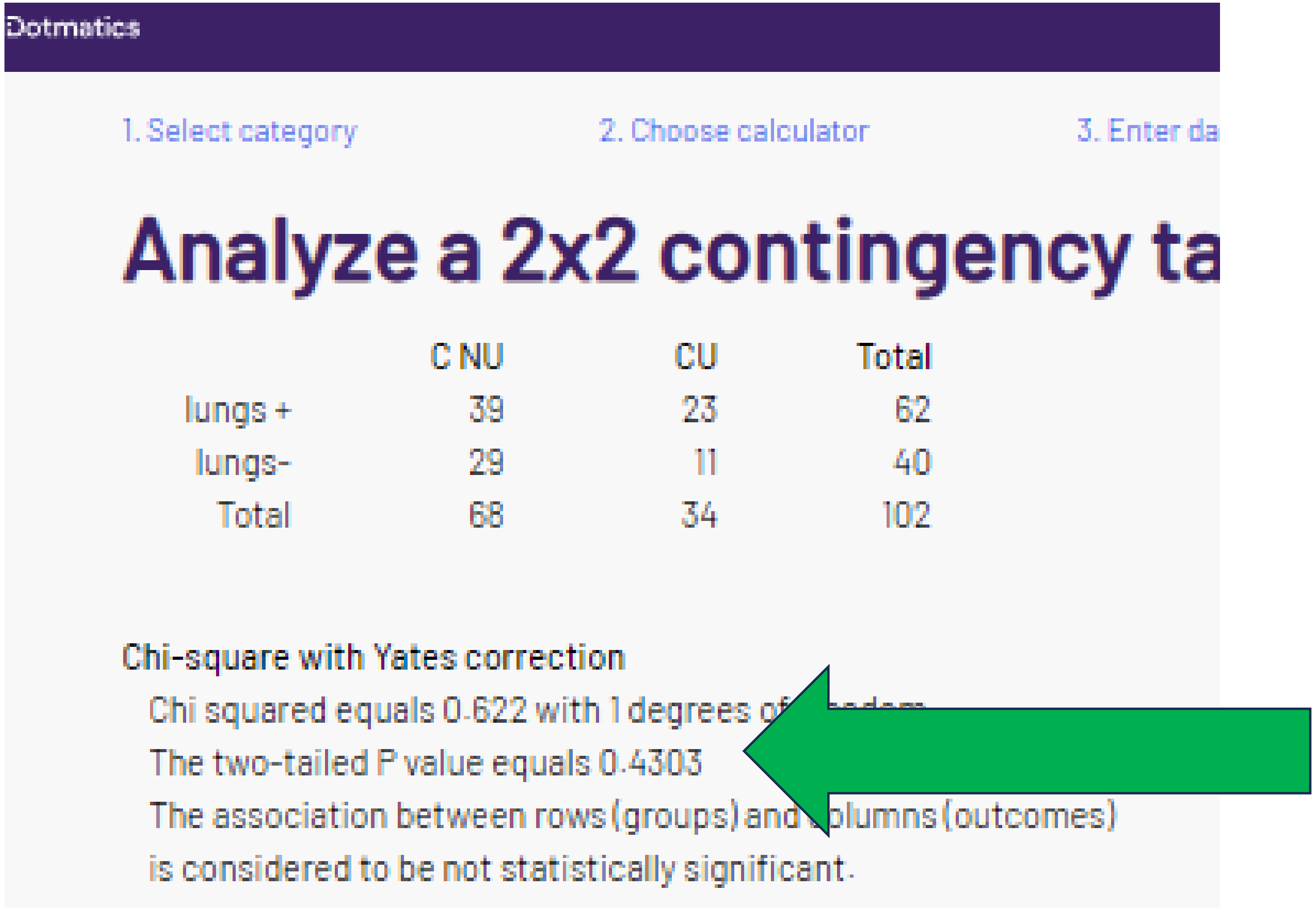
Dotmatics

1. Select category 2. Choose calculator 3. Enter da

Analyze a 2x2 contingency ta

| | C NU | CU | Total |
|---|---|---|---|
| lungs + | 39 | 23 | 62 |
| lungs- | 29 | 11 | 40 |
| Total | 68 | 34 | 102 |

Chi-square with Yates correction

Chi squared equals 0.622 with 1 degrees of [illegible]

The two-tailed P value equals 0.4303

The association between rows (groups) and [illegible]olumns (outcomes)

is considered to be not statistically significant.

With the smallest N per cell = 11, Yates correction is typically **not** employed. However, the p-value can be reproduced.

### R. “corrected” chi-sq or Fishers p = .2157

#### Enter your data

Enter the labels and number of subjects actually observ[illegible]
means.

| | | |
|---|---|---|
| | C NU | CU |
| liver + | 13 | 11 |
| liver- | 55 | 23 |

#### Which test

- ○ Fisher's exact test
- ● Chi-square
- ○ Chi-square with Yates' correction

#### P value Tails

- ● Two-tailed (recommended)
- ○ One-tailed

Calculate | Clear The Form

GraphPad
by Dotmatics

1. Select category 2. Choose calculator 3. Enter data 4. Vi[illegible]

### Analyze a 2x2 contingency table

| | C NU | CU | Total |
|---|---|---|---|
| liver + | 13 | 11 | 24 |
| liver- | 55 | 23 | 78 |
| Total | 68 | 34 | 102 |

**Chi-square without Yates correction**

Chi squared equals 2.207 with 1 degrees [illegible]
The two-tailed P value equals 0.1374
The association between rows (groups) an[illegible]olumns (outcomes)
is considered to be not statistically significant.

Note large >= 11 cell values.

### R. “corrected” chi-sq or Fishers p = .2157

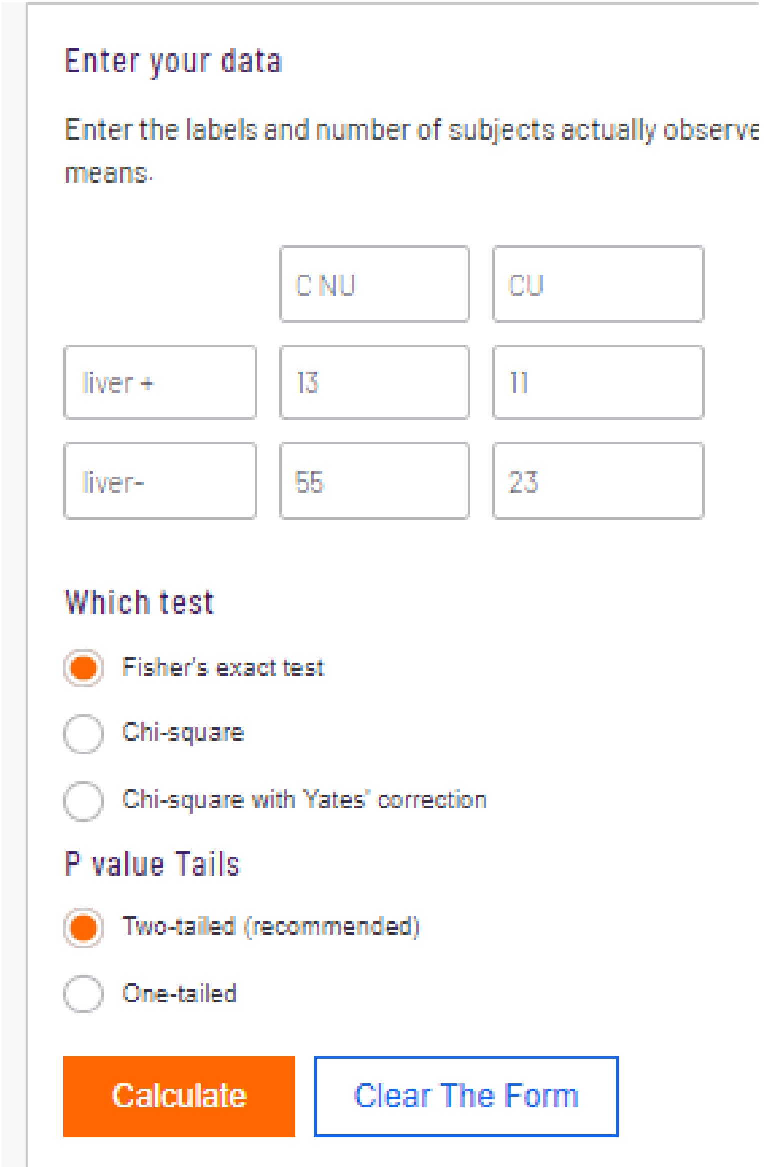


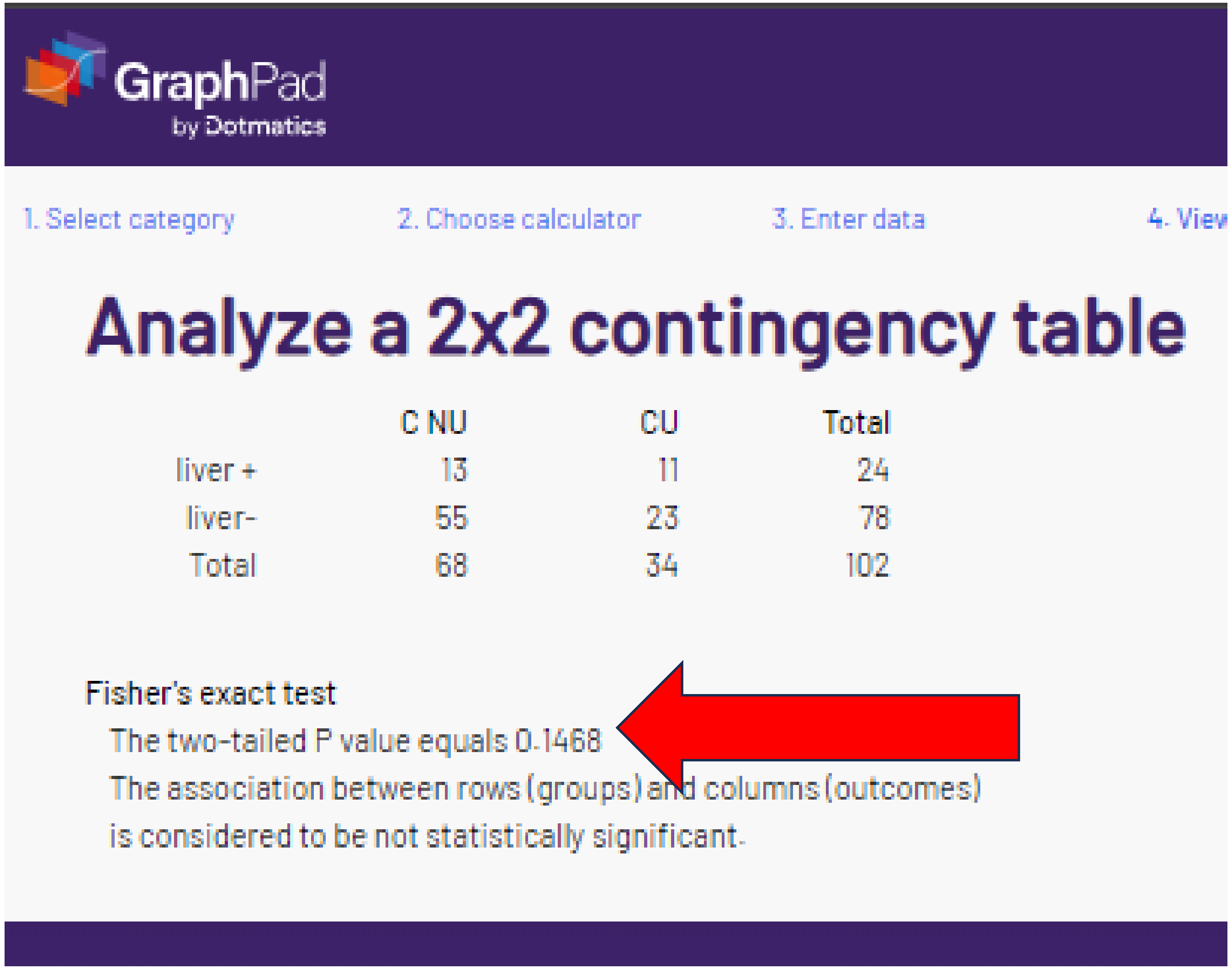


The reported p-values could **not** be verified.

### R. “corrected” chi-sq or Fishers p = .2157

Enter your data

Enter the labels and number of subjects actually observ
means.

| | C NU | CU |
|---|---|---|
| liver + | 13 | 11 |
| liver- | 55 | 23 |

Which test

- Fisher's exact test
- Chi-square
- Chi-square with Yates' correction (selected)

P value Tails

- Two-tailed (recommended) (selected)
- One-tailed

Calculate | Clear The Form

GraphPad
by Dotmatics

1. Select category 2. Choose calculator 3. Enter data 4. View

#### Analyze a 2x2 contingency table

| | C NU | CU | Total |
|---|---|---|---|
| liver + | 13 | 11 | 24 |
| liver- | 55 | 23 | 78 |
| Total | 68 | 34 | 102 |

Chi-square with Yates correction

Chi squared equals 1.532 with 1 degrees of freedom.
The two-tailed P value equals 0.2157
The association between rows (groups) and columns (outcomes)
is considered to be not statistically significant.

With the smallest N per cell = 11, Yates is **not** appropriate.
However, the p value is reproduced with this incorrect statistic.

**Table 1.** *Cont.* (page 2)

| | Characteristics | Cannabis Non-Users N = 68 | | Cannabis Users N = 34 | | *p*-Value | P values: Chi-sq | Fisher's | Chi-sq w/Yates |
|---|---|---|---|---|---|---|---|---|---|
| | Immunotherapy given as—N (%) | | | | | | | | |
| S | First line of treatment | 31 (45.5) | 45.6 | 8 (23.5) | 23.5 | 0.05178 | .0307 | .0334 | .0518 |
| | Second line of treatment or more | 37 (54.4) | 54.4 | 26 (76.4) | 76.5 | 0.05178 | | | |
| | Checkpoint therapy—N (%) | | | | | | | | |
| T | Anti PD1: Pembrolizumab or Nivolumab | 47 (69.1) | 69.1 | 29 (85.2) | 85.3 | 0.127 | .0772 | .0945 | .1270 |
| U | Ipilimumab and Nivolumab | 16 (23.5) | 23.5 | 4 (11.7) | 11.8 | 0.2517 | .1583 | .1934 | .2517 |
| V | Anti PDL-1: Durvalumab or Atezolizumab | 5 (7.3) | 7.4 | 1 (2.9) | 2.9 | 1 | ..3720 | .6607 | .6554 |

Key: green = % calculated (rounded) correctly

red = % calculated (rounding) incorrectly

P values calculated with GraphPad Prism. Although it is theoretically possible that the differences are due to software (Bel-Sala used SAS), we think this is exceedingly unlikely for simple non-parametric statistic like this. It is much more likely (based on values corresponding to 4 decimal places) that the authors simply ran a different statistic than was reported. Unfortunately, the statistic that was run (chi-square with Yates correction) is (again) only appropriate with a expected cell N of < 5 (Wikipedia) Or 0-9 (BMJ Guidance).

### S. “corrected” chi-sq or Fisher’s p value is .05178

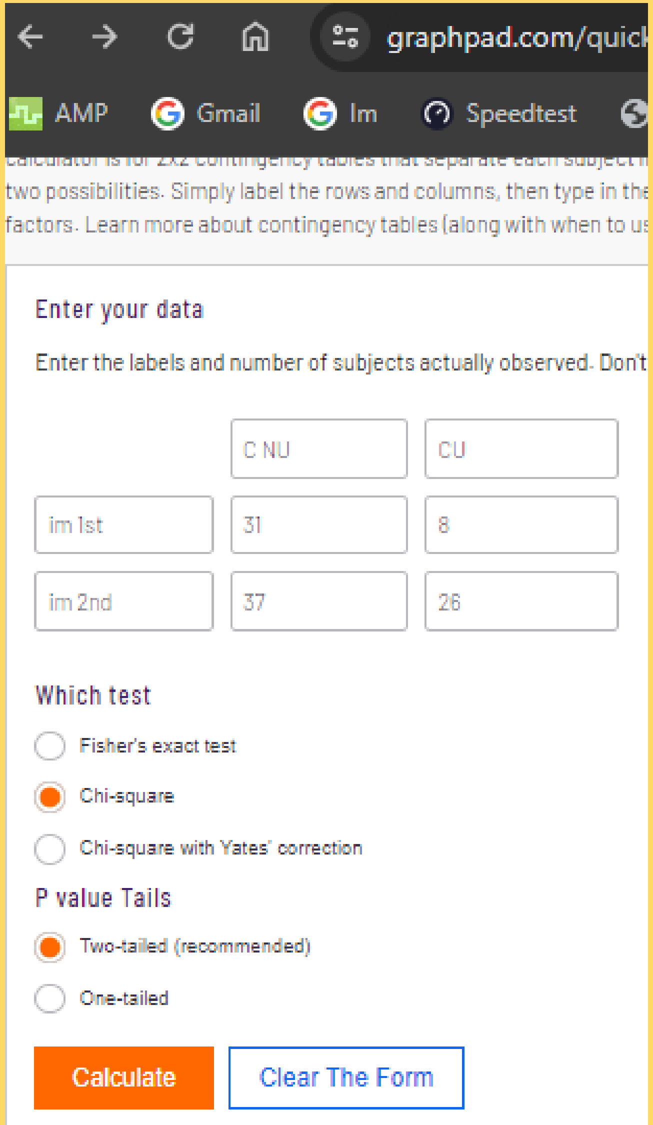


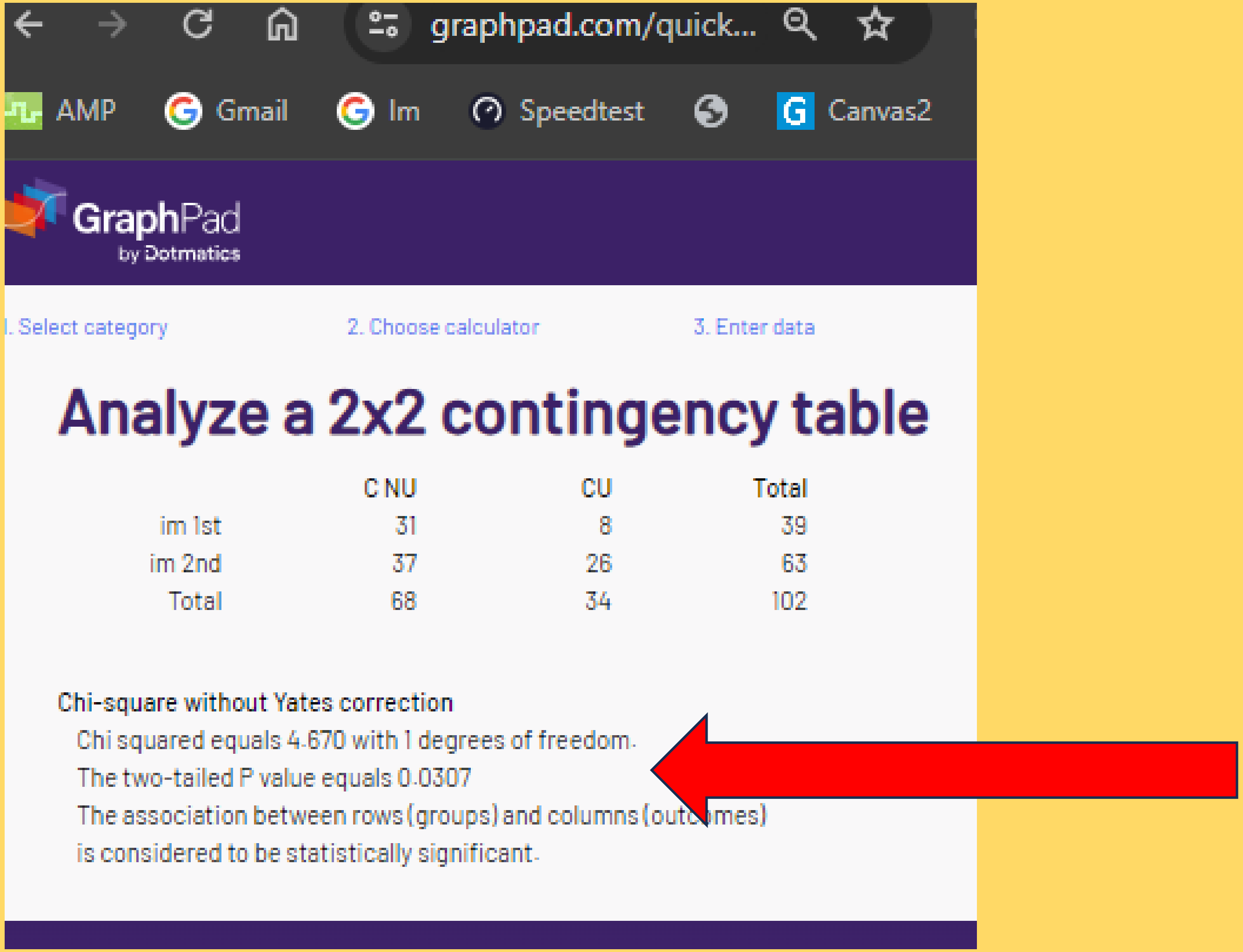


It is interesting that this is the only p value on Table 1 that the authors reported to 5 decimal places.

### S. “corrected” chi-sq or Fisher’s p value is .05178

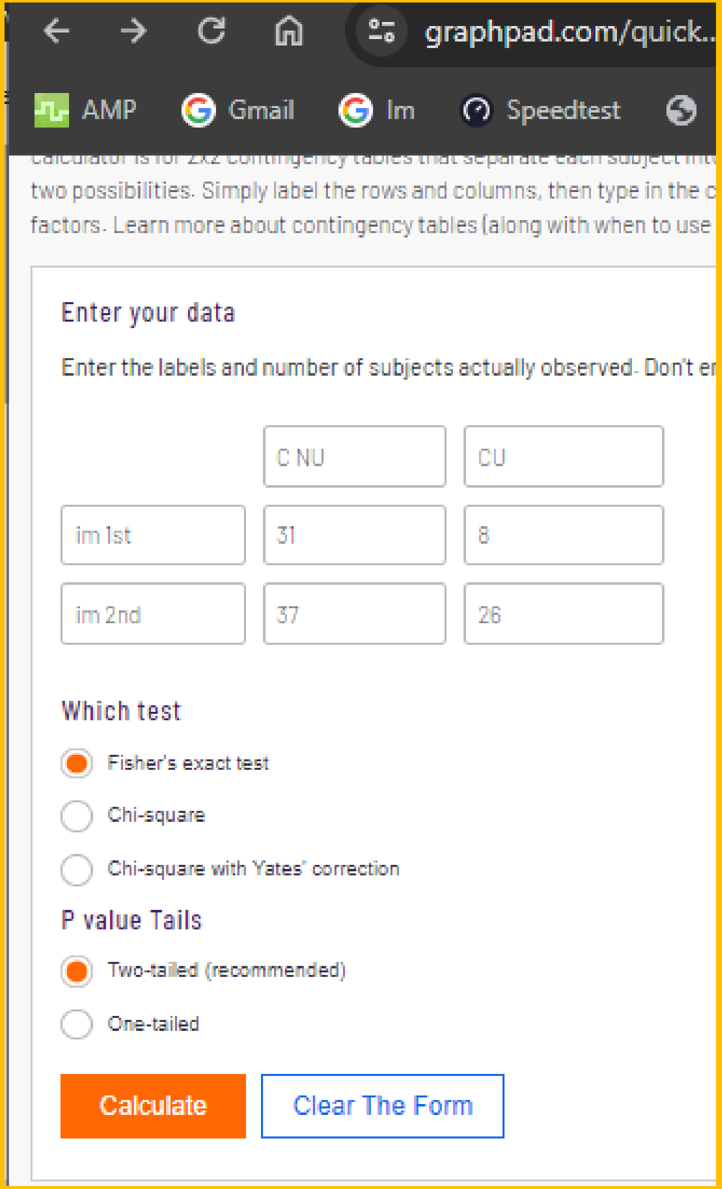


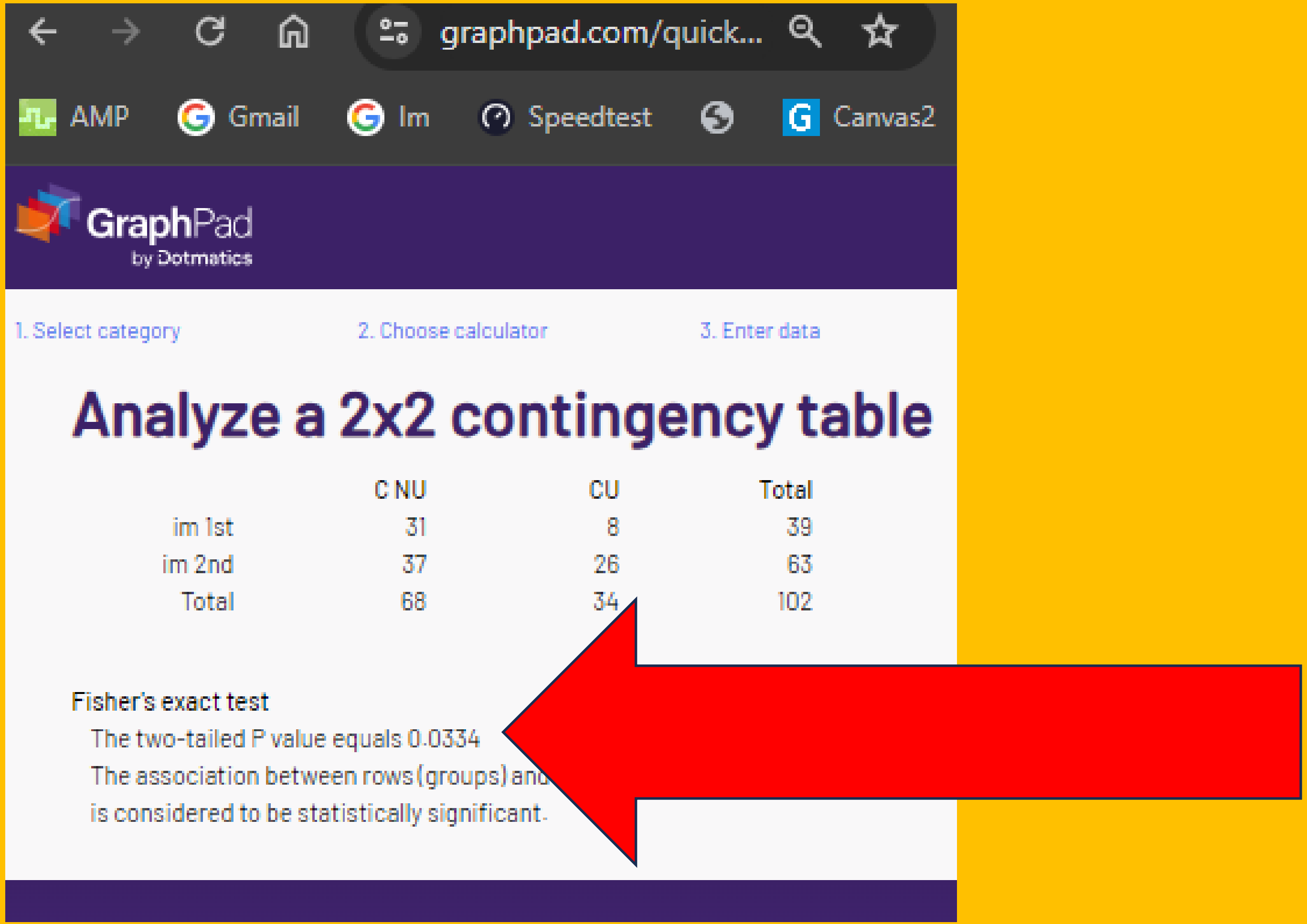


The reported p value for immunotherapy 1st vs 2nd could not be verified with either chi-square or Fisher’s exact test. As this manuscript is about immunotherapy, this significant ($p < .05$) difference could be an important confound. This was not addressed In the data-analysis (e.g. ANCOVA) or in the discussion section.

### S. “corrected” chi-sq or Fisher’s p value is .05178

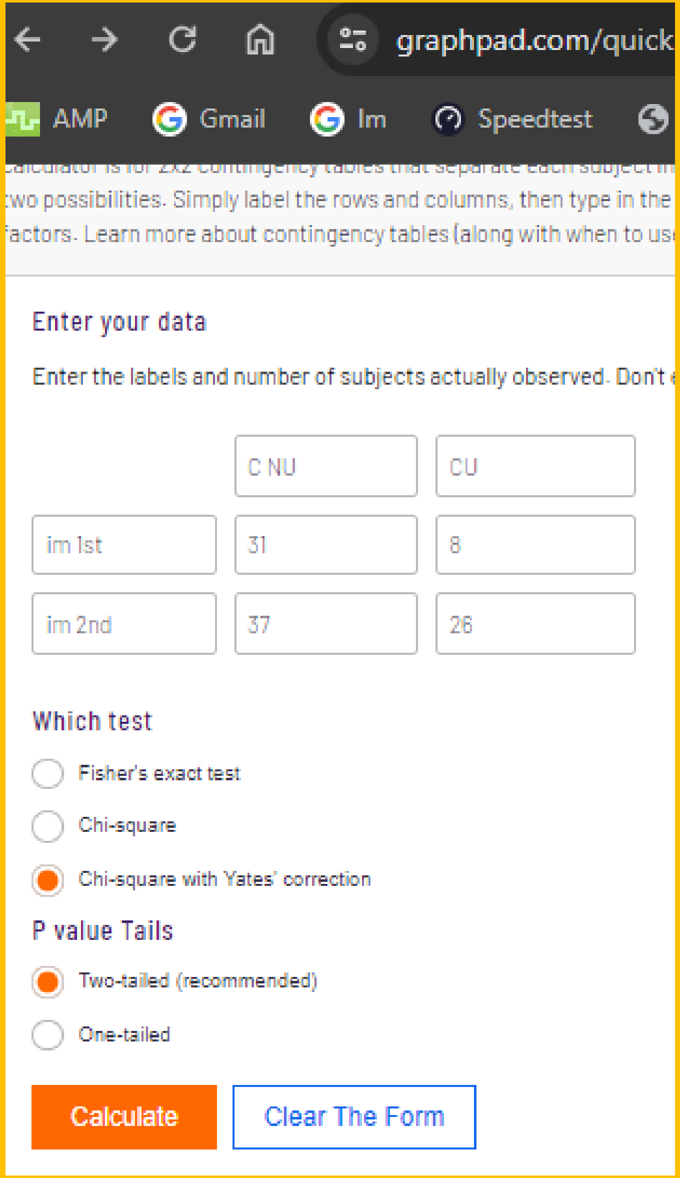


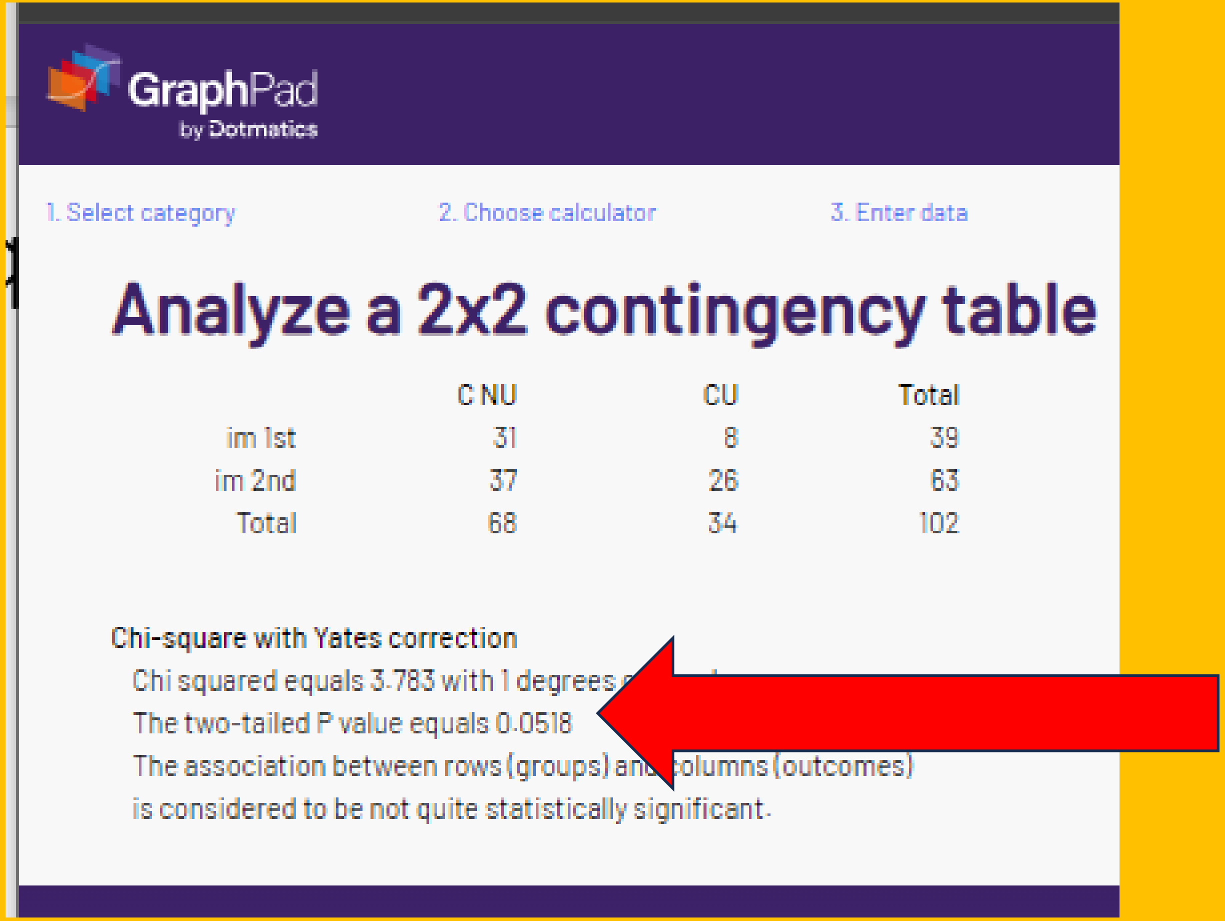


The smallest cell N is 8 so Yates would **not** be appropriate. Although Prism only reports to 4 decimal places, with rounding, the “corrected” p value is reproduced, albeit with a statistic that is both inappropriate (arguably) and not described in the methods.

### T. "corrected" chi-sq or Fisher's = .127

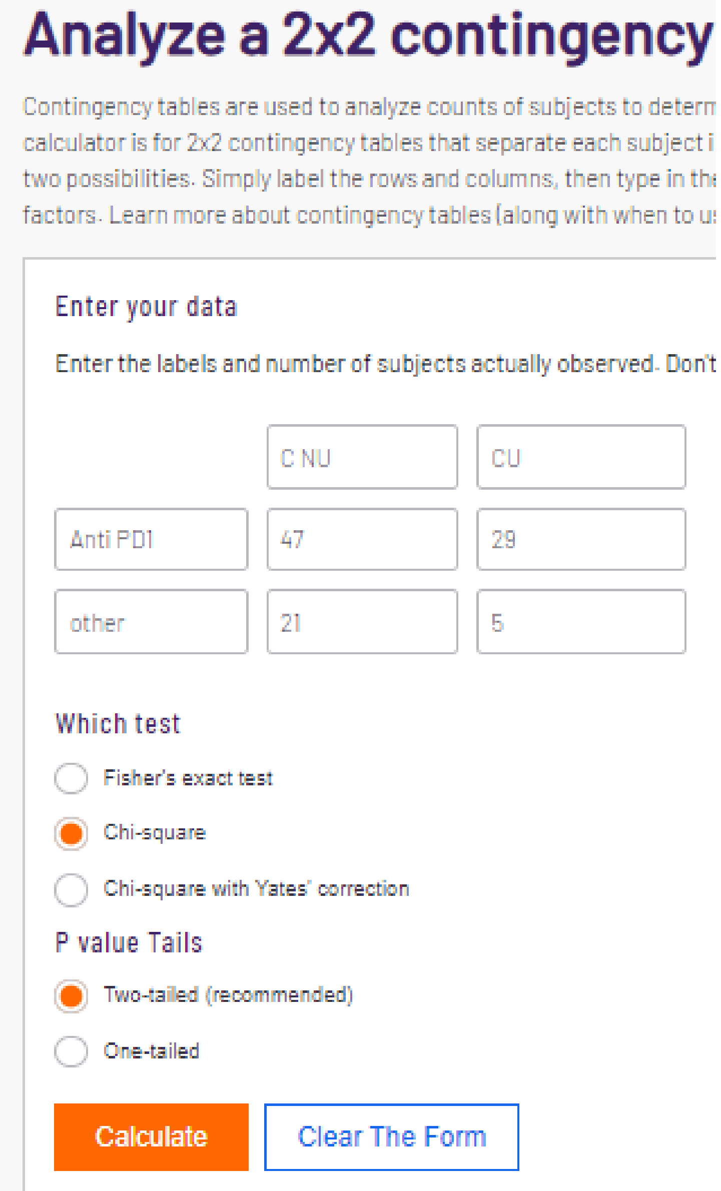


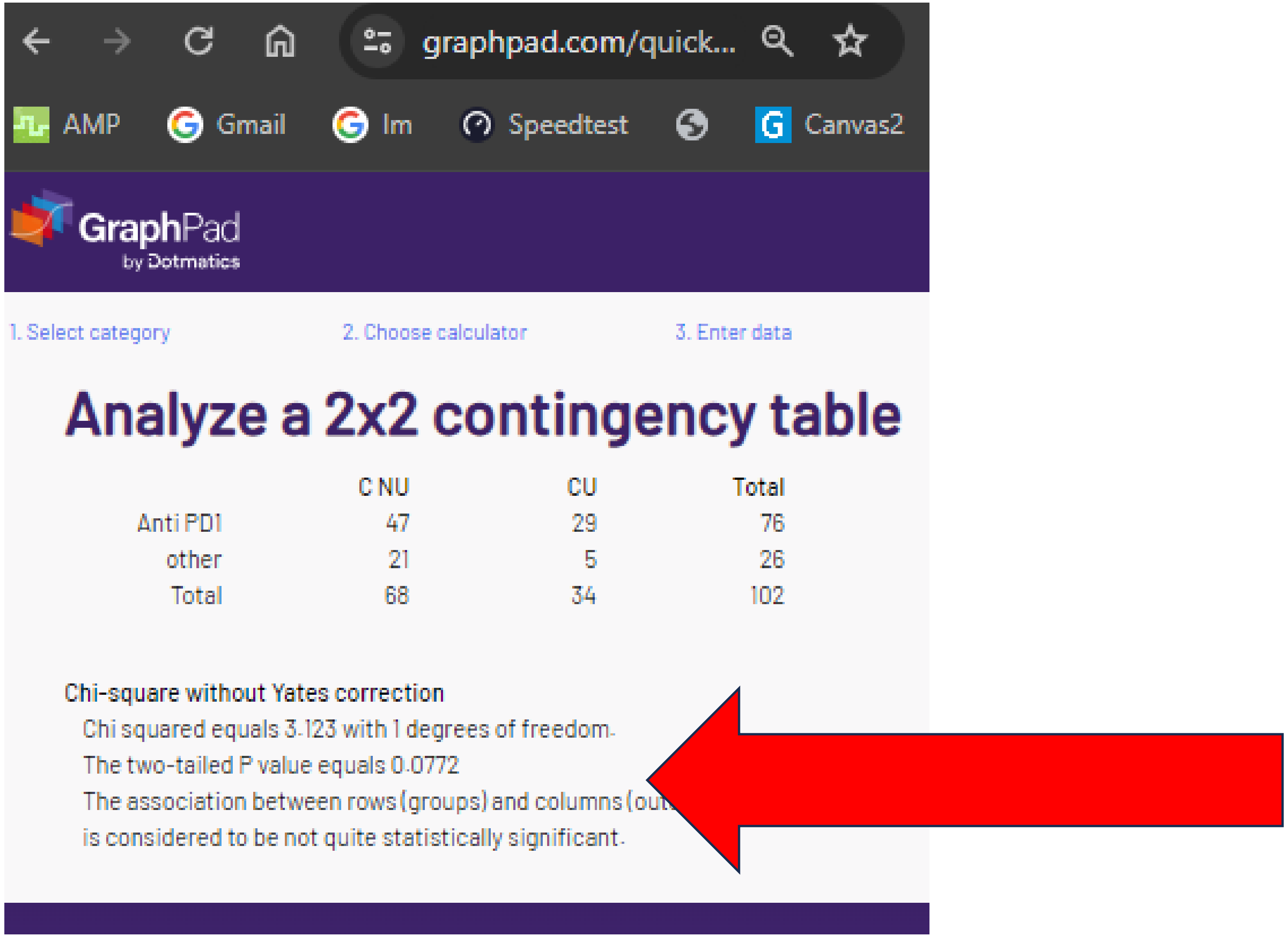

### T. “corrected” chi-sq or Fisher’s = .127

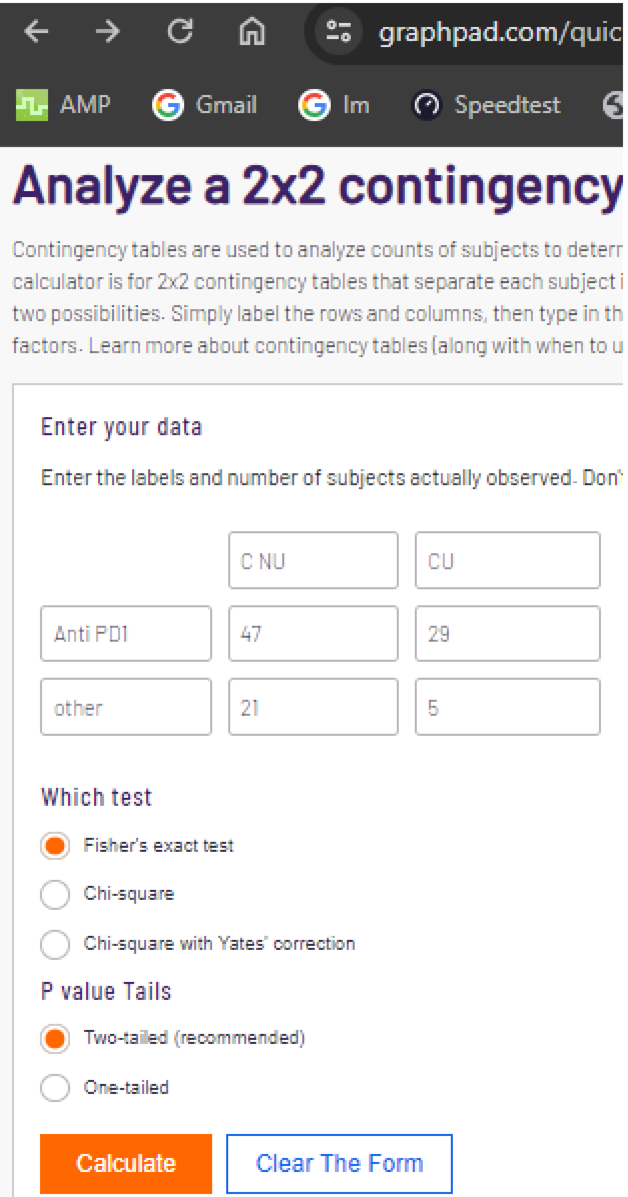


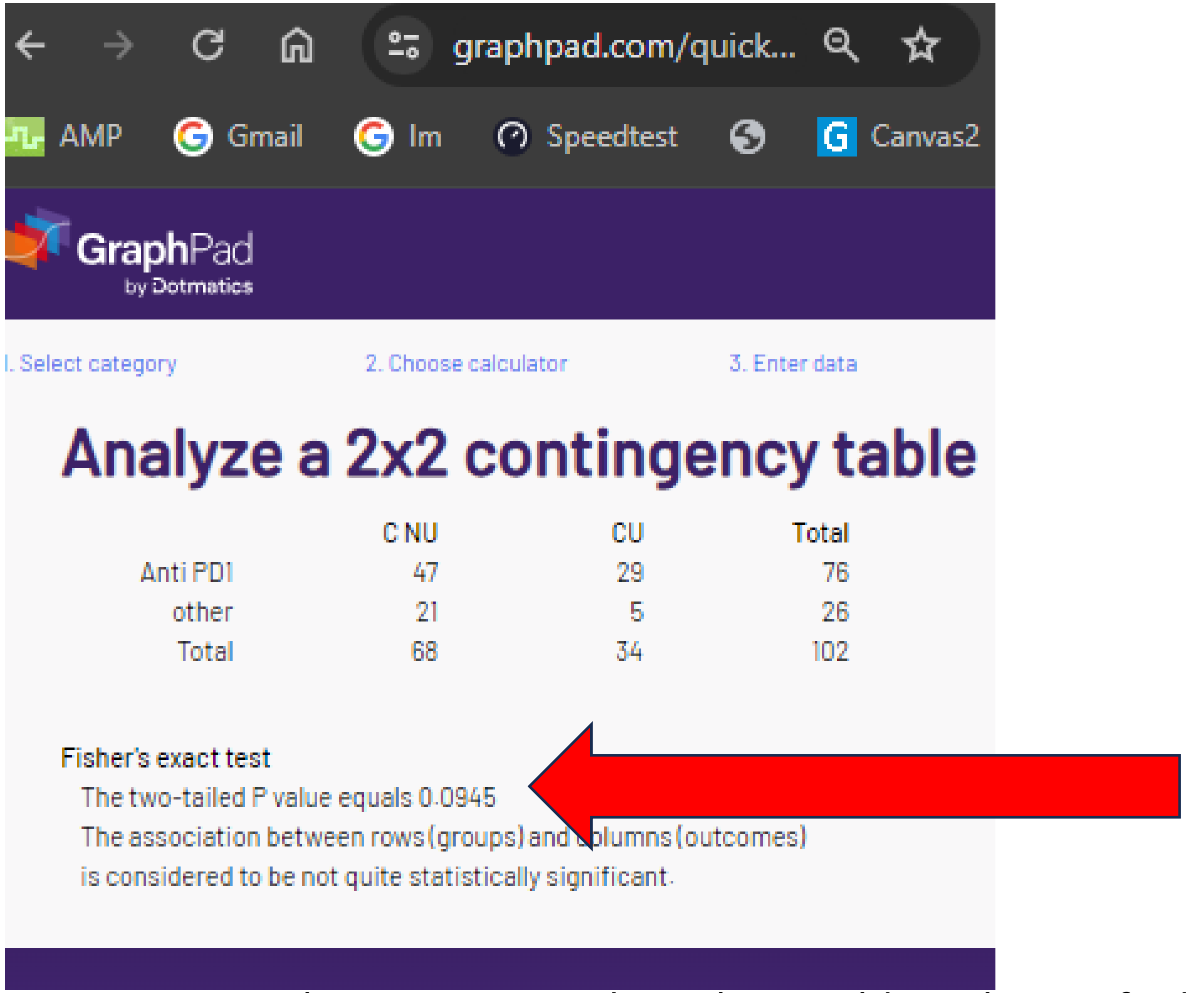


The for PD-1 immunotherapy reported p-value could not be verified with either chi-square or Fisher’s. A “trend” (p > .05 but p < .10) was identified with both statistics. However, this was not discussed in the manuscript as this near significant difference could change the interpretation of this immunotherapy study.

### T. “corrected” chi-sq or Fisher’s = .127

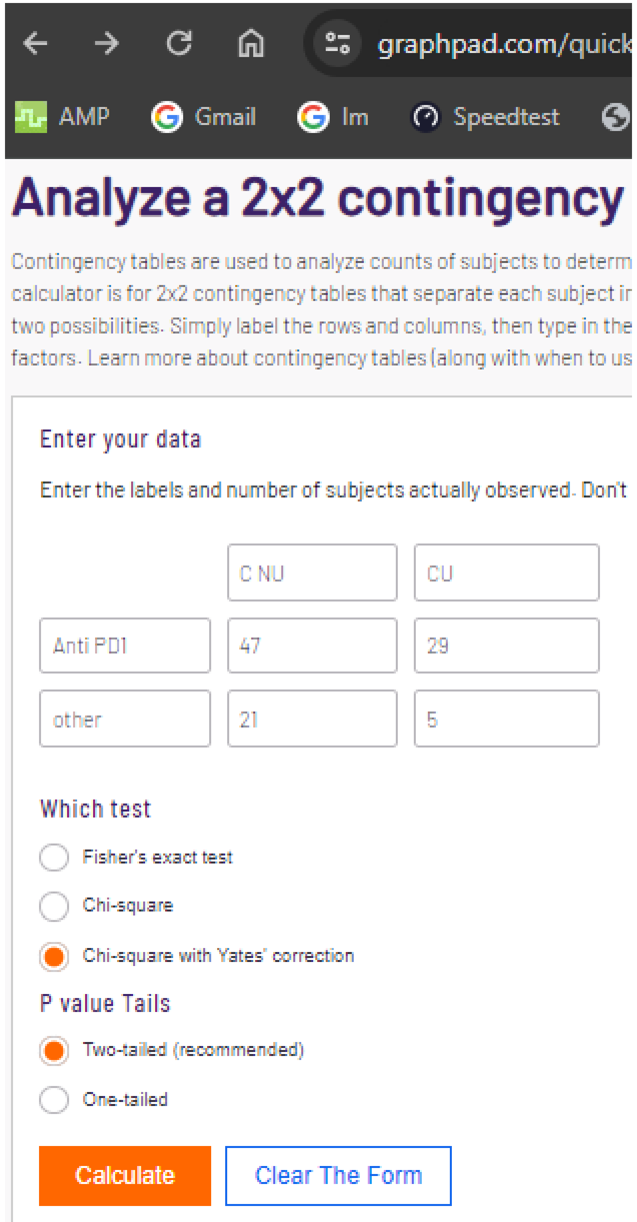


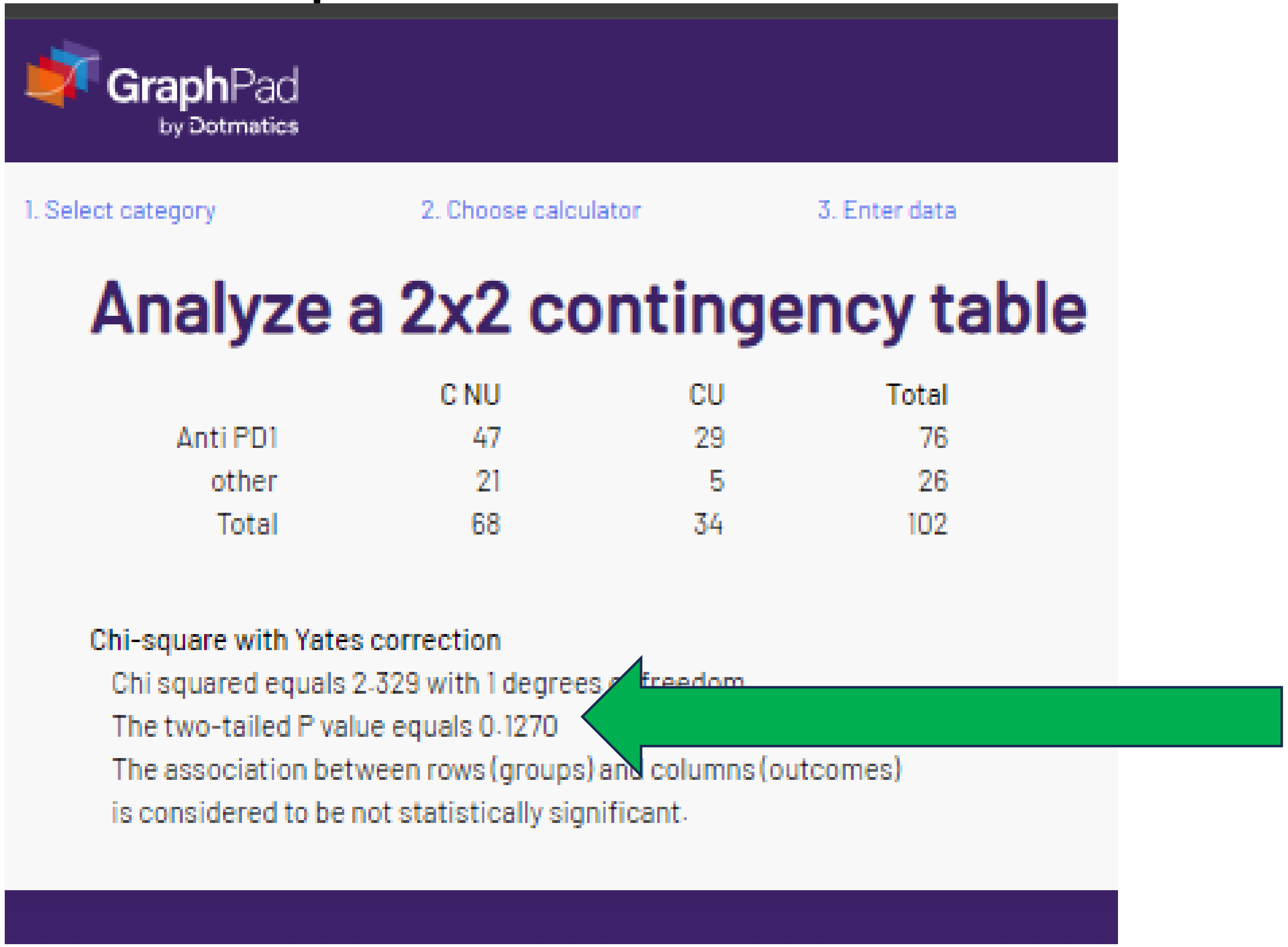


The smallest N per cell was 5 so Yates would (arguably) not be used.
However, the p-value was verified (again, with a statistic not listed in the methods section).

### U. “corrected” chi-sq or Fisher’s p = .2517

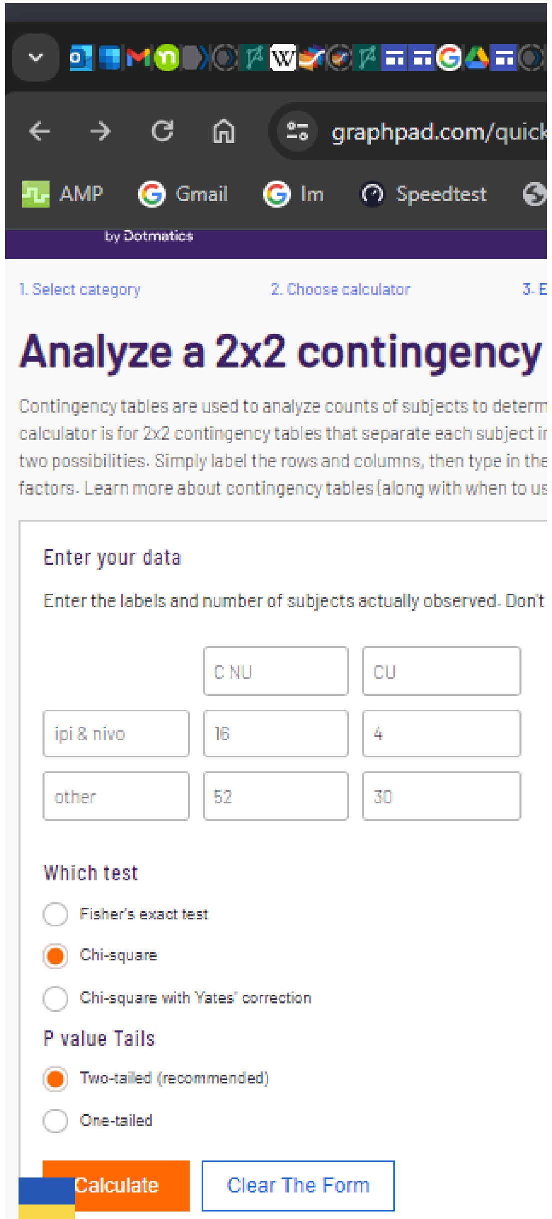


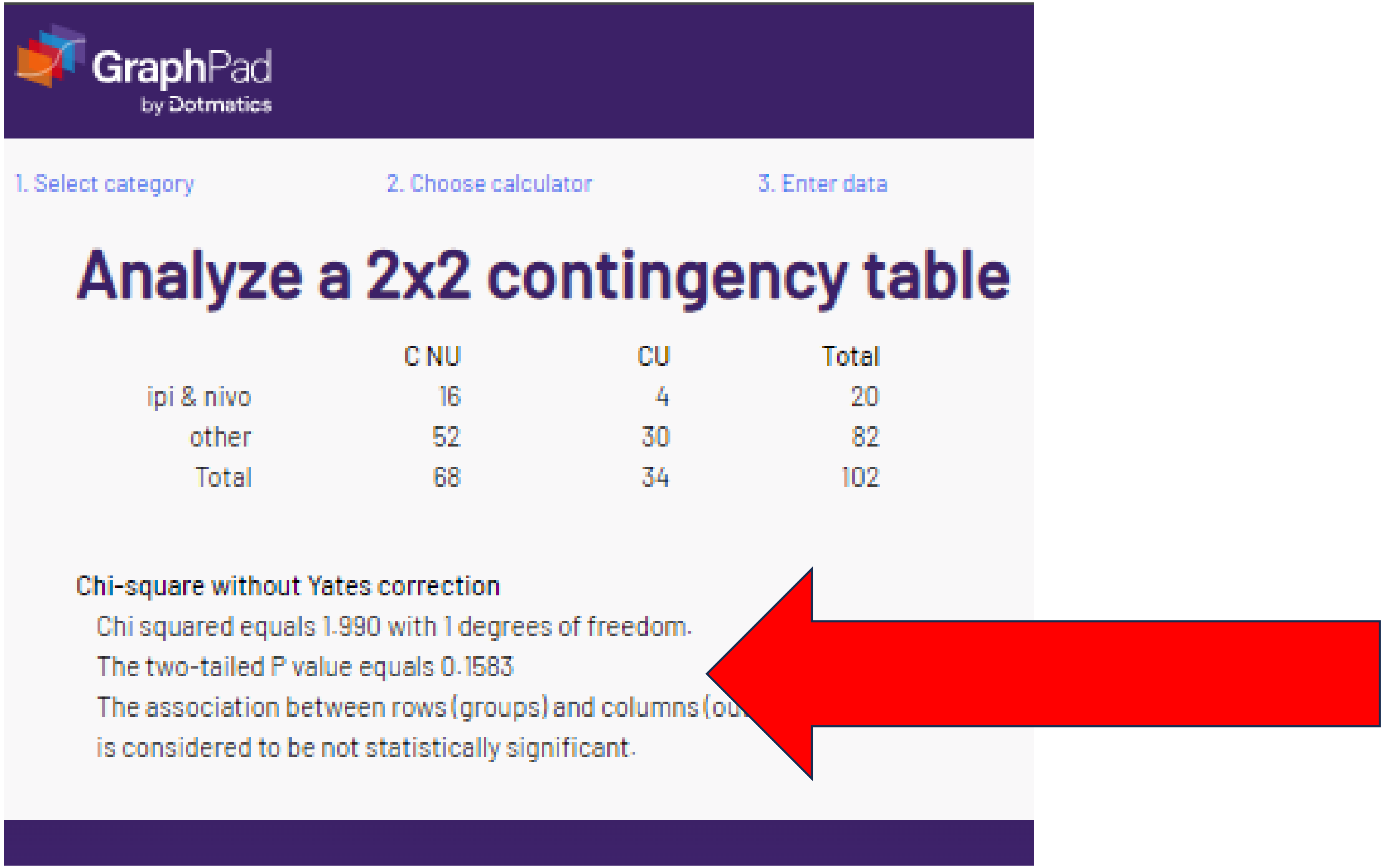

### U. "corrected" chi-sq or Fisher's p = .2517

#### Analyze a 2x2 contingency

Contingency tables are used to analyze counts of subjects to determ
calculator is for 2x2 contingency tables that separate each subject i
two possibilities. Simply label the rows and columns, then type in th
factors. Learn more about contingency tables (along with when to u

##### Enter your data

Enter the labels and number of subjects actually observed. Don't

| | C NU | CU |
|---|---|---|
| ipi & nivo | 16 | 4 |
| other | 52 | 30 |

**Which test**

- (selected) Fisher's exact test
- Chi-square
- Chi-square with Yates' correction

**P value Tails**

- (selected) Two-tailed (recommended)
- One-tailed

Calculate | Clear The Form

GraphPad
by Dotmatics

1. Select category 2. Choose calculator 3. Enter data

#### Analyze a 2x2 contingency table

| | C NU | CU | Total |
|---|---|---|---|
| ipi & nivo | 16 | 4 | 20 |
| other | 52 | 30 | 82 |
| Total | 68 | 34 | 102 |

**Fisher's exact test**

The two-tailed P value equals 0.1934

The association between rows (groups) a[illegible]olumns (outcomes)

is considered to be not statistically significant.

The reported p value for ipilimumab & nivolumab could not be verified with either chi-square or Fisher's.

### U. “corrected” chi-sq or Fisher’s p = .2517

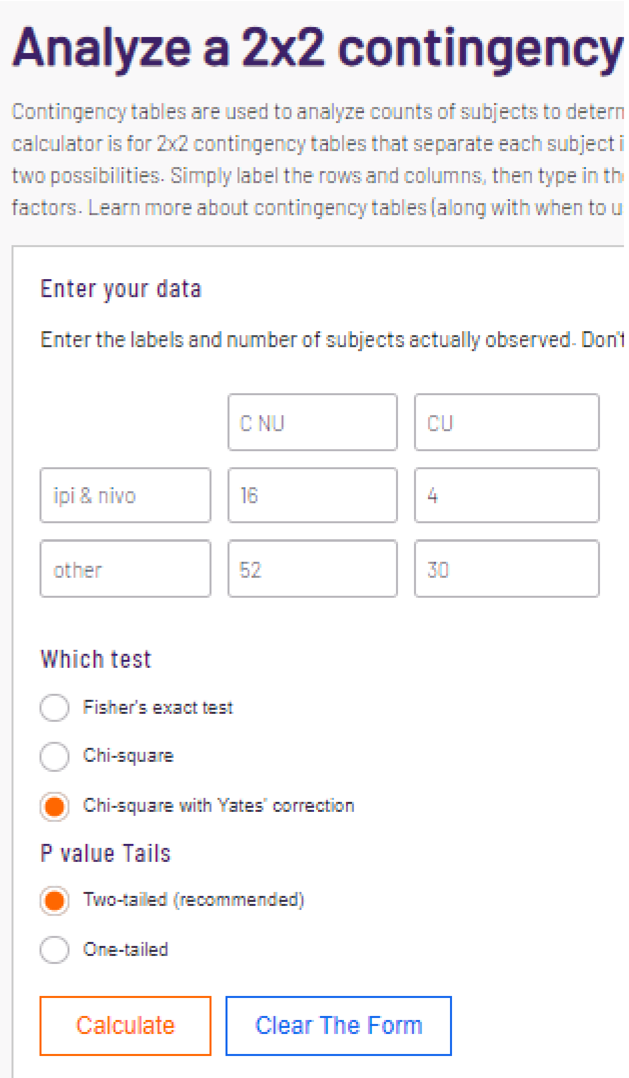


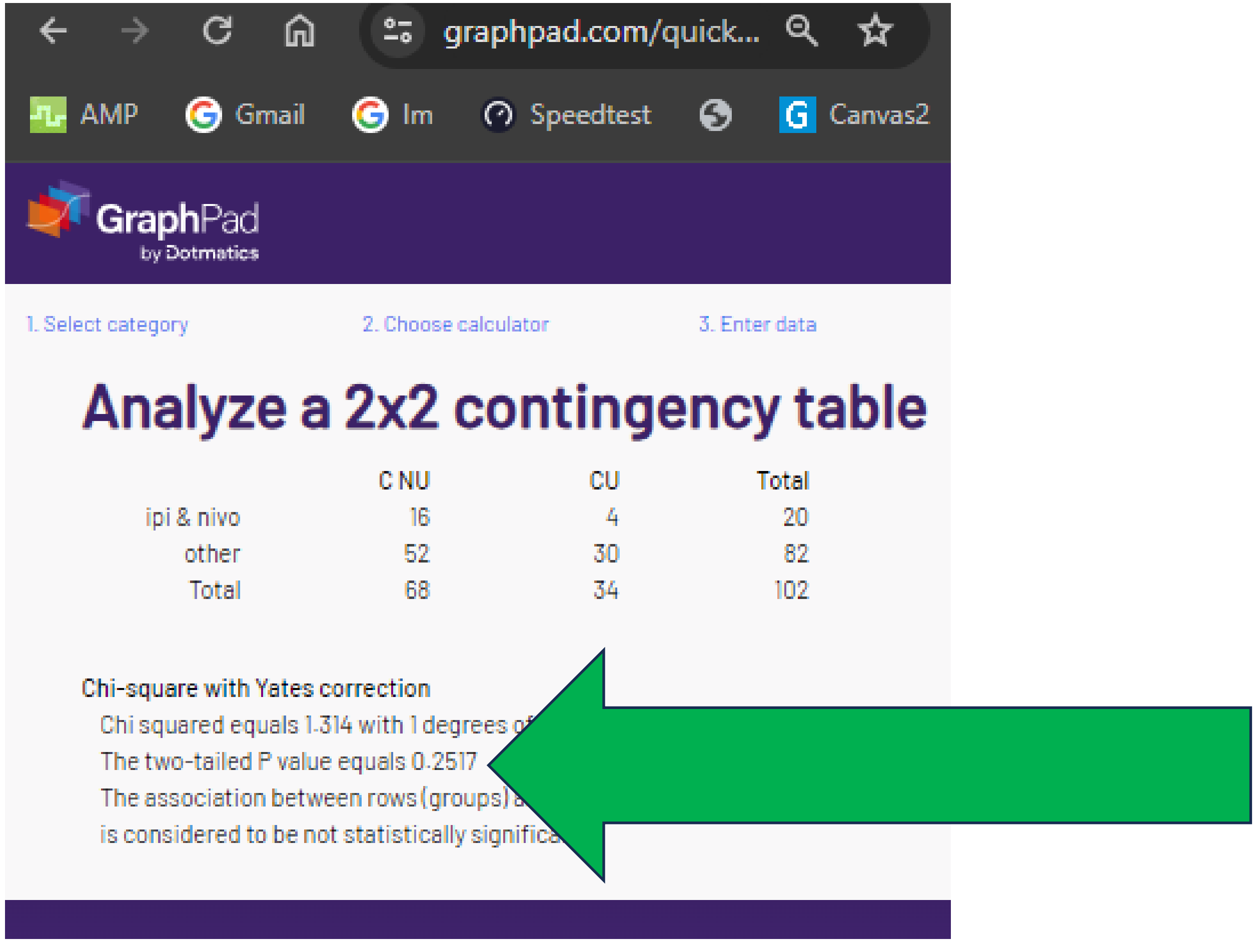


Although not listed in the methods section, due to the small N per cell (4), Yates correction would be appropriate here. The p value is verified.

### V. “Corrected” chi-sq or Fisher’s p = “1”

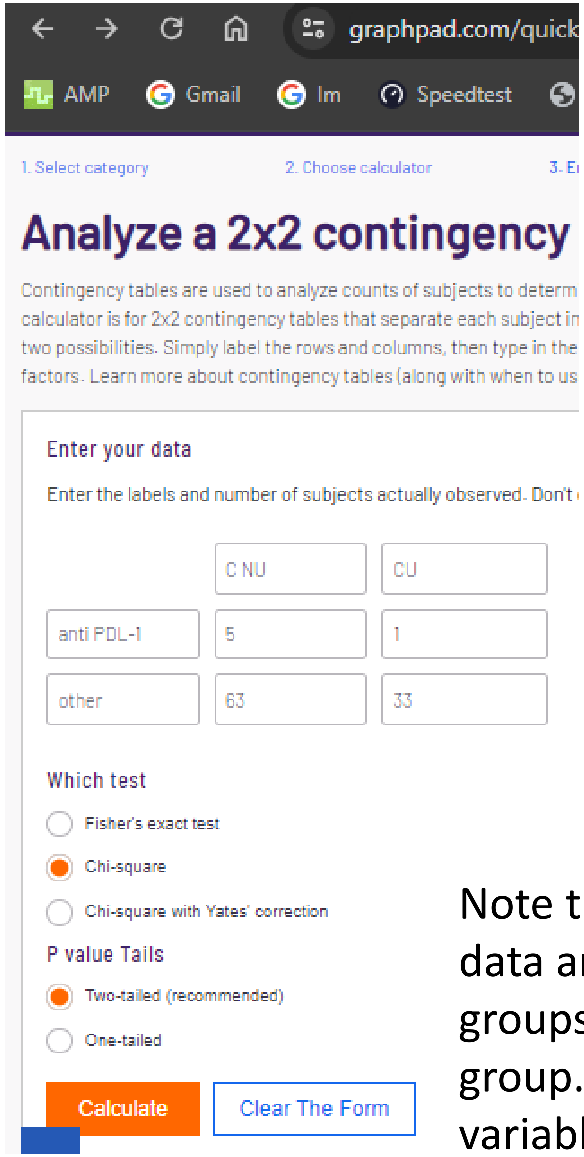


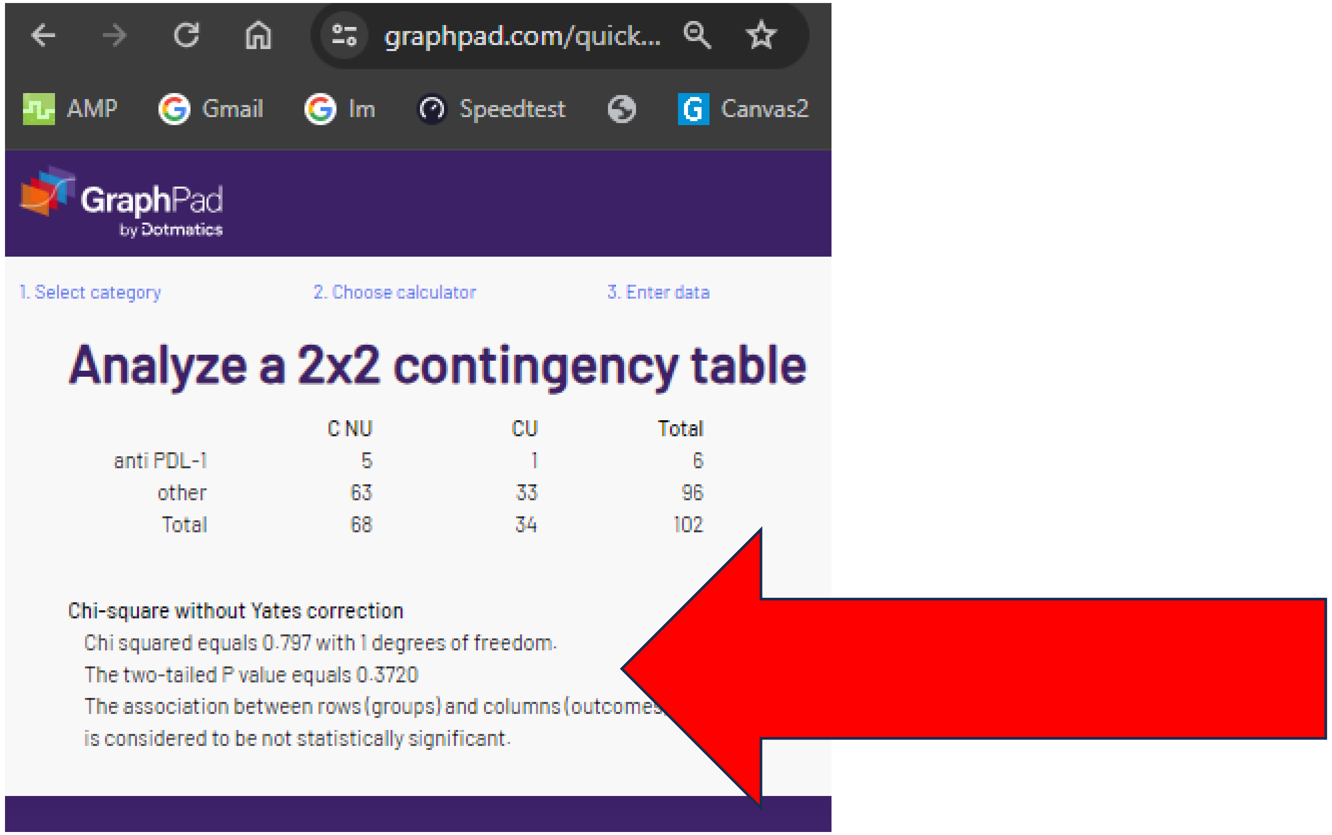


Note that a statistician that has worked with non-parametric data could just eyeball this data and intuitively know that the p value was **not** 1.000. That only happens when the % is equal for both groups. With 5 for 68 total in the CNU group, it would have to be 2.5 (i.e. half) per 34 total in the CU group. As this is an impossible outcome, there is clearly an error in the reporting for this immunotherapy variable.

### V. “Corrected” chi-sq or Fisher’s p = “1”

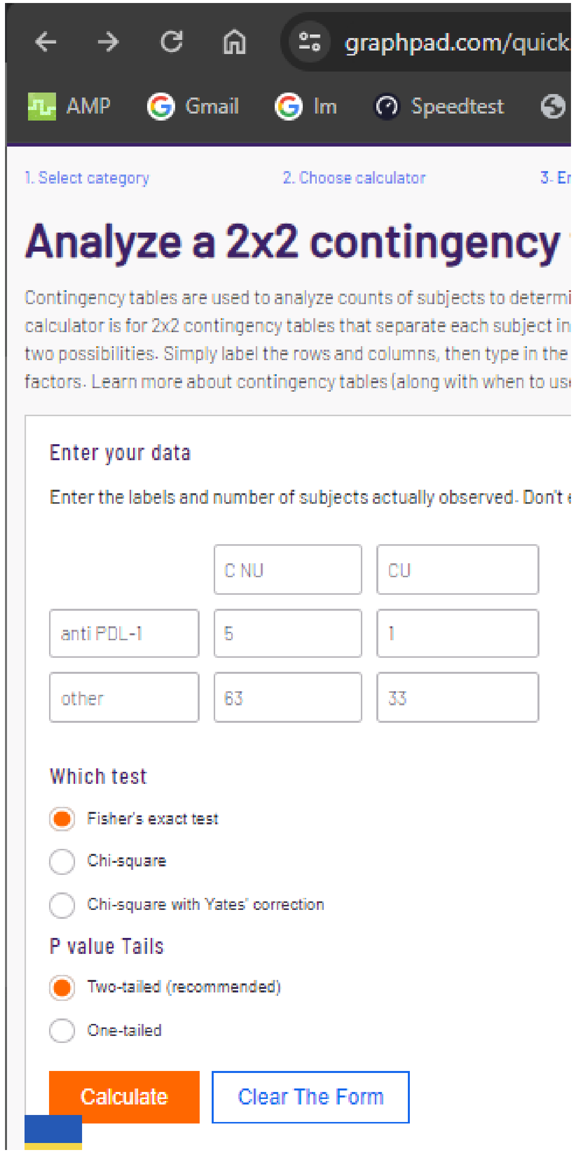


Select category 2. Choose calculator 3. Enter data

#### Analyze a 2x2 contingency table

| | C NU | CU | Total |
|---|---|---|---|
| anti PDL-1 | 5 | 1 | 6 |
| other | 63 | 33 | 96 |
| Total | 68 | 34 | 102 |

Fisher's exact test

The two-tailed P value equals 0.6607

The association between rows (groups) and columns (outcomes) is considered to be not statistically significant.

The “corrected” p value for anti-PDL-1 immunotherapy could **not** be verified with either chi-square or Fisher’s.

### V. “Corrected” chi-sq or Fisher’s p = “1”

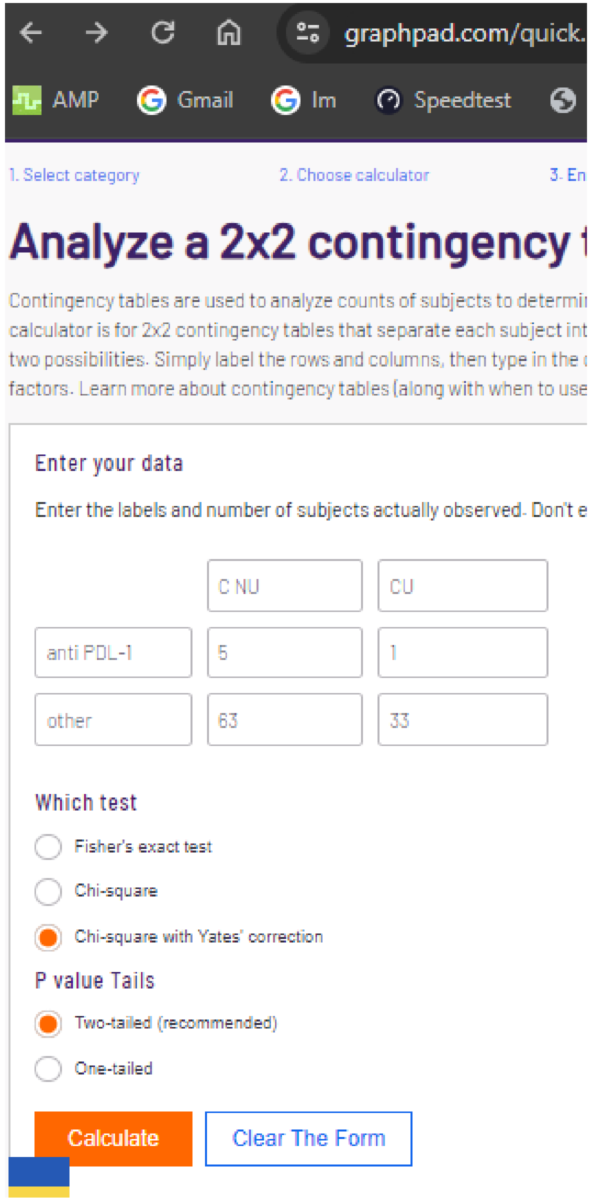


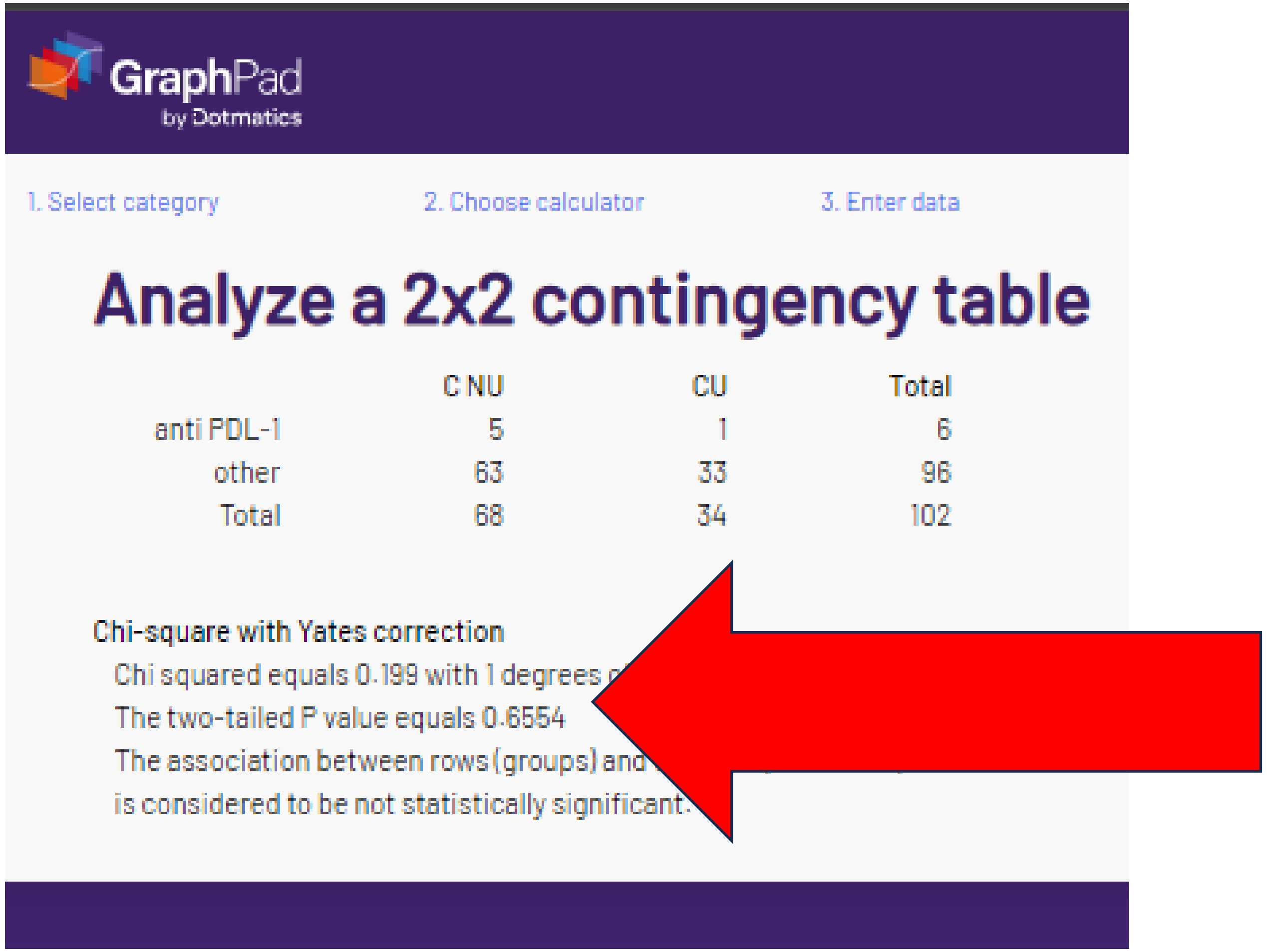


The reported p-value also was **not** verified with Yates.

### For further information

These concerns are also described at: https://pubpeer.com/publications/5AFA302155D8AE02603134F556A085?utm_source=Chrome&utm_medium=BrowserExtension&utm_campaign=Chrome

A series of $\chi^2$ tests or Fisher's exact tests (when the assumptions of the parametric $\chi^2$ test were not met) and nonparametric Mann–Whitney U tests were conducted to analyze the differences between patients' characteristics in both groups. Time to tumor progression (TTP) and overall survival (OS) was estimated using the Kaplan-Meier survival curve by group, and the log-rank test was computed to differentiate the survival curves between groups. Hazard ratios and the corresponding 95% CIs based on a Cox proportional hazards regression model were provided for multivariate analyses. We computed 2-tailed *p*-values, where $p < 0.05$ was considered a statistically significant result. Statistical analyses were performed using the SAS software package version 9.4 (SAS Institute, Cary, NC, USA).

### Note 2-tailed p values

Parametric: variables with normal distribution were compared by means of parametric tests, there is no such thing as a "parametric chi-square test"

### Summary of Table 2

- 0 of the 2 p values could be replicated despite trying 6 different ways
- 2 of the 12 percentage calculations contain rounding errors

**Table 2.** Abnormal laboratory tests before immunotherapy (according to local normal ranges).

| | Test | Cannabis: Nonusers $n = 68$ | Cannabis: Users $n = 34$ | $p$-Value |
|---|---|---|---|---|
| 1 | Lymphocytes ≤ 1.5 K/uL—N (%) | 35 (51) | 23 (67) 67.6=68 | 0.08 |
| | Blood count WBC ≤ 4.5 K/uL—N (%) | 7 (10) | 2 (6) | - |
| | Liver Function | | | |
| | Alanine Aminotransferase (ALT) > 45 | 7 (10) | 3 (9) | - |
| | Aspartate Aminotrasferase (AST) > 35 | 8 (12) | 5 (15) | - |
| 2 | Alkaline phosphatase level (ALKP) > 120 | 13 (19) | 11 (32) | 0.09 |
| | Renal Function—N (%) | | | |
| | Creatinine > 1.17 mL/min | 12 (17) 17.6 = 18 | 3 (9) | - |

Two-tailed
Ch-sq=.1199
F=.1412
Y=.1793
Ch-sq=.4590

Chi-sq=.1374
F=.1468
Y=.2157

One-tailed

1: C = .060, F = .0890, Y = .0896, 3: C= .0687, F = .1089, Y = .1079

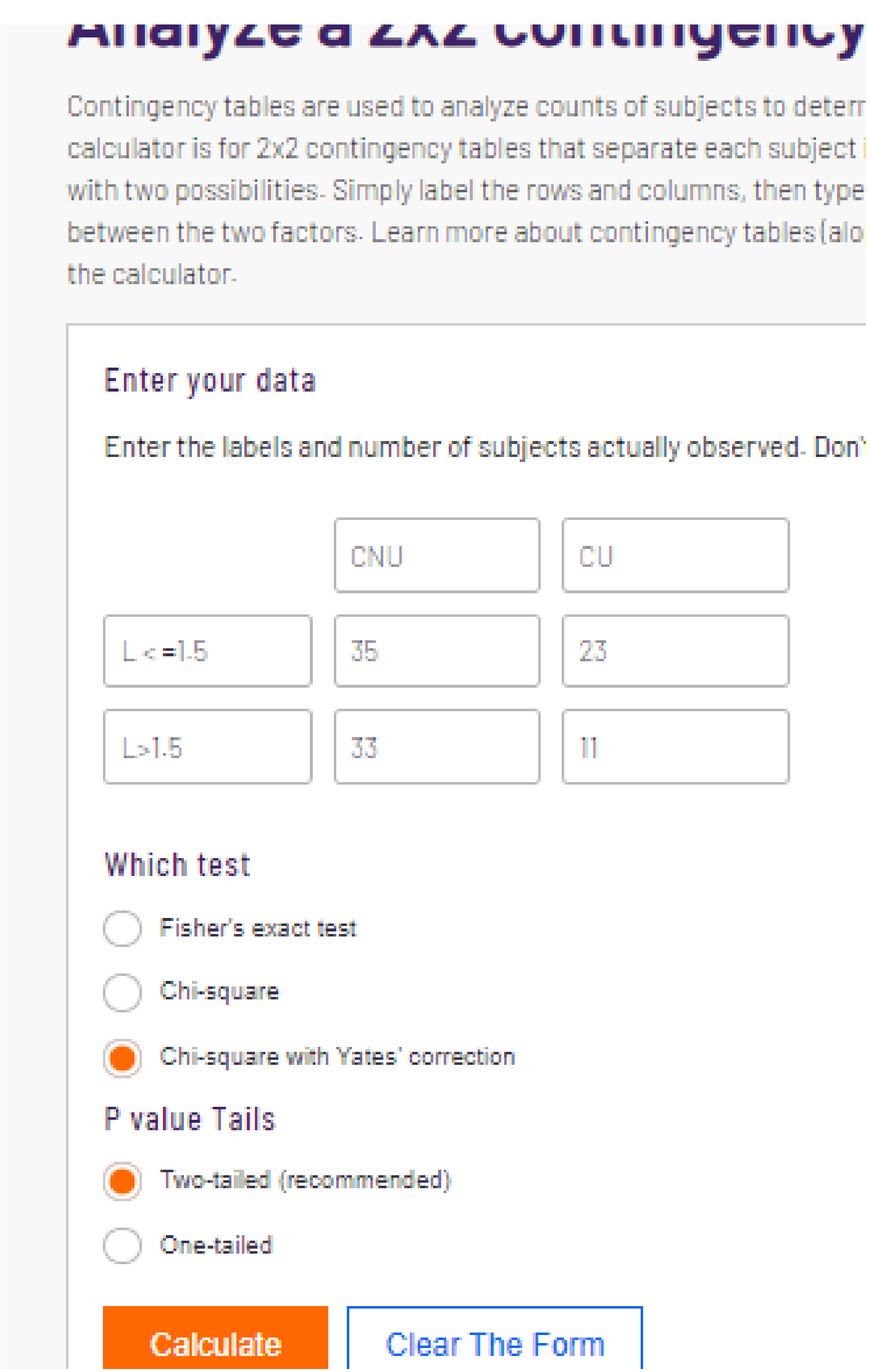
Analyze a 2x2 contingency
Contingency tables are used to analyze counts of subjects to deter
calculator is for 2x2 contingency tables that separate each subject
with two possibilities. Simply label the rows and columns, then type
between the two factors. Learn more about contingency tables (alo
the calculator.
Enter your data
Enter the labels and number of subjects actually observed. Don'
CNU
CU
L < =1.5
35
23
L>1.5
33
11
Which test
Fisher's exact test
Chi-square
Chi-square with Yates' correction
P value Tails
Two-tailed (recommended)
One-tailed
Calculate
Clear The Form

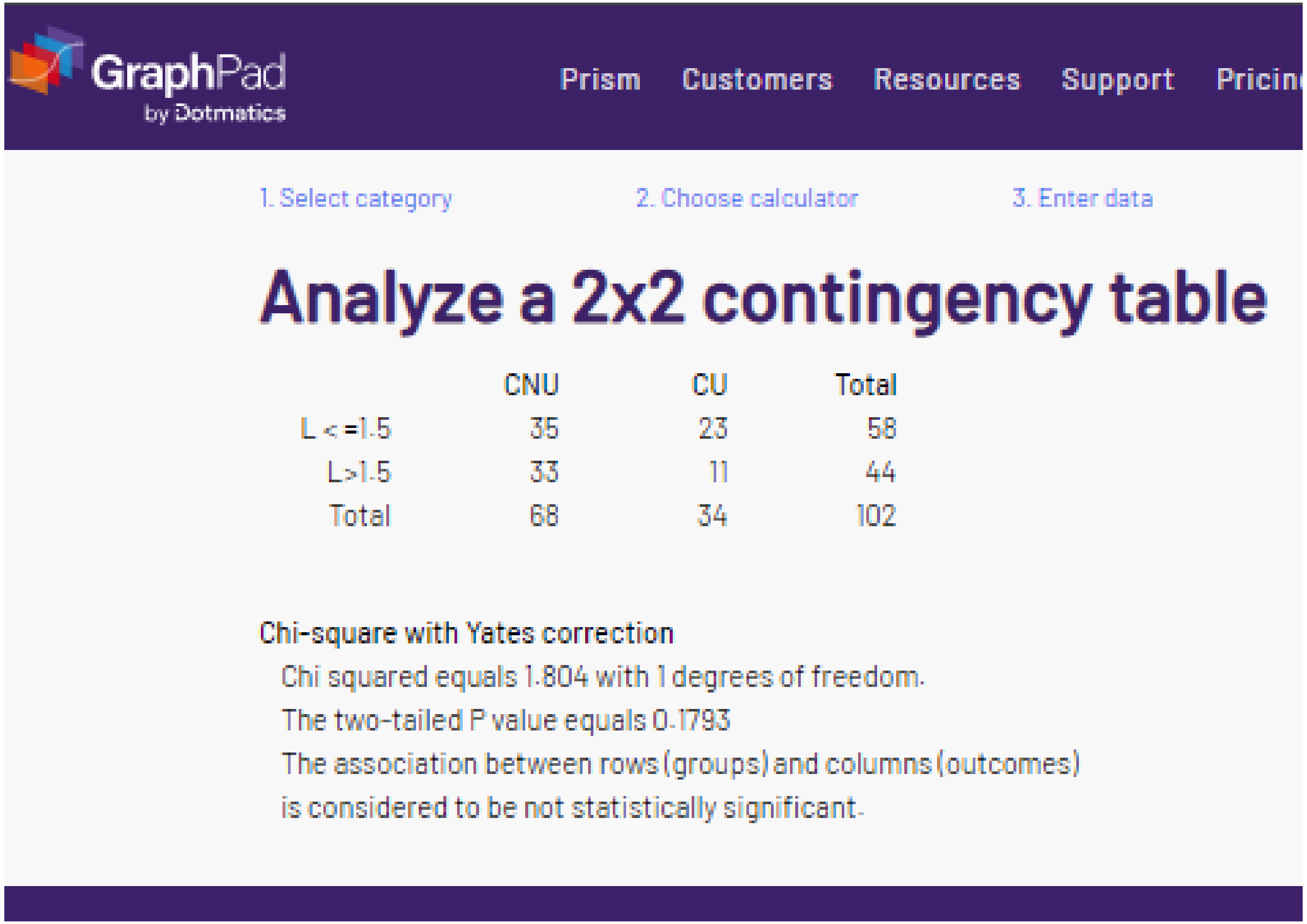
GraphPad
by Dotmatics
Prism
Customers
Resources
Support
Pricin
1. Select category
2. Choose calculator
3. Enter data
Analyze a 2x2 contingency table
CNU CU Total
L < =1.5 35 23 58
L>1.5 33 11 44
Total 68 34 102
Chi-square with Yates correction
Chi squared equals 1.804 with 1 degrees of freedom.
The two-tailed P value equals 0.1793
The association between rows (groups) and columns (outcomes)
is considered to be not statistically significant.

Contingency tables are used to analyze counts of subjects to deterr
calculator is for 2x2 contingency tables that separate each subject i
with two possibilities. Simply label the rows and columns, then type
between the two factors. Learn more about contingency tables (alo
the calculator.

##### Enter your data

Enter the labels and number of subjects actually observed. Don'

| | CNU | CU |
|---|---|---|
| L < =1.5 | 35 | 23 |
| L>1.5 | 33 | 11 |

##### Which test

- ( ) Fisher's exact test
- (•) Chi-square
- ( ) Chi-square with Yates' correction

##### P value Tails

- (•) Two-tailed (recommended)
- ( ) One-tailed

Calculate | Clear The Form

1. Select category 2. Choose calculator 3. Enter data

### Analyze a 2x2 contingency table

| | CNU | CU | Total |
|---|---|---|---|
| L < =1.5 | 35 | 23 | 58 |
| L>1.5 | 33 | 11 | 44 |
| Total | 68 | 34 | 102 |

Chi-square without Yates correction

Chi squared equals 2.418 with 1 degrees of freedom.

The two-tailed P value equals 0.1199

The association between rows (groups) and columns (outcomes) is considered to be not statistically significant.

#### Analyze a 2x2 continge[illegible]

Contingency tables are used to analyze counts of subjects to de[illegible] calculator is for 2x2 contingency tables that separate each subj[illegible] with two possibilities. Simply label the rows and columns, then [illegible] between the two factors. Learn more about contingency tables [illegible] the calculator.

##### Enter your data

Enter the labels and number of subjects actually observed. [illegible]

| | CNU | CU |
|---|---|---|
| L < =1.5 | 35 | 23 |
| L>1.5 | 33 | 11 |

##### Which test

- (●) Fisher's exact test
- ( ) Chi-square
- ( ) Chi-square with Yates' correction

##### P value Tails

- (●) Two-tailed (recommended)
- ( ) One-tailed

Calculate | Clear The Form

GraphPad by Dotmatics — Prism | Customers | Resources | Support | Pricing

1. Select category 2. Choose calculator 3. Enter data

### Analyze a 2x2 contingency table

| | CNU | CU | Total |
|---|---|---|---|
| L < =1.5 | 35 | 23 | 58 |
| L>1.5 | 33 | 11 | 44 |
| Total | 68 | 34 | 102 |

Fisher's exact test

The two-tailed P value equals 0.1412

The association between rows (groups) and columns (outcomes) is considered to be not statistically significant.

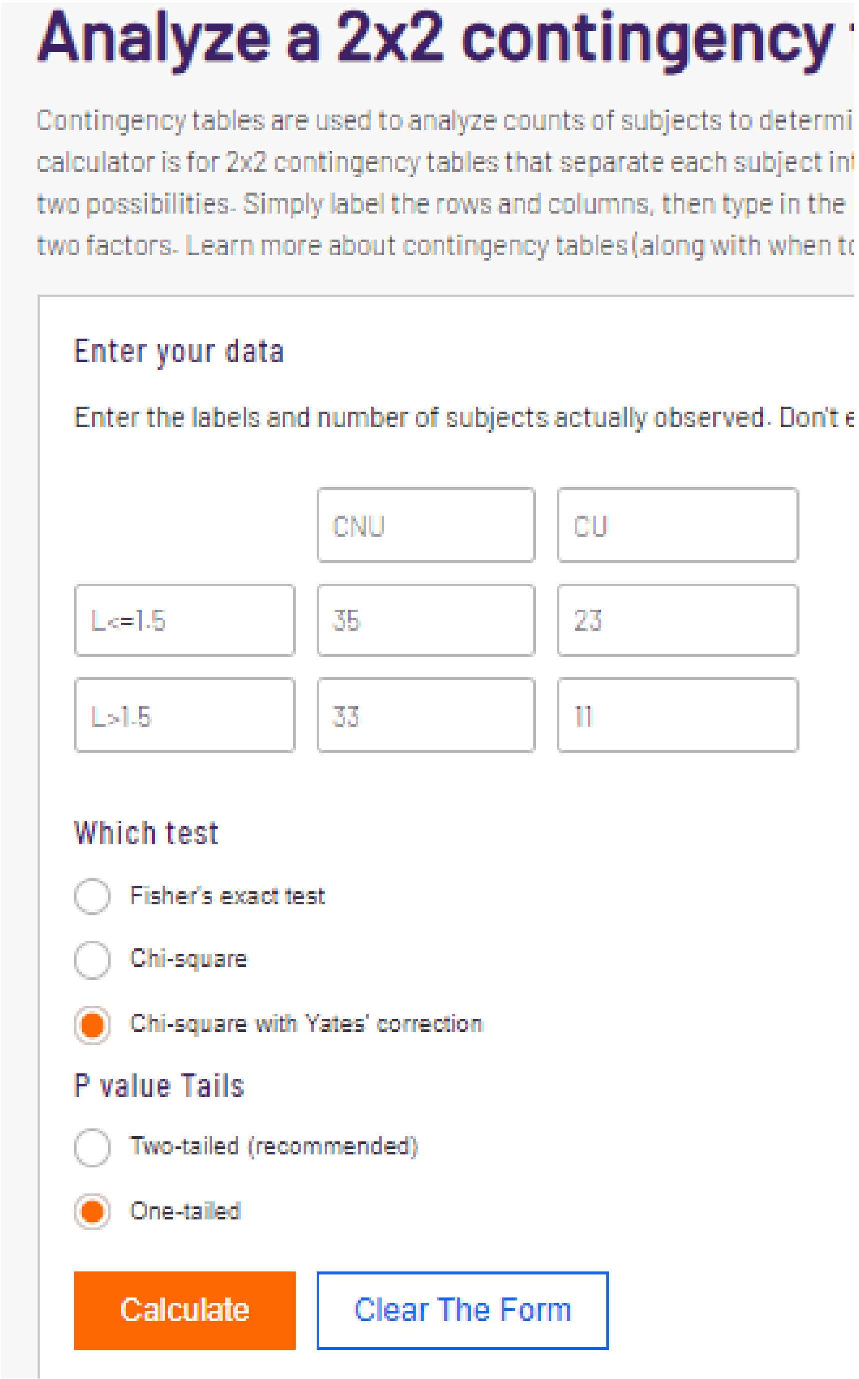


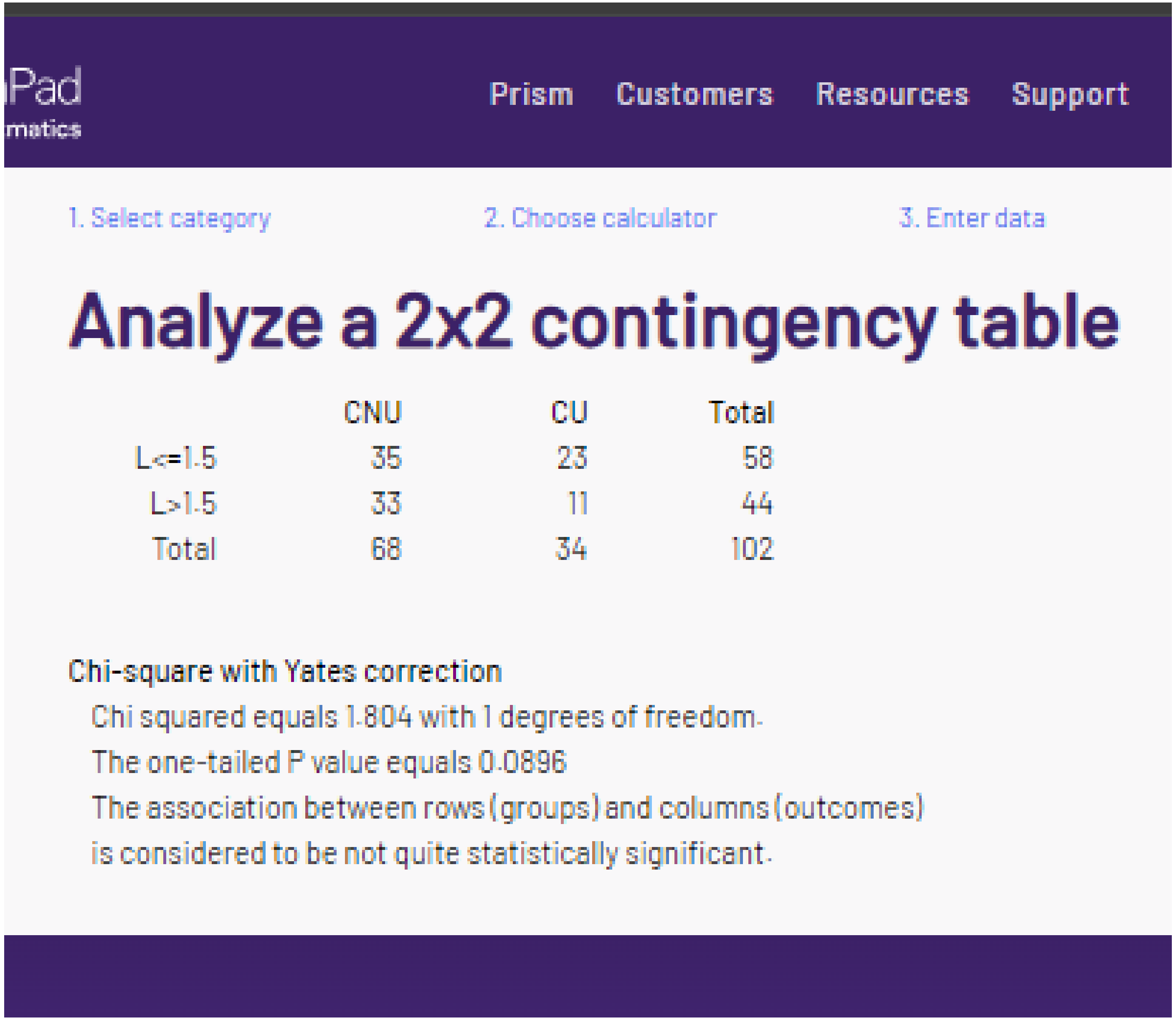


This p value <u>almost</u> corresponds with that reported in Table 2.

### Analyze a 2x2 contingency table

Contingency tables are used to analyze counts of subjects to determine if there is asso
calculator is for 2x2 contingency tables that separate each subject into one of four cate
with two possibilities. Simply label the rows and columns, then type in the counts for ea
between the two factors. Learn more about contingency tables (along with when to use
the calculator.

##### Enter your data

Enter the labels and number of subjects actually observed. Don't enter proportions,

| | | |
|---|---|---|
| | CNU | CU |
| ALKP > 120 | 13 | 11 |
| ALKP<=120 | 55 | 23 |

##### Which test

- ○ Fisher's exact test
- ○ Chi-square
- ● Chi-square with Yates' correction

##### P value Tails

- ○ Two-tailed (recommended)
- ● One-tailed

Calculate | Clear The Form

GraphPad by Dotmatics

Prism | Customers | Resources | Support | Pric

1. Select category | 2. Choose calculator | 3. Enter data

### Analyze a 2x2 contingency table

| | CNU | CU | Total |
|---|---|---|---|
| ALKP > 120 | 13 | 11 | 24 |
| ALKP<=120 | 55 | 23 | 78 |
| Total | 68 | 34 | 102 |

**Chi-square with Yates correction**

Chi squared equals 1.532 with 1 degrees of freedom.

The one-tailed P value equals 0.1079

The association between rows (groups) and columns (outcomes)
is considered to be not statistically significant.

Analyze a 2x2 continge

Contingency tables are used to analyze counts of subjects t
factors. This calculator is for 2x2 contingency tables that se
based on two factors, each with two possibilities. Simply lab
for each cell to test the relationship between the two factors
when to use each test) in the description below the calculat

Enter your data

Enter the labels and number of subjects actually observe
means.

| | CNU | CU |
|---|---|---|
| WBC<=4.5 | 7 | 2 |
| WBC>4.5 | 61 | 32 |

Which test

- Fisher's exact test
- Chi-square (selected)
- Chi-square with 'Yates' correction

P value Tails

- Two-tailed (recommended) (selected)
- One-tailed

Calculate | Clear The Form

GraphPad by Dotmatics

1. Select category  2. Choose calculator  3. Enter data  4. Vi

### Analyze a 2x2 contingency table

| | CNU | CU | Total |
|---|---|---|---|
| WBC<=4.5 | 7 | 2 | 9 |
| WBC>4.5 | 61 | 32 | 93 |
| Total | 68 | 34 | 102 |

Chi-square without Yates correction

Chi squared equals 0.548 with 1 degrees of freedom.

The two-tailed P value equals 0.4590

The association between rows (groups) and columns (outcomes) is considered to be not statistically significant.

### Analyze a 2x2 contingenc

Contingency tables are used to analyze counts of subjects to de
actors. This calculator is for 2x2 contingency tables that separ
ased on two factors, each with two possibilities. Simply label t
or each cell to test the relationship between the two factors. L
vhen to use each test) in the description below the calculator.

##### Enter your data

Enter the labels and number of subjects actually observed. I
means.

| | | |
|---|---|---|
| | CNU | CU |
| ALKP>120 | 13 | 11 |
| ALKP<=120 | 55 | 23 |

##### Which test

- ○ Fisher's exact test
- ● Chi-square
- ○ Chi-square with Yates' correction

##### P value Tails

- ● Two-tailed (recommended)
- ○ One-tailed

Calculate | Clear The Form

GraphPad
by Dotmatics

1. Select category 2. Choose calculator 3. Enter data 4. Vie

### Analyze a 2x2 contingency table

| | CNU | CU | Total |
|---|---|---|---|
| ALKP>120 | 13 | 11 | 24 |
| ALKP<=120 | 55 | 23 | 78 |
| Total | 68 | 34 | 102 |

**Chi-square without Yates correction**

Chi squared equals 2.207 with 1 degrees of freedom.

The two-tailed P value equals 0.1374

The association between rows (groups) and columns (outcomes)
is considered to be not statistically significant.

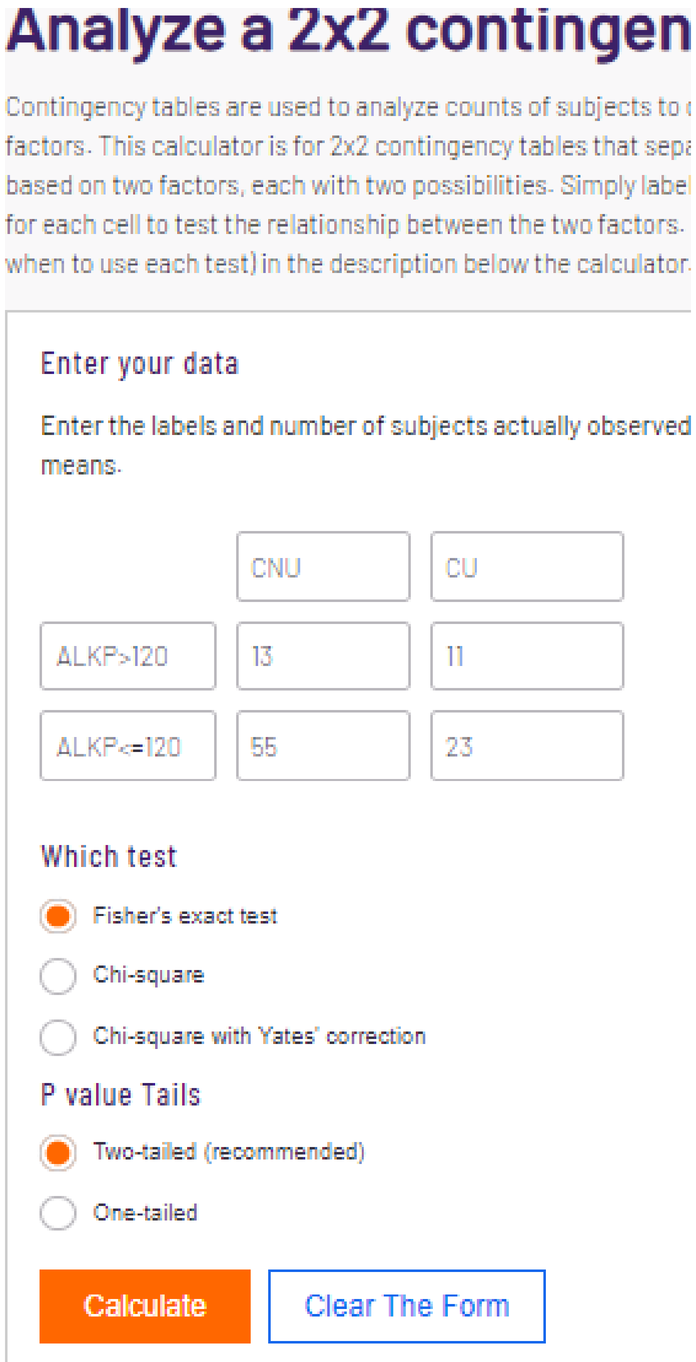
Analyze a 2x2 contingen
Contingency tables are used to analyze counts of subjects to
factors. This calculator is for 2x2 contingency tables that sepa
based on two factors, each with two possibilities. Simply label
for each cell to test the relationship between the two factors.
when to use each test) in the description below the calculator.
Enter your data
Enter the labels and number of subjects actually observed
means.
CNU
CU
ALKP>120
13
11
ALKP<=120
55
23
Which test
Fisher's exact test
Chi-square
Chi-square with Yates' correction
P value Tails
Two-tailed (recommended)
One-tailed
Calculate
Clear The Form

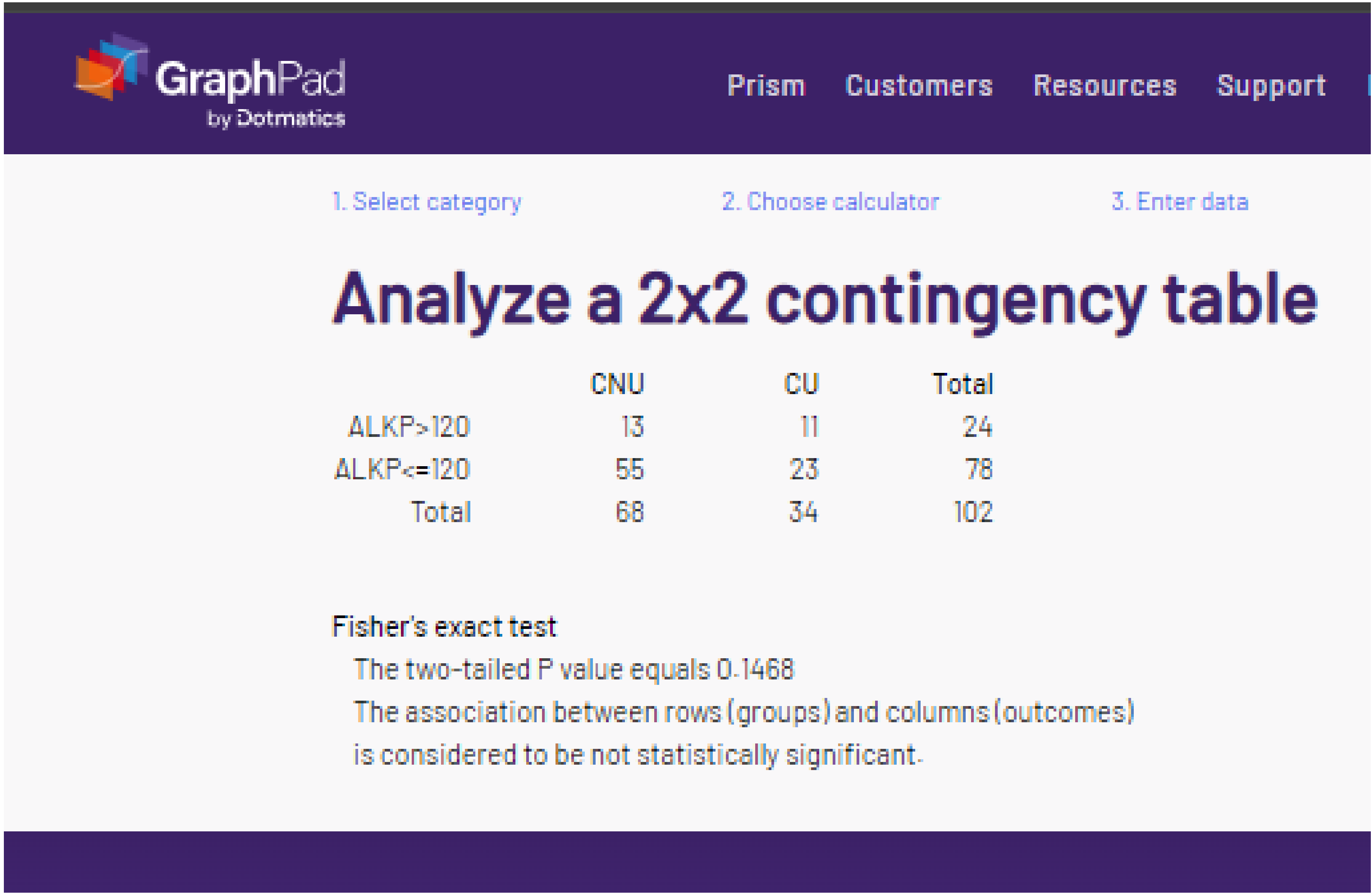
GraphPad
by Dotmatics
Prism
Customers
Resources
Support
1. Select category
2. Choose calculator
3. Enter data
Analyze a 2x2 contingency table
CNU CU Total
ALKP>120 13 11 24
ALKP<=120 55 23 78
Total 68 34 102
Fisher's exact test
The two-tailed P value equals 0.1468
The association between rows (groups) and columns (outcomes)
is considered to be not statistically significant.

### Analyze a 2x2 contingency table

Contingency tables are used to analyze counts of subjects to determine if there is assoc
calculator is for 2x2 contingency tables that separate each subject into one of four categ
two possibilities. Simply label the rows and columns, then type in the counts for each ce
two factors. Learn more about contingency tables (along with when to use each test) in t

#### Enter your data

Enter the labels and number of subjects actually observed. Don't enter proportions, p

| | CNU | CU |
|---|---|---|
| ALKP>120 | 13 | 11 |
| ALKP<=120 | 55 | 23 |

#### Which test

- ○ Fisher's exact test
- ○ Chi-square
- ● Chi-square with Yates' correction

#### P value Tails

- ● Two-tailed (recommended)
- ○ One-tailed

Calculate | Clear The Form

GraphPad by Dotmatics — Prism | Customers | Resources | Support

1. Select category 2. Choose calculator 3. Enter data

### Analyze a 2x2 contingency table

| | CNU | CU | Total |
|---|---|---|---|
| ALKP>120 | 13 | 11 | 24 |
| ALKP<=120 | 55 | 23 | 78 |
| Total | 68 | 34 | 102 |

**Chi-square with Yates correction**

Chi squared equals 1.532 with 1 degrees of freedom.
The two-tailed P value equals 0.2157
The association between rows (groups) and columns (outcomes)
is considered to be not statistically significant.
