## Supplementary material for "Immunotherapy and Cannabis: A Harmful Drug Interaction or Reefer Madness?": Taha statistical concerns

"Cannabis Impacts Tumor Response Rate to Nivolumab in Patients with Advanced Malignancies" has comments on PubPeer



Observational Study > Oncologist. 2019 Apr;24(4):549-554.
doi: 10.1634/theoncologist.2018-0383. Epub 2019 Jan 22.

### Cannabis Impacts Tumor Response Rate to Nivolumab in Patients with Advanced Malignancies

Tarek Taha [1], David Meiri [2] [3], Samira Talhamy [3], Mira Wollner [1], Avivit Peer [1], Gil Bar-Sela [4] [3]

Affiliations + expand

PMID: 30670598

PMCID: PMC6459234

1 comment on PubPeer (by: Phlegmariurus Pittieri)

1 comment on PubPeer (by: Phlegmariurus Pittieri)

DOI: 10.1634/theoncologist.2018-0383

FULL TEXT LINKS

OXFORD ACADEMIC

FREE Full text PMC

ACTIONS

Cite

Collections

SHARE

https://pubpeer.com/publications/85A8F34A338C7E670E8086BBAA31F4?utm_source=Chrome&utm_medium=BrowserExtension&utm_campaign=Chrome

**Table 2.** The demographic and medical characteristics of the study sample ($n$ = 140)

| Characteristics | Immunotherapy ($n$ = 89) | Immunotherapy + cannabis ($n$ = 51) | |
|---|---|---|---|
| Age, years | 67.7 ± 10.2 | 62 ± 10.6 | t(138)=3.1368, p=.0021 |
| Gender | | | |
| Male | 66 (74.2) | 40 (78.4) | |
| Female | 23 (25.8) | 11 (21.6) | |
| Type of disease | | | Chi-sq = 4.126, p=.0422 |
| Non-small cell cancer | 57 (64) | 41 (80.4) | CS with Y=3.384, p=.0658<br>Fisher's p=.0551 |
| Histology subtype | | | |
| Adeno | 43 (75) | 29 (71) 56.9% | |
| Squamous | 14 (25) 15.7%* | 12 (29) 23.5% | Chi-sq=33.177,p<.0001 |
| Other malignancies | 32 (36) | 10 (19.6) | Chi-sq=4.126, = p=.0422 |

3x2 chi-sq = 4.4105, p=.110221

The study includes "As shown in Table 2, no significant difference was found between the two groups in aspects of demographic and medical characteristics. Using their information, there are 4 significant group differences in these variables (red font). Using the N provided, the percentages (3) are also not accurate (e.g. *14/89 = 15.7 or 16%, not 25).

Enter up to 50 rows

Enter or paste up to 2000 rows

Enter mean, SEM and N

Enter mean, SD and N

Unpaired *t* test

Welch's unpaired *t* test (used

(You can only choose a paired *t* tes
enter individual values.)

##### 3. Enter data

Help me arrange the data

| | | |
|---|---|---|
| Label: | Immu | Immu + Can |
| Mean: | 67.7 | 62 |
| SD: | 10.2 | 10.6 |
| N: | 89 | 51 |

##### 4. View the results

Calculate Now

Clear Form

#### Unpaired *t* test results

**P value and statistical significance:**

The two-tailed P value equals 0.0021

By conventional criteria, this difference is considered to be very statistically significant.

**Confidence interval:**

The mean of Immu minus Immu + Can equals 5.700

95% confidence interval of this difference: From 2.107 to 9.293

**Intermediate values used in calculations:**

t = 3.1368

df = 138

standard error of difference = 1.817

**Review your data:**

| Group | Immu | Immu + Can |
|---|---|---|
| Mean | 67.700 | 62.000 |
| SD | 10.200 | 10.600 |
| SEM | 1.081 | 1.484 |
| N | 89 | 51 |

Here are the results of the age comparison. Note that the Immunotherapy + Cannabis was not reported to the tenths (62). As this team has difficulties with rounding, if a value of 62.9999 is entered, the p value is still significant (.0107).

https://www.graphpad.com/quickcalcs/ttest1/?format=SD

| NSCLC mutational status | N = 89 | | N = 51 | |
|---|---|---|---|---|
| EGFR mutation | 6 (10) | 6/57 = 10.5%=11%[1] | 1 (2) | |
| ALK | 0 (0) | | 0 (0) | |
| KRAS | 1 (1) | | 0 (0) | |
| No mutation | 50 (87) | 50/57=87.7=88% | 40 (97) | 40/41=97.56=98%? |
| Metastasis | | | | |
| Brain | 7 (19.1) | | 9 (17.6) | |
| Mediastinum | 55 (61.8) | | 22 (43.1) | |
| Liver | 13 (14.6) | | 7 (13.7) | |
| Adrenal | 21 (23.6) | | 10 (19.6) | |
| Bones | 28 (31.5) | | 21 (41.2) | |
| Other | 23 (25.8) | | 11 (21.6) | |
| Immunotherapy given as: | | | | |
| First line | 15 (16.8) | | 1 (2) | |
| Second line | 42 (47.2) | | 28 (54.9) | |
| Third line | 32 (36) | | 22 (43.1) | |

1. 6/89 = 6.7% or 7. However, 6 per 57 (total of all 4 groups) = 10.52% or 11. Although very minor, there appear to be 3 errors on this portion of the table

| | N = 89 | N = 51 | |
|---|---|---|---|
| Date of diagnosis of metastatic disease | | | |
| 2007–2012 | 12 (13.5) | 6 (12.2) | |
| 2013–2014 | 27 (30.3) | 14 (28.6) | |
| 2015–2016 | 50 (56.2) | 29 (59.2) | |
| Smoking | | | |
| No | 53 (59.6) | 22 (43.1) | chi-sq=3.512,p=.0609 |
| Yes | 36 (40.4) | 29 (56.9) | |
| NSCLC + smoking | 30/36 (83) | 25 (86) | Total = 29? |
| PS prior treatment | | | |
| 0 | 18 (26.1) | 8 (21.6) | |
| 1 | 28 (40.6) | 13 (35.1) | |
| 2 | 17 (24.6) | 10 (27) | |
| 3/4 | 6 (8.7) | 6 (16.2) | |

Data are presented as *n* (%).
Abbreviations: ALK, anaplastic lymphoma kinase; EGFR, epidermal growth factor receptor; NSCLC, non-small cell lung cancer; PS, performance status.

The chi-square on this is at the trend level. Although it is at the discretion of the lab, smoking is obviously a very important variable in oncology and this trend (arguably) should have been reported.

**Table 1.** Cannabis characteristics using high-performance liquid chromatography-diode array detector identification of phytocannabinoids for 37/59 patients

| Cannabis type | PR/CR/SD (*n* = 9) | PD (*n* = 28) | *p* value |
|---|---|---|---|
| THC ≥10 | 8 (29.6) | 19 (70.4) | .393 |
| THC <10 | 1 (10) | 9 (90) | |
| CBD ≥1 | 4 (16) | 21 (84) | .116 |
| CBD <1 | 5 (41.7) | 7 (58.3) | |

Data are presented as *n* (%).

Abbreviations: CBD, cannabidiol; CR, complete response; PD, progressive disease; PR, partial response; SD, stable disease; THC, tetrahydrocannabinol.

Chi-sq p = .2165
Chi-sq Yates
P = .4211
Fisher's p=.3932

Chi-sq p=.0885
Chi-sq Yates p=.1956
Fisher p=.1161

The methods section notes "Chi-square test was used to determine the difference between patients' characteristics in both groups". However, I believe they actually ran Fishers which is appropriate for small N/ cell. Although this is minor, the methods section should accurately describe the statistics that they ran.

**Table 3.** Reported side effects during the immunotherapy treatment period

| Side effects | Immunotherapy ($n$ = 89) | Immunotherapy + cannabis ($n$ = 51) | |
|---|---|---|---|
| Fatigue | 16 (18) | 10 (19) | 10/51=19.607=20% |
| Rash | 3 (3) | 4 (8) | |
| Pruritis | 2 (2) | 0 (0) | |
| Arthralgia | 2 (2) | 2 (3) | 2/51=3.92=4% |
| Abdominal pain | 1 (1) | 0 (0) | |
| Anorexia | 2 (2) | 5 (10) | |
| Hypothyroidism | 8 (9) | 3 (6) | |
| Diarrhea | 2 (2) | 2 (3) | 2/51=3.92=4% |
| No side effects | 53 (60) | 25 (50) | 25/51=49.01 = 49% |

Data are presented as *n* (%).

Again, these 4 errors are very minor (trivial even!) but show a lack of attention to detail by this lab

**Cannabis** impacts tumor response rate to **nivolumab** in patients with advanced malignancies

T **Taha**, D Meiri, S Talhamy, M Wollner, A Peer… - The …, 2019 - academic.oup.com

… treated with **nivolumab** in the years 2015–2016 at our hospital, and **cannabis** from six **cannabis**-… Included were 140 patients (89 **nivolumab** alone, 51 **nivolumab** plus **cannabis**) with …

Save Cite Cited by 120 Related articles All 11 versions

[HTML] nih.gov

Full View

1 comment on PubPeer (by: Phlegmariurus Pittieri)
